# An integrative multi-omics framework identifies epigenetic dysregulation of *HAND2* as a potential primary driver of impaired enteric neural crest cell differentiation in Hirschsprung Disease

**DOI:** 10.64898/2026.06.11.26354426

**Authors:** Stefanie Mellein, Nagarajan Paramasivam, Zuguang Gu, Ralph Roeth, Tanja Mederer, Heike Kuzan, Stephanie Rössler, Jutta Scheuerer, Felix Lasitschka, Constantin Schwab, Felix Sahm, Stefan Hamelmann, Rasul Khasanov, María Ángeles Tapia-Laliena, Lucas Wessel, Michael Boettcher, Leonie Carstensen, Beate Niesler, Britt-Sabina Löscher, Andre Franke, Kübra Narci, Daniel Hübschmann, Gudrun Rappold, Patrick Günther, Christian Schaaf, Philipp Romero

## Abstract

Hirschsprung disease (HSCR) is a congenital neurodevelopmental disorder characterized by segmental aganglionosis due to impaired developmental processes of enteric neural crest cells (NCCs). Despite being the leading genetic cause of functional intestinal obstruction in early childhood, HSCR represents a paradigmatic challenge in precision medicine: its multifactorial etiology, complex gene-environment interactions and limited resolution of single-modality analyses have long hindered mechanistic understanding and therapeutic translation.

Here, we applied an integrative multi⍰omics approach combining genetic, phenotypic, epigenomic and transcriptomic analyses of matched ganglionic and aganglionic formalin⍰fixed paraffin⍰embedded (FFPE) patient tissues, complemented by patient-specific *in vitro* models.

Beyond established genetic contributors, our integrative approach reveals novel regulatory pathways predominantly affecting enteric NCC differentiation, with convergent evidence pointing to epigenetic dysregulation as a primary disease mechanism. Notably, we identified over 1,300 differentially methylated positions between ganglionic and aganglionic FFPE samples, with *HAND2* emerging as a key candidate due to multiple hypermethylated sites and consistently reduced expression levels in aganglionic tissues and *in vitro* models, suggesting a potential role in HSCR pathophysiology.

We propose that our multi-omics approach offers a powerful and comprehensive framework for dissecting disease mechanisms. Beyond advancing biological understanding, this strategy holds promise for paving the way for molecularly informed patient stratification and supporting the development of personalized treatment and postoperative management strategies.

## Introduction

The enteric neuropathy Hirschsprung disease (HSCR, ORPHA: 388) is a congenital malformation characterized by the absence of the enteric nervous system (ENS), also known as aganglionosis, in distinct parts of the colon and rectum. The estimated incidence is 1:5,000 of live births (1). HSCR is caused by impairments in essential developmental biological processes affecting the proliferation, migration, differentiation or survival of enteric neural crest cells (ENCCs). Aganglionosis causes tonic contraction of the affected intestinal segment, with symptoms typically presenting shortly after birth and ranging from delayed passage of meconium and abdominal distension (megacolon) to pain, vomiting, altered bowel habits and severe enterocolitis, potentially leading to failure to thrive (2).

HSCR can either be familial (20%) or sporadic (80%) and is classified by the extent of aganglionosis (3) into short-segment aganglionosis (S-HSCR, 80%, distal rectum to rectosigmoid colon), long-segment aganglionosis (L-HSCR, 17%, sigmoid colon to descending colon or higher) and the most severe type characterized by a total colonic or intestinal aganglionosis (TCA, 3%) (4–5). HSCR is a multifactorial disease with complex pathoetiology and inheritance, variable expression and male predominance, representing the leading genetic cause of functional intestinal obstruction in early childhood (4,6). Genetic factors play a central role in the pathogenesis of HSCR, with about 20 susceptibility genes and loci identified (6–7), including the major susceptibility gene *RET* (8), which accounts for approximately 50% of familial and 20% of sporadic HSCR cases (6,9–11). Recent Genome-Wide Association Studies (GWAS), Genome-Wide Expression Studies (GWES) and Next-Generation Sequencing (NGS) have enabled the identification of new susceptibility genes and variants for HSCR (6). Phenotypic variability and incomplete penetrance in HSCR also suggest the involvement of epigenetic modifications or modifier genes (12).

Due to the complexity of HSCR and its genetic heterogeneity, underlying pathomechanisms are still not fully understood. The current treatment regime is focusing on the alleviation of symptoms by surgically removing the affected aganglionic segment. However, all children with HSCR are at risk for postoperative incontinence (up to 58.3%), enterocolitis (up to 44.9%), and obstructive symptoms (up to 58.8%), regardless of the type of surgical intervention (13–15). Consequently, these persisting symptoms can result in a massive and lifelong impairment of the patients’ quality of life.

Novel therapies targeting patient⍰specific cellular pathomechanisms can offer a promising strategy to restore normal neural innervation, to improve gastrointestinal (GI) function, and to re-establish gut homeostasis (16). The pathogenesis of HSCR is attributable to a multi-factorial interplay of genetic and epigenetic factors but probably also environmental variables (17–18). Focusing on a single factor is of limited use in interpreting the individual cause of the patient’s pathology. However, multi-level modeling can account for this complexity by considering influences at multiple hierarchical levels simultaneously, thereby facilitating the investigation of their interactions and collective contributions to disease development.

We therefore established a multi⍰level framework to dissect individual pathomechanisms and their interactions across various biological layers in HSCR, thereby providing a more comprehensive picture of HSCR. This approach integrated genomic and epigenomic analyses with detailed phenotyping of patients and parents, as well as expression profiling of patient samples, including formalin-fixed, paraffin-embedded (FFPE) samples and complementary *in vitro* models. Finally, epigenomics and transcriptomics data layers were jointly analyzed using multi⍰omics factor analysis. The integrative multi-level approach was applied to a cohort of 33 HSCR patients and identified already known features as clinical differences between S- and L-HSCR patients, as well as pathogenic variants in established HSCR genes. Differential methylation profiling revealed the identification about 1,300 differentially methylated positions (DMPs) between ganglionic and aganglionic FFPE tissues, highlighting the transcription factor *HAND2* and several HOX genes among annotated genes. Multiplex expression profiling revealed significantly reduced *HAND2* expression in aganglionic specimens, supporting its potential role in HSCR pathophysiology. Final multi⍰omics factor analysis confirmed shared regulatory mechanisms between DNA methylation and gene expression, supporting an integrated, multi⍰layered pathogenic model.

Beyond established mechanisms, we identified novel regulatory pathways in HSCR pathogenesis, primarily affecting ENCC differentiation, with integrative analyses suggesting a predominant role for epigenetic regulation.

## Results

This study aims to elucidate the complex interactions between different biological levels to create a more comprehensive and integrated disease profile of patients with HSCR and thus contribute to the development of novel and individualized treatment options (Figure 1).

**Figure 1.**
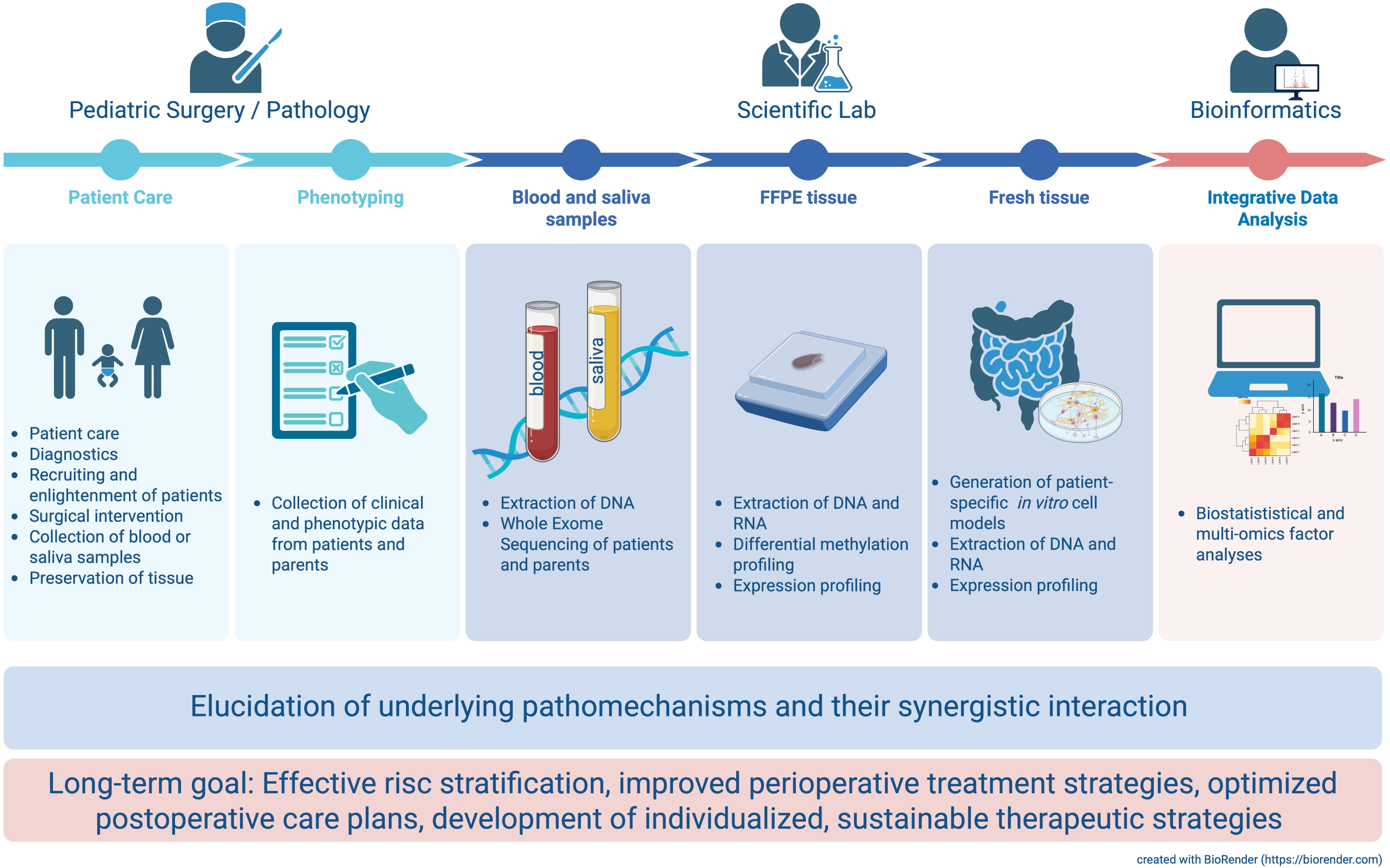
Multi-level analytical strategy to identify the molecular etiology of Hirschsprung disease.

### Clinical and demographic phenotyping of patients and parents reveal a significantly increased rate of associated congenital malformations in patients with long-segment HSCR

We present the evaluation of phenotypic data as well as the perioperative and postoperative course of all included patients, with a postoperative follow-up of one year after corrective surgery for HSCR. A total of 33 patients were included who underwent surgery between 2008 and 2020 in the pediatric surgery departments of the University Hospitals in Heidelberg (n = 29) and Mannheim (n = 4). Demographic data are shown in Suppl. Table 2A. In our cohort, associated anomalies comprised Trisomy 21, congenital kidney and heart malformations, along with a single case of Mowat-Wilson syndrome. Suppl. Table 2B shows the perioperative data of the operated patients. The transanal endorectal pull-through technique represents the standard surgical procedure in 93.9% of cases in the treating pediatric surgery departments. The length of aganglionosis is reported based on the affected bowel segments and as an actual value, with a mean of 18.4 cm, a minimum of 3 cm, and a maximum of 50 cm (Suppl. Table 2B). In this study, L-HSCR (n = 11, 33.3%) was defined as aganglionosis of more than 30 cm. Jirásek-Zuelzer-Wilson syndrome (n = 2, 6.1%) refers to aganglionosis of the entire colorectum. The discrepancy between the mean length of aganglionic segments (18.4 cm) and the resection length (30.1 cm) is explained by the standardized resection above the typically hypoganglionic transition zone in HSCR and the excision of pathologically dilated prestenotic colon segments.

In addition to recording of specific surgical complications such as colorectal anastomotic stenosis or dehiscence, complications that can occur independently of the surgical procedure, were also documented (Suppl. Table 2C). The children received elective and regular postoperative follow-up care. Both Hirschsprung⍰associated enterocolitis (HAEC) and constipation symptoms were treated immediately and, if necessary, in an inpatient setting. The duration of required treatment for constipation with retrograde irrigation or oral macrogol following corrective HSCR surgery was included in the average constipation period, which was 4.1 ± 2.9 months (Suppl. Table 2C). Of note, no significant differences were observed between S⍰ and L⍰HSCR patients in the incidence of HAEC or in postoperative constipation.

Patients with L-HSCR showed a significantly higher rate of associated congenital malformations (odds ratio = 3.7) compared to the rest of the cohort and were born on average earlier and with lower birth weight. Due to the need for longer bowel resections, children with L-HSCR showed, one year after surgery, a significantly higher stool frequency (24 ± 17.6 bowel movements per week) and softer stool consistency with an average of 5.1 on the Bristol Stool Scale, compared to non-L-HSCR patients (Suppl. Table 2D).

Following questionnaire return, data from 35 parents of the 33 HSCR patients were available for analysis, corresponding to a 53% response rate (Suppl. Table 2E). None of the parents was diagnosed with HSCR or a comparable disorder. However, constipation episodes during the one-year observation period prior to the survey were documented in 22.9% of cases (n=8), with a mean episode duration of 4.6 ± 4.5 months. Management of constipation involved retrograde irrigation in 12.5% of cases and oral macrogol therapy in 37.5% of respondents. More than three episodes of gastroenteritis during the observation period were reported by six parents (17.1%). At the time of the survey, nearly one-third of parents (n = 11/35, 31.4%) were following a specific diet, most commonly a ketogenic diet (36.4%), followed by a strict vegetarian diet (27.3%). Parents had a mean age of 39.3 years and an average BMI of 27.3. The mean Rintala Bowel Function Score was 19.4 ± 1.2 (maximum score: 20) and the mean age at attainment of continence was 2.8 ± 0.7 years. No cases of fecal incontinence were reported among the respondents. Overall, 20 parents (53.2%) rated their general health as “very good” and 15 (42.8%) as “good”. None of the parameters listed in Suppl. Table 2E differed significantly between male and female individuals.

### Whole exome sequencing enabled the identification of both established and novel genetic variants

Whole exome sequencing (WES) data were obtained from 29 trios (patients and parents) and 4 single patients (n = 33 patients). To identify genetic variants in the 20 established HSCR genes (6), but also in others, we filtered the WES data in three ways. In all figures, AlphaMissense-based (19) pathogenicity predictions are provided.

Initially, we filtered for rare genetic variants with a minor allele frequency (MAF) < 0.01 and in the 20 known HSCR-associated genes (6). Given their strong association with HSCR (20), also common and non-coding variants in *RET* and *SEMA3D* were included (Figure 2A). All analyzed patients (n = 33) are carrying genetic variants in at least one of the known HSCR genes. Of note, 31 from 33 patients (94%) are carrying rare and/or common SNVs in the *RET* gene and 20 (61%) in *SEMA3D* (Figure 2A). Focusing on other already known HSCR genes (6), 39% and 36% of the patients have rare variants in *ECE1* and *TCF4*, as well as 27% in *NTRK3* and 21% in *NRG1*. Remaining genes contain rare genetic variants in 6-15% of patients (Figure 2A). Furthermore, the data were filtered according to a MAF <0.001, the occurrence of either a stop-gained or missense variant in at least one patient and the inheritance model, considering compound heterozygous, homozygous, hemizygous and *de novo* variants (Figure 2B). According to the applied criteria, 17 genes were identified with missense and/or stop-gained variants in at least one patient. 21% of analyzed patients displayed missense variants in the titin gene (*TTN*). All other identified genes were found in 9% and 6% of patients (Figure 2B). 21 of 29 trio-sequenced patients carry one to four rare *de novo* variants. In two patients, we identified *de novo* variants in already known HSCR genes, *ZEB2* and *PHOX2B*. In addition, one patient carried a *de novo* variant in the *PIAS2* genes, which was already described by us (21). For getting a broader overview about the distribution of genetic variants in analyzed patients, we additionally filtered the data according to a MAF <0.0001 and the occurrence of missense variants, frameshift and inframe deletions and insertions as well as splice variants in at least two patients without considering the inheritance model (Suppl. Figure 1). 27% of patients carry genetic variants in *TTN* (Suppl. Figure 1). In about 18% of the patients, frameshift (n = 5) and missense variants (n = 1) in *MANEA* were identified. 15% of patients are harboring missense variants in the *MDN1* gene. All other variant-carrying genes were identified in 12% and 9% of analyzed patients (Suppl. Figure 1).

**Figure 2:**
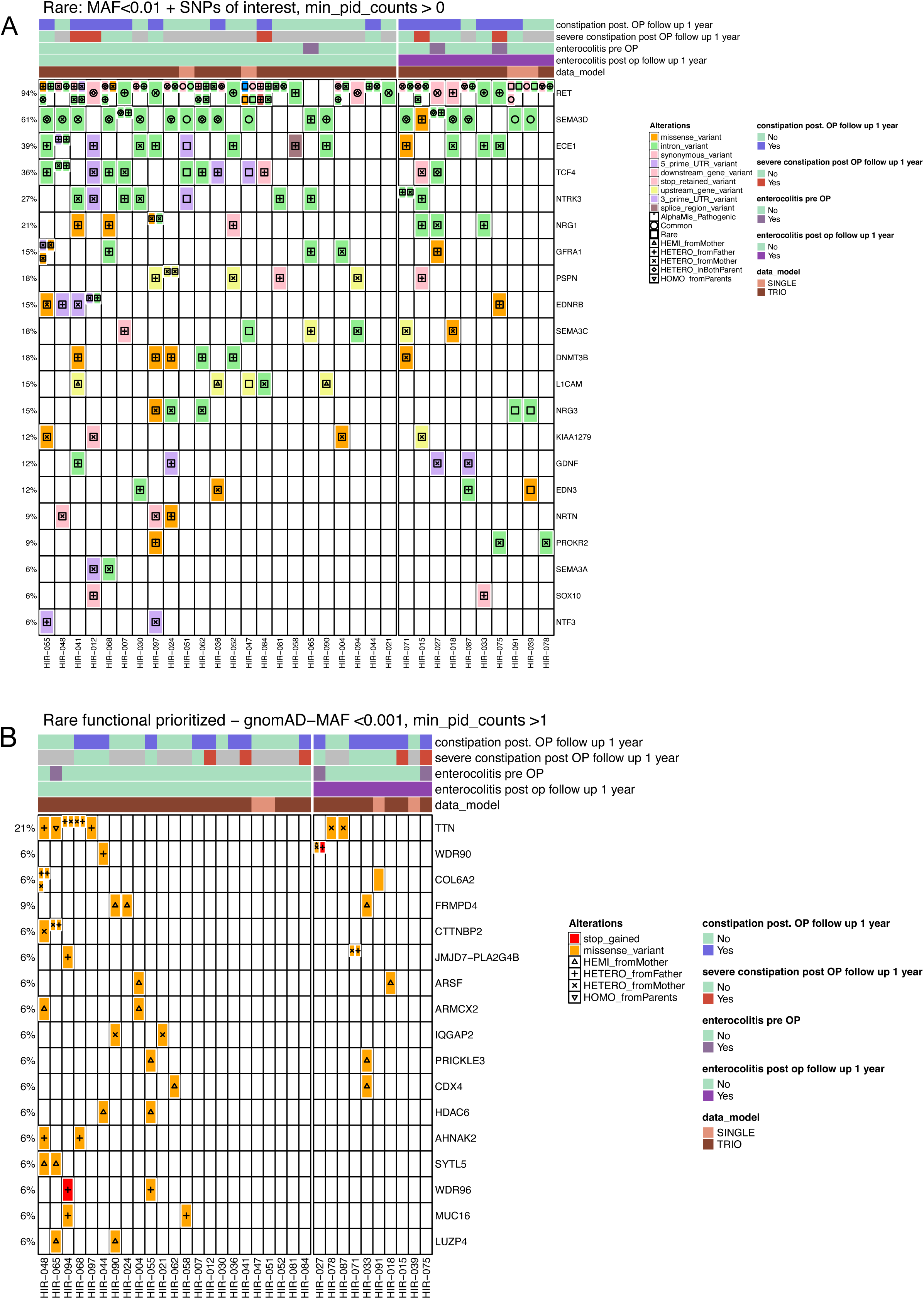
Mutational landscape of common and rare variants in HSCR patients. (A) Overview about rare variants (MAF<0.01) in already known HSCR genes, as well as common variants in *RET* and *SEMA3D*. (B) Occurrence of rare stop-gained and missense variants (MAF < 0.001) in at least one patient according to the inheritance model. The term “Data model” refers to “single” or “trio” whole exome sequencing.

To explore potential genotype–phenotype correlations, we included the phenotypic traits *constipation postoperatively (1 year after surgery)*, *severe constipation postoperatively (1 year after surgery)*, as well as *enterocolitis preoperatively* and *postoperatively (1 year after surgery)* in illustrated heatmaps. However, no obvious genotype-phenotype connections could be detected (Figure 2A, B; Suppl. Figure 1). Furthermore, we examined potential associations between genetic data and the phenotypic traits *sex*, *Trisomy 21* and the *extent of aganglionosis* (short-vs. long-segment HSCR). Also here, no genotype-phenotype correlations could be observed (data not shown).

Overall, no recurring genetic patterns or genotype-phenotype associations could be identified.

### Patient-derived aganglionic FFPE specimens exhibit a significantly distinct expression profile compared with ganglionic samples

To characterize the molecular composition and identify potential differences between affected and non-affected tissue segments, expression profiling of FFPE specimens was performed. FFPE preservation, while essential for long-term storage and histopathological assessment, results in RNA fragmentation and chemical modification, which could result in substantial technical challenges for RNAseq–based transcriptome profiling. At the time expression analyses were conducted, bulk RNAseq was not routinely applied to FFPE specimens, particularly not in the context of limited clinical material. Based on this, we have chosen the nCounter technology for expression profiling, as it is robust to RNA degradation, requires minimal input material and provides highly reproducible measurements across samples, making it particularly suitable for archived clinical specimens.

Corresponding ganglionic and aganglionic tissue-derived FFPE samples from 25 patients were comparatively analyzed. For getting a broad overview about changes in neuronal pathways, the *Neuropathology Panel* was used, which is composed of 770 genes involved in neurotransmission, neuron-glia interaction, neuroplasticity, metabolism, neuroinflammation and cell structure integrity (https://nanostring.com/ncounter-neuropathology/). To investigate also gut-, ENS-and HSCR-specific genes, we additionally applied a self-designed codeset (*Elements Codeset*), composed of 48 genes (Suppl. Table 1).

Using the *Neuropathology Panel*, 82 genes were significantly deregulated between aganglionic and ganglionic tissues. 72 genes achieved only nominally significant *p*-values, while seven and three genes obtained significant adjusted *p*-values for 1% FDR and 5% FDR, respectively (Suppl. Table 3). Applying the Elements Codeset, 13 genes were significantly deregulated, while two with nominal significance, as well as 9 and two with significant adjusted *p*-values for 1% FDR and 5% FDR, respectively (Suppl. Table 4). *UCHL1*, *VIP* and *NOS1* were available on both profiling sets. For simplicity, the results of FFPE samples analyzed with the *Neuropathology Panel* and the *Elements Codeset* will be described jointly (Figure 3A, B).

**Figure 3.**
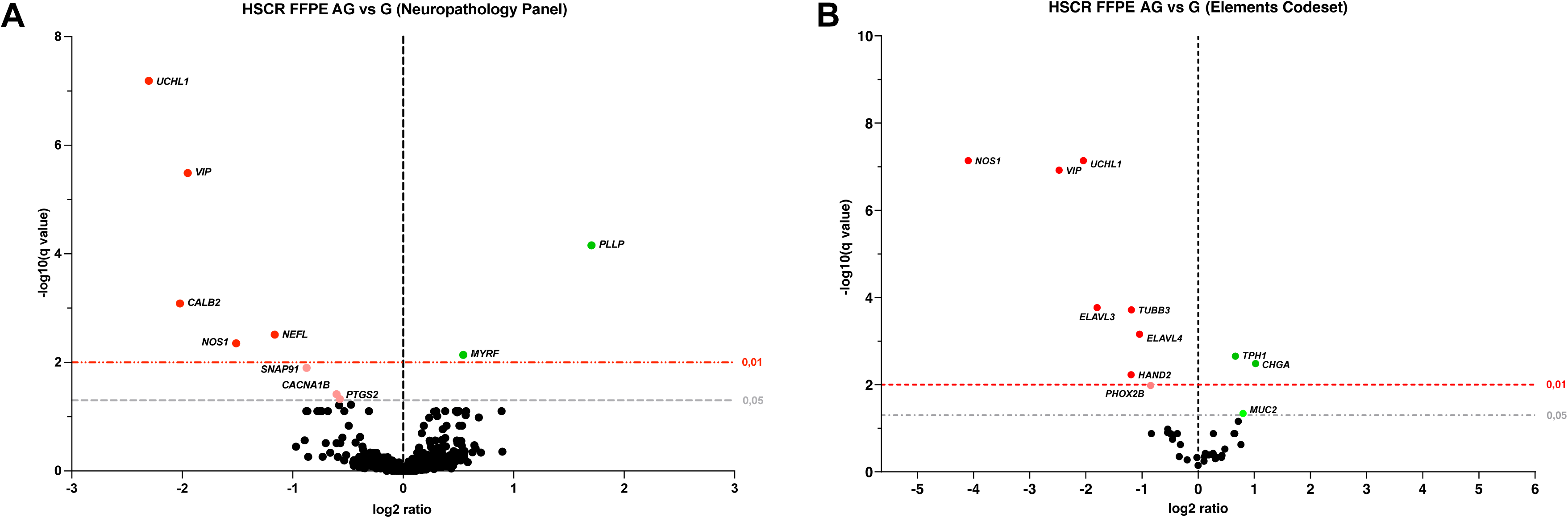
Volcano plot of significantly deregulated genes (after multiple testing) in aganglionic and ganglionic tissue-derived FFPE specimens. Statistically significant expression results obtained with the *Neuropathology Panel* (A) and the *Elements Codeset* (B) are shown. Nominal significant deregulated genes are shown in Suppl. Tables 5 (*Neuropathology Panel*) and 6 (*Elements Codeset*), respectively. Differential expression was tested using the Mann–Whitney U test with 1% FDR correction (indicated by the red line) via the two⍰stage Benjamini–Krieger–Yekutieli method (FDR 5% - gray line).

Independent of the applied profiling set, the neuronal markers *UCHL1* and *VIP* were amongst the most significantly downregulated genes in aganglionic compared to ganglionic specimens. According to an FDR < 1%, also *CALB2* (Figure 3A), and other neuronal markers such as *ELAVL3*, *ELAVL4*, *HAND2*, *TUBB3* (all Figure 3B), *NEFL* (Figure 3A) and *NOS1* (Figure 3A, B) were significantly downregulated. *NOS1*, displayed two different results in shown plots (Figure 3A, B), which can be explained by the usage of different probes in the applied profiling sets. However, in both cases, *NOS1* was amongst the highest significantly deregulated genes with an FDR <1%. Additionally, *CACNA1B*, *PTGS2, SNAP91* (Figure 3A) and *PHOX2B* (Figure 3B) displayed significantly reduced expression levels in aganglionic samples with an FDR <5%. Furthermore, aganglionic FFPE specimens displayed significantly higher expression levels for the epithelial markers *CHGA* and *TPH1* (FDR < 1%) as well as *MUC2* (FDR < 5%) (Figure 3B) but also for the myelination markers *MYRF* and *PLLP* (FDR < 1%) (Figure 3A). Results of significantly deregulated genes, including also nominal significance, are given in the Supplementary (Suppl. Table 3, Suppl. Table 4). Amongst nominal significantly deregulated genes, the four known HSCR genes, *RET*, *GDNF*, *NRG1* and *L1CAM* are listed.

### Assessment of patient-derived ganglionic and aganglionic cell culture models reveal no morphological changes on cellular level, while molecular differences are observed

In contrast to FFPE tissue, patient-specific cell models enable the modeling of the disease within an individual genetic context and allow for the detailed investigation of molecular processes at the cellular level. These cell models are based on enteric neurospheres, which form under optimal conditions from isolated epithelial, muscle, neuronal, stem and progenitor cells derived from respective tissue specimens (22).

Here, patient⍰specific cell culture models were established from freshly resected ganglionic and aganglionic gut specimens of prospectively enrolled patients. After 10 days in non-adherent culture, numerous enteric neurospheres were observed across all patient samples (n = 9), with no significant variation in their size or number relative to tissue origin (ganglionic or aganglionic) (Figure 4AI, BI). Most ganglionic and aganglionic enteric neurospheres were subjected to neuronal differentiation conditions. Overall, we successfully established differentiation cultures from nine patients. Also here, brightfield imaging did not reveal any noticeable differences in differentiation behavior or cell morphology (Figure 4AII, BII). Additionally, immunofluorescence analyses were performed for main occurring cell types as neuronal (TUBB3, MAP2), neuronal progenitor (P75, Nestin) and glial cells (S100ß). No relevant differences in the staining pattern of ganglionic or aganglionic cultures could be observed (Figure 4AIII-V, BIII-V).

**Figure 4.**
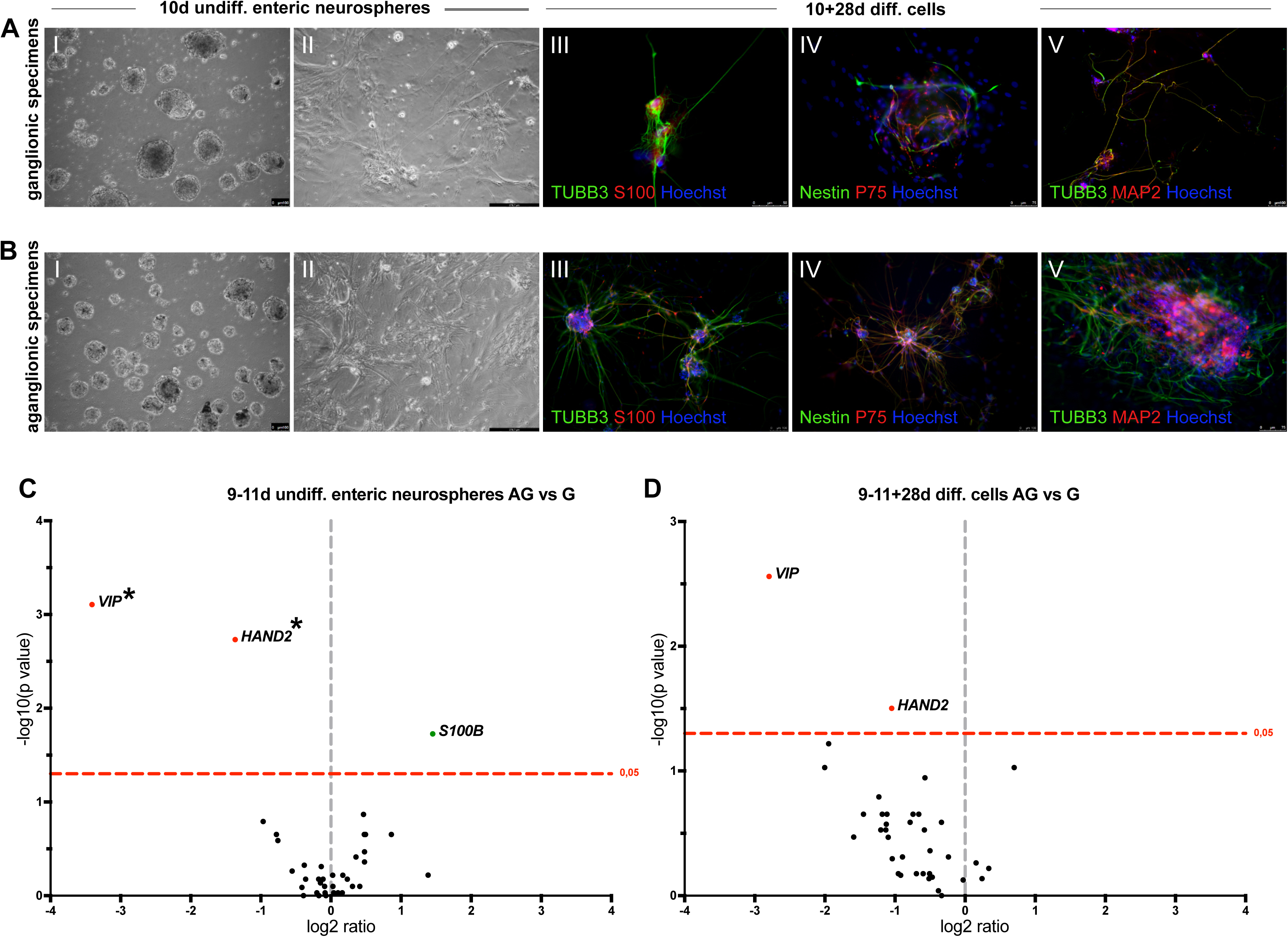
Comparative analyses of patient-derived *in vitro* samples. (A) Representative brightfield images of ganglionic tissue-derived undifferentiated enteric neurospheres (AI: 10d) and differentiated ENS-like cells (AII: 10+28d), as well as immunofluorescence staining of differentiated cells (A: III-V) applying the neuronal markers TUBB3 (green) and MAP2 (red), the glial cell marker S100ß (red) and the progenitor cell markers P75 (red) and Nestin (green). Nuclei are stained with Hoechst. (B) Corresponding representative brightfield images of aganglionic tissue-derived undifferentiated enteric neurospheres (BI: 10d) and differentiated cells (BII: 10+28d), as well as immunofluorescence staining of differentiated cells (B: III-V) according to the ganglionic samples (see A). (C) Volcano plot of undifferentiated aganglionic vs ganglionic enteric neurospheres at day +/- 10 applying the customized *Elements Codeset* (n=48 genes). (D) Volcano plot of differentiated aganglionic vs ganglionic cells at day +/- 10+28d applying the customized *Elements Codeset*. Nominal and adjusted *p*-values are shown. Black asterisks: significant according to adjusted *p*-value. undiff.: undifferentiated, diff.: differentiated, AG: aganglionic, G: ganglionic.

Additionally, comparative expression profiling of *in vitro* generated samples was performed. To ensure a level of comparability across analyses, we have also applied the nCounter technology. Due to limited input material, only the self-designed *Elements Codeset* (n = 48 genes) was applied (Figure 4C, D). Expression and statistical analyses were restricted to complete, patient-matched sample sets comprising ganglionic and aganglionic undifferentiated enteric neurospheres (9-11d) and differentiated cell models (9-11+28d). Expression profiling of 9-11d ganglionic and aganglionic enteric neurospheres displayed significantly altered expression levels in aganglionic specimens for *VIP* (*p*-value: 0.000782/*q*-value: 0.028723), *HAND2* (*p*-value: 0.001851/*q*-value: 0.034014) and *S100B* (*p*-value: 0.018758/*q*-value: 0.229782). While *VIP* and *HAND2* expression levels were significantly downregulated, *S100ß* levels were significantly upregulated in aganglionic specimens (Figure 4C).

Expression profiling of differentiation cell models at stage 9-11+28d revealed a significant downregulation of *HAND2* (*p*-value: 0.031469/*q*-value: 0.611276) and *VIP* (*p*-value: 0.002756/*q*-value: 0.107073). However, compared to undifferentiated specimens, a slight upregulation of *HAND2* and *VIP* could be observed for differentiated aganglionic samples (Figure 4D).

Based on the microscopic findings, we hypothesize that examined patients most likely suffer from differentiation defects of ENCC cells rather than migration defects, as aganglionic cells behaved in a similar way as their ganglionic counterparts. We therefore had a closer look on stem and neuronal progenitor marker expression in *in vitro* samples but also in FFPE specimens (Suppl. Figure 2). In FFPE samples, similar to slightly higher expression levels of *NES*, *NGFR*, *POU5F1* and *SOX10* could be observed in aganglionic compared to ganglionic samples*. In vitro* samples displayed similar results; however, expression levels of above-mentioned markers are constantly higher in undifferentiated aganglionic tissue-derived neurospheres than in ganglionic ones. In differentiated aganglionic samples, overall expression levels are decreasing, while they are slightly higher in aganglionic than in ganglionic samples (Suppl. Figure 2). In summary, our results suggest differentiation defects of ENCCs rather than migration defects.

### DNA methylation profiling of ganglionic and aganglionic FFPE specimens enabled the identification of differentially methylated positions and key genes

Following nCounter-based expression analyses, we conducted differential methylation profiling on the same ganglionic and aganglionic FFPE samples (n = 26 patients) to explore potential regulatory mechanisms. While expression profiling captures the functional transcriptional state, epigenetic modifications such as DNA methylation provide insight into upstream and potentially persistent regulatory changes. Analyzing both molecular layers in identical tissue specimens enables integrative interpretation while minimizing biological variability, thereby strengthening the robustness of the findings. We applied consensus *k*-means clustering on a total of 52 samples and identified a subset of 38 samples exhibiting stable classification between ganglionic and aganglionic groups. To enhance the robustness of subsequent analyses, differential methylation analysis was restricted to stably classified samples from groups 1 and 3 (Suppl. Figure 3).

In total, 1347 significantly differentially methylated positions (DMPs, *q*-value < 0.01, Figure 5A, Suppl. Table 5) could be identified. In Figure 5B the genomic distribution of identified DMPs is illustrated. The column bar graphs show the number of significantly methylated CpG probes categorized by gene-features, such as promoters or gene bodies (exons, introns, intergenic regions). The *y*-axis indicates the number of probes with potential differences between hyper- and hypomethylated sites. Hypo-(G>AG)- and hypermethylated (G<A) CpG probes are broadly distributed across the genome, with most sites located in intronic and intergenic regions. Over 300 DMPs map to exons, whereas approximately 100 are found in promoter regions (Figure 5B). Most DMPs lie ∼800 bp away from CpG islands (CGIs) and transcription start sites (TSS), although hypermethylated DMPs are closer to these features than hypomethylated ones (Figure 5C). As shown in Figure 5D, overall, more hypo-than hypermethylated DMPs could be identified.

**Figure 5:**
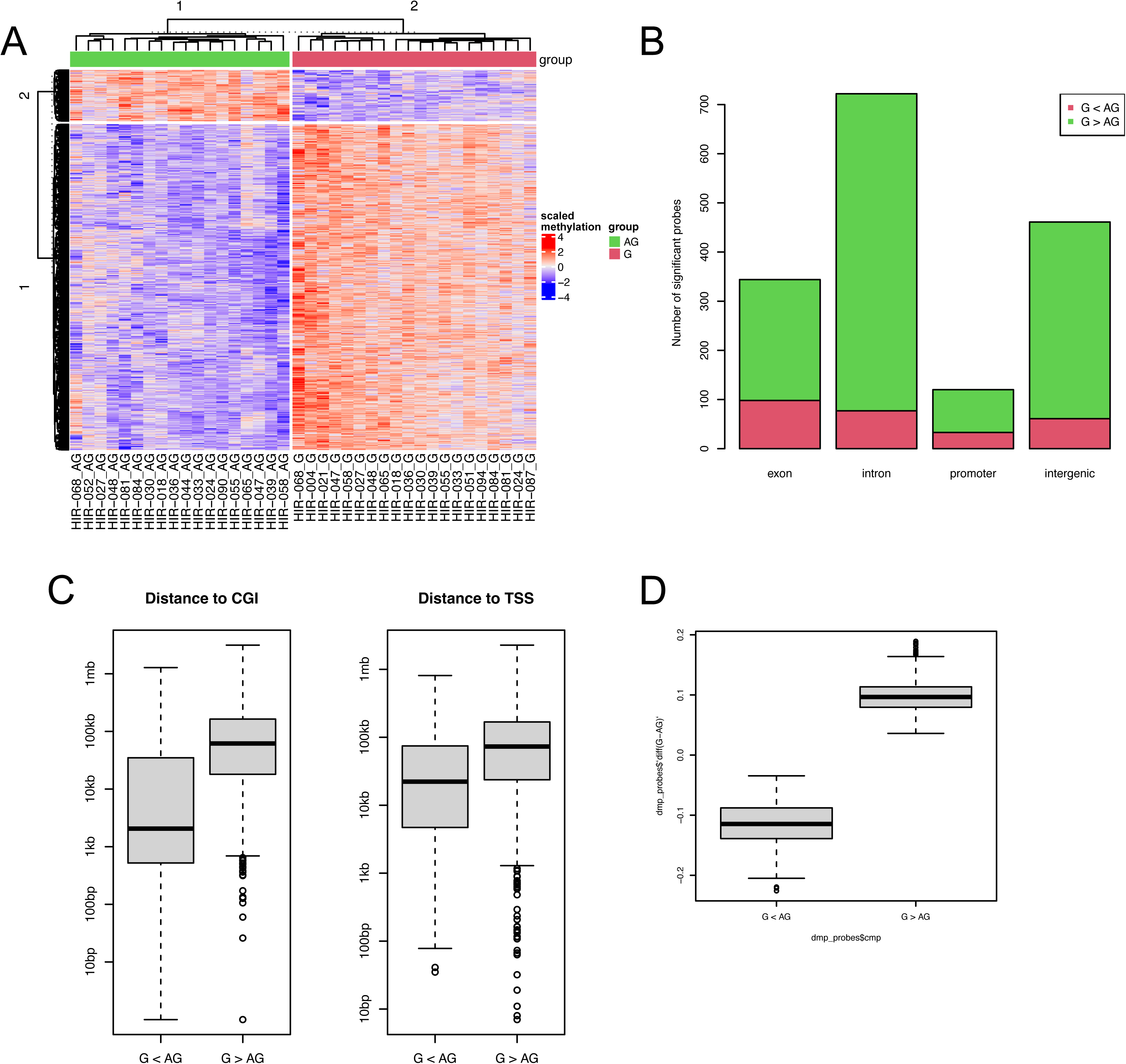
Differentially methylated positions (DMPs). (A) In total, 1,347 DMPs could be identified using the *dmpFinder* tool. (B, C) Numbers of significantly methylated CpG probes in B) CGI-features and C) gene-features. (D) Overview of hyper- and hypomethylated DMPs in aganglionic compared to ganglionic specimens. G < AG: hypermethylated in aganglionic tissue. G > AG: hypomethylated in aganglionic tissue.

To further characterize the methylation pattern of established HSCR-associated genes and to identify potential novel candidates, DMPs were mapped to the nearest gene based on its TSS. In total, 1,126 genes could be identified, including 1,004 potentially hypomethylated and 120 hypermethylated genes (Suppl. Table 5). To be able to identify genes, which have been already associated with HSCR, we initially aligned the gene list with established HSCR genes (6). Among hypermethylated genes, no known HSCR genes could be identified, whereas *EDNRB* and *GDNF* could be observed in potentially hypomethylated genes (Suppl. Table 5). To explore the biological significance of the observed methylation changes, we used the publicly available web platform *Metascape* (https://metascape.org) (23), offering comprehensive gene annotation and enrichment analysis capabilities.

Gene Ontology (GO)-based and pathway enrichment analysis revealed several developmental-related terms as *tube* (GO:0035239), *tissue* (GO:0048729) and *embryonic* (GO:0048598) *morphogenesis*, *brain development* (GO:0007420), or *regulation of nervous system development* (GO:0051960), *digestive tract development* (GO:0048565), but also *negative regulation of cell differentiation* (GO:0045596), *regulation of WNT signaling* (GO:0030111), *Signaling by GPCR* (R-HAS-372790), *Oxytocin signaling pathway* (hsa04921), *neural precursor cell proliferation* (GO:0061351), as well as *Neuronal System* (R-HSA-112316) and *Axon guidance* (hsa04360) and many more (Figure 6A). Finally, Metascape provided an enrichment analysis of diseases based on DisGeNET (https://disgenet.com/), which includes *Cleft Palate*, *Developmental Disabilities*, *Congenital Heart Defects*, and *Neurodevelopmental Disorders* amongst others (Figure 6B_Enrichment analysis).

**Figure 6:**
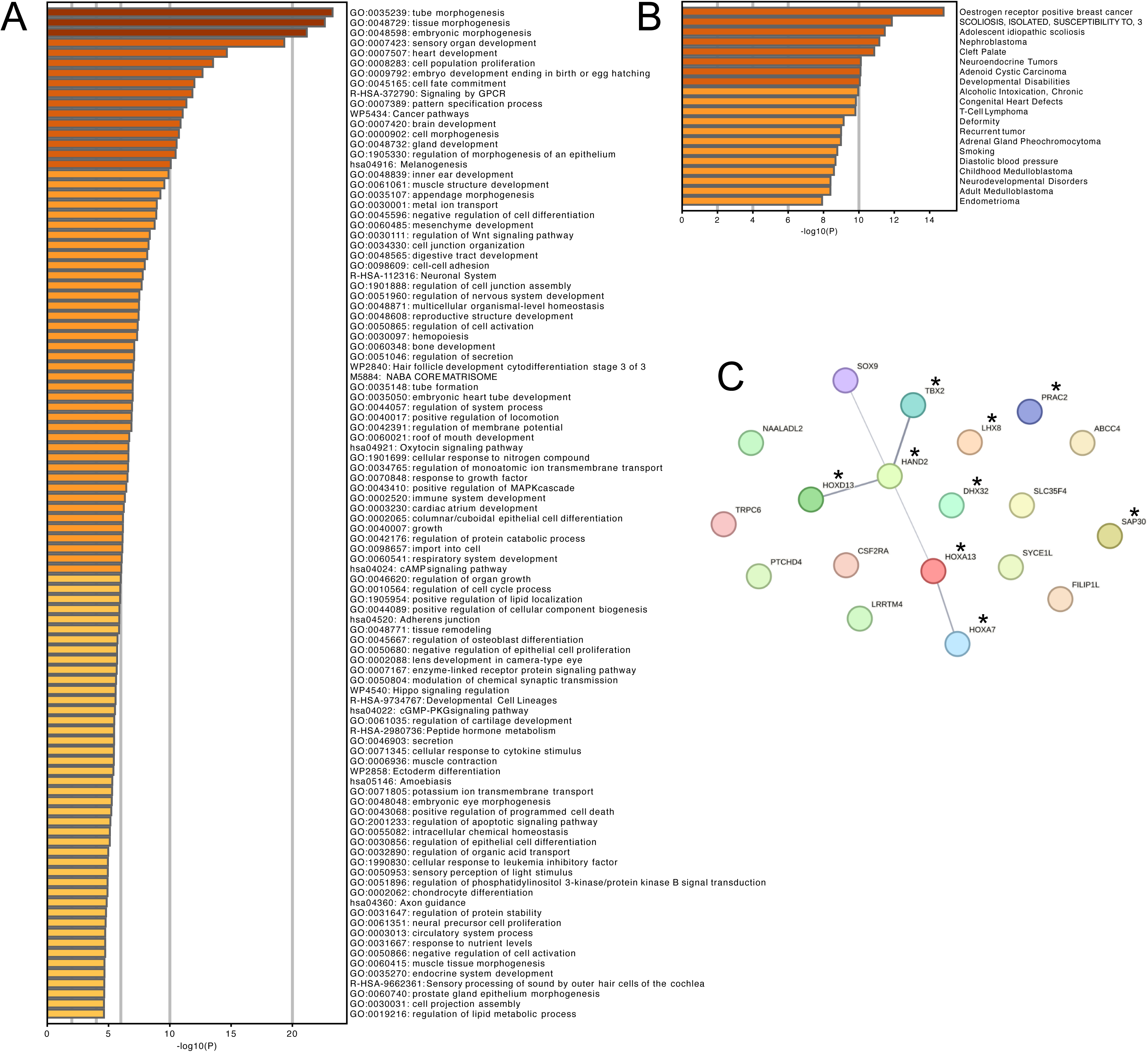
Metascape and STRING-based enrichment and network analyses. (A) Heatmap of GO term and pathway enrichment analysis of significantly hyper- and hypomethylated DMPs in aganglionic specimens. (B) STRING-network analysis of TOP20 DMP-related hyper- and hypomethylated genes. (C) Disease enrichment analysis. Asterisks indicate DMP-related hypermethylated genes.

To further explore potential protein–protein interactions, we conducted a network analysis using the STRING database (https://string-db.org/) (24) based on the TOP 20 DMP-assigned hyper- and hypomethylated genes (Table 3). A small network could be identified composed of several proteins as CSF2RA, EVX1, EVX2, HAND2, HOXA7, HOXA13, HOXB5, HOXD13, IKZF1, MEF2C, SOX9 and TBX2 (Figure 6C).

Since expression profiling of patient samples revealed significant downregulation of *HAND2* and differential DMP analysis identified *HAND2* among the top 20 DMP-associated hyper- and hypomethylated genes, we performed immunofluorescence staining to support these findings. Indeed, we could show that in aganglionic samples almost no expression of HAND2 was visible, while in ganglionic specimens HAND2-positive cells could be observed in submucosal and myenteric ganglia (Suppl. Figure 4).

### Multi-omics factor analysis identified five latent factors, indicating strong shared patterns across the integrated omics layers

A Multi-Omics Factor Analysis (MOFA) aids in recognizing the relationships between different types of biological data and understanding how they jointly influence disease onset or progression. Given their comparable data structures (AG vs G comparisons), results from gene expression analyses, using both the prebuild *Neuropathology Panel* and the self-designed *Elements Codeset*, alongside DMPs from methylation profiles, were incorporated into the integrative MOFA framework (Figure 7A). Selected clinical and phenotypic traits (Suppl. Table 6) of patients are used as covariates.

**Figure 7:**
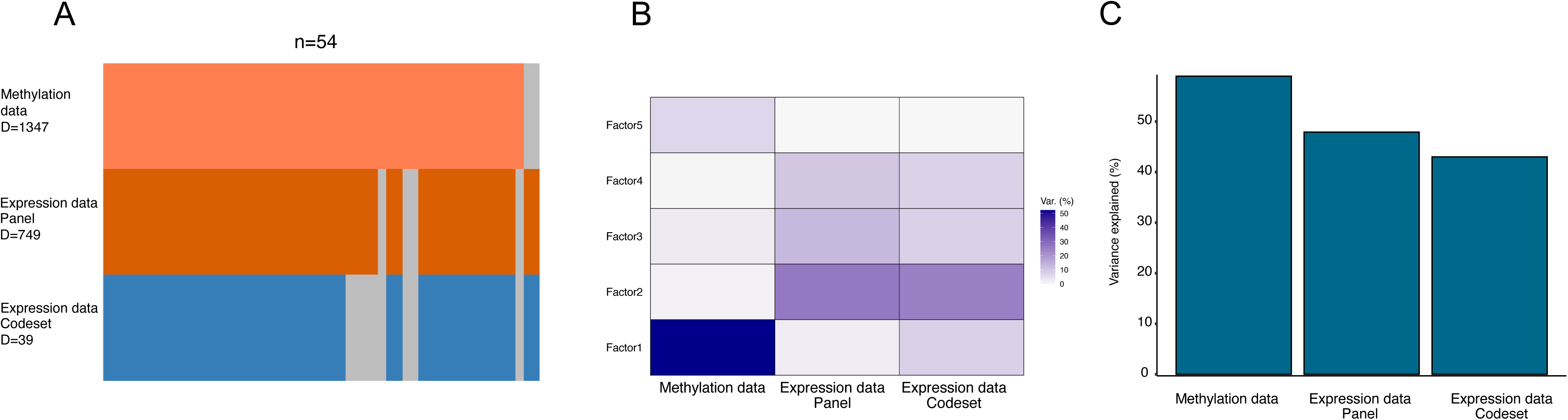
Multi-omics integrative analysis. (A) Data modalities. (B) The proportion of the total variance (R²) attributed to each factor for each data type. (C) Cumulative proportion of total variance. D: number of features.

In total, MOFA identified five latent factors with minimum explained variance of 43% in at least one data type (expression data – *Elements Codeset*) (Figure 7B, C). Factors 1, 2 and 3 are active in all analyzed data, indicating an important role in multiple molecular layers. Factor 4 is only active in two of the three data layers and Factor 5 only in a single data modality (Figure 7B). This distribution indicates that most of the biological signal is captured by Factor 1, suggesting the presence of strong shared patterns across the integrated omics layers. This dominant factor likely reflects major biological processes or experimental conditions influencing multiple data modalities, while the remaining factors may represent more specific or subtle sources of variation. In detail, Factor 1 methylation data explain about 49% of variance, while *Elements Codeset*-derived expression data explain 7% and *Neuropathology Panel*-derived expression data 2% of variance (Figure 7B). In total, the five factors explain about 58% of variation in the methylation, 48% in the *Neuropathology Panel*-derived expression data sets and 43% of variation in the *Elements Codeset*-derived expression data (Figure 7C).

The correlation of covariates (Suppl. Table 6) to identified factors from the MOFA is shown in Suppl. Figure 5. Factors 1, 2 and 3 are specific for the variance regarding the sample origin (*AG_G_class*), showing that ganglionic and aganglionic samples can be clearly distinguished from each other (Suppl. Figure 5A, B). Factor 1 further relates to the *length of the resected gut segment*, *the length of the aganglionosis*, *comorbidities* and *Trisomy 21*. Factor 2 is only active for the covariate *constipation_postoperatively_overall*. Factor 3 relates to *AG_G_class*, *enterocolitis_postoperative_1 year* and *long segment HSCR*. Factor 4 is only active for the *AG_G_class*, whereas Factor 5 is active for *birth weight* and *constipation_postoperatively_after 1 year, -overall* and *sex* (Suppl. Figure 5A).

In conclusion, the MOFA revealed five latent factors, suggesting the presence of robust shared molecular patterns across the integrated omics layers.

### Factor 1 features highlight epigenetic regulators of ENCC differentiation, with HAND2 linking methylation and expression signatures

Feature weights were plotted to visualize how individual methylation and expression features contribute to each factor. Significantly identified results of aganglionic FFPE specimens and for the most meaningful factor, Factor 1, are shown (Figure 8).

**Figure 8:**
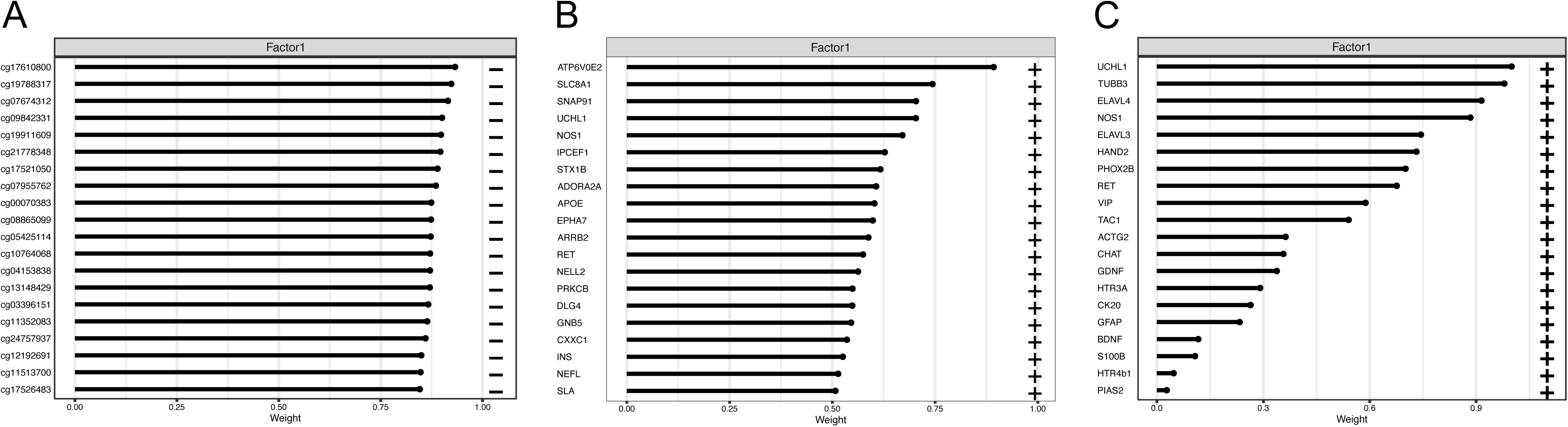
Top features identified via MOFA analysis. Factor 1 results for (A) methylation data, (B) for *Neuropathology Panel*-derived expression data and (C) for *Elements Codeset*-derived expression data.

Metascape and STRING analyses were conducted using all features from the respective methylation and expression analyses to further investigate the biological functions of the identified and annotated genes associated with them.

The strongest absolute weight in Factor 1 mapped to a CpG site in *HOXA13*. Further CpG sites could be mapped for example to *HAND2* and *EVX1*. While *EVX1* showed the shortest distance to a TSS, suggesting promoter-proximal methylation likely leading to transcriptional silencing (Figure 8A, Table 4), most of the other CpGs are located at considerable distances from TSS, which relates to distal, non-promoter methylation changes, potentially influencing enhancer or structural functions rather than directly controlling the start of transcription (Figure 8A, Table 4). Enriched GO terms and diseases, as *regulation of mitotic cell cycle* (GO:0007346) and *negative regulation of differentiation* (GO:0045596) as well as *Congenital Heart Defect*, *Developmental Delay* and *Cleft Palate* are given in Suppl. Figure 6A. Between HAND2, HOXA3, HOXA13, EVX1 and MEIS2 as well as SMAD7 and COL4A1 protein-protein interactions could be observed (Suppl. Figure 6A).

The strongest absolute weight in Factor 1 of the *Neuropathology Panel*-based expression dataset corresponds to *ATP6V0E2*, which was not described in the context of HSCR so far. Further interesting genes are *SLC8A1*, *SNAP91*, *UCHL1*, *NOS1, RET* and *NEFL*, which are all associated somehow to HSCR (Figure 8B, Table 4). Amongst enriched GO terms and pathways are *modulation of chemical synapse transmission* (GO:0050804), *neuron projection development* (GO:0031175), *regulation of neurotransmitter transport* (GO:0051588), *regulation of neuron apoptotic process* (GO:0043523) and *Calcium signaling pathway* (hsa04020). Amongst enriched diseases are *Peripheral Nervous System Disease*s, *Peripheral Neuropathy*, *Tauopathies* and some more. The STRING network analysis indicated that most of the genes, except *NELL2*, *RET*, *SLC8A1*, *CXXC1*, *STX1B*, *SLA*, *IPCEF1* and *EPHA7* are associated with each other (Suppl. Figure 6B).

In the *Elements Codeset-*based expression data, *UCHL1* showed the strongest absolute weight, closely followed by *TUBB3*. Additional genes potentially relevant for HSCR ethiology are *ELAVL4*, *NOS1, ELAVL3*, *HAND2*, *PHOX2B*, *RET*, *VIP*, *GDNF* and *PIAS2* (Figure 8C, Table 4). GO enrichment results included amongst others *cell morphogenesis involved in neuron differentiati*on (GO:0048667), *autonomic nervous system development* (GO:0048483), *regulation of neurotransmitter uptake* (GO:0051580) and *cell population proliferation* (GO:0008283). Disease-enrichment analysis identified besides *Peripheral Nervous System Diseases*, *Constipation*, *Peripheral Neuropathy* and *HSCR*. According to STRING network analysis, almost all markers exhibit interconnections, except ACTG2, KRT20 and PIAS2 (Suppl. Figure 6C).

In summary, most Factor 1-associated genes are linked to ENS development, particularly processes such as cell migration and differentiation.

## Discussion

Understanding HSCR pathomechanisms and advancing individualized therapies is of high relevance to both researchers and affected families, as reflected by over 7,800 PubMed-listed publications. However, due to its complexity and heterogeneity, underlying pathomechanisms are still not completely unraveled. The partial lack of detailed and comprehensive patient-specific information makes the development of novel treatment strategies difficult and tedious.

The application of multi-level/multi-omics approaches can provide valuable information from different biological layers to dissect and to investigate underlying pathomechanisms (25). Further, they can offer new avenues for future diagnosis and prevention strategies in HSCR as well as for personalized therapeutic interventions. A small number of multi-level/multi-omics studies have already been conducted (21,26–27), which have provided valuable insights into HSCR pathogenesis, but no study has examined such a large patient cohort across as many comprehensive biological layers as the presented study.

We employed a multi-level strategy, conducting a systematic analysis of each biological layer both independently and partially in combination through an integrative MOFA analysis. Our approach comprises phenotypic, genetic and epigenetic data, as well as expression profiling data from patient-derived paraffin-embedded tissue specimens and complementary *in vitro* models. A major strength of our study is the consistent analysis of the same set of patients and the consequent use of corresponding ganglionic and aganglionic patient-derived specimens. Furthermore, what sets our study apart from others, is the assessment of patient-derived FFPE and *in vitro* specimens.

In the first layer of our pipeline, we conducted phenotyping of patients (n = 33) and parents (n = 35). Our analyses revealed notable differences in congenital malformations, gestational, growth as well as stool parameters between S-HSCR and L-HSCR patients. Stool parameters as frequency and consistency can be altered in L-HSCR patients, as due to the surgical resection of long pieces, the transit time of stool can be shortened, which may lead to increased stool frequency and reduced water resorption from the stool. Higher rates of associated congenital malformations as Trisomy 21, as well as altered gestational and growth parameters in L-HSCR patients might be linked to a higher genetic variability in those patients.

None of the parents (n = 35) were diagnosed with HSCR or a comparable disorder. However, 8 out of 35 individuals reported constipation, while stool frequency and consistency were not conspicuous between parents with or without constipation.

WES analyses revealed the presence of inherited or *de novo* variants in at least one established HSCR gene (6), but also in novel HSCR-associated genes. In 94% of patients, rare or common variants in the major susceptibility gene *RET* could be identified. Variants in *RET* were also detected in unaffected parents with constipation, sometimes alongside *SEMA3D* variants, underscoring the incomplete penetrance and multifactorial nature of HSCR. These findings support previous reports indicating that variants in one or two ENS⍰related genes are insufficient for disease manifestation but may contribute to subclinical phenotypes as constipation. These findings are also in line with previous publications, emphasizing that the development of HSCR requires the presence of multiple variants across key developmental pathways (7,20). Variants in *MANEA* and *MDN1* were identified in 18% and 15% of patients, respectively. *MANEA* is involved in N⍰glycosylation (28), while *MDN1* encodes an AAA ATPase required for ribosome maturation (29). Neither gene has previously been directly linked to HSCR or ENS development. Overall, the absence of recurrent genetic patterns or clear genotype-phenotype associations underscores the remarkable heterogeneity of HSCR, while also highlighting the challenges inherent to exploring such diversity within a small cohort.

Within the next biological level, multiplexed expression profiling of ganglionic and aganglionic FFPE and *in vitro* samples was performed. FFPE preservation introduces challenges as RNA fragmentation and chemical modifications. At the time expression analyses were conducted, bulk RNAseq was not routinely applied to FFPE specimens. To ensure proper analysis of this compromised material, the nCounter technology was chosen, as it is very robust to RNA degradation. To address ENS⍰related molecular changes while also capturing epithelial, stem and smooth muscle cell-associated alterations, a pre-defined marker panel (*Neuropathology Panel)* and a customized *Elements Codeset* were used.

Expression profiling of FFPE specimens revealed significant downregulation in aganglionic samples of mature neuron markers such as *UCHL1*, *TUBB3*, *ELAVL3*, *ELAVL4* and others. One of the most interesting markers amongst them, is the transcription factor *HAND2*. Multiple studies have shown that Hand2 plays a pivotal role in the differentiation of enteric precursor cells into enteric neurons (30), as well as in neural network patterning in the developing ENS and neurotransmitter specification (30–31). Amongst others, D’Autréaux et□al. showed that *Hand2* haploinsufficiency in mice impairs ENS development, with reduced *Elavl4* (HuD) expression, also seen in our analysis, fewer nNOS⍰ and calretinin⍰positive neurons, and overall fewer, smaller ENS neurons (30). The deletion of *Hand2* in murine neural precursor cells even led to a complete loss of Vip and nNos (*Nos1*) (31). Its relevance to the HSCR pathophysiological network was emphasized e.g. by differential expression approaches showing decreased expression levels of *HAND2* in aganglionic compared to ganglionic tissues (32–33) and quite recently due to the identification of *HAND2*, together with three additional genes, as novel non-exonic susceptibility loci in a multi⍰ancestry GWAS meta⍰analysis [71□. None of these loci were detected in our patients, as they are not covered by exome sequencing.

nNOS (*NOS1*), significantly downregulated in our analyzed aganglionic specimens, is expressed by subsets of ENCCs that migrate along axon-like processes, where it contributes to the spatial and temporal coordination of NCC migration. Reduced or absent nNOS disrupts this guidance, resulting in a disorganized distribution of NCCs, neural precursors and neurons (31,34). In addition, nNOS is expressed by enteric nitrergic neurons, which regulate gut motility through nitric oxide–mediated smooth muscle relaxation. Loss of these neurons is a hallmark of enteric neuropathies, including HSCR (35–37). VIP, a key ENS neurotransmitter that inhibits colonic smooth muscle tone, was strongly downregulated in our analysis; early studies similarly reported reduced VIP levels in aganglionic HSCR bowel, implicating VIP deficiency contributes to impaired segmental relaxation (38).

Overall, these findings identify *HAND2* as a very promising candidate for follow⍰up studies, potentially highlighting differentiation defects of ENCCs in HSCR patients.

A major advantage of analyzing full□thickness FFPE tissue is the ability to capture molecular information from all constituent cell types, including neuronal, epithelial, and muscle cells. In addition to neuronal markers, *CALB2*, a Ca²□□binding protein expressed in enteric neurons (39) and used as a diagnostic marker for HSCR (40), was amongst the significantly downregulated genes and therefore functioning as a proof-of-concept marker. Other significantly downregulated markers are *CHGA*, *TPH1*, *MYRF* and *PLLP*. CHGA is expressed by enteroendocrine cells (41), as well as in enteric ganglia (42–43). TPH1 is expressed by enterochromaffin cells, which are involved in motility processes (44). Both markers are upregulated in assessed aganglionic specimens, which is in line with published data, claiming that expressing cell types are increased in aganglionic segments of HSCR patients (45), whereas elevated abundance of these cells may contribute to sustained contraction of the aganglionic bowel (45). Additionally, *MYRF* and *PLLP*, both linked to myelination-related programs, are significantly upregulated in aganglionic specimens. MYRF regulates myelin development, is required for oligodendrocyte differentiation (46) and is expressed in the postnatal intestine (47), while PLLP influences Notch signaling and intestinal epithelial differentiation. When PLLP is absent, Notch signaling is disrupted, which leads to defects in intestinal epithelial cell differentiation (48). Enteric glial cells are known to express genes typically associated with Schwann cells, oligodendrocytes, and other myelinating glia, despite the absence of myelination in the murine ENS (49). Additionally, a recent study described a microbiota–myelin axis, demonstrating that the gut microbiota modulates myelination by coordinating metabolic signaling, immune homeostasis, and neuroinflammatory responses (50). How MYRF⍰ and PLLP⍰associated pathways contribute to HSCR and whether they are involved in the pathogenesis of HSCR-associated enterocolitis characterized by epithelial barrier dysfunction, altered microbiota, impaired enteric innervation but also with intestinal epithelial barrier disruption (51), warrants further investigation.

While the nCounter approach yields informative data for predefined gene sets, it limits detection of broader transcriptional changes. To strongly link genetic and epigenetic alterations with downstream transcriptional effects, we plan to perform comprehensive RNA⍰seq approaches, particularly snPATHO⍰seq for single⍰cell profiling of FFPE tissues (52). Nevertheless, the nCounter technology remains well suited for RNA⍰seq validation.

In addition to FFPE samples, we established ganglionic and aganglionic cell culture models from prospectively enrolled patients, using undifferentiated enteric neurospheres as the basis for generating more mature neuronal⍰like cells. No obvious macroscopic differences were observed between undifferentiated and more advanced cell models, irrespective of their origin. Overall, it seems, that the application of appropriate supplements for initiation of neuronal differentiation let the aganglionic⍰derived cells closely resemble their ganglionic counterparts. On transcript level, we could show that *HAND2* expression increased during differentiation of aganglionic cells, indicating a potential reactivation of neuronal differentiation processes. As we repeatedly identified *HAND2* as a deregulated key marker in aganglionic specimens, we hypothesize that in most patients, ENCCs are successfully colonizing the developing gut but fail to mature into functional enteric neurons, remaining instead as dormant neuronal progenitors. This is in line with our expression findings and of a recent scRNA-seq study, in which the authors hypothesized that alterations in gene expression within the neural progenitor cell microenvironment, most likely driven by changes in mast cells, result in impaired differentiation (53). In this scenario, the investigated patients do not exhibit migration defects of ENCCs but rather defects in their differentiation. The application of suitable supplements under *in vitro* conditions, as observed in our experiments, seem to reactivate their neuronal differentiation program, at least to a certain extend.

Of course, this stage is far too early to speculate, how these findings could add to the development of novel treatments. Detailed analyses and more appropriate cell models, as 3D organoids, are needed to enable the detailed investigation of underlying cell type and microenvironment-dependent molecular mechanisms and potential interconnections. The restoration of a functional ENS in the affected tissue, by the application of neurogenic compounds, could improve intestinal motility to such an extent that typical symptoms might be alleviated and surgery may no longer be necessary, at least for patients clearly diagnosed with ENCC differentiation defects.

Finally, we assessed DNA methylation in patient FFPE samples. As epigenetic regulation is crucial for embryonic development and increasingly implicated in HSCR (54–56), we performed the first comparative methylation analysis of matched ganglionic and aganglionic FFPE tissues, enabling identification of disease⍰relevant individual epigenetic alterations. In total, 1,347 significantly DMPs could be identified, which have been mapped to the nearest gene based on its TSS, while 1,004 potentially hypomethylated and 120 hypermethylated genes could be identified.

GO term enrichment revealed pathways and diseases relevant mainly for developmental processes and the nervous system, but also for the epithelium, the immune system and for several others. Considering the congenital origin of HSCR and the involvement of NCCs, the enrichment of developmental and neuronal system□related pathways is expected and supports the validity of our approach. Given the breadth of obtained results, we will focus below on a subset of key findings.

Among the top enriched pathways, *Signaling by GPCR* emerged as particularly relevant. G protein-coupled receptors (GPCRs) are central regulators of enteric neurogenic control, affecting motility, barrier function, secretion, and immune signaling (57). We identified a hypomethylated DMP near the HSCR susceptibility gene *EDNRB* TSS (58), potentially enhancing its expression, which contrasts with the reduced EDNRB activity typically caused by HSCR⍰associated variants. Beyond EDNRB, hypomethylation of multiple GPCR genes suggests broader GPCR dysregulation may contribute to HSCR pathogenesis.

ENS development depends on tightly coordinated interactions between ENCCs and the gut microenvironment. Beyond primary ENS defects, HSCR is increasingly linked to intrinsic niche abnormalities that disrupt extracellular matrix (ECM) - cell interactions and stem cell signaling. Consistent with this, the *NABA Core Matrisome pathway* was enriched in our dataset, highlighting structural ECM components as key regulators of ENS development. In the intestine, the ECM provides dynamic mechanical and biochemical cues to regulate epithelial integrity, immune responses, and neuronal-glial interactions (59) as well as to guide ENCC migration, proliferation, and differentiation; disruption of these processes can impair ENCC colonization and could lead to distal aganglionosis (60). Together, our findings support a central role of the core matrisome in ENS patterning and HSCR pathogenesis.

Interestingly, the *Oxytocin Signaling Pathway* was enriched in our dataset. The neuropeptide and hormone oxytocin (OT) regulates central and peripheral functions via signaling through the oxytocin receptor (OTR). Studies suggest that OT/OTR-mediated signaling contributes to multiple GI functions as enteric neuronal activity, motility, intestinal inflammation, macromolecular permeability, mucosal homeostasis, colonic visceral perception and gut microbiota (61–62). The OT/OTR system acts as an anti⍰inflammatory and immunomodulatory pathway, a role underscored by the markedly increased inflammatory susceptibility observed in OTR knockout mice compared with wild⍰type controls (62). So far, OT/OTR signaling was not described in the context of HSCR or HAEC pathophysiology but is of high interest for follow-up studies.

The conducted STRING analysis revealed a small network composed of several molecules including HAND2, several HOX proteins (HOXA7/A13, HOXB5, HOXD13), as well as IKZF1 and CSF2RA. Most of these molecules are involved in developmental processes or in immune system activities. Again, HAND2 is amongst most promising results, as several hypermethylated DMPs near the TSS of *HAND2* could be identified. Hypermethylation of *HAND2* is in concordance with our expression analyses, which revealed a significant reduction of *HAND2* expression in FFPE and *in vitro* samples, likely a consequence of its hypermethylation. We could also underline these results by immunofluorescence staining of ganglionic and aganglionic FFPE specimens. So far, no differential methylation patterns have been described for *HAND2* in the context of HSCR. Further studies are planned to elucidate the mechanisms underlying these observations.

One of the most important molecules in development, are HOX genes. An enteric Hox code, defined by spatially and temporally coordinated Hox gene expression, regulates gut morphogenesis by providing positional cues for migrating cells and supporting differentiation of the enteric neuromusculature and the ENS (63–64). Due to methylation profiling, we could identify and map DMPs of the TOP 20 to TSS of *HOXA7*, *HOXA13*, *HOXB5* and *HOXD13*. Beyond their essential roles in embryonic development, HOXA13 and HOXB5 have also been implicated in HSCR etiology. *HOXA13* is a key regulator of hindgut development and important for ENCC migration during early ENS development (65–66). Reduced expression of the synergistic lncRNA *HOTTIP* and *HOXA13* impairs ENCC migration and may contribute to HSCR (66). HOXB5 is one of the most prominent HOX proteins implicated in ENS development and HSCR pathogenesis, as its conserved expression in the embryonic gut is closely associated with NCC migration and differentiation (63–64,67). During gut development, *HOXB5* is transiently expressed in migrating NCCs, downregulated during neuronal and glial differentiation, and re⍰activated in mature enteric neurons and glia, indicating a regulatory role in NCC development (67). Its involvement in HSCR is well documented: disruption of Hoxb5 in murine models reduces *Ret* expression, delays NCC migration, induces apoptosis of NCCs and leads to HSCR⍰like phenotypes, supporting RET as a downstream target (68–69). HOXB5 directly enhances *RET* transcription via interaction with the multispecies conserved sequence MCS+9.7 and by forming a transcriptional complex with NKX2.1 (70–71). In our cohort, known MCS+9.7 risk variants were absent (71), suggesting an alternative regulatory mechanism. We hypothesize that HOXB5 hypermethylation reduces HOXB5 expression, resulting in impaired RET activation and disturbed NCC development. This is supported by our expression data showing significant downregulation of *RET* and its ligand *GDNF* (72) in aganglionic tissues. However, further functional studies are required to confirm a causal link between *HOXB5* methylation and RET signaling.

Additionally, *CSF2RA* as well as *IKZF1* could be identified, which play important roles in the immune system. *CSF2RA* regulates the survival, functional activation and differentiation of macrophages. There is increasing evidence that the crosstalk between macrophages and the ENS is strongly associated with GI motility and intestinal homeostasis (73). IKZF1 is a zinc-finger transcription factor and associated with others in the regulation of myeloid, erythroid and lymphoid cell lineages as B-and T-cells (74). Both markers represent promising candidates for investigating the underlying pathomechanisms of HAEC.

In conclusion, differential methylation analysis revealed the significant demethylation of several transcription factors, as HAND2 and several HOX genes. To enable the identification of transcription factor binding sites and their target genes as well as to better estimate the impact on disease etiology, CUT&RUN analyses (Cleavage Under Targets & Release Using Nuclease) are planned (75).

To explore how the different biological layers may jointly influence or how a single biological layer is potentially dominating HSCR disease development, we finally performed a multi-omics factor analysis (MOFA). Given the heterogeneity of the datasets, we integrated comparative expression (*Neuropathology Panel*, *Elements Codeset*) and methylation profiling data both derived from FFPE samples. Selected clinical and phenotypic data were used as covariates. Results of methylation and expression profiling explain about 58% (methylation), 50% (*Neuropathology Panel*) and 40% (*Elements Codeset*) of variance between patients. A substantial fraction of the observed variability is non⍰random and reflects shared, structured patterns across multiple molecular layers. The MOFA emphasizes once again that a disturbed methylation pattern can massively contribute to the pathogenesis of HSCR. Identified methylation signatures may enable refined risk stratification in HSCR by serving as biomarkers for patient subgrouping, such as estimating individual susceptibility to enterocolitis or chronic constipation, or discriminating between distinct pathogenic mechanisms (e.g., migration versus differentiation defects). These insights could support more targeted perioperative management, optimized long-term follow-up strategies as well as the development of individualized treatment strategies.

The potential of the applied MOFA has not yet been fully exploited. To draw more detailed conclusions about the correlation between expression and methylation data, it is essential to perform a non-targeted expression profiling approach such as bulk RNAseq.

## Conclusion

The pathogenesis of HSCR is widely recognized as the result of a multifactorial interplay. Multi⍰level modeling frameworks offer a comprehensive strategy by integrating variables across hierarchical biological layers, thereby enabling systematic investigation of their interactions and combined contributions to disease development.

Here, we have presented a multi-level approach encompassing phenotypic, genetic, expression and methylation datasets of a cohort of 33 patients with sporadic HSCR. One highlight of the study was the altered molecular pattern of *HAND2* in aganglionic compared to ganglionic specimens, shown by several analyses. Its relevance to the HSCR pathophysiological network is underlined by differential methylation and expression profiling, resulting in hypermethylated DMPs near the *HAND2* TSS and significantly reduced *HAND2* expression levels in FFPE and *in vitro* samples, likely due to hypermethylation. *HAND2*⍰associated methylation changes have not yet been described in HSCR pathophysiology, highlighting this observation as a novel and potentially important finding. Another key novelty of our study was the use of matched ganglionic and aganglionic FFPE samples, fixed immediately after surgery and thus largely preserving disease-relevant molecular and epigenetic signatures, while also enabling retrospective analyses. This is followed by the use of patient⍰specific *in vitro* models, which complement tissue⍰based studies by enabling individual insights into disease⍰relevant responses.

At present, reliable patient stratification, whether for comorbidity risk, long⍰term outcomes or subgroup definition, is not yet possible. However, we identified potential novel regulatory pathways in HSCR pathogenesis, primarily affecting ENCC differentiation, with integrative analyses suggesting a predominant role for epigenetic regulation. To validate our findings and clearly identify the key molecular drivers within the pathological network of HSCR that may be suitable for personalized therapeutic approaches, multicenter studies are planned.

The described multi-level approach serves as a blueprint for future studies aiming to gain a detailed understanding of individual pathomechanisms, to predict the severity of the disease and possible post-surgery complications and to develop individual treatment strategies.

## Limitations

Several limitations should be acknowledged. First, our cohort of 33 patients, while one of the largest reported for matched ganglionic-aganglionic multi-omic profiling, remains modest for genome-wide methylation analyses. Further our observations require confirmation in an independent cohort. Second, bulk profiling of full-thickness FFPE specimens cannot resolve cell-type composition; the marked difference in neuronal cell content between ganglionic and aganglionic segments may contribute to the observed methylation and expression differences. We attempted to mitigate this through deconvolution analyses and consensus k-means based sample selection but acknowledge that single-cell or spatial profiling approaches will be required to disentangle cell-intrinsic regulation from compositional shifts.

Third, the principal mechanistic claim — that *HAND2* hypermethylation contributes to its transcriptional silencing and to impaired ENCC differentiation - remains correlative. Establishing causality would require orthogonal functional validation approaches, including re-expression of *HAND2* in patient-derived enteric neurospheres induced by a DNA methyltransferase inhibitor such as 5-aza-2′-deoxycytidine, targeted epigenetic perturbation of the *HAND2* promoter using the CRISPR/Cas technology, as well as CUT&RUN-based profiling of *HAND2* downstream targets. Fourth, the targeted nCounter approach restricts transcriptomic read-out to pre-defined gene sets and may miss broader regulatory programs; unbiased bulk and single-nucleus RNA-seq (snPATHO-seq) on the same FFPE specimens will be performed to address this gap. Finally, no independent validation cohort is included; a planned multicenter replication is intended to address this.

## Material & Methods

### Patient recruitment and sampling

Patients suffering from sporadic HSCR and their non-affected parents were recruited at the University Hospitals Heidelberg and Mannheim between 2008 and 2020. The enclosed parents provided written informed consent for genetic and molecular analyses for their affected child and for themselves (S509/2012, 08.02.2013; 2018-662N-MA, 27.12.2018: *Genomweite Sequenzierung bei Patienten mit Morbus Hirschsprung;* S-149/2007, S-208/2014: *Etablierung eines humanen Organoid-Modells des enterischen Nervensystems*, 08.05.2007: *Aufbau einer Gewebebank für molekularbiologische Untersuchungen von operationspflichtigen Erkrankungen im Kindesalter*; 2011-237N-MA, 23.03.2011, 16.10.2015, *Klassifizierung der Dysganglionose des Darmes bei M. Hirschsprung und verwandten Neurochristopathien*).

The diagnosis of HSCR was confirmed in all cases through full wall rectal biopsies taken 2.0, 3.0, and 5.0-6.0 cm above the dentate line under general anesthesia. From 2011 to 2016, HSCR diagnoses were exclusively based on Acetylcholine-Esterase (AChE) histochemistry. From 2017, HSCR diagnoses were performed exclusively using Calretinin immunohistochemistry. The intestinal samples were processed by specialized pediatric pathologists at the Universities of Heidelberg and Mannheim. Transanal endorectal pull-through (TERPT) according to De La Torre procedure was performed as the therapeutic procedure for HSCR in all patients.

From all patients and almost all parents, either EDTA blood or saliva samples (ORAgene saliva tests) were collected for DNA extraction followed by whole exome sequencing (WES). As part of standard clinical practice, tissue samples from each intestinal resection (ganglionic and aganglionic) were immediately transferred to the Institute of Pathology in Heidelberg, where they were formalin⍰fixed and paraffin⍰embedded (FFPE) for detailed histopathological evaluation and long⍰term storage. After routine diagnostics, part of the specimens was used for RNA and DNA extraction followed either by nCounter-based expression profiling or differential methylation analysis. Furthermore, remaining freshly resected tissue specimens (ganglionic and aganglionic; n = 9) obtained from prospectively recruited patients were used to generate patient-derived cell culture models (= *in vitro* samples). An overview of collected samples and conducted analyses is given in Table 1.

**Table 1:**
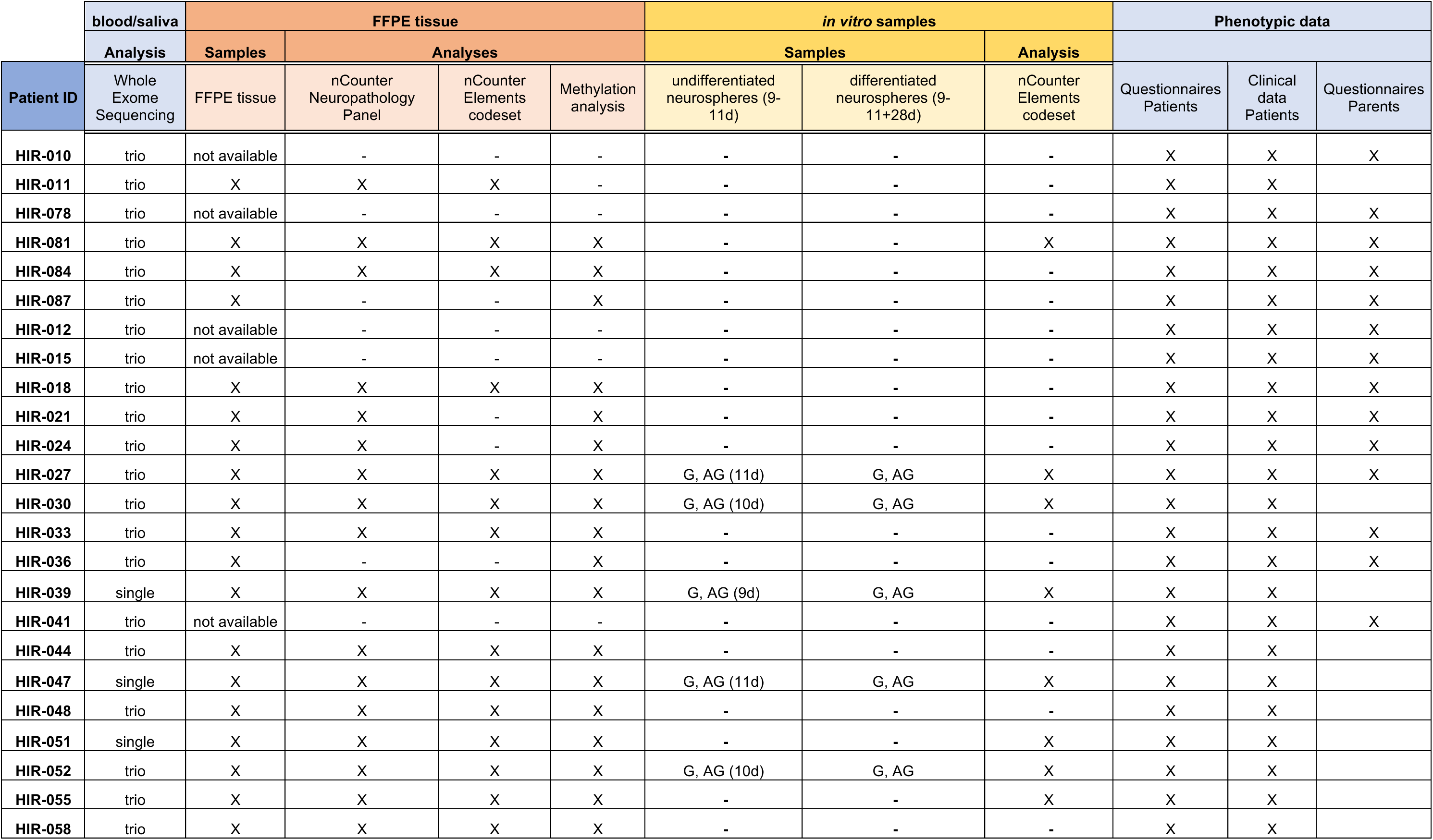
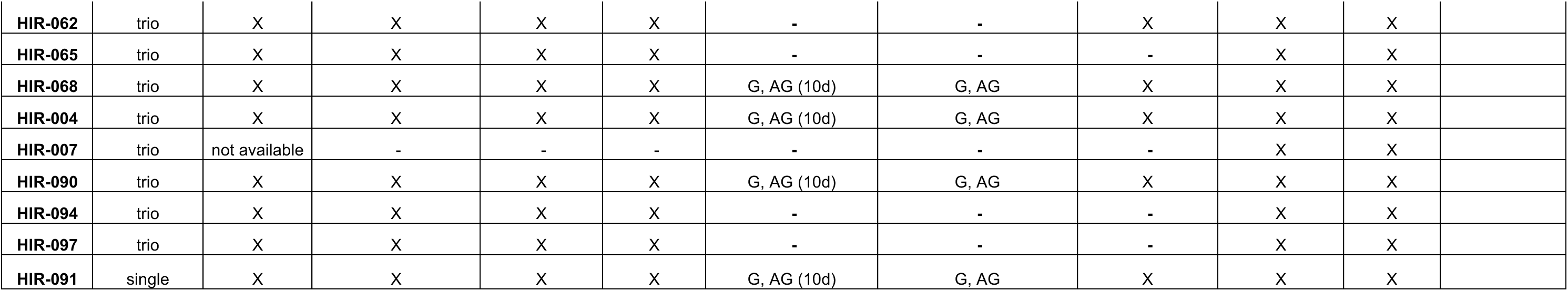
Overview patient samples.

| Patient ID | blood/saliva | FFPE tissue |  |  |  | in vitro samples |  |  | Phenotypic data |  |  |
| --- | --- | --- | --- | --- | --- | --- | --- | --- | --- | --- | --- |
|  | Analysis | Samples | Analyses |  |  | Samples |  | Analysis |  |  |  |
|  | Whole Exome Sequencing | FFPE tissue | nCounter Neuropathology Panel | nCounter Elements codeset | Methylation analysis | undifferentiated neurospheres (9-11d) | differentiated neurospheres (9-11+28d) | nCounter Elements codeset | Questionnaires Patients | Clinical data Patients | Questionnaires Parents |
| HIR-010 | trio | not available | - | - | - | - | - | - | X | X | X |
| HIR-011 | trio | X | X | X | - | - | - | - | X | X |  |
| HIR-078 | trio | not available | - | - | - | - | - | - | X | X | X |
| HIR-081 | trio | X | X | X | X | - | - | X | X | X | X |
| HIR-084 | trio | X | X | X | X | - | - | - | X | X | X |
| HIR-087 | trio | X | - | - | X | - | - | - | X | X | X |
| HIR-012 | trio | not available | - | - | - | - | - | - | X | X | X |
| HIR-015 | trio | not available | - | - | - | - | - | - | X | X | X |
| HIR-018 | trio | X | X | X | X | - | - | - | X | X | X |
| HIR-021 | trio | X | X | - | X | - | - | - | X | X | X |
| HIR-024 | trio | X | X | - | X | - | - | - | X | X | X |
| HIR-027 | trio | X | X | X | X | G, AG (11d) | G, AG | X | X | X | X |
| HIR-030 | trio | X | X | X | X | G, AG (10d) | G, AG | X | X | X |  |
| HIR-033 | trio | X | X | X | X | - | - | - | X | X | X |
| HIR-036 | trio | X | - | - | X | - | - | - | X | X | X |
| HIR-039 | single | X | X | X | X | G, AG (9d) | G, AG | X | X | X |  |
| HIR-041 | trio | not available | - | - | - | - | - | - | X | X | X |
| HIR-044 | trio | X | X | X | X | - | - | - | X | X |  |
| HIR-047 | single | X | X | X | X | G, AG (11d) | G, AG | X | X | X |  |
| HIR-048 | trio | X | X | X | X | - | - | - | X | X |  |
| HIR-051 | single | X | X | X | X | - | - | X | X | X |  |
| HIR-052 | trio | X | X | X | X | G, AG (10d) | G, AG | X | X | X |  |
| HIR-055 | trio | X | X | X | X | - | - | X | X | X |  |
| HIR-058 | trio | X | X | X | X | - | - | - | X | X |  |
| **HIR-062** | trio | X | X | X | X | - | - | X | X | X |  |
| **HIR-065** | trio | X | X | X | X | - | - | - | X | X |  |
| **HIR-068** | trio | X | X | X | X | G, AG (10d) | G, AG | X | X | X |  |
| **HIR-004** | trio | X | X | X | X | G, AG (10d) | G, AG | X | X | X |  |
| **HIR-007** | trio | not available | - | - | - | - | - | - | X | X |  |
| **HIR-090** | trio | X | X | X | X | G, AG (10d) | G, AG | X | X | X |  |
| **HIR-094** | trio | X | X | X | X | - | - | - | X | X |  |
| **HIR-097** | trio | X | X | X | X | - | - | - | X | X |  |
| **HIR-091** | single | X | X | X | X | G, AG (10d) | G, AG | X | X | X |  |

### Phenotyping

In addition to molecular analyses, deep phenotyping was performed of patients and parents. Therefore, a self-designed case form for all probands was used. The Rintala Bowel Function Score (BFS) (76) was assessed from the parents’ reports. It evaluates seven domains that address bowel control, symptoms of soiling, constipation and the social impact of the condition. The score ranges from 0 to 20, with a maximum possible score of 20 (77). The advantage of the Rintala BFS score is that it has been validated in healthy individuals (76,78). The self-designed case form for the patients was completed by the parents and the treating physician at predetermined intervals (1, 5 and 10 years after HSCR surgery). The case form included patient demographics and the presence of associated congenital anomalies. Additionally, age and weight at the time of survey, rectal biopsy and surgery, length of aganglionosis, intestinal resection, and hospitalization period were recorded. Long-segment HSCR (L-HSCR) was defined as an aganglionosis of more than 30 cm. Postoperative complications were also documented, with a follow-up period extending up to 10 years. These complications included among others enterocolitis and constipation. Enterocolitis was defined by the occurrence of at least three out of seven symptoms: diarrhea (stool frequency higher than three times per day), fever, meteorism, vomiting, elevated infectious parameters on blood sampling (CRP/leukocytes), need for hospitalization, and administration of antibiotics. Constipation was rated using the ROME III criteria (79). Stool frequency and stool consistency were conducted at the planned follow-up intervals of 1, 5, and 10 years after the HSCR surgery. Stool consistency was assessed according to the Bristol stool chart (1 = hard lumps, 2 = lumpy sausage, 3 = sausage with surface cracks, 4 = smooth sausage, 5 = soft blobs, 6 = mushy stool, 7 = watery stool) (79). The case form for the parents includes demographic data, congenital diseases, occurrence of more than 3 times enterocolitis in the last year, stool frequency and consistency in the average of the last 3 months, constipation, bowel function, stool continence, nutrition and self-assessment of the general state of health. The same definitions as for patients were used to collect the affiliation data.

Only data from the first year after surgery was included in the current study. Statistical analysis for all variables was performed using SPSS® Version 26.0. The findings were presented as numbers, percentages, means, odds ratio and standard deviations. The two-sided Fisher’s exact test and independent samples *t*-test were employed to assess differences between the groups, with statistical significance set at *p*-value < 0.05.

### Whole Exome Sequencing (WES)

#### DNA extraction

For whole exome sequencing (WES), DNA was extracted from either blood or saliva samples from 33 sporadic HSCR patients and their non-affected parents using the automated QIAsymphony instrument according to the manufacturer’s instructions. In two cases only the patients were sequenced and in one case the patient and the non-affected mother. In all other cases, trio WES was performed from the patient and both non-affected parents. All individuals are of Caucasian ancestry. DNA concentration was determined by the DeNovix spectrophoto- and fluorometer (DeNovix Inc., Wilmington, DE) and the Qubit fluorometer (Thermo Fisher Scientific, Waltham, MA, USA).

#### Data generation

After DNA extraction, WES was conducted on the NovaSeq 6000 Run S4 platform (Illumina) with the IDT xGen Exome Hybridization Panel v2 capture kit at the IKMB in Kiel.

The raw reads from each sample were aligned to genome build GRCh37 (version hs37d5) using *BWA mem* (version 0.7.15) (80) and duplicate reads were marked with *Sambamba* (version 0.6.5) (81). The small variants were called together for all the samples in the family using *Platypus* (version 0.8.1.1) (82). *VCFanno* (83) was employed to incorporate *gnomAD’s* (84) (v2.1) minor allele frequencies (MAF) and local controls’ variant frequencies (VFs) into the raw VCF obtained from *Platypus*. Additionally, variants within a designated list of SNPs and genes associated with HSCR were annotated for subsequent analysis (Table 2). Variants with a MAF > 0.0005 in *gnomAD* or a frequency >0.05 in the local control (3,910 GS and 1,198 WES samples) that were not labeled as variants of interest were excluded from further analysis as common or artifacts. The remaining rare variants were subsequently annotated with *Ensembl VEP* v104 (85), using Gencode v19. Variants were then filtered based on VEP annotations for ‘high’ and ‘moderate’ impact. For trio settings, these variants were prioritized by inheritance models (homozygous, compound homozygous, hemizygous, *de novo*); for single samples, all filtered ‘high’ and ‘moderate’ impact variants were used.

**Table 2:** HSCR-associated SNPs and genes. *according to (91).

| HSCR-associated SNPs |  | HSCR-associated genes* |
| --- | --- | --- |
| Gene | SNP | *DNMT3B* |
| *NRG1* | rs16879552 | *ECE1* |
| *NRG1* | rs16879576 | *EDN3* |
| *RET* | rs61844837 | *EDNRB* |
| *RET* | rs788243 | *GDNF* |
| *RET* | rs788274 | *GFRA1* |
| *RET* | rs788263 | *KIAA1279* |
| *RET* | rs788261 | *L1CAM* |
| *RET* | rs788260 | *NRG1* |
| *RET* | rs3004255 | *NRG3* |
| *RET* | rs2488279 | *NRTN* |
| *RET* | rs1547930 | *NTF3* |
| *RET* | rs4949062 | *NTRK3* |
| *RET* | rs3026693 | *PHOX2B* |
| *RET* | rs7069590 | *PROK1* |
| *RET* | rs2435357 | *PROKR1* |
| *RET* | rs12247456 | *PROKR2* |
| *RET* | rs7393733 | *PSPN* |
| *RET* | rs2506024 | *RET* |
| *RET* | rs78146028 | *SEMA3A* |
| *RET* | rs12764797 | *SEMA3C* |
| *RET* | rs2505515 | *SEMA3D* |
| *RET* | rs1800860 | *SOX10* |
| *RET* | rs11238441 | *TCF4* |
| *RET* | rs2472737 | *ZFHX1B* |
| *RET* | rs2505513 |  |
| *RET* | rs10793423 |  |
| *RET* | rs869184 |  |
| *RET* | rs953999 |  |
| *RET* | rs2506004 |  |
| *RET* | rs7074964 |  |
| *SEMA3* | rs10266471 |  |
| *SEMA3* | rs1228877 |  |
| *SEMA3* | rs1228937 |  |
| *SEMA3* | rs11766001 |  |
| *SEMA3* | rs35092834 |  |
| *SEMA3* | rs12707682 |  |
| *SEMA3* | rs6468008 |  |
| *SEMA3* | rs17560655 |  |

**Table 3:** TOP 10_identified DMPs.

|  | Chr | start | end | strand | nearest_tss | nearest_tss<br>symbol | nearest_tss<br>dist | p-value | q-value | cmp | diff.G.AG. |
| --- | --- | --- | --- | --- | --- | --- | --- | --- | --- | --- | --- |
| **TOP 10 hypermethylated DMPs** |  |  |  |  |  |  |  |  |  |  |  |
| cg11513700 | chr7 | 27205677 | 27205677 | - | ENSG00000106031.9 | HOXA13 | 5585 | 4,31198350847998E-09 | 9,78249331364004E-05 | G < AG | -0,180999664 |
| cg16980686 | chr1 | 75423226 | 75423226 | + | ENSG00000162624.15 | LHX8 | 294793 | 1,25262845380531E-08 | 0,000175038 | G < AG | -0,168128129 |
| cg19788317 | chr4 | 173508149 | 173508149 | + | ENSG00000164105.4 | SAP30 | 138181 | 2,44901044104206E-08 | 0,000246494 | G < AG | -0,197280682 |
| cg00070383 | chr4 | 173508223 | 173508223 | - | ENSG00000164107.9 | HAND2 | 22007 | 4,96605192221719E-08 | 0,00034142 | G < AG | -0,219442196 |
| cg24974365 | chr4 | 173527398 | 173527398 | - | ENSG00000164107.9 | HAND2 | 2832 | 5,03389171241708E-08 | 0,00034142 | G < AG | -0,088942693 |
| cg08195247 | chr2 | 176068558 | 176068558 | + | ENSG00000128714.6 | HOXD13 | 24162 | 4,84225643964017E-08 | 0,00034142 | G < AG | -0,182281677 |
| cg20913646 | chr10 | 126136744 | 126136744 | - | ENSG00000089876.12 | DHX32 | 240307 | 6,93938024352388E-08 | 0,000421289 | G < AG | -0,06195418 |
| cg22778379 | chr17 | 61402200 | 61402200 | + | ENSG00000121068.14 | TBX2 | 2358 | 7,83662045362926E-08 | 0,000447357 | G < AG | -0,121255624 |
| cg20725013 | chr7 | 27155417 | 27155417 | - | ENSG00000122592.8 | HOXA7 | 2520 | 8,77480011231745E-08 | 0,000474595 | G < AG | -0,159218234 |
| cg26491604 | chr17 | 48579502 | 48579502 | + | ENSG00000229637.4 | PRAC2 | 143682 | 9,63085468064795E-08 | 0,000494434 | G < AG | -0,188107976 |
| **TOP 10 hypomethylated DMPs** |  |  |  |  |  |  |  |  |  |  |  |
| cg22890023 | chr17 | 71276380 | 71276380 | + | ENSG00000125398.8 | SOX9 | 844639 | 5,51649394358124E-12 | 2,13470367361062E-06 | G > AG | 0,119383272 |
| cg14615547 | chr11 | 101687015 | 101687015 | - | ENSG00000137672.13 | TRPC6 | 185548 | 5,5813273055916E-12 | 2,13470367361062E-06 | G > AG | 0,171129081 |
| cg26591377 | chrX | 1017215 | 1017215 | + | ENSG00000198223.17 | CSF2RA | 251584 | 2,15380439465851E-11 | 4,5632340549136E-06 | G > AG | 0,10110753 |
| cg04569855 | chr3 | 99955414 | 99955414 | - | ENSG00000168386.19 | FILIP1L | 159100 | 2,3861768869697E-11 | 4,5632340549136E-06 | G > AG | 0,186092311 |
| cg19017635 | chr13 | 95100805 | 95100805 | - | ENSG00000125257.16 | ABCC4 | 200671 | 4,32247415244369E-11 | 5,90768584668151E-06 | G > AG | 0,133373875 |
| cg07638843 | chr14 | 57982303 | 57982303 | - | ENSG00000151812.15 | SLC35F4 | 108 | 4,63381340509489E-11 | 5,90768584668151E-06 | G > AG | 0,13657824 |
| cg08324750 | chr16 | 77272101 | 77272101 | + | ENSG00000205078.6 | SYCE1L | 72694 | 6,0396114476707E-11 | 6,59995631792094E-06 | G > AG | 0,134620031 |
| cg15147534 | chr6 | 48247905 | 48247905 | - | ENSG00000244694.8 | PTCHD4 | 136707 | 1,13292926302579E-10 | 1,08328544775505E-05 | G > AG | 0,09699643 |
| cg17528218 | chr2 | 77519923 | 77519923 | - | ENSG00000176204.14 | LRRTM4 | 73397 | 1,39394575562507E-10 | 1,1847684101815E-05 | G > AG | 0,099421791 |
| cg01295999 | chr3 | 175208544 | 175208544 | + | ENSG00000177694.16 | NAALADL2 | 769972 | 1,95938669817242E-10 | 1,49882268270561E-05 | G > AG | 0,117818355 |

**Table 4:**
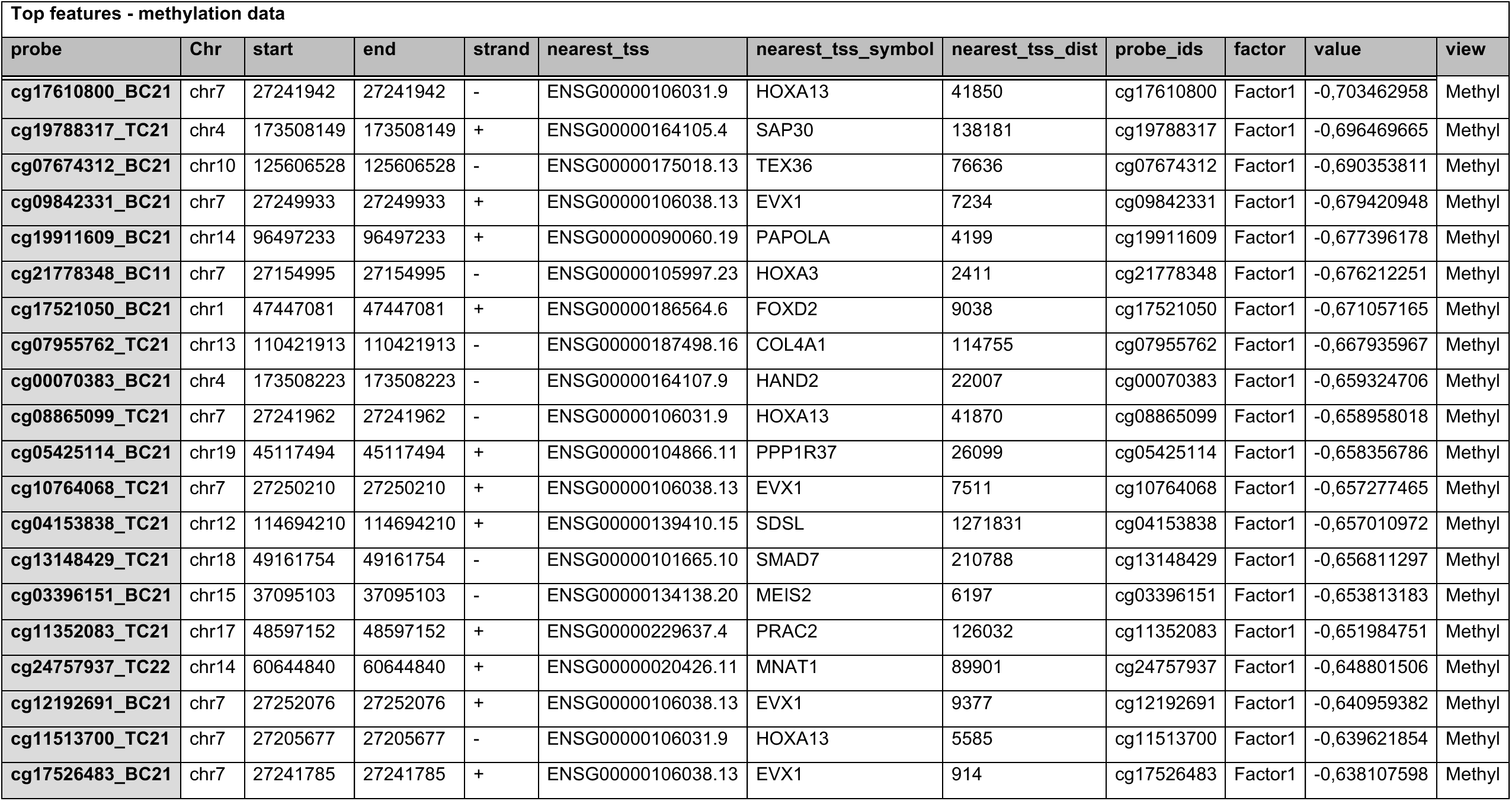
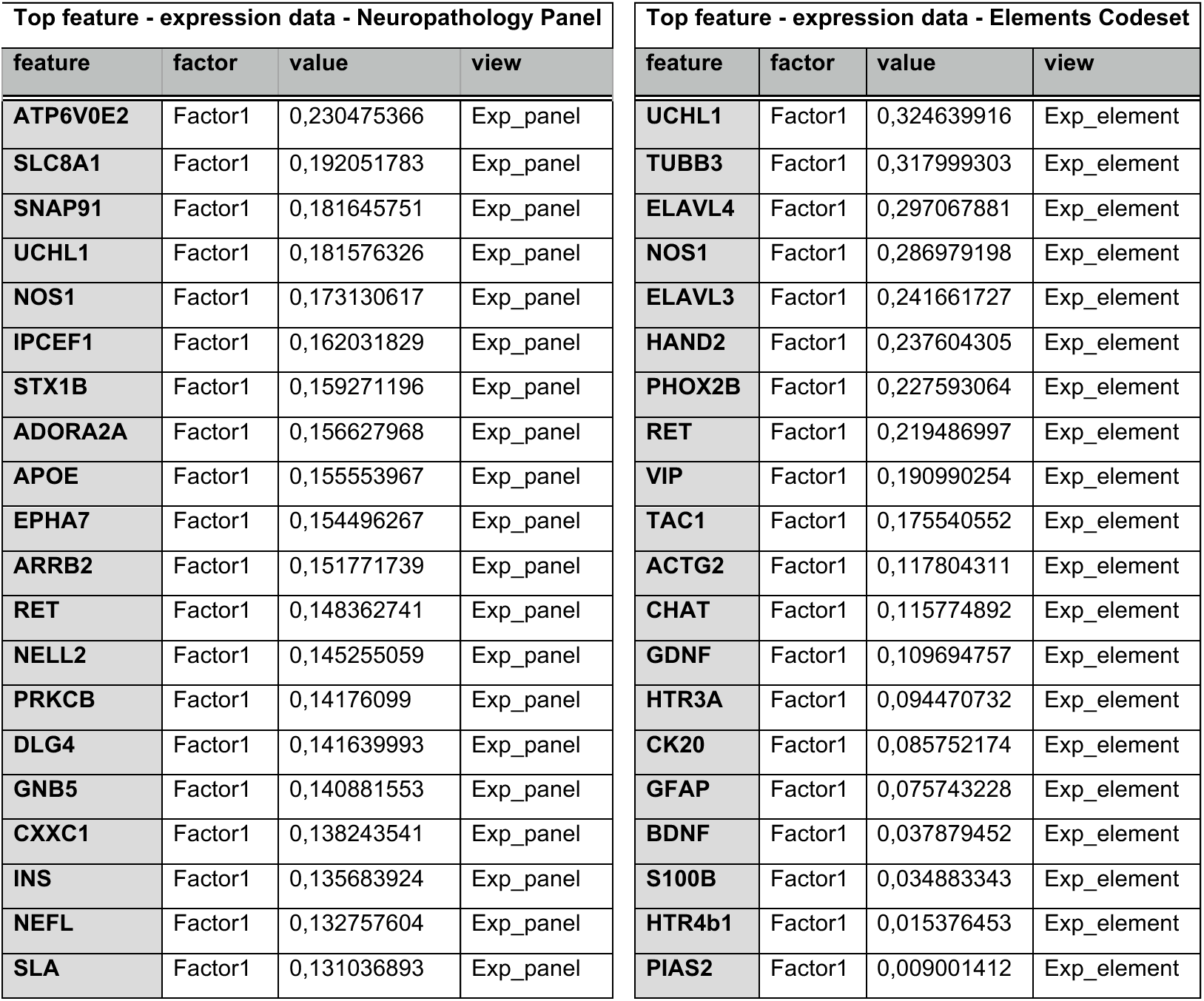
Top feature weights_Factor 1.

### Expression profiling of patient-derived FFPE and *in vitro* samples

#### Total RNA extraction

Expression profiling of patient-specific FFPE and *in vitro* samples was performed at the Expression & Spatial Profiling Core Facility in Heidelberg. Initially, FFPE specimens were sectioned into 10x 10µm sections and total RNA extraction was conducted using the Stratifyer XTRACT FFPE kit (Stratifyer Molecular Pathology GmbH, Cologne, Germany) based on the given instructions of the supplier.

For total RNA extraction from *in vitro* samples, cell pellets were first lysed in 1ml Trizol reagent and further processed using the RNAqueous-Micro kit (both Thermo Fisher Scientific, Waltham, MA, USA) according to the manufacturer’s instructions.

RNA concentration was determined by the DeNovix spectrophoto- and fluorometer (DeNovix Inc., Wilmington, DE) and the Qubit fluorometer (Thermo Fisher Scientific, Waltham, MA, USA). Quality assessment was performed with the Agilent 2100 Bioanalyzer (Agilent Technologies, Inc, Santa Clara, CA).

#### nCounter technology-based analyses

After quality and quantity control measurements, respective RNA samples were analyzed either by the nCounter Digital Analyzer GEN2 (*in vitro* samples) or its successor the nCounter SPRINT profiler (FFPE specimens) (both Bruker). For expression profiling, we applied the prebuild N*europathology Panel* (n = 770 genes; https://ncounter-neuropathology/) and the customized *Elements Codeset* (n = 48 genes; Suppl. Table 1) (both Bruker). FFPE samples were analyzed with both setups, while *in vitro* samples were only analyzed with the customized *Elements Codeset*. Normalization was carried out using the freely available nSolver Analysis Software (version 4.0, https://ncounter-analysis-solutions/) considering internal positive controls and the stably expressed reference genes. Stably expressed reference genes were chosen for normalization based on the geNorm method (v0.953, MS Excel Add-in, (86)). For determination of the background and detection limit, the threshold was set as the geometric mean of negative controls (NEG_A-F, included in the assay by default) + 2*standard deviation. Genes with numbers below this threshold were considered as background, while those above were regarded as specific signals. A gene was considered present in a group (ganglionic, aganglionic) if it surpassed this threshold in at least 50% of the group’s samples. Genes, not detected in all groups, were excluded from further analysis. Due to a low performance, FFPE sample sets of two patients need to be excluded from the *Elements Codeset*-based analysis (n = 23 patients).

Statistical analyses and visualizations were performed using the Prism software (version 9.3.1; GraphPad Software). Differential expression analysis was performed using the unpaired, nonparametric *Mann-Whitney U test*. Raw *p*-values were corrected by the *q*-value method (87) to control false positives in multiple hypothesis testing. *q*-values ≤ 0.05 (*), ≤ 0.01 (**) and ≤ 0.001 (***) were considered statistically significant.

#### Immunofluorescence staining

Comparative immunofluorescence staining of patients’ FFPE specimens for HAND2 is described in the Supplementary Methods section.

### Cell culture experiments

#### Undifferentiated enteric neurospheres

Freshly resected ganglionic and aganglionic gut specimens from nine prospective enrolled patients, were used for the generation of enteric neurospheres according to Schmitteckert et al. (22). In brief, ganglionic and aganglionic gut specimens were placed immediately after resection into ice⍰cold HBSS with penicillin/streptomycin (P/S; 10.000U ml^-1^, both Thermo Fisher Scientific). In the lab, tissues were kept in ice⍰cold HBSS+P/S, the mucosa and submucosa were removed, and samples were mechanically minced and enzymatically dissociated at 37□°C with Collagenase/Dispase (Clostridiopeptidase A, 800U mg^-1^; Sigma-Aldrich /Dispase II, 0.8U mg^-1^; Roche). After centrifugation, cells were resuspended in supplemented culture medium (Neurobasal A medium, GlutaMAX (100x), P/S (10.000U ml^-1^), N2 (100x), B27 minus vitamin A (50x), EGF; 20 ng ml^-1^, bFGF; 20 ng ml^-1^ (all Thermo Fisher Scientific), Amphoterizin B (250 µg ml^-1^), Metronidazol (50µg ml^-1^) (both Sigma-Aldrich) and seeded onto poly-L-Ornithin (0.01 %)/Laminin (2µg/cm^2^)/Fibronectin (2µg/cm^2^) (all from Sigma-Aldrich)-coated well plates. Medium was half⍰changed every 3–5□days, and EGF/bFGF were added daily.

#### Differentiated cell models

On day 10 of cultivation, a small portion of the formed ganglionic and aganglionic tissue-derived enteric neurospheres (= undifferentiated enteric neurospheres) was harvested for total RNA extraction and subsequent expression profiling. The remaining ones were dissociated with StemPro Accutase (Thermo Fisher Scientific) for 20 min at 37°C, gently triturated, centrifuged and resuspended in differentiation medium (Knockout DMEM/F-12, StemPro neural supplement (50x), GlutaMAX (100x) (all Thermo Fisher Scientific) and Tegaserod maleate (10nM) (Tocris Bioscience), which is inspired by the cultivation medium of H9 human neural stem cells (Thermo Fisher Scientific). The cell suspensions were respectively transferred onto Matrigel (growth factor-reduced Matrigel Matrix, Corning, Corning, NY, USA)-coated coverslips in 24-well plates and cultured for further 28 days. Two times a week a half medium change was performed.

Undifferentiated enteric neurospheres (10d) and differentiated cells (10+28d) from both tissue entities were comparatively assessed by bright field microscopy, nCounter expression profiling and/or immunofluorescence staining.

### Differential methylation profiling

#### Sample processing

For DNA methylation analysis, each FFPE block was sectioned in 10x 10µm sections and DNA was extracted using the Maxwell RSC FFPE DNA Purification Kit (Promega). Thereafter the Infinium HD FFPE DNA Restore Kit (Illumina) was applied to restore degraded FFPE DNA to an amplifiable state. Both kits were used in accordance with the company’s requirements. DNA samples were then analyzed using the EZ DNA Methylation Kit for bisulfite conversion of the DNA and the Infinium Methylation EPIC v2.0 BeadChip Kit (Illumina). All steps were conducted at the Department of Neuropathology in Heidelberg.

#### Data normalization

The methylation array was processed with the *minfi* package and the *IlluminaHumanMethylationEPICv2anno.20a1.hg38* annotation packages (88–89). Raw methylation signals were normalized using quantile normalization. Probes that are associated with SNPs were removed.

#### Identification of DMPs

The normalized beta values were initially uploaded into the *‘dmpFinder’* package (89). For this analysis, “AG” (aganglionic) and “G” (ganglionic) annotations were employed as categorical phenotypes to ascertain potential associations. A subset of 38 samples selected by consensus *k*-means clustering (90) that contribute to stable sample classifications were used for DMP detection. Probes satisfying *q*-value < 0.01were designated as DMPs. For gene ontology, pathway and disease enrichment analysis, DMPs were mapped to nearest transcription sites of protein-coding genes. Analyses were conducted using the gene annotation and analysis resource *Metascape* (https://metascape.org/) (23).

### Multi-Omics Factor analysis (MOFA)

A MOFA object was constructed utilizing the differentially methylated positions (DMPs), and the genes demonstrated significant differential expression based on the nCounter expression data from FFPE specimens (ready-to-use *Neuropathology Panel,* customized *Elements Codeset*). For the training parameters, the convergence mode was configured to “slow”, and the maximum number of iterations was set to 10,000. The total number of factors was specified as 5. With these defined training and model options, in conjunction with the patient data, the MOFA object was prepared, and the analysis was executed. To visually represent the results, the *correlate_factors_with_covariates* function was employed to generate the plot shown in Supplementary Figure 4, and the *plot_factor* function was utilized to create factor-specific scatter plots shown in Figure 8.

## Acknowledgements

We thank the patients and their families for their kind support and participation in our study. We further thank all involved surgeons and technicians for their valuable contribution to this study. We acknowledge the kind support of Matias Simons for proof-reading. We also thank Vladimir Benes and Laura Villacorta from the Genomics Core Facility at the EMBL (Heidelberg).

## Author contributions

SM, BN and PR designed the study. RS, MB, LW, LC, PG and PR recruited patients and parents and collected phenotypic data. RS, MB, PG, PR conducted surgeries. SR, FL, CS did the pathological examination of resected tissues. JS und MATL prepared samples for further analyses. SM performed the experiments. TM assisted in experimentation. HK and RR performed nCounter expression analyses, which SM and RR interpreted. BSL and AF performed whole exome sequencing analyses. FS and SH performed methylation profiling. NP, ZG, KN and DH performed bioinformatic analyses and helped with interpretation of findings. SM contributed to the data analysis, interpreted the data and wrote the manuscript, with editorial input from GR, CS and PR. All authors read, edited and approved the final manuscript.

## Funder information declared

This study was supported by the Heidelberg Stiftung Chirurgie (https://www.stiftung-chirurgie.de) (S. Mellein, B. Niesler, P. Romero), the Heinz and Heide Dürr Stiftung (https://www.heinzundheideduerrstiftung.de) (B. Niesler and P. Romero; 2017/2.2.1/04), the BILD hilft e. V. „Ein Herz für Kinder“ (B. Niesler and P. Romero) and the Ministry of Science, Research and Arts (MWK33-7532-55/8/9) (S. Mellein, B. Niesler). Additional support was provided by the Institute of Human Genetics (G. Rappold, C. Schaaf), Heidelberg University Hospital (https://www.heidelberg-university-hospital.com).

## Data availability

The data generated in this study are available via controlled access in the German Human Genome-Phenome Archive (GHGA, <u>data.ghga.de</u>) under the GHGA Accession https://data.ghga.de/study/GHGAS01342870485305. Further details, including the data access policy for the study, can be found there.

## Ethics approval statement

The Ethics Committees of the Medical Faculty Heidelberg and Mannheim gave ethical approval for this work. Details are given in the manuscript.

## References

1. Amiel J, Lyonnet S. Hirschsprung disease, associated syndromes, and genetics: a review. J Med Genet. Nov 2001;38(11):729–39. doi:10.1136/jmg.38.11.729

2. Heuckeroth RO. Hirschsprung disease - integrating basic science and clinical medicine to improve outcomes. Nat Rev Gastroenterol Hepatol. Mar 2018;15(3):152–167. doi:10.1038/nrgastro.2017.149

3. Brooks AS, et al. Studying the genetics of Hirschsprung’s disease: unraveling an oligogenic disorder. Clin Genet. Jan 2005;67(1):6–14. doi:10.1111/j.1399-0004.2004.00319.x

4. Fernandez RM, et al. Pathways systematically associated to Hirschsprung’s disease. Orphanet J Rare Dis. Dec 2 2013;8:187. doi:10.1186/1750-1172-8-187

5. Heanue TA, Pachnis V. Enteric nervous system development and Hirschsprung’s disease: advances in genetic and stem cell studies. Nat Rev Neurosci. Jun 2007;8(6):466–79. doi:10.1038/nrn2137

6. Luzon-Toro B, et al. What is new about the genetic background of Hirschsprung disease? Clin Genet. Jan 2020;97(1):114–124. doi:10.1111/cge.13615

7. Tilghman JM, et al. Molecular Genetic Anatomy and Risk Profile of Hirschsprung’s Disease. N Engl J Med. Apr 11 2019;380(15):1421–1432. doi:10.1056/NEJMoa1706594

8. Luo Y, et al. Close linkage with the RET protooncogene and boundaries of deletion mutations in autosomal dominant Hirschsprung disease. Hum Mol Genet. Nov 1993;2(11):1803–8. doi:10.1093/hmg/2.11.1803

9. Amiel J, et al. Hirschsprung disease, associated syndromes and genetics: a review. J Med Genet. Jan 2008;45(1):1–14. doi:10.1136/jmg.2007.053959

10. Serra A, et al. Analysis of RET, ZEB2, EDN3 and GDNF genomic rearrangements in central congenital hyperventilation syndrome patients by multiplex ligation-dependent probe amplification. Ann Hum Genet. Jul 2010;74(4):369–74. doi:10.1111/j.1469-1809.2010.00577.x

11. Theocharatos S, Kenny SE. Hirschsprung’s disease: current management and prospects for transplantation of enteric nervous system progenitor cells. Early Hum Dev. Dec 2008;84(12):801–4. doi:10.1016/j.earlhumdev.2008.09.007

12. Wallace AS, Anderson RB. Genetic interactions and modifier genes in Hirschsprung’s disease. World J Gastroenterol. Dec 7 2011;17(45):4937–44. doi:10.3748/wjg.v17.i45.4937

13. Chen Y, et al. Transanal endorectal pull-through versus transabdominal approach for Hirschsprung’s disease: a systematic review and meta-analysis. J Pediatr Surg. Mar 2013;48(3):642–51. doi:10.1016/j.jpedsurg.2012.12.036

14. De La Torre L, Langer JC. Transanal endorectal pull-through for Hirschsprung disease: technique, controversies, pearls, pitfalls, and an organized approach to the management of postoperative obstructive symptoms. Semin Pediatr Surg. May 2010;19(2):96–106. doi:10.1053/j.sempedsurg.2009.11.016

15. Romero P, et al. Outcome of transanal endorectal vs. transabdominal pull-through in patients with Hirschsprung’s disease. Langenbecks Arch Surg. Oct 2011;396(7):1027–33. doi:10.1007/s00423-011-0804-9

16. Ohkura T, et al. Updates and Challenges in ENS Cell Therapy for the Treatment of Neurointestinal Diseases. Biomolecules. Feb 16 2024;14(2)doi:10.3390/biom14020229

17. Diposarosa R, et al. Literature review: enteric nervous system development, genetic and epigenetic regulation in the etiology of Hirschsprung’s disease. Heliyon. Jun 2021;7(6):e07308. doi:10.1016/j.heliyon.2021.e07308

18. Heuckeroth RO, Schafer KH. Gene-environment interactions and the enteric nervous system: Neural plasticity and Hirschsprung disease prevention. Dev Biol. Sep 15 2016;417(2):188–97. doi:10.1016/j.ydbio.2016.03.017

19. Cheng J, et al. Accurate proteome-wide missense variant effect prediction with AlphaMissense. Science. Sep 22 2023;381(6664):eadg7492. doi:10.1126/science.adg7492

20. Kapoor A, et al. Multiple, independent, common variants at RET, SEMA3 and NRG1 gut enhancers specify Hirschsprung disease risk in European ancestry subjects. J Pediatr Surg. Dec 2021;56(12):2286–2294. doi:10.1016/j.jpedsurg.2021.04.010

21. Mederer T, et al. A complementary study approach unravels novel players in the pathoetiology of Hirschsprung disease. PLoS Genet. Nov 2020;16(11):e1009106. doi:10.1371/journal.pgen.1009106

22. Schmitteckert S, et al. Postnatal human enteric neurospheres show a remarkable molecular complexity. Neurogastroenterol Motil. Oct 2019;31(10):e13674. doi:10.1111/nmo.13674

23. Zhou Y, et al. Metascape provides a biologist-oriented resource for the analysis of systems-level datasets. Nat Commun. Apr 3 2019;10(1):1523. doi:10.1038/s41467-019-09234-6

24. Szklarczyk D, et al. The STRING database in 2023: protein-protein association networks and functional enrichment analyses for any sequenced genome of interest. Nucleic Acids Res. Jan 6 2023;51(D1):D638–D646. doi:10.1093/nar/gkac1000

25. Sergi CM, Hager J. Editorial: Hirschsprung disease: genetic susceptibility, disease mechanisms and innovative management in the multi-omics era. Front Pediatr. 2023;11:1274735. doi:10.3389/fped.2023.1274735

26. Lucena-Padros H, et al. Bioinformatics Prediction for Network-Based Integrative Multi-Omics Expression Data Analysis in Hirschsprung Disease. Biomolecules. Jan 30 2024;14(2)doi:10.3390/biom14020164

27. Tang W, et al. Multiple ’omics’-analysis reveals the role of prostaglandin E2 in Hirschsprung’s disease. Free Radic Biol Med. Feb 20 2021;164:390–398. doi:10.1016/j.freeradbiomed.2020.12.456

28. Sobala LF, et al. Structure of human endo-alpha-1,2-mannosidase (MANEA), an antiviral host-glycosylation target. Proc Natl Acad Sci U S A. Nov 24 2020;117(47):29595–29601. doi:10.1073/pnas.2013620117

29. Wen Q, et al. MDN1 variants cause susceptibility to epilepsy : For the China Epilepsy Gene 1.0 Project. Acta Epileptol. Mar 3 2025;7(1):17. doi:10.1186/s42494-025-00209-3

30. D’Autreaux F, et al. Hand2 is necessary for terminal differentiation of enteric neurons from crest-derived precursors but not for their migration into the gut or for formation of glia. Development. Jun 2007;134(12):2237–49. doi:10.1242/dev.003814

31. Lei J, Howard MJ. Targeted deletion of Hand2 in enteric neural precursor cells affects its functions in neurogenesis, neurotransmitter specification and gangliogenesis, causing functional aganglionosis. Development. Nov 2011;138(21):4789–800. doi:10.1242/dev.060053

32. Qin KW, et al. The research on screening differentially expressed genes in Hirschsprung’s disease by using Microarray. J Pediatr Surg. Nov 2013;48(11):2281–8. doi:10.1016/j.jpedsurg.2013.06.024

33. Xiao SJ, et al. Gene expression profiling coupled with Connectivity Map database mining reveals potential therapeutic drugs for Hirschsprung disease. J Pediatr Surg. Sep 2018;53(9):1716–1721. doi:10.1016/j.jpedsurg.2018.02.060

34. Hao MM, Young HM. Development of enteric neuron diversity. J Cell Mol Med. Jul 2009;13(7):1193–210. doi:10.1111/j.1582-4934.2009.00813.x

35. Cairns BR, et al. Automated computational analysis reveals structural changes in the enteric nervous system of nNOS deficient mice. Sci Rep. Aug 25 2021;11(1):17189. doi:10.1038/s41598-021-96677-x

36. Kusafuka T, Puri P. Altered mRNA expression of the neuronal nitric oxide synthase gene in Hirschsprung’s disease. J Pediatr Surg. Jul 1997;32(7):1054–8. doi:10.1016/s0022-3468(97)90398-5

37. Vanderwinden JM, et al. Nitric oxide synthase distribution in the enteric nervous system of Hirschsprung’s disease. Gastroenterology. Oct 1993;105(4):969–73. doi:10.1016/0016-5085(93)90938-9

38. Freund HR, et al. Reduced tissue content of vasoactive intestinal peptide in aganglionic colon of Hirschsprung’s disease. Am J Surg. Feb 1981;141(2):243–4. doi:10.1016/0002-9610(81)90167-7

39. Walters JR, et al. Calretinin and calbindin-D28k immunoreactivity in the human gastrointestinal tract. Gastroenterology. May 1993;104(5):1381–9. doi:10.1016/0016-5085(93)90346-e

40. Claxton HL, et al. The Diagnostic Value of Immunohistochemistry Markers in Hirschsprung Disease; A Systematic Review and Meta-analysis. J Pediatr Surg. Feb 2025;60(2):162010. doi:10.1016/j.jpedsurg.2024.162010

41. Engelstoft MS, et al. Research Resource: A Chromogranin A Reporter for Serotonin and Histamine Secreting Enteroendocrine Cells. Mol Endocrinol. Nov 2015;29(11):1658–71. doi:10.1210/me.2015-1106

42. Nikolić IT VP, V.; Petrović, A.; Mitić, D. Dynamics Of Differentiation Of Chromogranin A-Immunoreactive Endocrine Cells And Myenteric Plexus Of The Human Fetal Duodenum In The Third And The Fifth Month Of Development. Scientific Journal of the Faculty of Medicine in Niš. 2010:19–25.

43. Shen Z, et al. Immunocytochemical evidence for chromogranin A and B in neuronal elements in human gut. Regul Pept. May 26 1994;51(3):229–36. doi:10.1016/0167-0115(94)90069-8

44. Wei L, et al. Enterochromaffin Cells-Gut Microbiota Crosstalk: Underpinning the Symptoms, Pathogenesis, and Pharmacotherapy in Disorders of Gut-Brain Interaction. J Neurogastroenterol Motil. Jul 30 2022;28(3):357–375. doi:10.5056/jnm22008

45. Soeda J, et al. Mucosal neuroendocrine cell abnormalities in the colon of patients with Hirschsprung’s disease. J Pediatr Surg. Jul 1992;27(7):823–7. doi:10.1016/0022-3468(92)90374-g

46. Huang H, et al. MYRF: A Mysterious Membrane-Bound Transcription Factor Involved in Myelin Development and Human Diseases. Neurosci Bull. Jun 2021;37(6):881–884. doi:10.1007/s12264-021-00678-9

47. Baldarelli RM, et al. The mouse Gene Expression Database (GXD): 2021 update. Nucleic Acids Res. Jan 8 2021;49(D1):D924–D931. doi:10.1093/nar/gkaa914

48. Rodriguez-Fraticelli AE, et al. Developmental regulation of apical endocytosis controls epithelial patterning in vertebrate tubular organs. Nat Cell Biol. Mar 2015;17(3):241–50. doi:10.1038/ncb3106

49. Rao M, et al. Enteric glia express proteolipid protein 1 and are a transcriptionally unique population of glia in the mammalian nervous system. Glia. Nov 2015;63(11):2040–2057. doi:10.1002/glia.22876

50. Li Q, et al. Gut microbiota and myelination: Crosstalk across the lifespan and microbiota-based modulation strategies. Microbiol Res. Nov 2025;300:128286. doi:10.1016/j.micres.2025.128286

51. Duci M, et al. Postoperative Hirschsprung’s associated enterocolitis (HAEC): transition zone as putative histopathological predictive factor. J Clin Pathol. Jan 17 2025;78(2):111–116. doi:10.1136/jcp-2023-209129

52. Wang T, et al. snPATHO-seq, a versatile FFPE single-nucleus RNA sequencing method to unlock pathology archives. Commun Biol. Oct 16 2024;7(1):1340. doi:10.1038/s42003-024-07043-2

53. Tarapcsak S, et al. Single-cell RNA sequencing in Hirschsprung’s disease tissues reveals lack of neuronal differentiation in the aganglionic colon segment. bioRxiv. Jul 4 2025;doi:10.1101/2025.07.01.662516

54. Torroglosa A, et al. Epigenetics in ENS development and Hirschsprung disease. Dev Biol. Sep 15 2016;417(2):209–16. doi:10.1016/j.ydbio.2016.06.017

55. Torroglosa A, et al. Epigenetic Mechanisms in Hirschsprung Disease. Int J Mol Sci. Jun 26 2019;20(13)doi:10.3390/ijms20133123

56. Villalba-Benito L, et al. Genome-wide analysis of DNA methylation in Hirschsprung enteric precursor cells: unraveling the epigenetic landscape of enteric nervous system development. Clin Epigenetics. Mar 9 2021;13(1):51. doi:10.1186/s13148-021-01040-6

57. Carbone SE, et al. G protein-coupled receptor trafficking and signaling: new insights into the enteric nervous system. Am J Physiol Gastrointest Liver Physiol. Apr 1 2019;316(4):G446–G452. doi:10.1152/ajpgi.00406.2018

58. Puffenberger EG, et al. A missense mutation of the endothelin-B receptor gene in multigenic Hirschsprung’s disease. Cell. Dec 30 1994;79(7):1257–66. doi:10.1016/0092-8674(94)90016-7

59. Naba A, et al. The matrisome: in silico definition and in vivo characterization by proteomics of normal and tumor extracellular matrices. Mol Cell Proteomics. Apr 2012;11(4):M111 014647. doi:10.1074/mcp.M111.014647

60. Ji Y, et al. Roles of Enteric Neural Stem Cell Niche and Enteric Nervous System Development in Hirschsprung Disease. Int J Mol Sci. Sep 7 2021;22(18)doi:10.3390/ijms22189659

61. Liu H, et al. Oxytocin/Oxytocin Receptor Signalling in the Gastrointestinal System: Mechanisms and Therapeutic Potential. Int J Mol Sci. Oct 11 2024;25(20)doi:10.3390/ijms252010935

62. Welch MG, et al. Oxytocin regulates gastrointestinal motility, inflammation, macromolecular permeability, and mucosal maintenance in mice. Am J Physiol Gastrointest Liver Physiol. Oct 15 2014;307(8):G848–62. doi:10.1152/ajpgi.00176.2014

63. Krumlauf R. Hox genes in vertebrate development. Cell. Jul 29 1994;78(2):191–201. doi:10.1016/0092-8674(94)90290-9

64. Pitera JE, et al. Coordinated expression of 3’ hox genes during murine embryonal gut development: an enteric Hox code. Gastroenterology. Dec 1999;117(6):1339–51. doi:10.1016/s0016-5085(99)70284-2

65. Doodnath R, et al. The spatio-temporal patterning of Hoxa9 and Hoxa13 in the developing zebrafish enteric nervous system. Pediatr Surg Int. Feb 2012;28(2):115–21. doi:10.1007/s00383-011-2992-3

66. Xie H, et al. Long none coding RNA HOTTIP/HOXA13 act as synergistic role by decreasing cell migration and proliferation in Hirschsprung disease. Biochem Biophys Res Commun. Aug 7 2015;463(4):569–74. doi:10.1016/j.bbrc.2015.05.096

67. Fu M, et al. HOXB5 expression is spatially and temporarily regulated in human embryonic gut during neural crest cell colonization and differentiation of enteric neuroblasts. Dev Dyn. Sep 2003;228(1):1–10. doi:10.1002/dvdy.10350

68. Kam MK, et al. Perturbation of Hoxb5 signaling in vagal and trunk neural crest cells causes apoptosis and neurocristopathies in mice. Cell Death Differ. Feb 2014;21(2):278–89. doi:10.1038/cdd.2013.142

69. Lui VC, et al. Perturbation of hoxb5 signaling in vagal neural crests down-regulates ret leading to intestinal hypoganglionosis in mice. Gastroenterology. Apr 2008;134(4):1104–15. doi:10.1053/j.gastro.2008.01.028

70. Zhu J, et al. HOXB5 cooperates with NKX2-1 in the transcription of human RET. PLoS One. 2011;6(6):e20815. doi:10.1371/journal.pone.0020815

71. Zhu JJ, et al. HOXB5 binds to multi-species conserved sequence (MCS+9.7) of RET gene and regulates RET expression. Int J Biochem Cell Biol. Jun 2014;51:142–9. doi:10.1016/j.biocel.2014.04.013

72. Uesaka T, et al. GDNF signaling levels control migration and neuronal differentiation of enteric ganglion precursors. J Neurosci. Oct 9 2013;33(41):16372–82. doi:10.1523/JNEUROSCI.2079-13.2013

73. Hamilton JA. GM-CSF-Dependent Inflammatory Pathways. Front Immunol. 2019;10:2055. doi:10.3389/fimmu.2019.02055

74. Feng L, et al. Multifaceted roles of IKZF1 gene, perspectives from bench to bedside. Front Oncol. 2024;14:1383419. doi:10.3389/fonc.2024.1383419

75. Skene PJ, Henikoff S. An efficient targeted nuclease strategy for high-resolution mapping of DNA binding sites. Elife. Jan 16 2017;6doi:10.7554/eLife.21856

76. Rintala RJ, Lindahl H. Is normal bowel function possible after repair of intermediate and high anorectal malformations? J Pediatr Surg. Mar 1995;30(3):491–4. doi:10.1016/0022-3468(95)90064-0

77. Thakkar HS, et al. Functional outcomes in Hirschsprung disease: A single institution’s 12-year experience. J Pediatr Surg. Feb 2017;52(2):277–280. doi:10.1016/j.jpedsurg.2016.11.023

78. Kyrklund K, et al. Evaluation of bowel function and fecal continence in 594 Finnish individuals aged 4 to 26 years. Dis Colon Rectum. Jun 2012;55(6):671–6. doi:10.1097/DCR.0b013e31824c77e4

79. Chumpitazi BP, et al. Bristol Stool Form Scale reliability and agreement decreases when determining Rome III stool form designations. Neurogastroenterol Motil. Mar 2016;28(3):443–8. doi:10.1111/nmo.12738

80. Li H. Aligning sequence reads, clone sequences and assembly contigs with BWA-MEM. arXiv preprint 2013;arXiv:1303.3997

81. Tarasov A, et al. Sambamba: fast processing of NGS alignment formats. Bioinformatics. Jun 15 2015;31(12):2032–4. doi:10.1093/bioinformatics/btv098

82. Rimmer A, et al. Integrating mapping-, assembly- and haplotype-based approaches for calling variants in clinical sequencing applications. Nat Genet. Aug 2014;46(8):912–918. doi:10.1038/ng.3036

83. Pedersen BS, et al. Vcfanno: fast, flexible annotation of genetic variants. Genome Biol. Jun 1 2016;17(1):118. doi:10.1186/s13059-016-0973-5

84. Karczewski KJ, et al. The mutational constraint spectrum quantified from variation in 141,456 humans. Nature. May 2020;581(7809):434–443. doi:10.1038/s41586-020-2308-7

85. McLaren W, et al. The Ensembl Variant Effect Predictor. Genome Biol. Jun 6 2016;17(1):122. doi:10.1186/s13059-016-0974-4

86. Andersen CL, et al. Normalization of real-time quantitative reverse transcription-PCR data: a model-based variance estimation approach to identify genes suited for normalization, applied to bladder and colon cancer data sets. Cancer Res. Aug 1 2004;64(15):5245–50. doi:10.1158/0008-5472.CAN-04-0496

87. Storey JD, Tibshirani R. Statistical significance for genomewide studies. Proc Natl Acad Sci U S A. Aug 5 2003;100(16):9440–5. doi:10.1073/pnas.1530509100

88. Aryee MJ, et al. Minfi: a flexible and comprehensive Bioconductor package for the analysis of Infinium DNA methylation microarrays. Bioinformatics. May 15 2014;30(10):1363–9. doi:10.1093/bioinformatics/btu049

89. Fortin JP, et al. Preprocessing, normalization and integration of the Illumina HumanMethylationEPIC array with minfi. Bioinformatics. Feb 15 2017;33(4):558–560. doi:10.1093/bioinformatics/btw691

90. Gu Z, et al. cola: an R/Bioconductor package for consensus partitioning through a general framework. Nucleic Acids Res. Feb 22 2021;49(3):e15. doi:10.1093/nar/gkaa1146

