## Supplementary for "An integrative multi-omics framework identifies epigenetic dysregulation of *HAND2* as a potential primary driver of impaired enteric neural crest cell differentiation in Hirschsprung Disease"

**Supplementary Methods**

**Immunofluorescence staining**

Differentiated enteric neurospheres (ganglionic, aganglionic) were harvested for immunofluorescence staining after 28 days of differentiation (10+28d). The staining procedure was performed according to Muthuswamy et al. [1] with slight changes. In brief, after aspiration of media, adherent cells were fixed in 5% Formalin/1x PBS for 30 min at room temperature (RT). Fixed samples were then washed three times with 1x PBS/Glycine (10x PBS/Glycine 500ml: 38 g NaCl, 9.38 g Na_2_HPO_4_, 2.07 g NaH_2_PO_4_, 37.5 g Glycine, pH 7.4, filter sterilize) (all from Sigma Aldrich, St. Louis, MO, USA). Thereafter, samples were permeabilized using 0.5% Triton X-100 in 1x PBS for 5 min at RT. After three additional washing steps with 1x IF wash (10x IF wash 500ml: 38g NaCl, 9.38g Na_2_HPO_4_, 2.07g NaH_2_PO_4_, 2.5g NaN_3_, 5g BSA (Fraction V), 10 ml Triton X-100, 2.5 ml Tween-20, pH 7.4, filter sterilize) (all from Sigma Aldrich, St. Louis, MO, USA), samples were blocked with 10% goat serum (Thermo Fisher Scientific, Waltham, MA, USA)/1x IF wash for 1 h at RT and stained with primary antibodies diluted in blocking solution overnight at 4°C. All subsequent steps were performed in a humidified chamber. The next day, samples were washed three times in 1x IF wash and stained with respective secondary antibodies, which were diluted in blocking solution, for 1h at RT. Subsequently, cells were washed and incubated with the nuclear counterstain Hoechst 33342 (1:5000 in 1x PBS; Thermo Fisher Scientific, Waltham, MA, USA) for 7 min at RT, washed once in 1x IF wash and once in MilliPore water before mounting on glass slides using Vectashield mounting medium (Vector Laboratories, Inc., Newark, CA, USA). Fluorescent stainings were observed with the Leica fluorescent microscope DMI4000B (camera: Leica DFC7000T; both Leica, Wetzlar, Germany). The following primary and secondary antibodies are used: ms anti NESTIN, rb anti S100ß, rb anti TUBB3 (all Abcam, Cambridge, UK), rb anti P75 and ms anti TUBB3 (both Promega Corporation, Madison, WI, USA), rb anti MAP2 (Cell Signaling Technology, Danvers, MA, USA); goat anti-mouse IgG, AF488 and goat anti-mouse IgG, AF4568 (both Thermo Fisher Scientific, Waltham, MA, USA).

**Supplementary Figures**

**Suppl. Figure 1:** **WES results.** (A) Occurrence of rare genetic variants (MAF of <0.0001) including missense and frameshift variants, inframe deletions and insertions as well as splice variants in at least two patients without considering the inheritance model. The term “Data model” refers to “single” or “trio” whole exome sequencing.

**Suppl. Figure 2: Expression analyses of FFPE and *in vitro* samples focusing on stem and neuronal progenitor markers.**

**Suppl. Figure 3: Result of consensus *k*-means clustering of patient-specific DNA methylation profiles.** AG: aganglionic, G. ganglionic.

**Suppl. Figure 4: IF staining of HAND2.** Representative immunofluorescence staining of HAND2 (red) and TUBB3 (green) in ganglionic and aganglionic specimens from 4 patients. Nuclei are counterstained with Hoechst 33342 (blue).

**Suppl. Figure 5: Correlation of factors with covariates.** (A) Overview of factor correlation. (B) Correlation with patient samples’ origin - ganglionic, aganglionic. AG: aganglionic, G. ganglionic.

**Suppl. Figure 6: Network and enrichment analyses for feature weights of Factor 1.** (A) STRING network and Metascape-based GO term and disease enrichment heatmaps of CpG-related genes for Factor 1. (B) STRING network and Metascape enrichment heatmaps (GO terms, diseases) from *Neuropathology Panel* dataset-derived genes for Factor 1. (C) STRING network and GO term and disease enrichment heatmaps from *Elements Codeset* dataset-derived genes for Factor 1.

**Supplementary Tables**

**Suppl. Table 1: Elements Codeset.**

**Suppl. Table 2: Phenotypic data.**

**Suppl. Table 3: WES data_selected HSCR genes and SNPs.**

**Suppl. Table 4: WES data_MAF <0.001**

**Suppl. Table 5: WES data_MAF <0.0001**. *De novo* variants are marked in blue

**Suppl. Table 6: Expression profiling of FFPE specimens_Neuropathology panel.**

**Suppl. Table 7: Expression profiling of FFPE specimens_Elements codeset.**

**Suppl. Table 8: Expression profiling of *in vitro* specimens_Elements codeset.**

**Suppl. Table 9: Detailed list of DMPs.**

**Suppl. Table 10: Metascape results_hypo-hypermethyl. DMPs.**

**Suppl. Table 11: Covariates for MOFA.**

**Suppl. Table 12: Metascape results_Factor 1_CpGs_feature weights.**

**Suppl. Table 13: Metascape results_Factor 1_expression profiling_Neuropathology Panel_feature weights.**

**Suppl. Table 14: Metascape results_Factor 1_expression profiling_Elements Codeset_feature weights.**

1.	Muthuswamy, S.K., et al., *ErbB2, but not ErbB1, reinitiates proliferation and induces luminal repopulation in epithelial acini.* Nat Cell Biol, 2001. **3**(9): p. 785-92.
