## Supplementary figures and images for "An integrative multi-omics framework identifies epigenetic dysregulation of *HAND2* as a potential primary driver of impaired enteric neural crest cell differentiation in Hirschsprung Disease"

### Suppl. Figure 1

#### Rare functional – gnomAD–MAF < 0.0001, min_pid_counts > 2

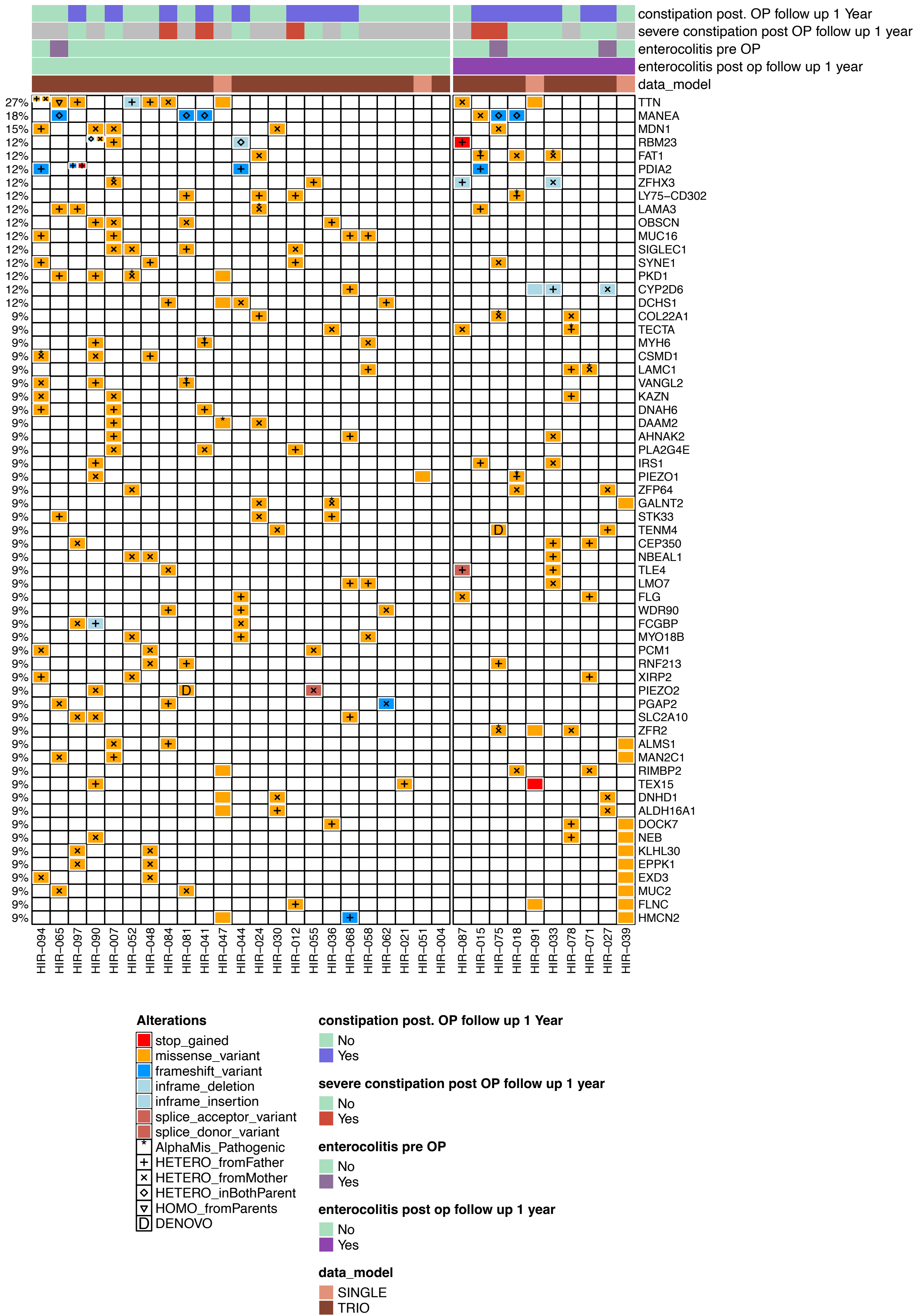

### Suppl. Figure 2

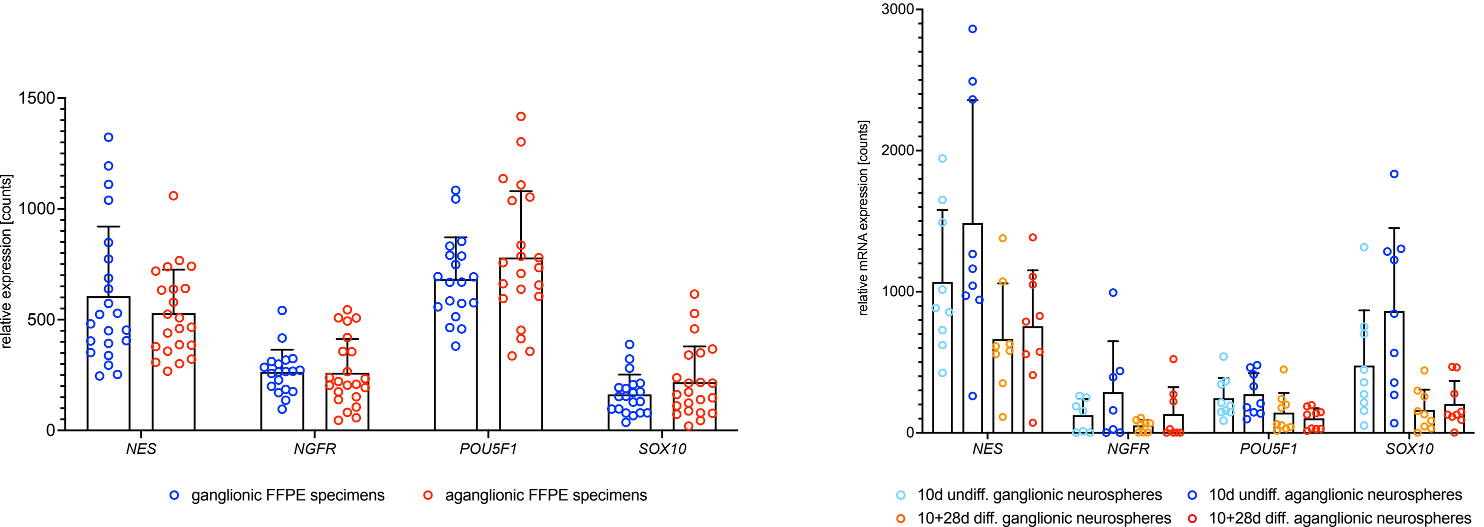

### Suppl. Figure 3

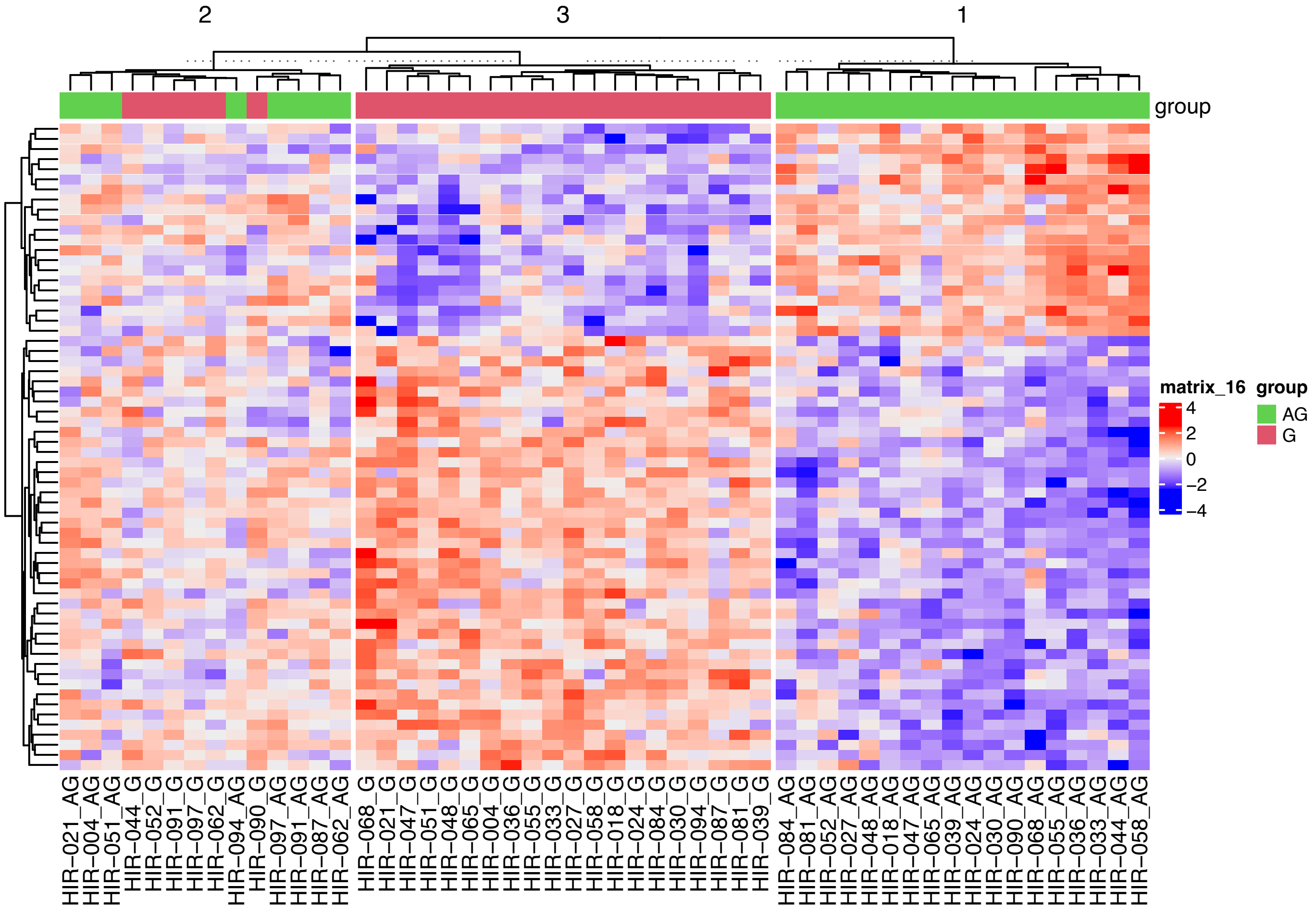

### Suppl. Figure 4

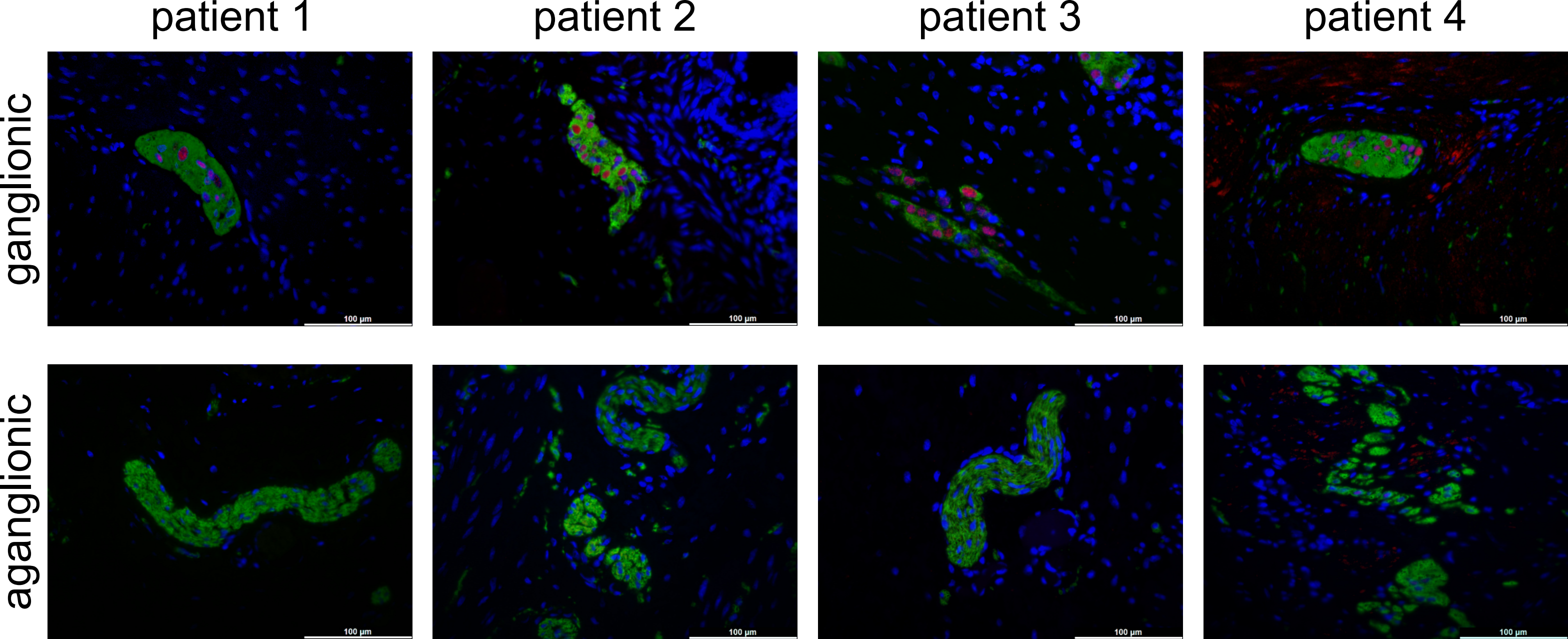

### Suppl. Figure 5

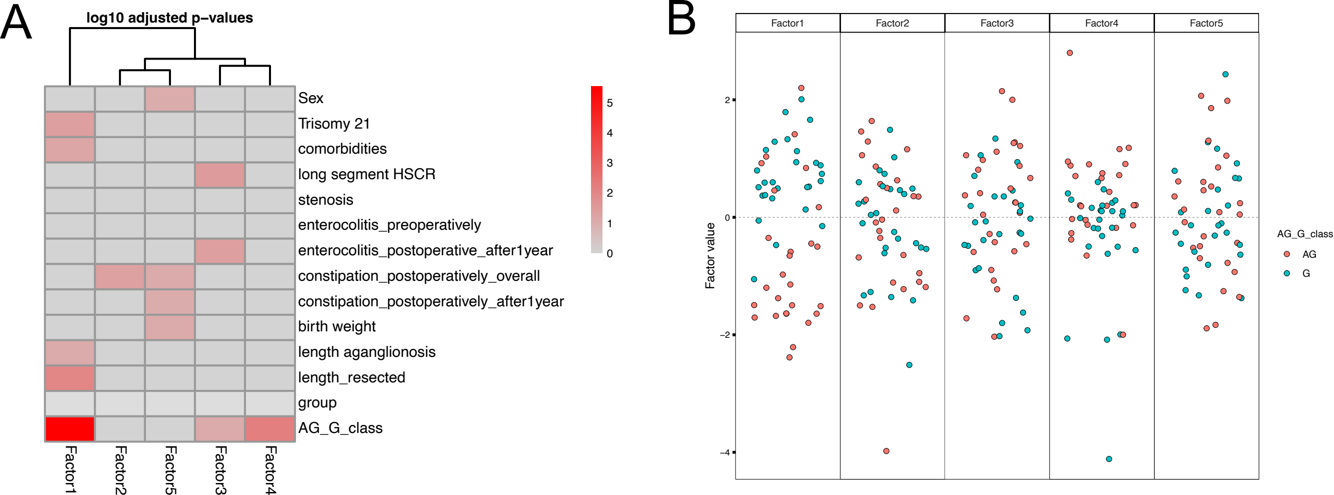

### Suppl. Figure 6

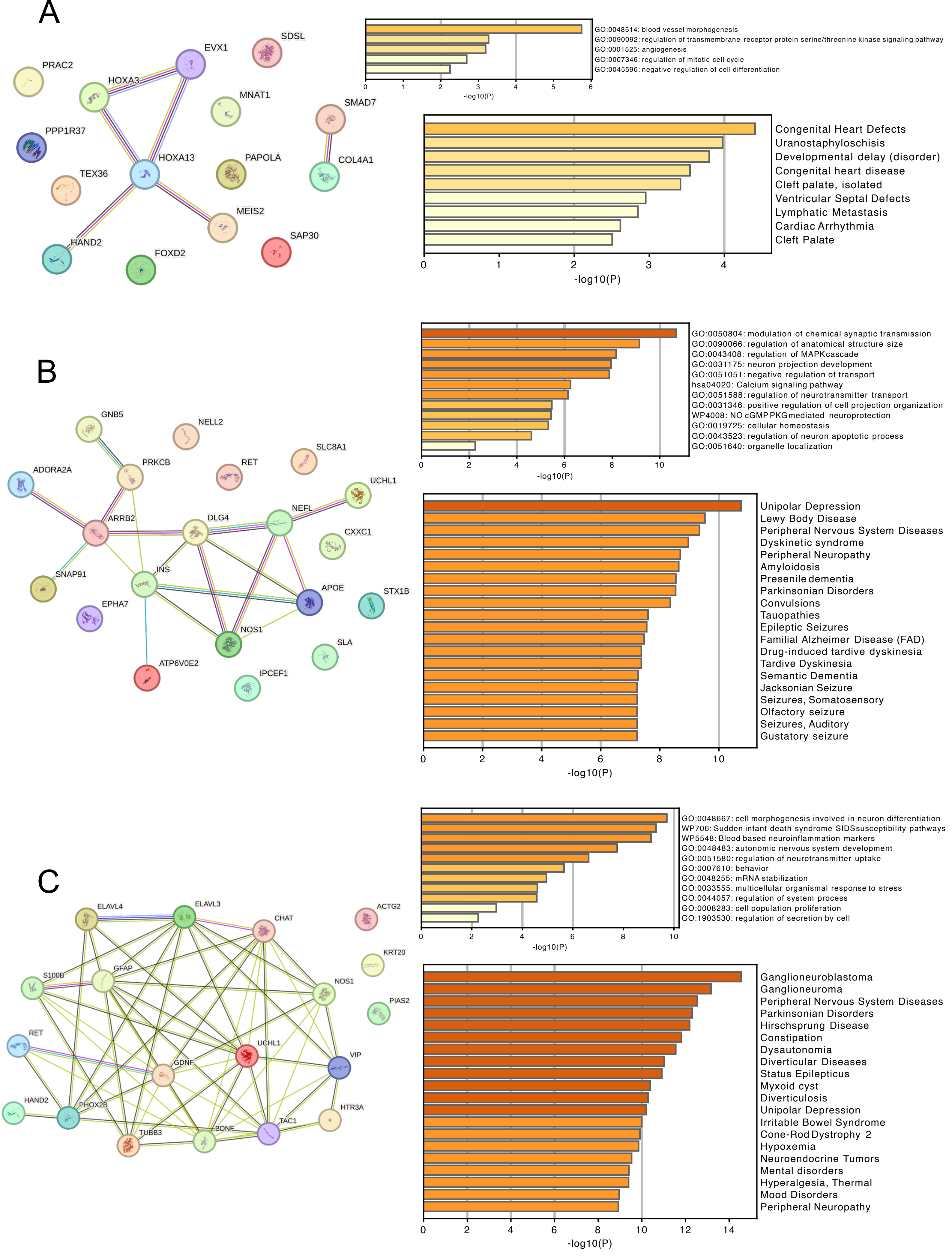
