## Supplementary material for "An integrative multi-omics framework identifies epigenetic dysregulation of *HAND2* as a potential primary driver of impaired enteric neural crest cell differentiation in Hirschsprung Disease": Suppl. Table 1

**Suppl. Table 1:** Elements Codeset.

| Target Gene | Theme | Accession Number | Target Sequence |
|---|---|---|---|
| ***ARF1*** | reference genes | NM_001024227.1 | CAATTCTGCATGGTCACAGTAGAGATCCCCGCAACTCGCTTGTCCTTGGGTCACC |
| | | | CTGCATTCCATAGCCATGTGCTTGTCCCTGTGCTCCCACGGTTCC |
| ***HPRT1*** | reference genes | NM_000194.1 | TGTGATGAAGGAGATGGGAGGCCATCACATTGTAGCCCTCTGTGTGCTCAAGGGG |
| | | | GGCTATAAATTCTTTGCTGACCTGCTGGATTACATCAAAGCACTG |
| ***PGK1*** | reference genes | NM_000291.3 | ATTGTCAAAGACCTAATGTCCAAAGCTGAGAAGAATGGTGTGAAGATTACCTTGCCT |
| | | | GTTGACTTTGTCACTGCTGACAAGTTTGATGAGAATGCCAAGA |
| ***PPIA*** | reference genes | NM_021130.2 | GGAATATTGAAAATGTAGGCAGCAACTGGGCATGGTGGCTCACTGTCTGTAATGTAT |
| | | | TACCTGAGGCAGAAGACCACCT |
| ***SDHA*** | reference genes | NM_004168.2 | AGTACATTGAAGGGAGATTGGCACCTAGTGGCTGGGAGCTTGCCAGGAACCCAGTG |
| | | | GCCAGGGAGCGTGGCACTTACCTTTGTCCCTTGCTTCATTCTTG |
| ***SNX17*** | reference genes | NM_014748.2 | CTTTCCTTGTCCCCTGGGCTGGCTGCACAGAGGATTGCCCCTTCTCTTTTCAGAGCT |
| | | | GGCCCTCGATGCCAAATTAGCATTTAGTATTTTGCACAAAGTC |
| ***UBB*** | reference genes | NM_018955.2 | CACCTGGTCCTGCGCCTGAGGGGTGGCTGTTAATTCTTCAGTCATGGCATTCGCAGT |
| | | | GCCCAGTGATGGCATTACTCTGCACTATAGCCATTTGCCCCAA |
| ***LGR5*** | stem & progenitor cell marker | NM_003667.2 | CTTATGCTTACCAGTGCTGTGCATTTGGAGTGTGTGAGAATGCCTATAAGATTTCTAA |
| | | | TCAATGGAATAAAGGTGACAACAGCAGTATGGACGACCTTCA |
| ***NES*** | stem & progenitor cell marker | NM_006617.1 | CAGAGAATCACAAATCACTGAGGTCTTTAGAAGAACAGGACCAAGAGACATTGAGAA |
| | | | CTCTTGAAAAAGAGACTCAACAGCGACGGAGGTCTCTAGGGGA |
| ***POU5F1*** | stem & progenitor cell marker | NM_002701.4 | AAGTTCTTCATTCACTAAGGAAGGAATTGGGAACACAAAGGGTGGGGGCAGGGGAGT |
| | | | TTGGGGCAACTGGTTGGAGGGAAGGTGAAGTTCAATGATGCTC |
| ***P75NTR*** | stem & progenitor cell marker | NM_002507.3 | TGAAGAAAAGTGGGCCAGTGTGGGAATGCGGCAAGAAGGAATTGACTTCGACTGTGA |
| | | | CCTGTGGGGATTTCTCCCAGCTCTAGACAACCCTGCAAAGGAC |
| ***SOX10*** | stem & progenitor cell marker | NM_006941.3 | GGCCGTGTCTCCCACTCAGGGGCTGAGAGTAGCTTTGAGGAGCCTCATTGGGGAG |
| | | | TGGGGGGTTCGAGGGACTTAGTGGAGTTCTCATCCCTTCAATGCC |
| ***ASCL1*** | neuronal cell marker | NM_004316.3 | ATAGTAACTCCCATCACCTCTAACACGCACAGCTGAAAGTTCTTGCTCGGGTCCCTT |
| | | | CACCTCCTCGCCCTTTCTTAAAGTGCAGTTCTTAGCCCTCTAG |
| ***ELAVL3*** | neuronal cell marker | NM_001420.3 | AGTCTCTTCGGCAGCATTGGCGACATCGAGTCCTGCAAGTTGGTTCGGGACAAGATC |
| | | | ACAGGGCAGAGCCTTGGCTACGGGTTTGTGAACTATTCTGACC |
| ***ELAVL4*** | neuronal cell marker | NM_021952.2 | GCCTCAGGTGTCAAATGGTCCGACATCCAATACAAGCAATGGACCCTCCAGCAACAAC |
| | | | AGAAACTGTCCTTCTCCCATGCAAACAGGGGCAACCACAGAT |
| ***HAND2*** | neuronal cell marker | NM_021973.2 | TCCATATTTTAATACGAAGAGGACACTCCCGTGTGGTAAGGGATCCCGTCGTCTCATAG |
| | | | ATTCTGTGTGCGTGAATGTTCCCTCTTGGCTGTGTAGACAC |
| ***PHOX2B*** | neuronal cell marker | NM_003924.3 | AAGGCACACACACACGTTTGCAGGGGTGTCTCGGTTTGCATTTCTGTTGGAATGATCCG |
| | | | AACTGGACTCACATCCTGTATGGTGGATGGACTGTATATTG |
| ***RET*** | neuronal cell marker | NM_020630.4 | AGGAGCCAGGGTCGGATTCCAGTTAAATGGATGGCAATTGAATCCCTTTTTGATCATATC |
| | | | TACACCACGCAAAGTGATGTATGGTCTTTTGGTGTCCTGC |
| ***TUBB3*** | neuronal cell marker | NM_006086.2 | GCCGCCCTCCTGCAGTATTTATGGCCTCGTCCTCCCCACCTAGGCCACGTGTGAGCTGC |
| | | | TCCTGTCTCTGTCTTATTGCAGCTCCAGGCCTGACGTTTTA |
| ***UCHL1*** | neuronal cell marker | NM_004181.3 | GAGCAAAATGCTTTGAAAAGAATGAGGCCATACAGGCAGCCCATGATGCCGTGGCACAG |
| | | | GAAGGCCAATGTCGGGTAGATGACAAGGTGAATTTCCATTT |
| ***BDNF*** | signaling molecules | NM_170732.4 | CAGGGTGATGCTCAGTAGTCAAGTGCCTTTGGAGCCTCCTCTTCTCTTTCTGCTGGAGGA |
| | | | ATACAAAAAATTACCTAGATGCTGCAAACATGTCCATGAGG |

| | | | |
|---|---|---|---|
| ***CGRP*** | signaling molecules | NM_001033953.2 | GGAAAGGCTCCATGGAAGACATACATATAGGCATCCTTCTTGATACTGAAAACTATCTTCTT<br>TGTTTGAAAGGAACTATTGCTAAATGCAGAACAAGCTC |
| ***CHAT*** | signaling molecules | NM_020549.4 | TCATTAATTTCCGCCGTCTCAGTGAGGGGGATCTGTTCACTCAGTTGAGAAAGATAGTCAAA<br>ATGGCTTCCAACGAGGACGAGCGTTTGCCTCCAATTGG |
| ***GDNF*** | signaling molecules | NM_000514.2 | CACTGACTTGGGTCTGGGCTATGAAACCAAGGAGGAACTGATTTTTAGGTACTGCAGCGGC<br>TCTTGCGATGCAGCTGAGACAACGTACGACAAAATATTG |
| ***NOS1*** | signaling molecules | NM_001204218.1 | GCGCTGGTGGAGATCAATATCGCGGTTCTCTATAGCTTCCAGAGTGACAAAGTGACCATTGT<br>TGACCATCACTCCGCCACCGAGTCCTTCATTAAGCACA |
| ***NPY*** | signaling molecules | NM_000905.2 | AGAGATATGGAAAACGATCCAGCCCAGAGACACTGATTTCAGACCTCTTGATGAGAGAAAGC<br>ACAGAAAATGTTCCCAGAACTCGGCTTGAAGACCCTGC |
| ***TAC1*** | signaling molecules | NM_003182.2 | GTCCGTCGCAAAATCCAACATGAAAATCCTCGTGGCCTTGGCAGTCTTTTTTCTTGTCTCCAC<br>TCAGCTGTTTGCAGAAGAAATAGGAGCCAATGATGAT |
| ***VIP*** | signaling molecules | NM_003381.2 | CACCCTATTATGATGTATCCAGAAATGCCAGGCATGCTGATGGAGTTTTCACCAGTGACTTCA<br>GTAAACTCTTGGGTCAACTTTCTGCCAAAAAGTACCT |
| ***HTR3A*** | serotonergic marker | NM_000869.5 | TTTCACAACTTTGCTTTTAGGTTGAAGGCAAAACCAACTCTCTACTACACAGGCCTGATAACTC<br>TGTACGAGGCTTCTCTAACCCCTAGTGTCTTTTTTT |
| ***HTR3B*** | serotonergic marker | NM_006028.4 | GACATTGAAAGATACCCTGACCTTCCCTATGTTTATGTGAACTCATCTGGGACCATTGAGAACT<br>ATAAGCCCATCCAGGTGGTCTCTGCGTGCAGTTTAG |
| ***HTR3E*** | serotonergic marker | NM_182589.2 | CCCCTTTCCTGAGTACCAACTATCATATCCCCAAAGATGACTGAGTCTCTGCTGTATTCCATGT<br>ATCCCAATCCGGTCCTGCTGATCAATTCCAATCCCA |
| ***HTR4a*** | serotonergic marker | NM_001040169.2 | TTAATGGATCCACACATGTACTAAGGTACACCGTTCTGCACAGGGGACATCATCAGGAACTCG<br>AGAAACTGCCCATACACAATGACCCAGAATCCCTGGA |
| ***HTR4b*** | serotonergic marker | NM_000870.6 | GCAGGGCAGCGGGGACCACCGGGGCTGGGGGCTGTTGAGCCCGTGGAGTCCGGCTCGGTT<br>GGGGAGAAGGACGATGCGCGGCGAGCCCAGGTGATCCGGG |
| ***HTR4b_2*** | serotonergic marker | Niesler_S_HTR4b_2.1 | GCCTCCGAAAGAGGGCCAGGTCCTAAGCTGCTGCTTGTGCGCGACTGCACCCGTTCTGGAAC<br>TGAAACCGACAGGAAGAAACTTTGGAATAAGGAAGAGA |
| ***SLC6A4*** | serotonergic marker | NM_001045.2 | CCCAGAGATCAATTGGGATCCTTGGCAGATGGACATCAGTGTCATTTACTAACCAGCAGGATGG<br>AGACGACGCCCTTGAATTCTCAGAAGCAGCTATCAG |
| ***TPH1*** | serotonergic marker | NM_004179.2 | TTGGCTGAACCTAGTTTTGCCCAATTCTCCCAAGAAATTGGCTTGGCTTCTCTTGGCGCTTC<br>AGAGGAGGCTGTTCAAAAACTGGCAACGTGCTACTTTT |
| ***TPH2*** | serotonergic marker | NM_173353.3 | ATGTAGGTTGCGTTGACCTTGTAGAACCTGAGTTATGACAAGCTTCCTGAAGTATTTTGGAA<br>GATAGTACTTCCGGAAAGGACATTAGGAAAGACTAAAC |
| ***GFAP*** | glial cell marker | NM_002055.4 | AAGCAGATGAAGCCACCCTGGCCCGTCTGGATCTGGAGAGGAAGATTGAGTCGCTGGAGG<br>AGGAGATCCGGTTCTTGAGGAAGATCCACGAGGAGGAGGT |
| ***S100B*** | glial cell marker | NM_006272.1 | AGAAGGCCATGGTGGCCCTCATCGACGTTTTCCACCAATATTCTGGAAGGGAGGGAGACAA<br>GCACAAGCTGAAGAAATCCGAACTCAAGGAGCTCATCAA |
| ***ALPi*** | epithelial cell marker | NM_001631.3 | CAGCAAGGCTCAGGACAGCAAAGCCTACACGTCCATCCTGTACGGCAATGGCCCGGGCTA<br>CGTGTTCAACTCAGGCGTGCGACCAGACGTGAATGAGAGC |
| ***CHGA*** | epithelial cell marker | NM_001275.3 | CTGCGCCGGGCAAGTCACTGCGCTCCCTGTGAACAGCCCTATGAATAAAGGGGATACCGA<br>GGTGATGAAATGCATCGTTGAGGTCATCTCCGACACACTT |
| ***CK20*** | epithelial cell marker | NM_019010.1 | ACAGTCGTGCAAGAAGTAGTGGATGGCAAGGTCGTGTCATCTGAAGTCAAAGAGGTGGAAG<br>AAAATATCTAAATAGCTACCAGAAGGAGATGCTGCTGAG |
| ***DEFA5*** | epithelial cell marker | NM_021010.1 | TATTGCCGAACCGGCCGTTGTGCTACCCGTGAGTCCCTCTCCGGGGTGTGTGAAATCAGTG<br>GCCGCCTCTACAGACTCTGCTGTCGCTGAGCTTCCTAGA |

| | | | |
|---|---|---|---|
| ***MUC2*** | epithelial cell marker | NM_002457.2 | GGAGCAGCTAGGCCAGAAGGTGCAGTGTGATGTCTCTGTTGGGTTCATTTGCAAGAATGAAG |
| | | | ACCAGTTTGGAAATGGACCATTTGGACTGTGTTACGAC |
| ***SI*** | epithelial cell marker | NM_001041.3 | TAGGAAAAGCAACGGTAAAACTTTGTTTGACACCAGCATTGGTCCCTTAGTGTACTCTGACCA |
| | | | GTACTTACAGATCTCAACCCGTCTTCCAAGTGATTAT |
| ***VIL*** | epithelial cell marker | NM_007127.1 | GATGTCCTGGAAGAGTTTCAACCGAGGGGATGTTTTCCTCCTGGACCTTGGGAAGCTTATCA |
| | | | TCCAGTGGAATGGACCGGAAAGCACCCGTATGGAGAGA |
| ***ACTG2*** | smooth muscle cell marker | NM_001615.3 | TTATTGGCATGGAGTCCGCTGGAATTCATGAGACAACCTACAATTCCATCATGAAGTGTGACA |
| | | | TTGACATCCGTAAGGACTTATATGCCAACAATGTCCT |
