## Supplementary material for "An integrative multi-omics framework identifies epigenetic dysregulation of *HAND2* as a potential primary driver of impaired enteric neural crest cell differentiation in Hirschsprung Disease": Suppl. Table 2

**Suppl. Table 2:** Phenotypic data. SD = standard deviation; HSCR =Hirschsprung disease; TERPT= transanal endorectal pullthrough; BMI = Body-Mass-Index; n.s.= not significant; OR = odds ratio; CI = confidence interval.

| **(A) Demographic data of HSCR patients** | | |
|---|---|---|
| | **n=33** | **%** |
| **sex** | | |
| - male | 31 | 93.9 |
| - female | 2 | 6.0 |
| **associated congenital malformations** | 14 | 42.4 |
| - Trisomy 21 | 10 | 30.3 |
| - congenital anomalies of the kidney | 9 | 27.2 |
| - congenital heart defects | 2 | 6.0 |
| | **mean** | **SD** |
| birth weight (g) | 3048.9 | 652.3 |
| weeks of gestation | 37.9 | 2.8 |

| **(B) Perioperative data of HSCR patients** | | |
|---|---|---|
| | **n=33** | **%** |
| **operation procedures** | | |
| - TERPT | 31 | 93.9 |
| - Soave procedure | 1 | 3.0 |
| - Rehbein procedure | 1 | 3.0 |
| **long-segment HSCR** | 11 | 33.3 |
| **region of aganglionosis** | | |
| - Rectosigmoid | 18 | 54.5 |
| - Colon | 13 | 39.3 |
| - Jirásek-Zuelzer-Wilson-Syndrom | 2 | 6.0 |
| | **mean** | **SD** |
| age of HSCR operation (month) | 8.8 | 10.6 |
| weight at HSCR operation (kg) | 6.9 | 3.0 |
| length of aganglionosis (cm) | 18.4 | 14.4 |
| length of intestinal resection (cm) | 30.0 | 13.1 |
| length of hospital stay (days) | 13.4 | 11.2 |

| **(D) Comparison long-segment vs non-long-segment HSCR** | | | | | |
|---|---|---|---|---|---|
| | **long-segment (n=11)** | **non-long-segment (n=22)** | ***p*-value** | **OR** | **95%-CI** |
| Birth weight (g), mean ± SD | 2770±817.4 | 3188±518.8 | 0.08 | | |
| Weeks of gestation, mean ± SD | 36.5±4.4 | 38.6±1.2 | 0.04 | | |
| Associated congenital malformations, n (%) | 7 (63.6%) | 7 (31.8%) | 0.08 | 3.7 | 0.8-17.1 |
| Length of intestinal resection (cm), mean ± SD | 41.6±14.7 | 24.3±7.2 | 0.003 | | |
| Constipation postoperative, n (%) | 6 (54.5%) | 11 (50%) | n.s. | 0.8 | 0.2-3.5 |
| Enterocolitis postoperative, n (%) | 2 (18.2%) | 8 (36.4%) | n.s. | 0.3 | 0.06-2.2 |
| Stool frequency (per week), mean ± SD | 24±17.6 | 13.6±8.8 | 0.03 | | |
| Stool consistency, mean ± SD | 5.1±0.7 | 4.5±0.5 | 0.007 | | |

| **(E) Clinical Data of parents** | | |
|---|---|---|
| **sex** | **n=35** | **%** |
| - male | 17 | 48.6 |
| - female | 8 | 51.4 |
| **conditions** | | |
| - constipation | 8 | 22.9 |
| - Enterocolitis | 6 | 17.1 |
| - diet | 11 | 31.4 |
| | **mean** | **SD** |
| age | 39.3 | 7.8 |
| BMI | 27.3 | 6.6 |
| stool frequency (per week) | 10.2 | 6.6 |
| stool consistency | 3.5 | 1 |
| age of bowel control | 2.8 | 0.7 |
| Rintala score | 19.4 | 1.2 |

| (C) Postoperative complications after corrective HSCR surgery | | |
|---|---|---|
| | **n=33** | **%** |
| Enterocolitis postoperative | 10 | 30.3 |
| constipation postoperative | 16 | 48.5 |
| | **mean** | **SD** |
| duration of constipation (month) | 4.1 | 2.8 |
| stool frequency (per week) | 17 | 13.1 |
| stool consistency (Bristol stool scale) | 4.7 | 0.7 |
