## Supplementary material for "An integrative multi-omics framework identifies epigenetic dysregulation of *HAND2* as a potential primary driver of impaired enteric neural crest cell differentiation in Hirschsprung Disease": Suppl. Table 3

**Suppl. Table 3:** Expression profiling of FFPE specimens_Neuropathology panel.

| Genes | p value | q value | -log10(q value) |
|---|---|---|---|
| *UCHL1* | <0,000001 | <0,000001 | 7,187 |
| *VIP* | <0,000001 | 0,000003 | 5,489 |
| *PLLP* | <0,000001 | 0,00007 | 4,156 |
| *CALB2* | 0,000004 | 0,000823 | 3,085 |
| *NEFL* | 0,00002 | 0,003081 | 2,511 |
| *NOS1* | 0,000035 | 0,004448 | 2,352 |
| *MYRF* | 0,000067 | 0,007269 | 2,139 |
| *SNAP91* | 0,000133 | 0,012611 | 1,899 |
| *CACNA1B* | 0,000459 | 0,038591 | 1,414 |
| *PTGS2* | 0,000626 | 0,047324 | 1,325 |
| *ATP6V0E2* | 0,000874 | 0,060089 | 1,221 |
| *NELL2* | 0,000976 | 0,061547 | 1,211 |
| *INA* | 0,0015 | 0,07949 | 1,1 |
| *EPHA3* | 0,001532 | 0,07949 | 1,1 |
| *EPHA2* | 0,001612 | 0,07949 | 1,1 |
| *GRIA4* | 0,001863 | 0,07949 | 1,1 |
| *CERS6* | 0,001968 | 0,07949 | 1,1 |
| *STX1B* | 0,002001 | 0,07949 | 1,1 |
| *PIK3CB* | 0,002208 | 0,07949 | 1,1 |
| *LAMA2* | 0,002606 | 0,07949 | 1,1 |
| *EPHA7* | 0,00263 | 0,07949 | 1,1 |
| *NEO1* | 0,002648 | 0,07949 | 1,1 |
| *SLC8A1* | 0,002705 | 0,07949 | 1,1 |
| *SLC2A1* | 0,002728 | 0,07949 | 1,1 |
| *SYT1* | 0,00273 | 0,07949 | 1,1 |
| *GNAI1* | 0,002823 | 0,07949 | 1,1 |
| *TF* | 0,002837 | 0,07949 | 1,1 |
| *CDK5* | 0,002974 | 0,080345 | 1,095 |
| *CX3CR1* | 0,003188 | 0,083171 | 1,08 |
| *CDS1* | 0,003732 | 0,094113 | 1,026 |
| *CDK7* | 0,004009 | 0,097827 | 1,01 |
| *NRG1* | 0,004384 | 0,103645 | 0,9845 |
| *TBPL1* | 0,004576 | 0,104899 | 0,9792 |
| *CTSE* | 0,006623 | 0,147212 | 0,8321 |
| *CCND1* | 0,007064 | 0,147212 | 0,8321 |
| *LCLAT1* | 0,007064 | 0,147212 | 0,8321 |
| *GRIA3* | 0,007207 | 0,147212 | 0,8321 |
| *SCN1A* | 0,007395 | 0,147212 | 0,8321 |
| *ARHGAP44* | 0,009013 | 0,170461 | 0,7684 |
| *MBP* | 0,009013 | 0,170461 | 0,7684 |
| *AP3S1* | 0,010935 | 0,20177 | 0,6951 |
| *GDNF* | 0,013092 | 0,235808 | 0,6274 |
| *RET* | 0,013831 | 0,243326 | 0,6138 |
| *GNPTAB* | 0,014363 | 0,246938 | 0,6074 |
| *LPAR1* | 0,016067 | 0,270097 | 0,5685 |
| *CACNA1C* | 0,016642 | 0,273689 | 0,5627 |

| | |
|---|---|
| 1% q-value | adjusted Significance |
| 5% q-value | adjusted Significance |
| P value | nominal Significance |

known HSCR genes

| | | | |
|---|---|---|---|
| *NWD1* | 0,017376 | 0,275401 | 0,56 |
| *CHL1* | 0,017474 | 0,275401 | 0,56 |
| *BCL2L1* | 0,017942 | 0,277001 | 0,5575 |
| *ACVRL1* | 0,018948 | 0,281063 | 0,5512 |
| *ENTPD2* | 0,018948 | 0,281063 | 0,5512 |
| *GFPT1* | 0,021059 | 0,301261 | 0,5211 |
| *CADPS* | 0,021106 | 0,301261 | 0,5211 |
| *CXCL16* | 0,022249 | 0,306199 | 0,514 |
| *FAS* | 0,022262 | 0,306199 | 0,514 |
| *SMYD1* | 0,023471 | 0,306593 | 0,5134 |
| *GNAO1* | 0,023497 | 0,306593 | 0,5134 |
| *L1CAM* | 0,023506 | 0,306593 | 0,5134 |
| *FN1* | 0,026171 | 0,335567 | 0,4742 |
| *PLA2G2A* | 0,027439 | 0,343315 | 0,4643 |
| *CHRNA7* | 0,027683 | 0,343315 | 0,4643 |
| *ANG* | 0,028881 | 0,352393 | 0,453 |
| *PLEKHO2* | 0,029517 | 0,354434 | 0,4505 |
| *IL6* | 0,030243 | 0,357473 | 0,4468 |
| *KIAA1161* | 0,031532 | 0,366984 | 0,4354 |
| *EMCN* | 0,032642 | 0,371567 | 0,43 |
| *OXR1* | 0,032909 | 0,371567 | 0,43 |
| *TENM2* | 0,034843 | 0,387107 | 0,4122 |
| *SRC* | 0,035308 | 0,387107 | 0,4122 |
| *ACHE* | 0,037416 | 0,404352 | 0,3932 |
| *KRAS* | 0,040879 | 0,429719 | 0,3668 |
| *EGF* | 0,040899 | 0,429719 | 0,3668 |
| *GALC* | 0,041611 | 0,431204 | 0,3653 |
| *GAL3ST1* | 0,042375 | 0,433192 | 0,3633 |
| *MPZ* | 0,043718 | 0,44096 | 0,3556 |
| *MAG* | 0,045496 | 0,452855 | 0,344 |
| *CAMK4* | 0,047162 | 0,458873 | 0,3383 |
| *PLCB3* | 0,047452 | 0,458873 | 0,3383 |
| *DCX* | 0,04792 | 0,458873 | 0,3383 |
| *PRKCA* | 0,049264 | 0,462103 | 0,3353 |
| *GPR4* | 0,050149 | 0,462103 | 0,3353 |
| *DGKB* | 0,050883 | 0,462103 | 0,3353 |
