## Supplementary material for "An integrative multi-omics framework identifies epigenetic dysregulation of *HAND2* as a potential primary driver of impaired enteric neural crest cell differentiation in Hirschsprung Disease": Suppl. Table 4

**Suppl. Table 4:** Expression profiling of FFPE specimens_Elements codeset.

| Genes | p value | q value | -log10(q value) |
|---|---|---|---|
| *nNOS* | <0.000001 | <0.000001 | 7,139 |
| *UCHL1* | <0.000001 | <0.000001 | 7,139 |
| *VIP* | <0.000001 | <0.000001 | 6,921 |
| *ELAVL3* | 0,000023 | 0,000171 | 3,766 |
| *TUBB3* | 0,000032 | 0,000193 | 3,715 |
| *ELAVL4* | 0,000139 | 0,000702 | 3,154 |
| *TPH1* | 0,000513 | 0,002222 | 2,653 |
| *CHGA* | 0,000868 | 0,003289 | 2,483 |
| *HAND2* | 0,00176 | 0,005924 | 2,227 |
| *PHOX2B* | 0,003449 | 0,010449 | 1,981 |
| *MUC2* | 0,016601 | 0,045729 | 1,34 |
| *Sucrose-Isomaltase* | 0,027434 | 0,069272 | 1,159 |
| *RET* | 0,045023 | 0,104937 | 0,9791 |

| | |
|---|---|
| 1% q-value | adjusted Significance |
| 5% q-value | adjusted Significance |
| P value | nominal Significance |

known HSCR genes
