## Supplementary material for "An integrative multi-omics framework identifies epigenetic dysregulation of *HAND2* as a potential primary driver of impaired enteric neural crest cell differentiation in Hirschsprung Disease": Suppl. Table 5

**Suppl. Table 9:** Detailed list of DMPs.

| | Chr | start | end | width | strand | nearest_tss | nearest_tss symbol | nearest_tss dist | p_value | q_value | cmp | diff.G.AG. |
|---|---|---|---|---|---|---|---|---|---|---|---|---|
| **cg11513700** | chr7 | 27205677 | 27205677 | 1 | - | ENSG00000106031.9 | HOXA13 | 5585 | 4,31198350847998E-09 | 9,78249331364004E-05 | G < AG | -0,180999664 |
| **cg16980686** | chr1 | 75423226 | 75423226 | 1 | + | ENSG00000162624.15 | LHX8 | 294793 | 1,25262845380531E-08 | 0,000175038 | G < AG | -0,168128129 |
| **cg19788317** | chr4 | 173508149 | 173508149 | 1 | + | ENSG00000164105.4 | SAP30 | 138181 | 2,44901044104206E-08 | 0,000246494 | G < AG | -0,197280682 |
| **cg00070383** | chr4 | 173508223 | 173508223 | 1 | - | ENSG00000164107.9 | HAND2 | 22007 | 4,96605192221719E-08 | 0,00034142 | G < AG | -0,219442196 |
| **cg24974365** | chr4 | 173527398 | 173527398 | 1 | - | ENSG00000164107.9 | HAND2 | 2832 | 5,03389171241708E-08 | 0,00034142 | G < AG | -0,088942693 |
| **cg08195247** | chr2 | 176068558 | 176068558 | 1 | + | ENSG00000128714.6 | HOXD13 | 24162 | 4,84225643964017E-08 | 0,00034142 | G < AG | -0,182281677 |
| **cg20913646** | chr10 | 126136744 | 126136744 | 1 | - | ENSG00000089876.12 | DHX32 | 240307 | 6,93938024352388E-08 | 0,000421289 | G < AG | -0,06195418 |
| **cg22778379** | chr17 | 61402200 | 61402200 | 1 | + | ENSG00000121068.14 | TBX2 | 2358 | 7,83662045362926E-08 | 0,000447357 | G < AG | -0,121255624 |
| **cg20725013** | chr7 | 27155417 | 27155417 | 1 | - | ENSG00000122592.8 | HOXA7 | 2520 | 8,77480011231745E-08 | 0,000474595 | G < AG | -0,159218234 |
| **cg26491604** | chr17 | 48579502 | 48579502 | 1 | + | ENSG00000229637.4 | PRAC2 | 143682 | 9,63085468064795E-08 | 0,000494434 | G < AG | -0,188107976 |
| **cg08082810** | chr14 | 60641673 | 60641673 | 1 | + | ENSG00000020426.11 | MNAT1 | 93068 | 1,27496298336738E-07 | 0,000593162 | G < AG | -0,153870081 |
| **cg16756004** | chr10 | 14085186 | 14085186 | 1 | + | ENSG00000282246.1 | RP11-295P9.13 | 475140 | 1,29116885710868E-07 | 0,000594585 | G < AG | -0,075184624 |
| **cg00588920** | chr8 | 72004912 | 72004912 | 1 | + | ENSG00000182674.6 | KCNB2 | 532312 | 1,40162740096894E-07 | 0,000624866 | G < AG | -0,100710029 |
| **cg10839823** | chr4 | 173508219 | 173508219 | 1 | - | ENSG00000164107.9 | HAND2 | 22011 | 1,62940621913938E-07 | 0,000692448 | G < AG | -0,185030206 |
| **cg03838927** | chr2 | 176111317 | 176111317 | 1 | - | ENSG00000174279.4 | EVX2 | 27403 | 2,08886999456724E-07 | 0,000819421 | G < AG | -0,14020433 |
| **cg12192691** | chr7 | 27252076 | 27252076 | 1 | + | ENSG00000106038.13 | EVX1 | 9377 | 2,32653132996202E-07 | 0,000875937 | G < AG | -0,105648342 |
| **cg00690402** | chr17 | 48592204 | 48592204 | 1 | - | ENSG00000120075.6 | HOXB5 | 1576 | 2,36404166676444E-07 | 0,000877845 | G < AG | -0,16237071 |
| **cg11010578** | chr7 | 50383580 | 50383580 | 1 | + | ENSG00000185811.19 | IKZF1 | 79513 | 2,50025619461205E-07 | 0,000897915 | G < AG | -0,110559128 |
| **cg09842331** | chr7 | 27249933 | 27249933 | 1 | + | ENSG00000106038.13 | EVX1 | 7234 | 2,77333290334017E-07 | 0,00094272 | G < AG | -0,134225086 |
| **cg11352083** | chr17 | 48597152 | 48597152 | 1 | + | ENSG00000229637.4 | PRAC2 | 126032 | 2,70285591699187E-07 | 0,00094272 | G < AG | -0,136667124 |
| **cg21778348** | chr7 | 27154995 | 27154995 | 1 | - | ENSG00000105997.23 | HOXA3 | 2411 | 2,89632261765777E-07 | 0,000963273 | G < AG | -0,137144745 |
| **cg17526483** | chr7 | 27241785 | 27241785 | 1 | + | ENSG00000106038.13 | EVX1 | 914 | 2,99101931142184E-07 | 0,00098909 | G < AG | -0,122152706 |
| **cg08396193** | chr7 | 27154090 | 27154090 | 1 | - | ENSG00000105997.23 | HOXA3 | 1506 | 3,00737131249782E-07 | 0,00098909 | G < AG | -0,101777512 |
| **cg07986943** | chr2 | 176067851 | 176067851 | 1 | + | ENSG00000128714.6 | HOXD13 | 24869 | 3,11352091061596E-07 | 0,000998353 | G < AG | -0,172655471 |
| **cg05822888** | chr17 | 48579530 | 48579530 | 1 | + | ENSG00000229637.4 | PRAC2 | 143654 | 3,12560719488742E-07 | 0,000998353 | G < AG | -0,202442059 |
| **cg19911609** | chr14 | 96497233 | 96497233 | 1 | + | ENSG00000090060.19 | PAPOLA | 4199 | 3,41331525012857E-07 | 0,001041727 | G < AG | -0,133200452 |
| **cg07674312** | chr10 | 125606528 | 125606528 | 1 | - | ENSG00000175018.13 | TEX36 | 76636 | 3,72441827223308E-07 | 0,001087395 | G < AG | -0,134022841 |

| | | | | | | | | | | | | |
|---|---|---|---|---|---|---|---|---|---|---|---|---|
| **cg00921669** | chr5 | 51465248 | 51465248 | 1 | - | ENSG00000170571.12 | EMB | 1021999 | 3,74329470080212E-07 | 0,001088751 | G < AG | -0,128293124 |
| **cg03396151** | chr15 | 37095103 | 37095103 | 1 | - | ENSG00000134138.20 | MEIS2 | 6197 | 3,92467031404559E-07 | 0,001120208 | G < AG | -0,166492154 |
| **cg04922154** | chr7 | 27244729 | 27244729 | 1 | + | ENSG00000106038.13 | EVX1 | 2030 | 4,61700875067155E-07 | 0,001261342 | G < AG | -0,119746729 |
| **cg14884929** | chr17 | 48580294 | 48580294 | 1 | + | ENSG00000229637.4 | PRAC2 | 142890 | 4,81773397671327E-07 | 0,001297641 | G < AG | -0,122331947 |
| **cg10620616** | chr7 | 27243986 | 27243986 | 1 | - | ENSG00000106031.9 | HOXA13 | 43894 | 4,88932016924462E-07 | 0,001300346 | G < AG | -0,113353799 |
| **cg23096689** | chr16 | 86566108 | 86566108 | 1 | + | ENSG00000176692.8 | FOXC2 | 720 | 5,09665886417701E-07 | 0,001335158 | G < AG | -0,083273358 |
| **cg22964918** | chr7 | 27246456 | 27246456 | 1 | - | ENSG00000106031.9 | HOXA13 | 46364 | 5,31917621128347E-07 | 0,001351786 | G < AG | -0,225888543 |
| **cg27028202** | chr6 | 165686828 | 165686828 | 1 | - | ENSG00000112541.18 | PDE10A | 301251 | 5,72086201973155E-07 | 0,001396696 | G < AG | -0,109088803 |
| **cg27198632** | chr4 | 173508112 | 173508112 | 1 | + | ENSG00000164105.4 | SAP30 | 138144 | 5,77062236711264E-07 | 0,001396696 | G < AG | -0,159708079 |
| **cg26708220** | chr15 | 37095376 | 37095376 | 1 | - | ENSG00000134138.20 | MEIS2 | 5924 | 5,82408933315556E-07 | 0,001400977 | G < AG | -0,117564418 |
| **cg03051777** | chr12 | 114693984 | 114693984 | 1 | + | ENSG00000139410.15 | SDSL | 1271605 | 6,08456801249159E-07 | 0,001449956 | G < AG | -0,147094925 |
| **cg15348720** | chr12 | 20404782 | 20404782 | 1 | + | ENSG00000172572.7 | PDE3A | 36246 | 6,46016837503388E-07 | 0,001497476 | G < AG | -0,100276485 |
| **cg27138072** | chr21 | 43600811 | 43600811 | 1 | - | ENSG00000160207.9 | HSF2BP | 58678 | 7,04949864345049E-07 | 0,001590701 | G < AG | -0,12168903 |
| **cg26484116** | chr22 | 46735157 | 46735157 | 1 | - | ENSG00000100422.14 | CERK | 3096 | 7,28094397569908E-07 | 0,001607324 | G < AG | -0,133729722 |
| **cg10764068** | chr7 | 27250210 | 27250210 | 1 | + | ENSG00000106038.13 | EVX1 | 7511 | 7,49832430558125E-07 | 0,001607324 | G < AG | -0,115264701 |
| **cg27218796** | chr4 | 173526977 | 173526977 | 1 | - | ENSG00000164107.9 | HAND2 | 3253 | 7,43208069511118E-07 | 0,001607324 | G < AG | -0,120974522 |
| **cg17062109** | chr17 | 48592667 | 48592667 | 1 | - | ENSG00000120075.6 | HOXB5 | 1113 | 7,40900862708672E-07 | 0,001607324 | G < AG | -0,110268428 |
| **cg19229651** | chr7 | 41911521 | 41911521 | 1 | - | ENSG00000122641.11 | INHBA | 205686 | 7,2968133319822E-07 | 0,001607324 | G < AG | -0,112993525 |
| **cg09554952** | chr17 | 3127587 | 3127587 | 1 | - | ENSG00000183024.4 | OR1G1 | 35 | 7,22891480961076E-07 | 0,001607324 | G < AG | -0,071377788 |
| **cg20556517** | chr19 | 18429520 | 18429520 | 1 | + | ENSG00000130511.16 | SSBP4 | 10657 | 7,50336685986241E-07 | 0,001607324 | G < AG | -0,189372322 |
| **cg06355422** | chr22 | 49620001 | 49620001 | 1 | - | ENSG00000100425.18 | BRD1 | 207512 | 7,93162079291298E-07 | 0,001657401 | G < AG | -0,063207795 |
| **cg00538017** | chr6 | 63319965 | 63319965 | 1 | + | ENSG00000198225.5 | FKBP1C | 108520 | 8,43272033066455E-07 | 0,00172475 | G < AG | -0,089045988 |
| **cg10760958** | chr15 | 96412115 | 96412115 | 1 | + | ENSG00000185551.15 | NR2F2 | 86178 | 8,64389773749366E-07 | 0,001740028 | G < AG | -0,15356766 |
| **cg06799670** | chr4 | 87074183 | 87074183 | 1 | + | ENSG00000172493.23 | AFF1 | 139182 | 8,76379667725337E-07 | 0,001750345 | G < AG | -0,107099288 |
| **cg26187080** | chr8 | 143460693 | 143460693 | 1 | - | ENSG00000182759.4 | MAFA | 29960 | 1,14147378951374E-06 | 0,002054505 | G < AG | -0,057374464 |
| **cg00772428** | chr15 | 75902388 | 75902388 | 1 | + | ENSG00000167196.14 | FBXO22 | 1487 | 1,20746416055194E-06 | 0,002153015 | G < AG | -0,10297867 |
| **cg12814766** | chr14 | 70783707 | 70783707 | 1 | - | ENSG00000006432.16 | MAP3K9 | 25828 | 1,38486740779141E-06 | 0,002354105 | G < AG | -0,069076241 |
| **cg17863743** | chr10 | 99233830 | 99233830 | 1 | + | ENSG00000119946.11 | CNNM1 | 95525 | 1,41152720179015E-06 | 0,002378283 | G < AG | -0,107824574 |
| **cg10101521** | chr2 | 176068404 | 176068404 | 1 | + | ENSG00000128714.6 | HOXD13 | 24316 | 1,42777624306074E-06 | 0,002384651 | G < AG | -0,147270339 |
| **cg05678033** | chr12 | 114666786 | 114666786 | 1 | + | ENSG00000139410.15 | SDSL | 1244407 | 1,46079594388841E-06 | 0,002403072 | G < AG | -0,096523361 |
| **cg11369049** | chr7 | 90690671 | 90690671 | 1 | - | ENSG00000283267.1 | FAM237B | 369362 | 1,49723886636861E-06 | 0,002436819 | G < AG | -0,098393066 |

| | | | | | | | | | | | | |
|---|---|---|---|---|---|---|---|---|---|---|---|---|
| **cg01593801** | chr1 | 170680690 | 170680690 | 1 | - | ENSG00000075945.13 | KIFAP3 | 595481 | 1,57034404077002E-06 | 0,002502555 | G < AG | -0,084285522 |
| **cg08391745** | chr16 | 86575530 | 86575530 | 1 | - | ENSG00000103248.19 | MTHFSD | 20294 | 1,6983828846242E-06 | 0,002608773 | G < AG | -0,123820254 |
| **cg10805666** | chr7 | 27225194 | 27225194 | 1 | - | ENSG00000106031.9 | HOXA13 | 25102 | 1,73740433667562E-06 | 0,002642184 | G < AG | -0,158949728 |
| **cg10802856** | chr7 | 27137435 | 27137435 | 1 | - | ENSG00000106004.5 | HOXA5 | 6247 | 1,80638862939938E-06 | 0,002709387 | G < AG | -0,171037613 |
| **cg11564083** | chr7 | 110279990 | 110279990 | 1 | + | ENSG00000173114.13 | LRRN3 | 811015 | 1,99454955429035E-06 | 0,002922836 | G < AG | -0,104078602 |
| **cg21710669** | chr16 | 72404789 | 72404789 | 1 | + | ENSG00000140829.12 | DHX38 | 311177 | 2,07557473532216E-06 | 0,003007008 | G < AG | -0,04699178 |
| **cg21535606** | chr1 | 47446268 | 47446268 | 1 | + | ENSG00000186564.6 | FOXD2 | 8225 | 2,10942741919797E-06 | 0,003023686 | G < AG | -0,204962219 |
| **cg00090182** | chr3 | 42148670 | 42148670 | 1 | + | ENSG00000182606.17 | TRAK1 | 134869 | 2,10163180399997E-06 | 0,003023686 | G < AG | -0,085724706 |
| **cg02897230** | chr4 | 173508390 | 173508390 | 1 | + | ENSG00000164105.4 | SAP30 | 138422 | 2,19578247684872E-06 | 0,00312203 | G < AG | -0,181600612 |
| **cg17521050** | chr1 | 47447081 | 47447081 | 1 | + | ENSG00000186564.6 | FOXD2 | 9038 | 2,27599265644854E-06 | 0,003194511 | G < AG | -0,159401226 |
| **cg04153838** | chr12 | 114694210 | 114694210 | 1 | + | ENSG00000139410.15 | SDSL | 1271831 | 2,28035528091521E-06 | 0,003194773 | G < AG | -0,117605217 |
| **cg17610800** | chr7 | 27241942 | 27241942 | 1 | - | ENSG00000106031.9 | HOXA13 | 41850 | 2,293203879218E-06 | 0,003201048 | G < AG | -0,158522004 |
| **cg01047586** | chr2 | 176068679 | 176068679 | 1 | - | ENSG00000174279.4 | EVX2 | 15235 | 2,4509909100391E-06 | 0,003318359 | G < AG | -0,155655436 |
| **cg14526718** | chr2 | 176068112 | 176068112 | 1 | - | ENSG00000174279.4 | EVX2 | 15802 | 2,49566686751487E-06 | 0,003355092 | G < AG | -0,199573207 |
| **cg05425114** | chr19 | 45117494 | 45117494 | 1 | + | ENSG00000104866.11 | PPP1R37 | 26099 | 2,5859630469883E-06 | 0,003440207 | G < AG | -0,115517863 |
| **cg15466224** | chr7 | 152344493 | 152344493 | 1 | - | ENSG00000055609.21 | KMT2C | 92152 | 2,66041704916242E-06 | 0,003478756 | G < AG | -0,116859796 |
| **cg27629992** | chr5 | 77959305 | 77959305 | 1 | + | ENSG00000085365.18 | SCAMP1 | 401305 | 2,72381634169995E-06 | 0,003513756 | G < AG | -0,102893012 |
| **cg20801476** | chr7 | 27241846 | 27241846 | 1 | + | ENSG00000106038.13 | EVX1 | 853 | 2,79768828117187E-06 | 0,003543174 | G < AG | -0,142720479 |
| **cg08865099** | chr7 | 27241962 | 27241962 | 1 | - | ENSG00000106031.9 | HOXA13 | 41870 | 2,99006954820462E-06 | 0,003713049 | G < AG | -0,163172975 |
| **cg23766724** | chr7 | 27133012 | 27133012 | 1 | - | ENSG00000197576.14 | HOXA4 | 2231 | 3,01072149328658E-06 | 0,003720575 | G < AG | -0,117246967 |
| **cg14513210** | chr9 | 116279505 | 116279505 | 1 | - | ENSG00000148219.18 | ASTN2 | 1135566 | 3,07179406435587E-06 | 0,00376563 | G < AG | -0,088215004 |
| **cg07328317** | chr4 | 173508721 | 173508721 | 1 | - | ENSG00000164107.9 | HAND2 | 21509 | 3,32591574897019E-06 | 0,003944406 | G < AG | -0,16186071 |
| **cg26621770** | chr12 | 114693642 | 114693642 | 1 | - | ENSG00000135111.16 | TBX3 | 9466 | 3,41844508352133E-06 | 0,003968015 | G < AG | -0,123609751 |
| **cg15683295** | chr7 | 27205923 | 27205923 | 1 | + | ENSG00000106038.13 | EVX1 | 36776 | 3,54060686718742E-06 | 0,004057533 | G < AG | -0,153771112 |
| **cg06514399** | chr1 | 85571998 | 85571998 | 1 | + | ENSG00000142871.18 | CCN1 | 8762 | 3,61095055598529E-06 | 0,004083429 | G < AG | -0,094973864 |
| **cg11958644** | chr5 | 131536729 | 131536729 | 1 | + | ENSG00000158985.14 | CDC42SE2 | 291237 | 3,68886489080339E-06 | 0,004131447 | G < AG | -0,13598372 |
| **cg24456654** | chr4 | 41752108 | 41752108 | 1 | + | ENSG00000109133.13 | TMEM33 | 183020 | 3,82873590552249E-06 | 0,004226222 | G < AG | -0,050290071 |
| **cg11362221** | chr7 | 89623249 | 89623249 | 1 | + | ENSG00000164647.9 | STEAP1 | 531206 | 3,96824811665266E-06 | 0,004302004 | G < AG | -0,091893697 |
| **cg24403057** | chr7 | 5507229 | 5507229 | 1 | + | ENSG00000075618.18 | FSCN1 | 85586 | 4,01582944330813E-06 | 0,004326603 | G < AG | -0,085799633 |
| **cg12230006** | chr8 | 16461911 | 16461911 | 1 | - | ENSG00000038945.15 | MSR1 | 105580 | 4,10472216657322E-06 | 0,004409952 | G < AG | -0,076743563 |
| **cg23930856** | chr6 | 50844011 | 50844011 | 1 | - | ENSG00000180872.7 | DEFB112 | 794081 | 4,11222490693289E-06 | 0,004411816 | G < AG | -0,107096026 |

| | | | | | | | | | | | | |
|---|---|---|---|---|---|---|---|---|---|---|---|---|
| **cg20284629** | chr1 | 47445347 | 47445347 | 1 | - | ENSG00000123473.17 | STIL | 130454 | 4,14999872679412E-06 | 0,004438069 | G < AG | -0,105584499 |
| **cg07937443** | chr4 | 173521392 | 173521392 | 1 | - | ENSG00000164107.9 | HAND2 | 8838 | 4,28292647881283E-06 | 0,004499124 | G < AG | -0,079287321 |
| **cg08162910** | chr5 | 77957939 | 77957939 | 1 | + | ENSG00000085365.18 | SCAMP1 | 402671 | 4,42616741744984E-06 | 0,004593994 | G < AG | -0,100295446 |
| **cg09829551** | chr16 | 86565505 | 86565505 | 1 | - | ENSG00000103248.19 | MTHFSD | 10269 | 4,57966227644987E-06 | 0,004693305 | G < AG | -0,129113563 |
| **cg17712241** | chr6 | 165686826 | 165686826 | 1 | - | ENSG00000112541.18 | PDE10A | 301253 | 4,68869672292976E-06 | 0,004757007 | G < AG | -0,088732193 |
| **cg06894970** | chr4 | 101924360 | 101924360 | 1 | + | ENSG00000153064.12 | BANK1 | 513075 | 4,70281666173599E-06 | 0,004758459 | G < AG | -0,076538888 |
| **cg01572694** | chr17 | 48580193 | 48580193 | 1 | - | ENSG00000182742.6 | HOXB4 | 1842 | 4,70163325513651E-06 | 0,004758459 | G < AG | -0,14394884 |
| **cg17603788** | chr12 | 45908160 | 45908160 | 1 | - | ENSG00000139218.18 | SCAF11 | 83961 | 4,74587979233474E-06 | 0,004788182 | G < AG | -0,136749929 |
| **cg13972343** | chr2 | 176067421 | 176067421 | 1 | + | ENSG00000128714.6 | HOXD13 | 25299 | 4,76782161470394E-06 | 0,004796299 | G < AG | -0,145862865 |
| **cg06502892** | chr7 | 103432812 | 103432812 | 1 | - | ENSG00000170615.15 | SLC26A5 | 13396 | 4,77156417742127E-06 | 0,004796299 | G < AG | -0,14724724 |
| **cg09935792** | chr17 | 9159412 | 9159412 | 1 | - | ENSG00000141506.15 | PIK3R5 | 193699 | 4,79840827998123E-06 | 0,00480251 | G < AG | -0,067559316 |
| **cg18741219** | chr13 | 50992471 | 50992471 | 1 | + | ENSG00000136104.21 | RNASEH2B | 82725 | 5,12993385147183E-06 | 0,005011643 | G < AG | -0,123377324 |
| **cg06284231** | chr14 | 38255050 | 38255050 | 1 | + | ENSG00000139874.6 | SSTR1 | 47147 | 5,26530220028937E-06 | 0,005104773 | G < AG | -0,103412904 |
| **cg18949192** | chr12 | 114696475 | 114696475 | 1 | + | ENSG00000139410.15 | SDSL | 1274096 | 5,33581108876853E-06 | 0,005134089 | G < AG | -0,121732753 |
| **cg02998883** | chr4 | 169555947 | 169555947 | 1 | + | ENSG00000109572.14 | CLCN3 | 56685 | 5,34888167371503E-06 | 0,0051402 | G < AG | -0,092154062 |
| **cg07625849** | chr17 | 48580616 | 48580616 | 1 | - | ENSG00000182742.6 | HOXB4 | 2265 | 5,40305376792223E-06 | 0,005161327 | G < AG | -0,1534276 |
| **cg18483404** | chr10 | 122832216 | 122832216 | 1 | + | ENSG00000203795.3 | FAM24A | 78393 | 5,8395840207577E-06 | 0,005433804 | G < AG | -0,121550587 |
| **cg06674986** | chr7 | 89622031 | 89622031 | 1 | + | ENSG00000164647.9 | STEAP1 | 532424 | 5,84620066664164E-06 | 0,005433804 | G < AG | -0,082301452 |
| **cg14928241** | chr7 | 122855261 | 122855261 | 1 | - | ENSG00000081803.16 | CADPS2 | 31499 | 6,04973740894684E-06 | 0,005544991 | G < AG | -0,079400963 |
| **cg09731313** | chr2 | 212968853 | 212968853 | 1 | + | ENSG00000144451.19 | SPAG16 | 315525 | 6,14342505728924E-06 | 0,005614553 | G < AG | -0,067461193 |
| **cg27470066** | chr17 | 61408418 | 61408418 | 1 | - | ENSG00000253506.3 | NACA2 | 182802 | 6,15555341188781E-06 | 0,005618924 | G < AG | -0,105535944 |
| **cg25851147** | chr21 | 37862703 | 37862703 | 1 | - | ENSG00000157542.11 | KCNJ6 | 258643 | 6,21054124021521E-06 | 0,005648896 | G < AG | -0,111861025 |
| **cg13936452** | chr1 | 47445664 | 47445664 | 1 | + | ENSG00000186564.6 | FOXD2 | 7621 | 6,28241858085065E-06 | 0,005687223 | G < AG | -0,162389656 |
| **cg01913455** | chr17 | 61401709 | 61401709 | 1 | - | ENSG00000253506.3 | NACA2 | 189511 | 6,31942628827313E-06 | 0,005713963 | G < AG | -0,119773034 |
| **cg16196175** | chr7 | 27249501 | 27249501 | 1 | - | ENSG00000106031.9 | HOXA13 | 49409 | 6,37107600318982E-06 | 0,005733383 | G < AG | -0,09783703 |
| **cg07955762** | chr13 | 110421913 | 110421913 | 1 | - | ENSG00000187498.16 | COL4A1 | 114755 | 6,47556815285006E-06 | 0,005782057 | G < AG | -0,15300006 |
| **cg04197548** | chr10 | 73245187 | 73245187 | 1 | + | ENSG00000138286.15 | FAM149B1 | 77069 | 6,92738459279117E-06 | 0,005969389 | G < AG | -0,089618462 |
| **cg04528477** | chr2 | 176067730 | 176067730 | 1 | + | ENSG00000128714.6 | HOXD13 | 24990 | 7,00242477000676E-06 | 0,005979986 | G < AG | -0,221076863 |
| **cg25945246** | chr21 | 44581116 | 44581116 | 1 | + | ENSG00000215454.6 | KRTAP10-4 | 7393 | 7,28327243133473E-06 | 0,006101412 | G < AG | -0,107671598 |
| **cg01692978** | chr16 | 86827168 | 86827168 | 1 | - | ENSG00000103248.19 | MTHFSD | 271932 | 7,35687303878394E-06 | 0,006123901 | G < AG | -0,087480728 |
| **cg10192138** | chr1 | 68378842 | 68378842 | 1 | - | ENSG00000116745.7 | RPE65 | 71113 | 7,71516900972822E-06 | 0,006325486 | G < AG | -0,037877637 |

| | | | | | | | | | | | | |
|---|---|---|---|---|---|---|---|---|---|---|---|---|
| **cg24210813** | chr17 | 48592282 | 48592282 | 1 | + | ENSG00000229637.4 | PRAC2 | 130902 | 7,84596601212247E-06 | 0,006378035 | G < AG | -0,16331839 |
| **cg22274074** | chr7 | 27181220 | 27181220 | 1 | - | ENSG00000253293.5 | HOXA10 | 958 | 7,89296992128599E-06 | 0,006401541 | G < AG | -0,133382387 |
| **cg11910375** | chr7 | 27155279 | 27155279 | 1 | + | ENSG00000106038.13 | EVX1 | 87420 | 8,08634774660359E-06 | 0,006490671 | G < AG | -0,148640024 |
| **cg01121566** | chr7 | 27206182 | 27206182 | 1 | - | ENSG00000106031.9 | HOXA13 | 6090 | 8,31222492058487E-06 | 0,006600293 | G < AG | -0,183467995 |
| **cg18148179** | chr3 | 181710510 | 181710510 | 1 | + | ENSG00000181449.4 | SOX2 | 1414 | 8,26598379828759E-06 | 0,006600293 | G < AG | -0,126065446 |
| **cg03352153** | chr6 | 63320524 | 63320524 | 1 | - | ENSG00000146166.17 | LGSN | 540 | 8,67063695793053E-06 | 0,006726303 | G < AG | -0,086241993 |
| **cg00479745** | chr14 | 106874586 | 106874586 | 1 | + | ENSG00000184986.11 | TMEM121 | 1348004 | 8,69168264196455E-06 | 0,006726303 | G < AG | -0,098444772 |
| **cg13148429** | chr18 | 49161754 | 49161754 | 1 | - | ENSG00000101665.10 | SMAD7 | 210788 | 8,72088266364304E-06 | 0,006734431 | G < AG | -0,137268841 |
| **cg05516617** | chr7 | 27182070 | 27182070 | 1 | - | ENSG00000253293.5 | HOXA10 | 1808 | 8,80199851037998E-06 | 0,006766877 | G < AG | -0,096195559 |
| **cg04681554** | chr7 | 27243790 | 27243790 | 1 | + | ENSG00000106038.13 | EVX1 | 1091 | 8,85962523656284E-06 | 0,006779583 | G < AG | -0,076460473 |
| **cg09574499** | chr7 | 27129343 | 27129343 | 1 | + | ENSG00000106038.13 | EVX1 | 113356 | 8,91077568043019E-06 | 0,006789095 | G < AG | -0,083800428 |
| **cg23512098** | chr17 | 72479902 | 72479902 | 1 | - | ENSG00000133195.11 | SLC39A11 | 612811 | 9,01556426370455E-06 | 0,00684847 | G < AG | -0,0758238 |
| **cg24757937** | chr14 | 60644840 | 60644840 | 1 | + | ENSG00000020426.11 | MNAT1 | 89901 | 9,18863900469587E-06 | 0,006918112 | G < AG | -0,101291792 |
| **cg06217451** | chr1 | 108079741 | 108079741 | 1 | + | ENSG00000186086.18 | NBPF6 | 370540 | 9,45269944471183E-06 | 0,007056526 | G < AG | -0,108505915 |
| **cg20392696** | chr7 | 116479121 | 116479121 | 1 | + | ENSG00000105974.13 | CAV1 | 45872 | 9,52560954746608E-06 | 0,007077665 | G < AG | -0,083958444 |
| **cg06014207** | chr3 | 194579300 | 194579300 | 1 | + | ENSG00000185112.6 | FAM43A | 106582 | 9,79826428110634E-06 | 0,007205937 | G < AG | -0,0627051 |
| **cg23373556** | chr7 | 18506288 | 18506288 | 1 | - | ENSG00000229937.9 | PRPS1L1 | 478441 | 9,83224871605994E-06 | 0,007205937 | G < AG | -0,10015736 |
| **cg24628013** | chr6 | 29612590 | 29612590 | 1 | - | ENSG00000204681.11 | GABBR1 | 21387 | 9,86499801658929E-06 | 0,00720743 | G < AG | -0,034500611 |
| **cg10566978** | chr11 | 34433097 | 34433097 | 1 | - | ENSG00000166016.6 | ABTB2 | 75086 | 9,96991326945686E-06 | 0,007242577 | G < AG | -0,079564417 |
| **cg05612778** | chr3 | 142022710 | 142022710 | 1 | + | ENSG00000069849.11 | ATP1B3 | 146587 | 1,00410254346251E-05 | 0,007266632 | G < AG | -0,10626216 |
| **cg01639409** | chr1 | 177128988 | 177128988 | 1 | - | ENSG00000152092.16 | ASTN1 | 35986 | 1,00707648881525E-05 | 0,007273632 | G < AG | -0,123086799 |
| **cg23555848** | chr18 | 51152479 | 51152479 | 1 | - | ENSG00000176624.12 | MEX3C | 65855 | 1,01868560154818E-05 | 0,007323668 | G < AG | -0,131602888 |
| **cg05986745** | chr13 | 30453744 | 30453744 | 1 | + | ENSG00000236444.5 | UBE2L5 | 31257 | 1,02922911909348E-05 | 0,00737327 | G < AG | -0,067707311 |
| **cg09683440** | chr16 | 83836322 | 83836322 | 1 | + | ENSG00000230989.7 | HSBP1 | 28345 | 1,04040277254247E-05 | 0,00742398 | G < AG | -0,134294516 |
| **cg16598356** | chr9 | 132380065 | 132380065 | 1 | + | ENSG00000188523.9 | CFAP77 | 29977 | 1,05020000062972E-05 | 0,00745218 | G < AG | -0,039735578 |
| **cg00116234** | chr9 | 18474245 | 18474245 | 1 | + | ENSG00000178031.18 | ADAMTSL1 | 567683 | 1,05451894598644E-05 | 0,007467948 | G < AG | -0,135796 |
| **cg25674620** | chr4 | 150209702 | 150209702 | 1 | + | ENSG00000170390.16 | DCLK2 | 131258 | 1,09032666355165E-05 | 0,007616915 | G < AG | -0,084039057 |
| **cg11417123** | chr2 | 201396924 | 201396924 | 1 | - | ENSG00000155749.13 | FLACC1 | 39525 | 1,09387269381107E-05 | 0,007621747 | G < AG | -0,051446048 |
| **cg21848211** | chr12 | 26156187 | 26156187 | 1 | + | ENSG00000123096.12 | SSPN | 34197 | 1,1181096744066E-05 | 0,007740201 | G < AG | -0,077969575 |
| **cg06933370** | chr15 | 37095322 | 37095322 | 1 | + | ENSG00000186073.14 | CDIN1 | 515697 | 1,12407869556239E-05 | 0,007748341 | G < AG | -0,127378658 |
| **cg15798385** | chr7 | 27241950 | 27241950 | 1 | - | ENSG00000106031.9 | HOXA13 | 41858 | 1,13694009940759E-05 | 0,007813984 | G < AG | -0,144979373 |

| | | | | | | | | | | | | |
|---|---|---|---|---|---|---|---|---|---|---|---|---|
| **cg09757709** | chr11 | 93731555 | 93731555 | 1 | + | ENSG00000182919.15 | C11orf54 | 10035 | 1,13930515140154E-05 | 0,007816193 | G < AG | -0,056024931 |
| **cg01541645** | chr14 | 60637611 | 60637611 | 1 | - | ENSG00000126778.12 | SIX1 | 20649 | 1,18525564788724E-05 | 0,007988151 | G < AG | -0,074815454 |
| **cg24588381** | chr13 | 27977008 | 27977008 | 1 | + | ENSG00000139515.6 | PDX1 | 57009 | 1,19774265813698E-05 | 0,008046864 | G < AG | -0,110473882 |
| **cg09704116** | chr17 | 48589596 | 48589596 | 1 | - | ENSG00000120075.6 | HOXB5 | 4184 | 1,22256305828407E-05 | 0,008146283 | G < AG | -0,155366196 |
| **cg02658990** | chr2 | 150981978 | 150981978 | 1 | - | ENSG00000184898.7 | RBM43 | 279886 | 1,22991146393501E-05 | 0,008188115 | G < AG | -0,137561498 |
| **cg26514961** | chr12 | 94173008 | 94173008 | 1 | + | ENSG00000136040.9 | PLXNC1 | 24432 | 1,24453031969587E-05 | 0,008242398 | G < AG | -0,13135431 |
| **cg00924017** | chr1 | 109643405 | 109643405 | 1 | + | ENSG00000168765.17 | GSTM4 | 12693 | 1,28073062424784E-05 | 0,008381543 | G < AG | -0,12170941 |
| **cg22781048** | chr17 | 61555241 | 61555241 | 1 | - | ENSG00000253506.3 | NACA2 | 35979 | 1,29142741351014E-05 | 0,008428931 | G < AG | -0,072507306 |
| **cg16551732** | chr4 | 168877936 | 168877936 | 1 | - | ENSG00000145439.12 | CBR4 | 132340 | 1,29307924161594E-05 | 0,008432517 | G < AG | -0,161631299 |
| **cg22610399** | chr2 | 38723088 | 38723088 | 1 | - | ENSG00000115875.19 | SRSF7 | 28407 | 1,30455680466803E-05 | 0,008492885 | G < AG | -0,083509901 |
| **cg26674479** | chr7 | 27241796 | 27241796 | 1 | + | ENSG00000106038.13 | EVX1 | 903 | 1,32409873993006E-05 | 0,008548902 | G < AG | -0,095527852 |
| **cg21773724** | chr14 | 60658459 | 60658459 | 1 | + | ENSG00000020426.11 | MNAT1 | 76282 | 1,32457005667171E-05 | 0,008548902 | G < AG | -0,120684227 |
| **cg03161037** | chr17 | 48730947 | 48730947 | 1 | + | ENSG00000229637.4 | PRAC2 | 7763 | 1,32722557427803E-05 | 0,008548902 | G < AG | -0,075791223 |
| **cg02132714** | chr17 | 48579328 | 48579328 | 1 | - | ENSG00000182742.6 | HOXB4 | 977 | 1,35143441772558E-05 | 0,008629315 | G < AG | -0,122637353 |
| **cg12284098** | chr8 | 2045778 | 2045778 | 1 | + | ENSG00000036448.10 | MYOM2 | 733 | 1,3525876453562E-05 | 0,008629315 | G < AG | -0,105297222 |
| **cg12999453** | chr17 | 61404958 | 61404958 | 1 | - | ENSG00000253506.3 | NACA2 | 186262 | 1,39346468156855E-05 | 0,008799735 | G < AG | -0,089888539 |
| **cg05365255** | chr7 | 32155543 | 32155543 | 1 | + | ENSG00000105778.19 | AVL9 | 339882 | 1,4293899816238E-05 | 0,008897902 | G < AG | -0,107725381 |
| **cg17818338** | chr12 | 76158625 | 76158625 | 1 | - | ENSG00000187109.15 | NAP1L1 | 73889 | 1,50088629832976E-05 | 0,009119104 | G < AG | -0,111962252 |
| **cg00507008** | chr12 | 114693616 | 114693616 | 1 | - | ENSG00000135111.16 | TBX3 | 9440 | 1,51923213473274E-05 | 0,009201336 | G < AG | -0,124795086 |
| **cg01774894** | chr17 | 48597033 | 48597033 | 1 | - | ENSG00000120075.6 | HOXB5 | 3253 | 1,54256928999748E-05 | 0,009299402 | G < AG | -0,153795393 |
| **cg14943837** | chr22 | 30308829 | 30308829 | 1 | + | ENSG00000187860.11 | CCDC157 | 47805 | 1,55732691169678E-05 | 0,009321355 | G < AG | -0,128827847 |
| **cg01729108** | chr12 | 121257787 | 121257787 | 1 | + | ENSG00000135124.15 | P2RX4 | 47927 | 1,56121905275957E-05 | 0,009337345 | G < AG | -0,081297556 |
| **cg05574357** | chr12 | 114696557 | 114696557 | 1 | + | ENSG00000139410.15 | SDSL | 1274178 | 1,58213374817668E-05 | 0,009400944 | G < AG | -0,130444712 |
| **cg07498531** | chr1 | 40949190 | 40949190 | 1 | - | ENSG00000171790.15 | SLFNL1 | 74048 | 1,58291363649668E-05 | 0,009400944 | G < AG | -0,081609778 |
| **cg22841821** | chr17 | 66677731 | 66677731 | 1 | - | ENSG00000091583.11 | APOH | 421205 | 1,59798907955211E-05 | 0,009446472 | G < AG | -0,057452364 |
| **cg11112560** | chr5 | 73343945 | 73343945 | 1 | + | ENSG00000145741.16 | BTF3 | 154462 | 1,64708856698046E-05 | 0,0096178 | G < AG | -0,056694025 |
| **cg26723054** | chr16 | 85616916 | 85616916 | 1 | - | ENSG00000131153.9 | GINS2 | 73158 | 1,6522384846079E-05 | 0,009634972 | G < AG | -0,103086711 |
| **cg22741264** | chr12 | 114667714 | 114667714 | 1 | - | ENSG00000135111.16 | TBX3 | 16462 | 1,66908665538325E-05 | 0,009687095 | G < AG | -0,121232819 |
| **cg07768763** | chr16 | 81082009 | 81082009 | 1 | + | ENSG00000166454.10 | ATMIN | 46168 | 1,73607848304223E-05 | 0,009918654 | G < AG | -0,114565771 |
| **cg19517864** | chr20 | 32064966 | 32064966 | 1 | - | ENSG00000088356.6 | PDRG1 | 112919 | 1,74075834605603E-05 | 0,009929784 | G < AG | -0,048988204 |
| **cg14097408** | chr9 | 109954189 | 109954189 | 1 | - | ENSG00000188959.10 | C9orf152 | 253971 | 1,7512249210731E-05 | 0,00995978 | G < AG | -0,075430327 |

| | | | | | | | | | | | | |
|---|---|---|---|---|---|---|---|---|---|---|---|---|
| **cg22567585** | chr15 | 36888886 | 36888886 | 1 | + | ENSG00000186073.14 | CDIN1 | 309261 | 1,75767544780431E-05 | 0,009985416 | G < AG | -0,081131441 |
| **cg22890023** | chr17 | 71276380 | 71276380 | 1 | + | ENSG00000125398.8 | SOX9 | 844639 | 5,51649394358124E-12 | 2,13470367361062E-06 | G > AG | 0,119383272 |
| **cg14615547** | chr11 | 101687015 | 101687015 | 1 | - | ENSG00000137672.13 | TRPC6 | 185548 | 5,5813273055916E-12 | 2,13470367361062E-06 | G > AG | 0,171129081 |
| **cg26591377** | chrX | 1017215 | 1017215 | 1 | + | ENSG00000198223.17 | CSF2RA | 251584 | 2,15380439465851E-11 | 4,5632340549136E-06 | G > AG | 0,10110753 |
| **cg04569855** | chr3 | 99955414 | 99955414 | 1 | - | ENSG00000168386.19 | FILIP1L | 159100 | 2,3861768869697E-11 | 4,5632340549136E-06 | G > AG | 0,186092311 |
| **cg19017635** | chr13 | 95100805 | 95100805 | 1 | - | ENSG00000125257.16 | ABCC4 | 200671 | 4,32247415244369E-11 | 5,90768584668151E-06 | G > AG | 0,133373875 |
| **cg07638843** | chr14 | 57982303 | 57982303 | 1 | - | ENSG00000151812.15 | SLC35F4 | 108 | 4,63381340509489E-11 | 5,90768584668151E-06 | G > AG | 0,13657824 |
| **cg08324750** | chr16 | 77272101 | 77272101 | 1 | + | ENSG00000205078.6 | SYCE1L | 72694 | 6,0396114476707E-11 | 6,59995631792094E-06 | G > AG | 0,134620031 |
| **cg15147534** | chr6 | 48247905 | 48247905 | 1 | - | ENSG00000244694.8 | PTCHD4 | 136707 | 1,13292926302579E-10 | 1,08328544775505E-05 | G > AG | 0,09699643 |
| **cg17528218** | chr2 | 77519923 | 77519923 | 1 | - | ENSG00000176204.14 | LRRTM4 | 73397 | 1,39394575562507E-10 | 1,1847684101815E-05 | G > AG | 0,099421791 |
| **cg01295999** | chr3 | 175208544 | 175208544 | 1 | + | ENSG00000177694.16 | NAALADL2 | 769972 | 1,95938669817242E-10 | 1,49882268270561E-05 | G > AG | 0,117818355 |
| **cg19017920** | chr13 | 95128368 | 95128368 | 1 | + | ENSG00000134873.10 | CLDN10 | 305235 | 2,49237802883857E-10 | 1,73321057645449E-05 | G > AG | 0,143437354 |
| **cg09989818** | chr4 | 74197257 | 74197257 | 1 | - | ENSG00000081041.9 | CXCL2 | 98060 | 5,61250989896208E-10 | 3,57771692511421E-05 | G > AG | 0,156483882 |
| **cg22889444** | chr13 | 107022091 | 107022091 | 1 | - | ENSG00000134884.15 | ARGLU1 | 453953 | 7,85401250588426E-10 | 3,57920389221606E-05 | G > AG | 0,100934024 |
| **cg08207013** | chr5 | 89232528 | 89232528 | 1 | - | ENSG00000081189.16 | MEF2C | 328270 | 7,62245348290471E-10 | 3,57920389221606E-05 | G > AG | 0,131487047 |
| **cg17836965** | chr12 | 79086706 | 79086706 | 1 | - | ENSG00000177425.11 | PAWR | 604259 | 6,50894115343883E-10 | 3,57920389221606E-05 | G > AG | 0,127238204 |
| **cg25412357** | chr13 | 111858148 | 111858148 | 1 | + | ENSG00000182968.5 | SOX1 | 209000 | 6,7525876093817E-10 | 3,57920389221606E-05 | G > AG | 0,100134653 |
| **cg18152517** | chr7 | 135231004 | 135231004 | 1 | - | ENSG00000105875.15 | WDR91 | 19448 | 7,95436029994707E-10 | 3,57920389221606E-05 | G > AG | 0,094768907 |
| **cg06358636** | chr6 | 133159691 | 133159691 | 1 | + | ENSG00000112319.20 | EYA4 | 80822 | 1,01304031600582E-09 | 4,30511072543773E-05 | G > AG | 0,112078421 |
| **cg19645721** | chr18 | 21367303 | 21367303 | 1 | + | ENSG00000141449.15 | GREB1L | 125062 | 1,10109008821632E-09 | 4,43301656171091E-05 | G > AG | 0,174468273 |
| **cg14216672** | chr1 | 3349359 | 3349359 | 1 | - | ENSG00000162591.16 | MEGF6 | 262150 | 1,22182870833407E-09 | 4,45327286792618E-05 | G > AG | 0,145184444 |
| **cg12365177** | chr7 | 117277880 | 117277880 | 1 | - | ENSG00000105989.10 | WNT2 | 45273 | 1,22255526387487E-09 | 4,45327286792618E-05 | G > AG | 0,10124424 |
| **cg24969528** | chr20 | 10741299 | 10741299 | 1 | + | ENSG00000149346.15 | SLX4IP | 305995 | 1,43835029738217E-09 | 5,00117542207458E-05 | G > AG | 0,127895887 |
| **cg05953296** | chr12 | 130027148 | 130027148 | 1 | + | ENSG00000111432.5 | FZD10 | 135310 | 1,58456514622015E-09 | 5,27002122785956E-05 | G > AG | 0,143244318 |
| **cg12830291** | chr14 | 80974095 | 80974095 | 1 | - | ENSG00000100629.17 | CEP128 | 14577 | 2,01361910956435E-09 | 6,16122993701969E-05 | G > AG | 0,143993974 |
| **cg05106567** | chr9 | 32742018 | 32742018 | 1 | + | ENSG00000188133.6 | TMEM215 | 41521 | 1,9634378828111E-09 | 6,16122993701969E-05 | G > AG | 0,167656013 |
| **cg11132229** | chr8 | 133952755 | 133952755 | 1 | + | ENSG00000104415.14 | CCN4 | 761717 | 2,31128003014811E-09 | 6,800006385243E-05 | G > AG | 0,114142225 |
| **cg03404911** | chr2 | 120453528 | 120453528 | 1 | + | ENSG00000163083.6 | INHBB | 107393 | 2,4151659461195E-09 | 6,84247648987699E-05 | G > AG | 0,08497912 |
| **cg03779078** | chr7 | 45514820 | 45514820 | 1 | + | ENSG00000164742.16 | ADCY1 | 59319 | 2,74079702213793E-09 | 7,38815350832312E-05 | G > AG | 0,145664868 |
| **cg06414254** | chr4 | 164199205 | 164199205 | 1 | + | ENSG00000248771.5 | SMIM31 | 554858 | 2,80094000700494E-09 | 7,38815350832312E-05 | G > AG | 0,137246048 |
| **cg13804685** | chr14 | 23876751 | 23876751 | 1 | - | ENSG00000129535.13 | NRL | 238260 | 3,43859549090396E-09 | 8,48495409504716E-05 | G > AG | 0,150628685 |

| | | | | | | | | | | | | |
|---|---|---|---|---|---|---|---|---|---|---|---|---|
| **cg04981423** | chr6 | 153215004 | 153215004 | 1 | - | ENSG00000091844.8 | RGS17 | 83721 | 3,39817438468495E-09 | 8,48495409504716E-05 | G > AG | 0,101942928 |
| **cg03353172** | chr20 | 17735360 | 17735360 | 1 | + | ENSG00000125888.15 | BANF2 | 41689 | 3,92236699076801E-09 | 9,37623208377892E-05 | G > AG | 0,158676342 |
| **cg04196458** | chr21 | 42449113 | 42449113 | 1 | - | ENSG00000160188.10 | RSPH1 | 47134 | 4,34808850190079E-09 | 9,78249331364004E-05 | G > AG | 0,142149731 |
| **cg06755262** | chr6 | 14824206 | 14824206 | 1 | - | ENSG00000047579.20 | DTNBP1 | 838853 | 5,72425223411245E-09 | 0,000122695 | G > AG | 0,096384285 |
| **cg18239109** | chr5 | 126054933 | 126054933 | 1 | + | ENSG00000155324.10 | GRAMD2B | 305198 | 6,09510779295764E-09 | 0,000122695 | G > AG | 0,108766358 |
| **cg16186064** | chr22 | 36856059 | 36856059 | 1 | + | ENSG00000100365.16 | NCF4 | 4928 | 6,06588091307365E-09 | 0,000122695 | G > AG | 0,119729368 |
| **cg06372353** | chr4 | 146174483 | 146174483 | 1 | - | ENSG00000151612.18 | ZNF827 | 235659 | 5,7809711173839E-09 | 0,000122695 | G > AG | 0,171795721 |
| **cg21568669** | chr18 | 21369243 | 21369243 | 1 | - | ENSG00000141446.11 | ESCO1 | 231642 | 6,30282046617588E-09 | 0,000123623 | G > AG | 0,136161521 |
| **cg08081757** | chr22 | 30878251 | 30878251 | 1 | + | ENSG00000253352.10 | TUG1 | 90993 | 6,57937010945793E-09 | 0,000125821 | G > AG | 0,087077063 |
| **cg08180229** | chr2 | 5982682 | 5982682 | 1 | + | ENSG00000176887.7 | SOX11 | 290299 | 7,08074070526516E-09 | 0,000132107 | G > AG | 0,080133457 |
| **cg24681960** | chr4 | 160300109 | 160300109 | 1 | + | ENSG00000109756.9 | RAPGEF2 | 1197097 | 7,48158723554922E-09 | 0,000133948 | G > AG | 0,102760787 |
| **cg00106273** | chr12 | 65609495 | 65609495 | 1 | - | ENSG00000156076.10 | WIF1 | 488189 | 7,52965940127218E-09 | 0,000133948 | G > AG | 0,121079705 |
| **cg05461178** | chr18 | 75132327 | 75132327 | 1 | - | ENSG00000180011.7 | ZADH2 | 76813 | 8,02755912275272E-09 | 0,00013956 | G > AG | 0,15154292 |
| **cg09667837** | chr6 | 67677077 | 67677077 | 1 | - | ENSG00000188107.15 | EYS | 1969850 | 8,73922900120063E-09 | 0,000147035 | G > AG | 0,125336501 |
| **cg11152513** | chr2 | 168901241 | 168901241 | 1 | + | ENSG00000152254.11 | G6PC2 | 49 | 8,84192938112743E-09 | 0,000147035 | G > AG | 0,131092972 |
| **cg12369028** | chr18 | 25815824 | 25815824 | 1 | + | ENSG00000154611.15 | PSMA8 | 318027 | 1,02953722991513E-08 | 0,000167562 | G > AG | 0,082878855 |
| **cg12363133** | chr17 | 9798607 | 9798607 | 1 | - | ENSG00000184544.12 | DHRS7C | 7014 | 1,09827329495762E-08 | 0,000172075 | G > AG | 0,110533178 |
| **cg22533771** | chr17 | 41530796 | 41530796 | 1 | + | ENSG00000173812.11 | EIF1 | 158088 | 1,10734925033948E-08 | 0,000172075 | G > AG | 0,093969639 |
| **cg15700583** | chr5 | 82552482 | 82552482 | 1 | + | ENSG00000152422.16 | XRCC4 | 525015 | 1,12475736813247E-08 | 0,000172075 | G > AG | 0,119446945 |
| **cg00422932** | chr11 | 3380770 | 3380770 | 1 | + | ENSG00000129744.3 | ART1 | 264357 | 1,22572485205571E-08 | 0,000175038 | G > AG | 0,103577708 |
| **cg19225008** | chr13 | 113882482 | 113882482 | 1 | - | ENSG00000183087.15 | GAS6 | 18405 | 1,19784648488414E-08 | 0,000175038 | G > AG | 0,16697666 |
| **cg21959330** | chr4 | 44157644 | 44157644 | 1 | - | ENSG00000183783.7 | KCTD8 | 291166 | 1,27172793340965E-08 | 0,000175038 | G > AG | 0,123134744 |
| **cg08803700** | chr16 | 57239381 | 57239381 | 1 | - | ENSG00000102934.10 | PLLP | 45292 | 1,28141475825055E-08 | 0,000175038 | G > AG | 0,162161599 |
| **cg08540973** | chr13 | 45853148 | 45853148 | 1 | - | ENSG00000215475.5 | SIAH3 | 1394 | 1,20097704430609E-08 | 0,000175038 | G > AG | 0,111010655 |
| **cg08418664** | chr22 | 25660548 | 25660548 | 1 | + | ENSG00000133454.16 | MYO18B | 81595 | 1,41999734611141E-08 | 0,000190565 | G > AG | 0,154672477 |
| **cg17509180** | chr21 | 16305946 | 16305946 | 1 | + | ENSG00000155313.16 | USP25 | 575965 | 1,55940059914205E-08 | 0,000205665 | G > AG | 0,118457967 |
| **cg13309826** | chr17 | 10533819 | 10533819 | 1 | + | ENSG00000170222.12 | ADPRM | 163774 | 1,6194455000835E-08 | 0,000208849 | G > AG | 0,087420812 |
| **cg25851417** | chr12 | 91001614 | 91001614 | 1 | - | ENSG00000083782.8 | EPYC | 3413 | 1,6381528631108E-08 | 0,000208849 | G > AG | 0,185148148 |
| **cg09572449** | chr6 | 51474659 | 51474659 | 1 | - | ENSG00000170927.15 | PKHD1 | 612955 | 1,7134206483306E-08 | 0,000213068 | G > AG | 0,118552722 |
| **cg00284731** | chr3 | 78765721 | 78765721 | 1 | - | ENSG00000169855.20 | ROBO1 | 1002278 | 1,72695170005024E-08 | 0,000213068 | G > AG | 0,189242485 |
| **cg00689835** | chr2 | 168901108 | 168901108 | 1 | + | ENSG00000152254.11 | G6PC2 | 182 | 1,905814724007E-08 | 0,000224973 | G > AG | 0,114586384 |

| | | | | | | | | | | | | |
|---|---|---|---|---|---|---|---|---|---|---|---|---|
| **cg24856447** | chr12 | 75848675 | 75848675 | 1 | + | ENSG00000139278.10 | GLIPR1 | 367923 | 1,87514459312667E-08 | 0,000224973 | G > AG | 0,16876546 |
| **cg11814946** | chr2 | 160254582 | 160254582 | 1 | - | ENSG00000115221.12 | ITGB6 | 54268 | 1,91167175186488E-08 | 0,000224973 | G > AG | 0,118628711 |
| **cg23975840** | chr12 | 116605090 | 116605090 | 1 | + | ENSG00000258102.5 | MAP1LC3B2 | 56986 | 2,07174972280966E-08 | 0,000228662 | G > AG | 0,065088287 |
| **cg20959675** | chr4 | 98728359 | 98728359 | 1 | + | ENSG00000164024.12 | METAP1 | 267299 | 2,09247951108913E-08 | 0,000228662 | G > AG | 0,100449491 |
| **cg20850544** | chr15 | 96020420 | 96020420 | 1 | + | ENSG00000185551.15 | NR2F2 | 305517 | 1,99458191927038E-08 | 0,000228662 | G > AG | 0,094726056 |
| **cg21255147** | chr10 | 3503406 | 3503406 | 1 | + | ENSG00000067057.18 | PFKP | 437074 | 2,01408409989905E-08 | 0,000228662 | G > AG | 0,102109971 |
| **cg12536880** | chr15 | 91295934 | 91295934 | 1 | + | ENSG00000185518.12 | SV2B | 195985 | 2,0800018025106E-08 | 0,000228662 | G > AG | 0,113137844 |
| **cg00422192** | chr2 | 13022407 | 13022407 | 1 | + | ENSG00000071575.12 | TRIB2 | 305498 | 2,16413018535622E-08 | 0,000233161 | G > AG | 0,113061838 |
| **cg22890018** | chr17 | 71276150 | 71276150 | 1 | + | ENSG00000125398.8 | SOX9 | 844869 | 2,22755231366331E-08 | 0,000236604 | G > AG | 0,1470486 |
| **cg12939519** | chr6 | 35162449 | 35162449 | 1 | - | ENSG00000124678.20 | TCP11 | 13838 | 2,25795602218675E-08 | 0,000236604 | G > AG | 0,156678761 |
| **cg05242561** | chr15 | 82853385 | 82853385 | 1 | - | ENSG00000186628.12 | FSD2 | 47314 | 2,30413430754716E-08 | 0,00023818 | G > AG | 0,087585361 |
| **cg06780190** | chr4 | 84319337 | 84319337 | 1 | + | ENSG00000163624.6 | CDS1 | 263789 | 2,43102171475083E-08 | 0,000246494 | G > AG | 0,084116865 |
| **cg26892353** | chr3 | 181329368 | 181329368 | 1 | - | ENSG00000205981.8 | DNAJC19 | 339593 | 2,60095334681794E-08 | 0,000248698 | G > AG | 0,146603124 |
| **cg21184428** | chr2 | 160140537 | 160140537 | 1 | - | ENSG00000115221.12 | ITGB6 | 59777 | 2,594776059723E-08 | 0,000248698 | G > AG | 0,080013168 |
| **cg03668686** | chr1 | 213966379 | 213966379 | 1 | + | ENSG00000117707.16 | PROX1 | 16801 | 2,54859185756962E-08 | 0,000248698 | G > AG | 0,10650689 |
| **cg13754514** | chr9 | 72578441 | 72578441 | 1 | + | ENSG00000165091.18 | TMC1 | 56834 | 2,54264920467373E-08 | 0,000248698 | G > AG | 0,186311655 |
| **cg06192451** | chr17 | 69175414 | 69175414 | 1 | - | ENSG00000154262.13 | ABCA6 | 33518 | 2,72627790090325E-08 | 0,000257463 | G > AG | 0,148898946 |
| **cg17016145** | chr6 | 141199617 | 141199617 | 1 | + | ENSG00000203733.5 | GJE1 | 933472 | 2,96012793250113E-08 | 0,000276138 | G > AG | 0,090381867 |
| **cg02250187** | chr8 | 48876211 | 48876211 | 1 | - | ENSG00000019549.13 | SNAI2 | 45530 | 2,99917015426726E-08 | 0,00027641 | G > AG | 0,103572394 |
| **cg16156230** | chr11 | 30310621 | 30310621 | 1 | + | ENSG00000152219.5 | ARL14EP | 12482 | 3,13133075306623E-08 | 0,00028176 | G > AG | 0,108902416 |
| **cg17937897** | chr6 | 91694500 | 91694500 | 1 | + | ENSG00000135355.4 | GJA10 | 1800032 | 3,24113667919372E-08 | 0,00028176 | G > AG | 0,097443391 |
| **cg03293447** | chr1 | 42190260 | 42190260 | 1 | - | ENSG00000197273.4 | GUCA2A | 25514 | 3,21795008823866E-08 | 0,00028176 | G > AG | 0,080800543 |
| **cg01673931** | chr3 | 49119213 | 49119213 | 1 | + | ENSG00000185909.15 | KLHDC8B | 52384 | 3,18913636568036E-08 | 0,00028176 | G > AG | 0,134705958 |
| **cg11329482** | chr12 | 91099560 | 91099560 | 1 | + | ENSG00000187510.11 | PLEKHG7 | 1603282 | 3,24606145741847E-08 | 0,00028176 | G > AG | 0,098137113 |
| **cg07782669** | chr8 | 72731597 | 72731597 | 1 | - | ENSG00000164764.11 | SBSPON | 392492 | 3,27822945947078E-08 | 0,00028176 | G > AG | 0,117093922 |
| **cg20428112** | chr5 | 31023243 | 31023243 | 1 | + | ENSG00000113361.13 | CDH6 | 170442 | 3,52392802824242E-08 | 0,000299512 | G > AG | 0,080351491 |
| **cg15441396** | chr9 | 32842650 | 32842650 | 1 | - | ENSG00000137074.20 | APTX | 182481 | 3,60312455833143E-08 | 0,000302878 | G > AG | 0,112737287 |
| **cg23243492** | chr3 | 194494263 | 194494263 | 1 | - | ENSG00000133657.16 | ATP13A3 | 4102 | 3,67622629219564E-08 | 0,000305664 | G > AG | 0,129801788 |
| **cg20094706** | chr13 | 106700744 | 106700744 | 1 | - | ENSG00000134884.15 | ARGLU1 | 132606 | 3,82173339381957E-08 | 0,000314346 | G > AG | 0,156647082 |
| **cg22463921** | chr13 | 40065032 | 40065032 | 1 | - | ENSG00000183722.9 | LHFPL6 | 461503 | 3,99316109695157E-08 | 0,000324952 | G > AG | 0,138974076 |
| **cg11410281** | chr7 | 96921596 | 96921596 | 1 | - | ENSG00000105880.7 | DLX5 | 103355 | 4,08446574291109E-08 | 0,000325457 | G > AG | 0,107992816 |

| | | | | | | | | | | | | |
|---|---|---|---|---|---|---|---|---|---|---|---|---|
| **cg06671215** | chr2 | 54541500 | 54541500 | 1 | + | ENSG00000115306.16 | SPTBN1 | 85184 | 4,04877807789294E-08 | 0,000325457 | G > AG | 0,086178219 |
| **cg21617067** | chr5 | 79757423 | 79757423 | 1 | - | ENSG00000177034.17 | MTX3 | 233840 | 4,2198896072559E-08 | 0,000331694 | G > AG | 0,104987849 |
| **cg25469009** | chr1 | 218809644 | 218809644 | 1 | - | ENSG00000215817.7 | ZC3H11B | 803502 | 4,24945726435132E-08 | 0,000331694 | G > AG | 0,080910729 |
| **cg23450214** | chr2 | 223797314 | 223797314 | 1 | - | ENSG00000152056.17 | AP1S3 | 40714 | 5,12933822290187E-08 | 0,00034142 | G > AG | 0,15084614 |
| **cg01184676** | chr4 | 174214278 | 174214278 | 1 | + | ENSG00000164118.13 | CEP44 | 69451 | 5,13283125677229E-08 | 0,00034142 | G > AG | 0,134728993 |
| **cg13131223** | chr8 | 130521295 | 130521295 | 1 | + | ENSG00000132294.15 | EFR3A | 1382797 | 4,83582742193406E-08 | 0,00034142 | G > AG | 0,064144133 |
| **cg13932370** | chr12 | 70598943 | 70598943 | 1 | + | ENSG00000135643.5 | KCNMB4 | 232654 | 4,41956491762499E-08 | 0,00034142 | G > AG | 0,153186704 |
| **cg20566475** | chr10 | 124589703 | 124589703 | 1 | + | ENSG00000107902.14 | LHPP | 127881 | 5,02686314433856E-08 | 0,00034142 | G > AG | 0,121665665 |
| **cg17410895** | chr3 | 66785534 | 66785534 | 1 | - | ENSG00000144749.14 | LRIG1 | 284270 | 4,47565852576134E-08 | 0,00034142 | G > AG | 0,151321633 |
| **cg10828127** | chr21 | 40178887 | 40178887 | 1 | + | ENSG00000183036.11 | PCP4 | 311450 | 5,08990840058995E-08 | 0,00034142 | G > AG | 0,105530297 |
| **cg15177644** | chr9 | 6215375 | 6215375 | 1 | - | ENSG00000137040.10 | RANBP6 | 199749 | 4,74896108356816E-08 | 0,00034142 | G > AG | 0,118817852 |
| **cg01290136** | chr4 | 52925849 | 52925849 | 1 | + | ENSG00000128045.7 | RASL11B | 63533 | 5,07725099254584E-08 | 0,00034142 | G > AG | 0,1244728 |
| **cg20849255** | chr8 | 19495415 | 19495415 | 1 | + | ENSG00000104611.12 | SH2D4A | 181723 | 4,5656440379023E-08 | 0,00034142 | G > AG | 0,148598432 |
| **cg20340511** | chr15 | 45285901 | 45285901 | 1 | - | ENSG00000138606.19 | SHF | 84725 | 4,76836235876235E-08 | 0,00034142 | G > AG | 0,090623275 |
| **cg10178274** | chr9 | 14316957 | 14316957 | 1 | + | ENSG00000164975.15 | SNAPC3 | 1105746 | 4,59807912859089E-08 | 0,00034142 | G > AG | 0,155177554 |
| **cg11117908** | chr4 | 91087324 | 91087324 | 1 | - | ENSG00000145335.17 | SNCA | 1249008 | 4,53030945564141E-08 | 0,00034142 | G > AG | 0,108819114 |
| **cg12883521** | chr8 | 97261879 | 97261879 | 1 | - | ENSG00000180543.5 | TSPYL5 | 16050 | 5,08700147431191E-08 | 0,00034142 | G > AG | 0,153594333 |
| **cg12229408** | chr8 | 16363063 | 16363063 | 1 | - | ENSG00000038945.15 | MSR1 | 204428 | 5,31936706055622E-08 | 0,000350778 | G > AG | 0,152602039 |
| **cg10968631** | chr14 | 69519304 | 69519304 | 1 | - | ENSG00000267909.3 | CCDC177 | 55568 | 5,37253848816978E-08 | 0,000351256 | G > AG | 0,146958042 |
| **cg08977462** | chr5 | 178581945 | 178581945 | 1 | - | ENSG00000050767.18 | COL23A1 | 8449 | 5,53236821197183E-08 | 0,000355627 | G > AG | 0,102120259 |
| **cg06085726** | chr1 | 6500983 | 6500983 | 1 | - | ENSG00000171680.23 | PLEKHG5 | 19092 | 5,4958235058216E-08 | 0,000355627 | G > AG | 0,098098561 |
| **cg06854048** | chr10 | 67793864 | 67793864 | 1 | - | ENSG00000183230.18 | CTNNA3 | 30226 | 6,40506707026596E-08 | 0,000398335 | G > AG | 0,064680387 |
| **cg08287652** | chr5 | 96051272 | 96051272 | 1 | + | ENSG00000164292.13 | RHOBTB3 | 337751 | 6,38389696709897E-08 | 0,000398335 | G > AG | 0,071125411 |
| **cg06569266** | chr6 | 25831983 | 25831983 | 1 | + | ENSG00000146039.11 | SLC17A4 | 77285 | 6,26680705151546E-08 | 0,000398335 | G > AG | 0,163808888 |
| **cg07025874** | chr21 | 42042801 | 42042801 | 1 | + | ENSG00000177398.19 | UMODL1 | 20157 | 6,31579951115476E-08 | 0,000398335 | G > AG | 0,09630533 |
| **cg14043540** | chr7 | 101527864 | 101527864 | 1 | + | ENSG00000160963.14 | COL26A1 | 164990 | 6,93557930532524E-08 | 0,000421289 | G > AG | 0,119241259 |
| **cg20471017** | chr11 | 94304652 | 94304652 | 1 | - | ENSG00000123901.9 | GPR83 | 96768 | 6,8732948511862E-08 | 0,000421289 | G > AG | 0,123186586 |
| **cg10388077** | chr7 | 138204995 | 138204995 | 1 | + | ENSG00000122787.15 | AKR1D1 | 202672 | 7,10376972790939E-08 | 0,000427873 | G > AG | 0,113074237 |
| **cg27224143** | chr8 | 87044429 | 87044429 | 1 | - | ENSG00000170289.13 | CNGB3 | 300753 | 7,31507375857844E-08 | 0,000437158 | G > AG | 0,080627589 |
| **cg16073408** | chr7 | 44088619 | 44088619 | 1 | + | ENSG00000106624.11 | AEBP1 | 15725 | 7,70006580075605E-08 | 0,000444123 | G > AG | 0,113588244 |
| **cg10427769** | chr6 | 166705246 | 166705246 | 1 | + | ENSG00000213066.13 | CEP43 | 294070 | 7,70811244200404E-08 | 0,000444123 | G > AG | 0,088260868 |

| | | | | | | | | | | | | | |
|---|---|---|---|---|---|---|---|---|---|---|---|---|---|
| **cg11339888** | chr2 | 172927882 | 172927882 | 1 | + | ENSG00000091436.17 | MAP3K20 | 147552 | 7,72191725590121E-08 | 0,000444123 | | G > AG | 0,118907783 |
| **cg16902747** | chr22 | 35549453 | 35549453 | 1 | - | ENSG00000198125.13 | MB | 88499 | 7,60642544173996E-08 | 0,000444123 | | G > AG | 0,136939598 |
| **cg22380324** | chr17 | 29057337 | 29057337 | 1 | + | ENSG00000179761.12 | PIPOX | 106825 | 7,64944309513078E-08 | 0,000444123 | | G > AG | 0,079274636 |
| **cg04616525** | chr18 | 21315752 | 21315752 | 1 | + | ENSG00000141449.15 | GREB1L | 73511 | 7,91849277184606E-08 | 0,000448682 | | G > AG | 0,123710927 |
| **cg00711366** | chr8 | 67732089 | 67732089 | 1 | + | ENSG00000046889.19 | PREX2 | 219956 | 8,05608346860891E-08 | 0,000453122 | | G > AG | 0,118148278 |
| **cg11259937** | chr17 | 19709874 | 19709874 | 1 | + | ENSG00000072210.19 | ALDH3A2 | 61739 | 8,32951544317562E-08 | 0,000461712 | | G > AG | 0,109324962 |
| **cg13809736** | chr9 | 79663442 | 79663442 | 1 | - | ENSG00000156052.11 | GNAQ | 1631630 | 8,31651906340522E-08 | 0,000461712 | | G > AG | 0,105284124 |
| **cg08655871** | chr4 | 79050254 | 79050254 | 1 | + | ENSG00000138756.19 | BMP2K | 273913 | 8,87215841189725E-08 | 0,000474595 | | G > AG | 0,157697994 |
| **cg20381334** | chr5 | 124310875 | 124310875 | 1 | + | ENSG00000151292.17 | CSNK1G3 | 798777 | 8,83345347166976E-08 | 0,000474595 | | G > AG | 0,109008721 |
| **cg06263430** | chr4 | 10521758 | 10521758 | 1 | + | ENSG00000169676.6 | DRD5 | 740125 | 8,84951960834765E-08 | 0,000474595 | | G > AG | 0,084230231 |
| **cg16068519** | chr8 | 19495518 | 19495518 | 1 | + | ENSG00000104611.12 | SH2D4A | 181826 | 8,69059680115186E-08 | 0,000474595 | | G > AG | 0,133219208 |
| **cg07329095** | chr6 | 70183652 | 70183652 | 1 | - | ENSG00000112280.18 | COL9A1 | 119433 | 8,95834440703081E-08 | 0,000475878 | | G > AG | 0,113020039 |
| **cg05085472** | chr3 | 71163960 | 71163960 | 1 | - | ENSG00000114861.23 | FOXP1 | 420030 | 9,13016234039487E-08 | 0,00048166 | | G > AG | 0,120586542 |
| **cg23213339** | chr18 | 7941889 | 7941889 | 1 | + | ENSG00000173482.17 | PTPRM | 375108 | 9,43191497800321E-08 | 0,00048876 | | G > AG | 0,097596802 |
| **cg13101347** | chr18 | 8469116 | 8469116 | 1 | + | ENSG00000206418.5 | RAB12 | 140320 | 9,45642827535402E-08 | 0,00048876 | | G > AG | 0,105172752 |
| **cg06785806** | chr7 | 26653595 | 26653595 | 1 | + | ENSG00000086300.16 | SNX10 | 361734 | 9,33646521106911E-08 | 0,00048876 | | G > AG | 0,089601784 |
| **cg04680436** | chr2 | 241900517 | 241900517 | 1 | + | ENSG00000188011.5 | RTP5 | 30918 | 9,70667931111884E-08 | 0,000495005 | | G > AG | 0,087143002 |
| **cg22086461** | chr8 | 76431493 | 76431493 | 1 | - | ENSG00000164751.15 | PEX2 | 569552 | 9,90352661641052E-08 | 0,000501699 | | G > AG | 0,124963323 |
| **cg14690649** | chr12 | 31731721 | 31731721 | 1 | - | ENSG00000151743.11 | AMN1 | 2599 | 1,01712417337402E-07 | 0,00050803 | | G > AG | 0,113064564 |
| **cg22233451** | chr17 | 15630953 | 15630953 | 1 | - | ENSG00000241322.11 | CDRT1 | 11248 | 1,02044421598541E-07 | 0,00050803 | | G > AG | 0,07298182 |
| **cg04148163** | chr10 | 63682529 | 63682529 | 1 | - | ENSG00000171988.19 | JMJD1C | 160678 | 1,02277403797956E-07 | 0,00050803 | | G > AG | 0,111866864 |
| **cg23451405** | chr19 | 38060398 | 38060398 | 1 | + | ENSG00000105738.11 | SIPA1L3 | 153191 | 1,03329100112679E-07 | 0,000509942 | | G > AG | 0,128181041 |
| **cg17673665** | chr21 | 34774933 | 34774933 | 1 | + | ENSG00000159212.13 | CLIC6 | 105940 | 1,11844950092905E-07 | 0,000548431 | | G > AG | 0,071175102 |
| **cg10785556** | chr7 | 25385644 | 25385644 | 1 | + | ENSG00000050344.9 | NFE2L3 | 766553 | 1,13838340705534E-07 | 0,00055465 | | G > AG | 0,083965098 |
| **cg07540258** | chr5 | 33328801 | 33328801 | 1 | + | ENSG00000113407.14 | TARS1 | 111894 | 1,16714195456316E-07 | 0,000565063 | | G > AG | 0,11507946 |
| **cg10809656** | chr7 | 27641542 | 27641542 | 1 | + | ENSG00000106052.14 | TAX1BP1 | 97788 | 1,19216317815892E-07 | 0,000573547 | | G > AG | 0,091595442 |
| **cg14931118** | chr2 | 212364232 | 212364232 | 1 | - | ENSG00000178568.15 | ERBB4 | 174610 | 1,25252947592258E-07 | 0,000592786 | | G > AG | 0,109741538 |
| **cg22714069** | chr17 | 55706885 | 55706885 | 1 | + | ENSG00000141179.14 | PCTP | 44165 | 1,24179287981388E-07 | 0,000592786 | | G > AG | 0,088801725 |
| **cg15819366** | chr8 | 58864484 | 58864484 | 1 | + | ENSG00000137575.12 | SDCBP | 311561 | 1,25540263846354E-07 | 0,000592786 | | G > AG | 0,113644806 |
| **cg08329397** | chr7 | 48250914 | 48250914 | 1 | + | ENSG00000179869.15 | ABCA13 | 79457 | 1,27422164020175E-07 | 0,000593162 | | G > AG | 0,159294062 |
| **cg00998593** | chr1 | 83441218 | 83441218 | 1 | + | ENSG00000142875.19 | PRKACB | 636843 | 1,27946186829331E-07 | 0,000593162 | | G > AG | 0,103084721 |

| | | | | | | | | | | | | |
|---|---|---|---|---|---|---|---|---|---|---|---|---|
| **cg06366833** | chr4 | 182789320 | 182789320 | 1 | + | ENSG00000151718.16 | WWC2 | 309936 | 1,29807560487534E-07 | 0,000594585 | G > AG | 0,134293695 |
| **cg10901805** | chr11 | 84923576 | 84923576 | 1 | + | ENSG00000171204.13 | TMEM126B | 704996 | 1,31364269936264E-07 | 0,000598133 | G > AG | 0,142566051 |
| **cg25494227** | chr12 | 10179015 | 10179015 | 1 | - | ENSG00000173391.9 | OLR1 | 6876 | 1,32607988295487E-07 | 0,000600224 | G > AG | 0,086694429 |
| **cg11972379** | chr2 | 52500470 | 52500470 | 1 | - | ENSG00000179915.24 | NRXN1 | 1274894 | 1,37031455504268E-07 | 0,000616597 | G > AG | 0,123689419 |
| **cg12831678** | chr8 | 90227311 | 90227311 | 1 | - | ENSG00000104327.7 | CALB1 | 131835 | 1,40502867117483E-07 | 0,000624866 | G > AG | 0,105738389 |
| **cg23132327** | chr11 | 80456555 | 80456555 | 1 | - | ENSG00000149256.16 | TENM4 | 1015524 | 1,42530502582909E-07 | 0,000630219 | G > AG | 0,086448625 |
| **cg26941801** | chr13 | 32308708 | 32308708 | 1 | + | ENSG00000139618.17 | BRCA2 | 6377 | 1,4773759102074E-07 | 0,000645778 | G > AG | 0,123235115 |
| **cg05439421** | chr5 | 3196496 | 3196496 | 1 | - | ENSG00000170561.13 | IRX2 | 444818 | 1,47694556616978E-07 | 0,000645778 | G > AG | 0,086809903 |
| **cg21188409** | chr3 | 180748252 | 180748252 | 1 | - | ENSG00000284862.3 | CCDC39 | 63309 | 1,5220217675495E-07 | 0,000661513 | G > AG | 0,130803022 |
| **cg19987900** | chr14 | 59377726 | 59377726 | 1 | - | ENSG00000181619.11 | GPR135 | 87617 | 1,56515856527835E-07 | 0,000672959 | G > AG | 0,136949685 |
| **cg14389442** | chr2 | 1231351 | 1231351 | 1 | - | ENSG00000130508.11 | PXDN | 513502 | 1,56595150439163E-07 | 0,000672959 | G > AG | 0,104885986 |
| **cg00027114** | chr7 | 42700887 | 42700887 | 1 | - | ENSG00000136197.13 | C7orf25 | 211419 | 1,61059995711856E-07 | 0,000688279 | G > AG | 0,081502105 |
| **cg14977361** | chr10 | 31951944 | 31951944 | 1 | - | ENSG00000165322.18 | ARHGAP12 | 23067 | 1,64329062124964E-07 | 0,000692479 | G > AG | 0,104740839 |
| **cg13100009** | chr8 | 126571863 | 126571863 | 1 | - | ENSG00000168672.4 | LRATD2 | 13384 | 1,65559502354863E-07 | 0,000692479 | G > AG | 0,103653877 |
| **cg08530632** | chr1 | 88571487 | 88571487 | 1 | + | ENSG00000065243.20 | PKN2 | 112734 | 1,65663785880516E-07 | 0,000692479 | G > AG | 0,076568007 |
| **cg02362978** | chr17 | 9646820 | 9646820 | 1 | + | ENSG00000154914.17 | USP43 | 2123 | 1,70388681041771E-07 | 0,000708358 | G > AG | 0,15116683 |
| **cg17452384** | chr4 | 86453084 | 86453084 | 1 | - | ENSG00000109339.24 | MAPK10 | 141542 | 1,71420693181593E-07 | 0,000708797 | G > AG | 0,092591604 |
| **cg10774926** | chr4 | 34270839 | 34270839 | 1 | + | ENSG00000197057.10 | DTHD1 | 2010776 | 1,82407242900292E-07 | 0,000750169 | G > AG | 0,101701749 |
| **cg18505325** | chr13 | 24194642 | 24194642 | 1 | + | ENSG00000240654.6 | C1QTNF9 | 112523 | 1,87065634724266E-07 | 0,000765213 | G > AG | 0,133197166 |
| **cg13403290** | chr11 | 3721106 | 3721106 | 1 | + | ENSG00000129744.3 | ART1 | 75979 | 1,88923694133675E-07 | 0,000768703 | G > AG | 0,132248266 |
| **cg06074555** | chr7 | 20221023 | 20221023 | 1 | + | ENSG00000105855.10 | ITGB8 | 109678 | 1,92090626033189E-07 | 0,000774062 | G > AG | 0,120605106 |
| **cg18763247** | chr2 | 12767787 | 12767787 | 1 | - | ENSG00000169006.7 | NTSR2 | 1097591 | 1,9256498027888E-07 | 0,000774062 | G > AG | 0,096760337 |
| **cg00469952** | chr21 | 35168898 | 35168898 | 1 | - | ENSG00000159200.18 | RCAN1 | 553784 | 1,93276490812348E-07 | 0,000774062 | G > AG | 0,081321398 |
| **cg04099688** | chr2 | 212227773 | 212227773 | 1 | - | ENSG00000178568.15 | ERBB4 | 311069 | 2,0302258861798E-07 | 0,00080886 | G > AG | 0,098835615 |
| **cg06449036** | chr12 | 48527857 | 48527857 | 1 | + | ENSG00000284723.2 | OR8S1 | 2226 | 2,0464726906337E-07 | 0,000811108 | G > AG | 0,14048693 |
| **cg20226073** | chr3 | 67875932 | 67875932 | 1 | - | ENSG00000172340.15 | SUCLG2 | 221319 | 2,08604957278412E-07 | 0,000819421 | G > AG | 0,09905828 |
| **cg19187308** | chr2 | 21468786 | 21468786 | 1 | - | ENSG00000084674.15 | APOB | 424712 | 2,1278249947247E-07 | 0,000830443 | G > AG | 0,072892853 |
| **cg14550716** | chr7 | 118005091 | 118005091 | 1 | + | ENSG00000128534.8 | LSM 8,00 | 179052 | 2,15809589394432E-07 | 0,00083375 | G > AG | 0,119252354 |
| **cg00755146** | chr8 | 48192304 | 48192304 | 1 | - | ENSG00000253729.8 | PRKDC | 232125 | 2,15103409236102E-07 | 0,00083375 | G > AG | 0,153127606 |
| **cg16014233** | chr11 | 10633744 | 10633744 | 1 | - | ENSG00000133800.9 | LYVE1 | 22054 | 2,19483235756747E-07 | 0,000843681 | G > AG | 0,093586614 |
| **cg14848461** | chr3 | 74011831 | 74011831 | 1 | + | ENSG00000255423.1 | EBLN2 | 950173 | 2,22175534484011E-07 | 0,00084976 | G > AG | 0,092026185 |

| | | | | | | | | | | | | |
|---|---|---|---|---|---|---|---|---|---|---|---|---|
| **cg09138220** | chr9 | 68832860 | 68832860 | 1 | + | ENSG00000187866.10 | PABIR1 | 52796 | 2,27279173529161E-07 | 0,000864955 | G > AG | 0,123178988 |
| **cg05051967** | chr3 | 66602735 | 66602735 | 1 | - | ENSG00000144749.14 | LRIG1 | 101471 | 2,3243231941344E-07 | 0,000875937 | G > AG | 0,10331669 |
| **cg12277566** | chr2 | 85781071 | 85781071 | 1 | - | ENSG00000168878.19 | SFTPB | 112329 | 2,33599997525076E-07 | 0,000875937 | G > AG | 0,113359948 |
| **cg24853285** | chr2 | 75620139 | 75620139 | 1 | + | ENSG00000115364.14 | MRPL19 | 26643 | 2,35819195140548E-07 | 0,000877845 | G > AG | 0,106489001 |
| **cg07209547** | chr7 | 16833567 | 16833567 | 1 | - | ENSG00000106541.12 | AGR2 | 133 | 2,43889187674984E-07 | 0,000884179 | G > AG | 0,117074849 |
| **cg25641388** | chr4 | 141576225 | 141576225 | 1 | + | ENSG00000164136.17 | IL15 | 60357 | 2,43032875060348E-07 | 0,000884179 | G > AG | 0,06615328 |
| **cg09308847** | chr16 | 29231281 | 29231281 | 1 | - | ENSG00000254206.5 | NPIPB11 | 172749 | 2,41967210355552E-07 | 0,000884179 | G > AG | 0,107207084 |
| **cg03601444** | chr18 | 10754210 | 10754210 | 1 | - | ENSG00000154864.13 | PIEZO2 | 395360 | 2,39636115984721E-07 | 0,000884179 | G > AG | 0,100141056 |
| **cg18379838** | chr6 | 134308128 | 134308128 | 1 | + | ENSG00000288529.1 | RP1-73H22.5 | 354825 | 2,43824645858949E-07 | 0,000884179 | G > AG | 0,105050127 |
| **cg09131332** | chr22 | 50043616 | 50043616 | 1 | - | ENSG00000138892.11 | TTLL8 | 13320 | 2,49418269332396E-07 | 0,000897915 | G > AG | 0,143868238 |
| **cg12767627** | chr5 | 123209292 | 123209292 | 1 | + | ENSG00000061455.11 | PRDM6 | 120052 | 2,52367602958115E-07 | 0,00090209 | G > AG | 0,119545824 |
| **cg12301872** | chr2 | 381528 | 381528 | 1 | + | ENSG00000143727.16 | ACP1 | 117389 | 2,58328821261194E-07 | 0,000918823 | G > AG | 0,081848429 |
| **cg05783444** | chr3 | 167886972 | 167886972 | 1 | - | ENSG00000114209.15 | PDCD10 | 152032 | 2,5945109498016E-07 | 0,000918823 | G > AG | 0,111698522 |
| **cg18202853** | chr12 | 120382027 | 120382027 | 1 | - | ENSG00000135097.7 | MSI1 | 12862 | 2,66307328015236E-07 | 0,000938758 | G > AG | 0,116550866 |
| **cg16035872** | chr11 | 13041398 | 13041398 | 1 | - | ENSG00000148925.11 | BTBD10 | 421900 | 2,73256446304903E-07 | 0,00094272 | G > AG | 0,137207954 |
| **cg13724533** | chr15 | 57445367 | 57445367 | 1 | + | ENSG00000128849.11 | CGNL1 | 69401 | 2,75602153271805E-07 | 0,00094272 | G > AG | 0,106925729 |
| **cg00584227** | chr1 | 36107640 | 36107640 | 1 | - | ENSG00000171812.13 | COL8A2 | 17583 | 2,78522923401293E-07 | 0,00094272 | G > AG | 0,077826478 |
| **cg07759714** | chr6 | 129341268 | 129341268 | 1 | + | ENSG00000196569.13 | LAMA2 | 458131 | 2,70600842689749E-07 | 0,00094272 | G > AG | 0,077569506 |
| **cg23787564** | chr6 | 112641093 | 112641093 | 1 | - | ENSG00000112769.20 | LAMA4 | 386153 | 2,73848055837789E-07 | 0,00094272 | G > AG | 0,105111716 |
| **cg12020736** | chr3 | 169134586 | 169134586 | 1 | + | ENSG00000085274.16 | MYNN | 638809 | 2,78225868294979E-07 | 0,00094272 | G > AG | 0,118889075 |
| **cg22439873** | chr10 | 28095932 | 28095932 | 1 | - | ENSG00000169126.16 | ODAD2 | 96852 | 2,74399334042865E-07 | 0,00094272 | G > AG | 0,116641086 |
| **cg21341499** | chr4 | 161763077 | 161763077 | 1 | - | ENSG00000168843.14 | FSTL5 | 400928 | 2,80412508177993E-07 | 0,000944934 | G > AG | 0,090479539 |
| **cg26485452** | chr1 | 193589908 | 193589908 | 1 | - | ENSG00000162630.6 | B3GALT2 | 403294 | 2,84227949830457E-07 | 0,000953591 | G > AG | 0,116150954 |
| **cg12456833** | chr20 | 23013720 | 23013720 | 1 | - | ENSG00000178726.7 | THBD | 35953 | 2,87526470337361E-07 | 0,000960445 | G > AG | 0,11079441 |
| **cg17536666** | chr20 | 52501186 | 52501186 | 1 | - | ENSG00000020256.20 | ZFP64 | 296877 | 3,01274036659045E-07 | 0,00098909 | G > AG | 0,108658633 |
| **cg23442245** | chr18 | 36643272 | 36643272 | 1 | - | ENSG00000134779.15 | TPGS2 | 185945 | 3,037157547655E-07 | 0,000992845 | G > AG | 0,087237558 |
| **cg19984262** | chr4 | 174214255 | 174214255 | 1 | + | ENSG00000164118.13 | CEP44 | 69474 | 3,06785879316745E-07 | 0,000994382 | G > AG | 0,160992378 |
| **cg13860754** | chr9 | 86046907 | 86046907 | 1 | + | ENSG00000135040.16 | NAA35 | 105762 | 3,0566273282374E-07 | 0,000994382 | G > AG | 0,134039131 |
| **cg15562860** | chr6 | 131001543 | 131001543 | 1 | + | ENSG00000118507.18 | AKAP7 | 133923 | 3,1584174865255E-07 | 0,000998353 | G > AG | 0,06715567 |
| **cg22253078** | chr2 | 211331674 | 211331674 | 1 | - | ENSG00000115365.12 | LANCL1 | 854021 | 3,14150822548153E-07 | 0,000998353 | G > AG | 0,092096059 |
| **cg04857926** | chr11 | 8313171 | 8313171 | 1 | - | ENSG00000166407.14 | LMO1 | 44454 | 3,13209868576786E-07 | 0,000998353 | G > AG | 0,12401883 |

| | | | | | | | | | | | | |
|---|---|---|---|---|---|---|---|---|---|---|---|---|
| **cg23700156** | chr3 | 131553602 | 131553602 | 1 | + | ENSG00000198585.12 | NUDT16 | 171932 | 3,1547169588898E-07 | 0,000998353 | G > AG | 0,11556011 |
| **cg11767349** | chr7 | 137756420 | 137756420 | 1 | + | ENSG00000122787.15 | AKR1D1 | 245903 | 3,17249521939035E-07 | 0,000998676 | G > AG | 0,11707608 |
| **cg16358698** | chr1 | 34379479 | 34379479 | 1 | - | ENSG00000121904.18 | CSMD2 | 213636 | 3,20291373136943E-07 | 0,00100412 | G > AG | 0,085118916 |
| **cg11978968** | chr2 | 76985804 | 76985804 | 1 | - | ENSG00000176204.14 | LRRTM4 | 607516 | 3,22952114327216E-07 | 0,001008329 | G > AG | 0,128489012 |
| **cg19197789** | chr2 | 112869528 | 112869528 | 1 | + | ENSG00000125571.10 | IL37 | 41636 | 3,26305061777759E-07 | 0,001014656 | G > AG | 0,161518448 |
| **cg17583307** | chr7 | 73351755 | 73351755 | 1 | - | ENSG00000146755.11 | TRIM50 | 23672 | 3,31495279554819E-07 | 0,001026622 | G > AG | 0,092087593 |
| **cg18606098** | chr4 | 103077560 | 103077560 | 1 | - | ENSG00000164038.16 | SLC9B2 | 8270 | 3,34731941150337E-07 | 0,001032466 | G > AG | 0,103887952 |
| **cg21832319** | chr9 | 120290074 | 120290074 | 1 | + | ENSG00000214654.8 | B3GALT9 | 502328 | 3,41819936632435E-07 | 0,001041727 | G > AG | 0,158101924 |
| **cg02523907** | chr2 | 21758230 | 21758230 | 1 | + | ENSG00000218819.6 | TDRD15 | 634263 | 3,39542579912021E-07 | 0,001041727 | G > AG | 0,133337429 |
| **cg08057475** | chr15 | 45511143 | 45511143 | 1 | - | ENSG00000104154.7 | SLC30A4 | 11613 | 3,47132388688625E-07 | 0,001053719 | G > AG | 0,125234806 |
| **cg07587639** | chr11 | 113895656 | 113895656 | 1 | + | ENSG00000149305.7 | HTR3B | 9020 | 3,5307559577508E-07 | 0,001067523 | G > AG | 0,107128516 |
| **cg17627261** | chr16 | 47074020 | 47074020 | 1 | + | ENSG00000166123.14 | GPT2 | 189659 | 3,56264260322975E-07 | 0,001072923 | G > AG | 0,143933493 |
| **cg22176779** | chr20 | 22220246 | 22220246 | 1 | - | ENSG00000125798.15 | FOXA2 | 365210 | 3,63519984248922E-07 | 0,001077801 | G > AG | 0,103072534 |
| **cg06059360** | chr3 | 42616126 | 42616126 | 1 | + | ENSG00000114857.18 | NKTR | 15472 | 3,62323066577161E-07 | 0,001077801 | G > AG | 0,071104279 |
| **cg23639898** | chr18 | 25327809 | 25327809 | 1 | + | ENSG00000154611.15 | PSMA8 | 806042 | 3,59764357304953E-07 | 0,001077801 | G > AG | 0,08851551 |
| **cg15501329** | chr6 | 157716122 | 157716122 | 1 | - | ENSG00000215712.11 | TMEM242 | 392520 | 3,61416605672008E-07 | 0,001077801 | G > AG | 0,106211749 |
| **cg22946767** | chr1 | 20413544 | 20413544 | 1 | - | ENSG00000162545.6 | CAMK2N1 | 72667 | 3,67970661077233E-07 | 0,001078457 | G > AG | 0,065932713 |
| **cg09919215** | chr10 | 124703542 | 124703542 | 1 | - | ENSG00000189319.14 | FAM53B | 40837 | 3,65582836808982E-07 | 0,001078457 | G > AG | 0,058449056 |
| **cg12421116** | chr1 | 63803886 | 63803886 | 1 | - | ENSG00000142856.17 | ITGB3BP | 210164 | 3,67370914789888E-07 | 0,001078457 | G > AG | 0,089304114 |
| **cg05565492** | chr1 | 41442903 | 41442903 | 1 | + | ENSG00000204060.7 | FOXO6 | 80982 | 3,80063088735156E-07 | 0,001097084 | G > AG | 0,094430611 |
| **cg11328826** | chr11 | 75496724 | 75496724 | 1 | + | ENSG00000149257.16 | SERPINH1 | 65331 | 3,79943072929765E-07 | 0,001097084 | G > AG | 0,085597735 |
| **cg12859046** | chr1 | 193186078 | 193186078 | 1 | + | ENSG00000134371.14 | CDC73 | 64096 | 3,87156437674501E-07 | 0,001113358 | G > AG | 0,163891764 |
| **cg01417037** | chr5 | 4244562 | 4244562 | 1 | - | ENSG00000170561.13 | IRX2 | 1492884 | 3,89408537600679E-07 | 0,001115641 | G > AG | 0,111272273 |
| **cg23762915** | chr13 | 106702645 | 106702645 | 1 | + | ENSG00000182346.21 | DAOA | 1236779 | 3,94001484662811E-07 | 0,001120407 | G > AG | 0,102608081 |
| **cg16731114** | chr12 | 8661246 | 8661246 | 1 | - | ENSG00000197614.11 | MFAP5 | 1643 | 3,99631926530187E-07 | 0,001132209 | G > AG | 0,110348574 |
| **cg07671026** | chr5 | 9529325 | 9529325 | 1 | - | ENSG00000112902.12 | SEMA5A | 16751 | 4,07981897811602E-07 | 0,0011516 | G > AG | 0,087161029 |
| **cg14068729** | chr13 | 48490764 | 48490764 | 1 | - | ENSG00000136161.13 | RCBTB2 | 42493 | 4,16544493950121E-07 | 0,001171447 | G > AG | 0,10625798 |
| **cg24253892** | chr1 | 177651291 | 177651291 | 1 | + | ENSG00000075391.17 | RASAL2 | 442812 | 4,19764392244381E-07 | 0,001176178 | G > AG | 0,12242418 |
| **cg19579544** | chr14 | 62006337 | 62006337 | 1 | - | ENSG00000182107.7 | TMEM30B | 724575 | 4,30654003432692E-07 | 0,001202287 | G > AG | 0,09567144 |
| **cg13492483** | chr9 | 21549601 | 21549601 | 1 | - | ENSG00000184995.7 | IFNE | 67287 | 4,32669235605484E-07 | 0,00120352 | G > AG | 0,14909294 |
| **cg06072571** | chr12 | 10313535 | 10313535 | 1 | + | ENSG00000134539.17 | KLRD1 | 87478 | 4,46260335641141E-07 | 0,001236828 | G > AG | 0,104518856 |

| | | | | | | | | | | | | |
|---|---|---|---|---|---|---|---|---|---|---|---|---|
| **cg11322056** | chr16 | 65667028 | 65667028 | 1 | - | ENSG00000140937.14 | CDH11 | 540915 | 4,50478000978865E-07 | 0,001239535 | G > AG | 0,081150267 |
| **cg02211200** | chr1 | 241558501 | 241558501 | 1 | + | ENSG00000117009.12 | KMO | 26368 | 4,49986747708286E-07 | 0,001239535 | G > AG | 0,084462175 |
| **cg18244459** | chr6 | 12365927 | 12365927 | 1 | + | ENSG00000078401.7 | EDN1 | 75567 | 4,54566612909954E-07 | 0,001246302 | G > AG | 0,144192604 |
| **cg04810284** | chr4 | 79050023 | 79050023 | 1 | - | ENSG00000163291.14 | PAQR3 | 110584 | 4,71832741231856E-07 | 0,001284434 | G > AG | 0,144544034 |
| **cg06992006** | chr17 | 15453714 | 15453714 | 1 | - | ENSG00000239704.11 | CDRT4 | 49895 | 4,80698803963362E-07 | 0,001297641 | G > AG | 0,085680107 |
| **cg12866368** | chr10 | 21184550 | 21184550 | 1 | - | ENSG00000078114.19 | NEBL | 108462 | 4,81379404527555E-07 | 0,001297641 | G > AG | 0,077881932 |
| **cg10036405** | chr1 | 168920980 | 168920980 | 1 | + | ENSG00000143153.13 | ATP1B1 | 184716 | 4,94676920165963E-07 | 0,001300346 | G > AG | 0,108226978 |
| **cg25387812** | chr1 | 44088985 | 44088985 | 1 | - | ENSG00000283039.1 | KLF18 | 52647 | 4,92066248041184E-07 | 0,001300346 | G > AG | 0,080825339 |
| **cg13100005** | chr8 | 126571692 | 126571692 | 1 | - | ENSG00000168672.4 | LRATD2 | 13213 | 4,90936586099856E-07 | 0,001300346 | G > AG | 0,09443821 |
| **cg15990826** | chr8 | 105527509 | 105527509 | 1 | - | ENSG00000147650.12 | LRP12 | 938250 | 4,91966235091757E-07 | 0,001300346 | G > AG | 0,148916374 |
| **cg14579056** | chr13 | 97279019 | 97279019 | 1 | - | ENSG00000165621.9 | OXGR1 | 284288 | 4,94551963502995E-07 | 0,001300346 | G > AG | 0,081871571 |
| **cg20261451** | chr15 | 38593860 | 38593860 | 1 | - | ENSG00000172575.12 | RASGRP1 | 28284 | 4,85221601573062E-07 | 0,001300346 | G > AG | 0,092759255 |
| **cg08640867** | chr3 | 181624159 | 181624159 | 1 | + | ENSG00000181449.4 | SOX2 | 87765 | 5,14045483438543E-07 | 0,001342036 | G > AG | 0,102569273 |
| **cg09699104** | chr6 | 72760501 | 72760501 | 1 | - | ENSG00000256980.5 | KHDC1L | 465270 | 5,27474550315968E-07 | 0,001351786 | G > AG | 0,103109781 |
| **cg02623448** | chr20 | 40511681 | 40511681 | 1 | - | ENSG00000204103.4 | MAFB | 177556 | 5,26670409736856E-07 | 0,001351786 | G > AG | 0,075806512 |
| **cg21469364** | chr4 | 115671901 | 115671901 | 1 | + | ENSG00000270394.4 | MTRNR2L13 | 626974 | 5,30341813429599E-07 | 0,001351786 | G > AG | 0,119363096 |
| **cg21660392** | chr17 | 68954404 | 68954404 | 1 | + | ENSG00000108946.15 | PRKAR1A | 442625 | 5,27713976494384E-07 | 0,001351786 | G > AG | 0,083197231 |
| **cg17546509** | chr6 | 25925822 | 25925822 | 1 | + | ENSG00000112343.11 | TRIM38 | 36979 | 5,21171889306125E-07 | 0,001351786 | G > AG | 0,129965771 |
| **cg15876825** | chr3 | 11610407 | 11610407 | 1 | - | ENSG00000144560.15 | VGLL4 | 160944 | 5,2882573509099E-07 | 0,001351786 | G > AG | 0,106260766 |
| **cg06814469** | chr1 | 89943643 | 89943643 | 1 | + | ENSG00000162664.17 | ZNF326 | 51466 | 5,215291586951E-07 | 0,001351786 | G > AG | 0,124771586 |
| **cg06024391** | chr7 | 134660382 | 134660382 | 1 | - | ENSG00000085662.14 | AKR1B1 | 201097 | 5,41195876271452E-07 | 0,001366287 | G > AG | 0,078048985 |
| **cg06260423** | chr4 | 10184330 | 10184330 | 1 | + | ENSG00000169676.6 | DRD5 | 402697 | 5,39706146200201E-07 | 0,001366287 | G > AG | 0,111513156 |
| **cg13479148** | chr1 | 3437869 | 3437869 | 1 | + | ENSG00000130762.15 | ARHGEF16 | 16795 | 5,47323141058937E-07 | 0,001377024 | G > AG | 0,036052333 |
| **cg05686867** | chr10 | 44176653 | 44176653 | 1 | + | ENSG00000198298.13 | ZNF485 | 570235 | 5,49049118270016E-07 | 0,001377024 | G > AG | 0,120054715 |
| **cg23470828** | chr17 | 20824056 | 20824056 | 1 | - | ENSG00000124422.12 | USP22 | 219705 | 5,52587504518846E-07 | 0,001378801 | G > AG | 0,065218234 |
| **cg04783353** | chr3 | 40691392 | 40691392 | 1 | + | ENSG00000172888.12 | ZNF621 | 166515 | 5,5336264820949E-07 | 0,001378801 | G > AG | 0,141035049 |
| **cg06217164** | chr13 | 77980685 | 77980685 | 1 | - | ENSG00000136160.17 | EDNRB | 5155 | 5,5554499147287E-07 | 0,001379744 | G > AG | 0,100281363 |
| **cg00475558** | chr7 | 94585081 | 94585081 | 1 | - | ENSG00000127990.19 | SGCE | 71492 | 5,61571439157675E-07 | 0,001390198 | G > AG | 0,100941715 |
| **cg07700524** | chr16 | 62954378 | 62954378 | 1 | + | ENSG00000179776.19 | CDH5 | 3412243 | 5,63675852179351E-07 | 0,001390906 | G > AG | 0,135198801 |
| **cg21455213** | chr16 | 47847587 | 47847587 | 1 | - | ENSG00000140798.16 | ABCC12 | 308432 | 5,68137183308485E-07 | 0,001394655 | G > AG | 0,091658523 |
| **cg08795956** | chr1 | 167563902 | 167563902 | 1 | - | ENSG00000143162.9 | CREG1 | 10096 | 5,68841632130161E-07 | 0,001394655 | G > AG | 0,055959737 |

| | | | | | | | | | | | | |
|---|---|---|---|---|---|---|---|---|---|---|---|---|
| **cg10273615** | chr7 | 137804419 | 137804419 | 1 | + | ENSG00000122787.15 | AKR1D1 | 197904 | 5,7727285845256E-07 | 0,001396696 | G > AG | 0,122095697 |
| **cg17868538** | chr6 | 25232646 | 25232646 | 1 | + | ENSG00000079691.18 | CARMIL1 | 46431 | 5,7650093482277E-07 | 0,001396696 | G > AG | 0,084468117 |
| **cg20820996** | chr1 | 33284248 | 33284248 | 1 | + | ENSG00000160094.15 | ZNF362 | 27757 | 5,78803564788358E-07 | 0,001396696 | G > AG | 0,071640516 |
| **cg07494294** | chr6 | 1063963 | 1063963 | 1 | - | ENSG00000112685.14 | EXOC2 | 370823 | 6,01112254825379E-07 | 0,001441435 | G > AG | 0,077863646 |
| **cg18312429** | chr13 | 103066580 | 103066580 | 1 | - | ENSG00000125255.7 | SLC10A2 | 162 | 6,08033006586684E-07 | 0,001449956 | G > AG | 0,120331432 |
| **cg06574757** | chr3 | 160870480 | 160870480 | 1 | + | ENSG00000163590.14 | PPM1L | 114879 | 6,1874831411454E-07 | 0,001469902 | G > AG | 0,136018438 |
| **cg05644508** | chr2 | 57882979 | 57882979 | 1 | - | ENSG00000115392.12 | FANCL | 358394 | 6,22787806639443E-07 | 0,001474917 | G > AG | 0,118598223 |
| **cg12525187** | chr10 | 67181080 | 67181080 | 1 | + | ENSG00000198739.11 | LRRTM3 | 255045 | 6,27476181868498E-07 | 0,001480841 | G > AG | 0,122258021 |
| **cg17009481** | chr5 | 59638319 | 59638319 | 1 | - | ENSG00000113448.19 | PDE4D | 883802 | 6,29160932718376E-07 | 0,001480841 | G > AG | 0,098693552 |
| **cg26243693** | chr22 | 32542996 | 32542996 | 1 | + | ENSG00000100225.18 | FBXO7 | 68321 | 6,34453258743213E-07 | 0,001488717 | G > AG | 0,078006981 |
| **cg25473044** | chr20 | 58141521 | 58141521 | 1 | + | ENSG00000124237.6 | C20orf85 | 9380 | 6,39786129998251E-07 | 0,001491323 | G > AG | 0,079999935 |
| **cg10504624** | chr7 | 435960 | 435960 | 1 | + | ENSG00000248767.2 | FOXL3 | 145791 | 6,40153157447925E-07 | 0,001491323 | G > AG | 0,094283665 |
| **cg12853277** | chr1 | 236747685 | 236747685 | 1 | - | ENSG00000119285.11 | HEATR1 | 143168 | 6,41412595620174E-07 | 0,001491323 | G > AG | 0,076446376 |
| **cg18124672** | chr12 | 23054908 | 23054908 | 1 | + | ENSG00000139163.16 | ETNK1 | 429834 | 6,62873883501714E-07 | 0,001531909 | G > AG | 0,09803651 |
| **cg23195891** | chr18 | 6249875 | 6249875 | 1 | - | ENSG00000154655.16 | L3MBTL4 | 165363 | 6,792536087871E-07 | 0,001561395 | G > AG | 0,096107048 |
| **cg18951877** | chr13 | 84669142 | 84669142 | 1 | - | ENSG00000178235.8 | SLITRK1 | 786667 | 6,79715043601756E-07 | 0,001561395 | G > AG | 0,116327605 |
| **cg17772110** | chr12 | 68977530 | 68977530 | 1 | + | ENSG00000135679.26 | MDM2 | 169354 | 6,89176809203782E-07 | 0,001577107 | G > AG | 0,102969175 |
| **cg26554042** | chr6 | 63862947 | 63862947 | 1 | + | ENSG00000118482.12 | PHF3 | 227146 | 6,94561356543904E-07 | 0,001577107 | G > AG | 0,076003667 |
| **cg25816367** | chr21 | 34957184 | 34957184 | 1 | - | ENSG00000159200.18 | RCAN1 | 342070 | 6,96863495018751E-07 | 0,001577107 | G > AG | 0,100612842 |
| **cg26881307** | chr2 | 5982666 | 5982666 | 1 | + | ENSG00000176887.7 | SOX11 | 290283 | 6,95371516678149E-07 | 0,001577107 | G > AG | 0,099601841 |
| **cg14458315** | chr9 | 98317025 | 98317025 | 1 | - | ENSG00000095383.20 | TBC1D2 | 61375 | 6,96456556476965E-07 | 0,001577107 | G > AG | 0,091356356 |
| **cg19051042** | chr17 | 41524067 | 41524067 | 1 | + | ENSG00000173812.11 | EIF1 | 164817 | 7,07036551466936E-07 | 0,001590717 | G > AG | 0,11348949 |
| **cg26753123** | chr4 | 181660139 | 181660139 | 1 | - | ENSG00000129187.15 | DCTD | 1257798 | 7,53969045079261E-07 | 0,001607324 | G > AG | 0,136463934 |
| **cg13865846** | chr18 | 45517652 | 45517652 | 1 | - | ENSG00000152223.15 | EPG5 | 449678 | 7,43516621573648E-07 | 0,001607324 | G > AG | 0,111786256 |
| **cg05211862** | chr14 | 77542587 | 77542587 | 1 | - | ENSG00000100593.18 | ISM2 | 43770 | 7,43731782898402E-07 | 0,001607324 | G > AG | 0,137226669 |
| **cg02749012** | chr3 | 135229297 | 135229297 | 1 | - | ENSG00000174611.12 | KY | 577660 | 7,20635646087821E-07 | 0,001607324 | G > AG | 0,121052648 |
| **cg13027963** | chr8 | 117883367 | 117883367 | 1 | + | ENSG00000164758.7 | MED30 | 362655 | 7,22375766825032E-07 | 0,001607324 | G > AG | 0,120997014 |
| **cg10135231** | chr6 | 136173888 | 136173888 | 1 | - | ENSG00000146410.12 | MTFR2 | 76448 | 7,41682620415648E-07 | 0,001607324 | G > AG | 0,103837137 |
| **cg07027506** | chr8 | 3375142 | 3375142 | 1 | + | ENSG00000036448.10 | MYOM2 | 1330097 | 7,43870393966426E-07 | 0,001607324 | G > AG | 0,084652992 |
| **cg14003693** | chr9 | 14308248 | 14308248 | 1 | - | ENSG00000147862.17 | NFIB | 90736 | 7,4697804419198E-07 | 0,001607324 | G > AG | 0,122382511 |
| **cg23655242** | chr1 | 236155245 | 236155245 | 1 | - | ENSG00000116962.15 | NID1 | 90135 | 7,27442514883492E-07 | 0,001607324 | G > AG | 0,075620606 |

| | | | | | | | | | | | | |
|---|---|---|---|---|---|---|---|---|---|---|---|---|
| **cg12389774** | chr8 | 31821810 | 31821810 | 1 | - | ENSG00000172733.12 | PURG | 788094 | 7,20178838982671E-07 | 0,001607324 | G > AG | 0,089778198 |
| **cg07949863** | chr20 | 17735271 | 17735271 | 1 | - | ENSG00000125844.16 | RRBP1 | 52975 | 7,54341260741344E-07 | 0,001607324 | G > AG | 0,104748878 |
| **cg02763462** | chr11 | 122229312 | 122229312 | 1 | + | ENSG00000154127.10 | UBASH3B | 426409 | 7,53548641549969E-07 | 0,001607324 | G > AG | 0,135501749 |
| **cg15143024** | chr7 | 91570327 | 91570327 | 1 | + | ENSG00000157240.4 | FZD1 | 305895 | 7,65204496878883E-07 | 0,001625942 | G > AG | 0,060624787 |
| **cg02642328** | chr8 | 8297003 | 8297003 | 1 | - | ENSG00000275342.5 | PRAG1 | 89437 | 7,72971335231473E-07 | 0,001637896 | G > AG | 0,095363302 |
| **cg17086139** | chr12 | 102634465 | 102634465 | 1 | - | ENSG00000017427.17 | IGF1 | 152720 | 7,78922538154266E-07 | 0,001641413 | G > AG | 0,159154543 |
| **cg17061156** | chr4 | 184020341 | 184020341 | 1 | + | ENSG00000173320.12 | STOX2 | 222650 | 7,78580823679585E-07 | 0,001641413 | G > AG | 0,085604407 |
| **cg18181923** | chr14 | 99215932 | 99215932 | 1 | - | ENSG00000127152.18 | BCL11B | 56266 | 7,84040320483671E-07 | 0,001647658 | G > AG | 0,08550978 |
| **cg25828346** | chr2 | 239140355 | 239140355 | 1 | + | ENSG00000233608.4 | TWIST2 | 292324 | 7,8884751063326E-07 | 0,001653219 | G > AG | 0,099610851 |
| **cg25844787** | chr7 | 55158516 | 55158516 | 1 | + | ENSG00000146648.20 | EGFR | 139500 | 7,95176587881009E-07 | 0,001657401 | G > AG | 0,125203087 |
| **cg07040325** | chr8 | 36783695 | 36783695 | 1 | + | ENSG00000215262.8 | KCNU1 | 628 | 8,11880397986262E-07 | 0,001685698 | G > AG | 0,09934897 |
| **cg22831463** | chr17 | 65610063 | 65610063 | 1 | + | ENSG00000108370.17 | RGS9 | 509252 | 8,13160131114483E-07 | 0,001685698 | G > AG | 0,109156414 |
| **cg19727165** | chr3 | 94177283 | 94177283 | 1 | - | ENSG00000178700.9 | DHFR2 | 113893 | 8,2995290721986E-07 | 0,001711235 | G > AG | 0,088227893 |
| **cg04958239** | chr3 | 55609627 | 55609627 | 1 | - | ENSG00000114251.15 | WNT5A | 119087 | 8,28292357459455E-07 | 0,001711235 | G > AG | 0,04742782 |
| **cg17187705** | chr13 | 35161163 | 35161163 | 1 | - | ENSG00000180660.8 | MAB21L1 | 315527 | 8,35047844933966E-07 | 0,001715534 | G > AG | 0,089012804 |
| **cg21066876** | chr4 | 95051315 | 95051315 | 1 | - | ENSG00000182168.15 | UNC5C | 497892 | 8,36523513076713E-07 | 0,001715534 | G > AG | 0,102935444 |
| **cg04096619** | chr5 | 9547483 | 9547483 | 1 | + | ENSG00000150753.12 | CCT5 | 702445 | 8,46976377461577E-07 | 0,001727707 | G > AG | 0,15995256 |
| **cg16822666** | chr17 | 37299735 | 37299735 | 1 | + | ENSG00000278505.5 | C17orf78 | 76249 | 8,49979656449962E-07 | 0,001729222 | G > AG | 0,113362082 |
| **cg00872683** | chr4 | 40395988 | 40395988 | 1 | + | ENSG00000174343.6 | CHRNA9 | 60656 | 8,57604557721775E-07 | 0,001740028 | G > AG | 0,091436476 |
| **cg07559178** | chr14 | 53369771 | 53369771 | 1 | - | ENSG00000100523.16 | DDHD1 | 216447 | 8,61695320251015E-07 | 0,001740028 | G > AG | 0,092889297 |
| **cg06866827** | chr1 | 3031900 | 3031900 | 1 | - | ENSG00000215912.12 | TTC34 | 230182 | 8,64275769308469E-07 | 0,001740028 | G > AG | 0,138535459 |
| **cg21570457** | chr10 | 31781191 | 31781191 | 1 | - | ENSG00000165322.18 | ARHGAP12 | 147686 | 8,71424704886358E-07 | 0,001749585 | G > AG | 0,082475632 |
| **cg16518145** | chr10 | 28876013 | 28876013 | 1 | - | ENSG00000150054.19 | MPP7 | 541526 | 8,73718491682709E-07 | 0,001749598 | G > AG | 0,083329237 |
| **cg12599765** | chr4 | 111808546 | 111808546 | 1 | + | ENSG00000174749.6 | FAM241A | 336907 | 8,80953002717654E-07 | 0,001754897 | G > AG | 0,106626698 |
| **cg15444081** | chr7 | 123766446 | 123766446 | 1 | + | ENSG00000106302.10 | HYAL4 | 62536 | 9,03112880252525E-07 | 0,001794368 | G > AG | 0,119589295 |
| **cg19095511** | chr13 | 103008491 | 103008491 | 1 | + | ENSG00000134899.24 | ERCC5 | 162661 | 9,05658865132941E-07 | 0,001794764 | G > AG | 0,089348657 |
| **cg13247463** | chr13 | 37173342 | 37173342 | 1 | + | ENSG00000120699.13 | EXOSC8 | 174527 | 9,09174395976218E-07 | 0,001795155 | G > AG | 0,091616106 |
| **cg03286617** | chr2 | 153479888 | 153479888 | 1 | + | ENSG00000144278.15 | GALNT13 | 392033 | 9,10549537272212E-07 | 0,001795155 | G > AG | 0,11898175 |
| **cg21170038** | chr17 | 73977000 | 73977000 | 1 | - | ENSG00000069188.17 | SDK2 | 332554 | 9,15155824559897E-07 | 0,001799598 | G > AG | 0,08642765 |
| **cg25794897** | chr21 | 33115983 | 33115983 | 1 | - | ENSG00000205929.11 | C21orf62 | 302239 | 9,19123916466252E-07 | 0,001802767 | G > AG | 0,088928133 |
| **cg07797660** | chr1 | 3322575 | 3322575 | 1 | + | ENSG00000130762.15 | ARHGEF16 | 132089 | 9,23589980997582E-07 | 0,001806894 | G > AG | 0,11679493 |

| | | | | | | | | | | | | |
|---|---|---|---|---|---|---|---|---|---|---|---|---|
| **cg14717436** | chr6 | 146410744 | 146410744 | 1 | - | ENSG00000146414.16 | SHPRH | 446320 | 9,38851920040529E-07 | 0,001832066 | G > AG | 0,092136814 |
| **cg10833393** | chr19 | 46147970 | 46147970 | 1 | - | ENSG00000188624.3 | IGFL3 | 23281 | 9,41963672104629E-07 | 0,001833461 | G > AG | 0,092287041 |
| **cg02218140** | chr1 | 242240249 | 242240249 | 1 | + | ENSG00000196289.7 | BECN2 | 282483 | 9,52520227718696E-07 | 0,001841898 | G > AG | 0,096592839 |
| **cg22233757** | chr19 | 46056663 | 46056663 | 1 | - | ENSG00000204869.8 | IGFL4 | 20456 | 9,55929823619274E-07 | 0,001841898 | G > AG | 0,100545004 |
| **cg18287241** | chr5 | 167392289 | 167392289 | 1 | - | ENSG00000120137.7 | PANK3 | 1187080 | 9,50932950322747E-07 | 0,001841898 | G > AG | 0,103168962 |
| **cg20299313** | chr20 | 46671226 | 46671226 | 1 | + | ENSG00000197496.6 | SLC2A10 | 38422 | 9,54367193515538E-07 | 0,001841898 | G > AG | 0,070825167 |
| **cg04131117** | chr2 | 216596529 | 216596529 | 1 | + | ENSG00000115457.10 | IGFBP2 | 36298 | 9,63640435089492E-07 | 0,001847448 | G > AG | 0,141889924 |
| **cg24803855** | chr4 | 182583176 | 182583176 | 1 | + | ENSG00000218336.9 | TENM3 | 439190 | 9,62229077868485E-07 | 0,001847448 | G > AG | 0,110046204 |
| **cg14750567** | chr17 | 64097240 | 64097240 | 1 | - | ENSG00000178607.17 | ERN 1,00 | 33580 | 9,78970461785462E-07 | 0,001862832 | G > AG | 0,138581224 |
| **cg11036959** | chr10 | 73946110 | 73946110 | 1 | - | ENSG00000148660.21 | CAMK2G | 71518 | 9,7896473033758E-07 | 0,001862832 | G > AG | 0,09917285 |
| **cg20307292** | chr2 | 98242132 | 98242132 | 1 | - | ENSG00000075568.17 | TMEM131 | 246183 | 9,77704626120948E-07 | 0,001862832 | G > AG | 0,105941555 |
| **cg10505571** | chr7 | 465244 | 465244 | 1 | - | ENSG00000197461.13 | PDGFA | 55053 | 9,83252964569391E-07 | 0,001866338 | G > AG | 0,0901422 |
| **cg08669766** | chr10 | 118257342 | 118257342 | 1 | + | ENSG00000170370.12 | EMX2 | 714898 | 1,00285983429367E-06 | 0,001894154 | G > AG | 0,108828202 |
| **cg19855622** | chr14 | 53209569 | 53209569 | 1 | + | ENSG00000198252.12 | STYX | 479404 | 1,0020171826311E-06 | 0,001894154 | G > AG | 0,107426685 |
| **cg00641105** | chr6 | 51514251 | 51514251 | 1 | - | ENSG00000170927.15 | PKHD1 | 573363 | 1,02861490427276E-06 | 0,001938014 | G > AG | 0,111767259 |
| **cg19587537** | chr15 | 95196208 | 95196208 | 1 | + | ENSG00000140563.15 | MCTP2 | 964671 | 1,03632556142493E-06 | 0,001947744 | G > AG | 0,105831814 |
| **cg07226436** | chr4 | 151807396 | 151807396 | 1 | + | ENSG00000164142.16 | FAM160A1 | 398221 | 1,03969975494046E-06 | 0,001949296 | G > AG | 0,103424728 |
| **cg25680635** | chr15 | 94925509 | 94925509 | 1 | + | ENSG00000140563.15 | MCTP2 | 693972 | 1,04922734670598E-06 | 0,00196235 | G > AG | 0,073971628 |
| **cg09698689** | chr8 | 85523760 | 85523760 | 1 | - | ENSG00000133742.14 | CA1 | 144745 | 1,0548115946732E-06 | 0,001962411 | G > AG | 0,093473769 |
| **cg00957695** | chr12 | 124174453 | 124174453 | 1 | - | ENSG00000119242.9 | CCDC92 | 201621 | 1,05944642234964E-06 | 0,001962411 | G > AG | 0,075898472 |
| **cg10436941** | chr18 | 14086177 | 14086177 | 1 | + | ENSG00000176136.6 | MC5R | 262029 | 1,05493074341368E-06 | 0,001962411 | G > AG | 0,078095917 |
| **cg10901380** | chr7 | 38306287 | 38306287 | 1 | + | ENSG00000010270.14 | STARD3NL | 128043 | 1,05952172372529E-06 | 0,001962411 | G > AG | 0,100798394 |
| **cg12024480** | chr11 | 97656648 | 97656648 | 1 | + | ENSG00000183340.7 | JRKL | 1266660 | 1,06814548047531E-06 | 0,001968849 | G > AG | 0,108840465 |
| **cg18121989** | chr8 | 133450029 | 133450029 | 1 | - | ENSG00000008513.16 | ST3GAL1 | 121912 | 1,06709429030759E-06 | 0,001968849 | G > AG | 0,106545133 |
| **cg18010348** | chr11 | 129284850 | 129284850 | 1 | + | ENSG00000043039.7 | BARX2 | 90997 | 1,07474803666556E-06 | 0,001976257 | G > AG | 0,132468521 |
| **cg17007772** | chr11 | 122494390 | 122494390 | 1 | + | ENSG00000154127.10 | UBASH3B | 161331 | 1,07934707142611E-06 | 0,001979954 | G > AG | 0,112359443 |
| **cg14097821** | chr11 | 40597784 | 40597784 | 1 | + | ENSG00000166181.13 | API5 | 2714178 | 1,08266599822667E-06 | 0,001981291 | G > AG | 0,084511433 |
| **cg23190689** | chr5 | 38468423 | 38468423 | 1 | + | ENSG00000164318.18 | EGFLAM | 210015 | 1,09074982127865E-06 | 0,001988286 | G > AG | 0,123263331 |
| **cg01707802** | chr5 | 125237187 | 125237187 | 1 | - | ENSG00000168916.16 | ZNF608 | 488379 | 1,09168660055798E-06 | 0,001988286 | G > AG | 0,078265841 |
| **cg27590148** | chrX | 132662968 | 132662968 | 1 | - | ENSG00000076770.15 | MBNL3 | 172999 | 1,09843508344539E-06 | 0,001995825 | G > AG | 0,149053642 |
| **cg06511395** | chr14 | 60003445 | 60003445 | 1 | + | ENSG00000131951.11 | LRRC9 | 83733 | 1,13286435108734E-06 | 0,002053504 | G > AG | 0,112478873 |

| | | | | | | | | | | | | |
|---|---|---|---|---|---|---|---|---|---|---|---|---|
| **cg20137991** | chr4 | 79050278 | 79050278 | 1 | + | ENSG00000138756.19 | BMP2K | 273937 | 1,13882921543941E-06 | 0,002054505 | G > AG | 0,178999962 |
| **cg12744758** | chr13 | 112913776 | 112913776 | 1 | + | ENSG00000126217.21 | MCF2L | 19399 | 1,13846310196588E-06 | 0,002054505 | G > AG | 0,087657348 |
| **cg23998629** | chr8 | 19495225 | 19495225 | 1 | - | ENSG00000147408.14 | CSGALNACT1 | 262805 | 1,18567267564575E-06 | 0,002129047 | G > AG | 0,149108275 |
| **cg04972669** | chr1 | 75343045 | 75343045 | 1 | - | ENSG00000137968.16 | SLC44A5 | 268072 | 1,19150375037289E-06 | 0,002134507 | G > AG | 0,129705186 |
| **cg11152829** | chr10 | 36188163 | 36188163 | 1 | - | ENSG00000177283.8 | FZD8 | 545866 | 1,20557742437195E-06 | 0,002153015 | G > AG | 0,064502987 |
| **cg00207329** | chr4 | 182212795 | 182212795 | 1 | - | ENSG00000129187.15 | DCTD | 705142 | 1,21333320110947E-06 | 0,002158449 | G > AG | 0,094297777 |
| **cg06632797** | chr11 | 69200748 | 69200748 | 1 | + | ENSG00000284713.1 | SMIM38 | 45271 | 1,24836829735448E-06 | 0,002215621 | G > AG | 0,073912159 |
| **cg26100256** | chr17 | 37274517 | 37274517 | 1 | - | ENSG00000278540.5 | ACACA | 132320 | 1,25373651835433E-06 | 0,002219998 | G > AG | 0,133173252 |
| **cg24411778** | chr5 | 60185660 | 60185660 | 1 | - | ENSG00000113448.19 | PDE4D | 336461 | 1,25910653186979E-06 | 0,002224358 | G > AG | 0,10631961 |
| **cg02985115** | chr18 | 36672573 | 36672573 | 1 | + | ENSG00000150477.15 | KIAA1328 | 156532 | 1,28299052796177E-06 | 0,002256131 | G > AG | 0,117063821 |
| **cg11340672** | chr8 | 126820264 | 126820264 | 1 | + | ENSG00000212993.5 | POU5F1B | 501918 | 1,28251782236218E-06 | 0,002256131 | G > AG | 0,100516573 |
| **cg23657388** | chr7 | 101528049 | 101528049 | 1 | + | ENSG00000160963.14 | COL26A1 | 165175 | 1,29094971974085E-06 | 0,00226492 | G > AG | 0,116962083 |
| **cg27582081** | chr3 | 119575969 | 119575969 | 1 | - | ENSG00000121594.12 | CD80 | 16354 | 1,30428288653274E-06 | 0,002277864 | G > AG | 0,081242442 |
| **cg17057626** | chr1 | 7538055 | 7538055 | 1 | - | ENSG00000049247.14 | UTS2 | 315458 | 1,30167476449727E-06 | 0,002277864 | G > AG | 0,07964133 |
| **cg09645658** | chr12 | 50196019 | 50196019 | 1 | - | ENSG00000139624.14 | CERS5 | 28485 | 1,32698338582264E-06 | 0,00231223 | G > AG | 0,124763309 |
| **cg03324951** | chr2 | 111396154 | 111396154 | 1 | + | ENSG00000153094.24 | BCL2L11 | 276777 | 1,33752735482363E-06 | 0,002315991 | G > AG | 0,11773907 |
| **cg01740356** | chr14 | 85703131 | 85703131 | 1 | - | ENSG00000054983.17 | GALC | 2290535 | 1,33822476855766E-06 | 0,002315991 | G > AG | 0,069483723 |
| **cg27543981** | chr7 | 132742280 | 132742280 | 1 | - | ENSG00000221866.9 | PLXNA4 | 93591 | 1,33521087420931E-06 | 0,002315991 | G > AG | 0,137841637 |
| **cg19898262** | chr3 | 41235487 | 41235487 | 1 | + | ENSG00000168036.18 | CTNNB1 | 40747 | 1,34137530571187E-06 | 0,002316203 | G > AG | 0,087751664 |
| **cg06055086** | chr8 | 105527662 | 105527662 | 1 | - | ENSG00000147650.12 | LRP12 | 938403 | 1,35215259522009E-06 | 0,002329554 | G > AG | 0,120070284 |
| **cg01027941** | chr3 | 56917570 | 56917570 | 1 | - | ENSG00000163947.12 | ARHGEF3 | 161760 | 1,37293406596475E-06 | 0,002347823 | G > AG | 0,132708799 |
| **cg26850677** | chr4 | 182789191 | 182789191 | 1 | - | ENSG00000129187.15 | DCTD | 128746 | 1,37036271930229E-06 | 0,002347823 | G > AG | 0,163184336 |
| **cg16783373** | chr5 | 60451545 | 60451545 | 1 | + | ENSG00000164182.12 | NDUFAF2 | 493631 | 1,37806350002408E-06 | 0,002347823 | G > AG | 0,105602619 |
| **cg26222722** | chr5 | 143445825 | 143445825 | 1 | - | ENSG00000113580.15 | NR3C1 | 10312 | 1,37446723252366E-06 | 0,002347823 | G > AG | 0,097895913 |
| **cg08187750** | chr12 | 130339984 | 130339984 | 1 | - | ENSG00000060709.15 | RIMBP2 | 376298 | 1,37810258356066E-06 | 0,002347823 | G > AG | 0,081464664 |
| **cg05496970** | chr2 | 239444820 | 239444820 | 1 | - | ENSG00000068024.17 | HDAC4 | 43165 | 1,39198388293886E-06 | 0,002356954 | G > AG | 0,094509484 |
| **cg04737625** | chr2 | 104514209 | 104514209 | 1 | + | ENSG00000198914.5 | POU3F3 | 339077 | 1,39270605319584E-06 | 0,002356954 | G > AG | 0,096298082 |
| **cg14033170** | chr7 | 28530414 | 28530414 | 1 | + | ENSG00000146592.17 | CREB5 | 231094 | 1,41086324156889E-06 | 0,002378283 | G > AG | 0,124014448 |
| **cg04943313** | chr3 | 53940106 | 53940106 | 1 | + | ENSG00000056736.10 | IL17RB | 93539 | 1,41614411307421E-06 | 0,002380818 | G > AG | 0,097338907 |
| **cg20584518** | chr15 | 71082989 | 71082989 | 1 | - | ENSG00000129028.9 | THAP10 | 190555 | 1,42191739320356E-06 | 0,002384651 | G > AG | 0,078380421 |
| **cg15547852** | chr6 | 11606506 | 11606506 | 1 | + | ENSG00000205269.6 | TMEM170B | 68758 | 1,42758438413672E-06 | 0,002384651 | G > AG | 0,127984886 |

| | | | | | | | | | | | | |
|---|---|---|---|---|---|---|---|---|---|---|---|---|
| **cg18050007** | chr12 | 108303148 | 108303148 | 1 | - | ENSG00000174600.14 | CMKLR1 | 36170 | 1,4400429168855E-06 | 0,002393393 | G > AG | 0,127507308 |
| **cg19540037** | chr2 | 130987925 | 130987925 | 1 | - | ENSG00000152102.18 | FAM168B | 105536 | 1,44239701255063E-06 | 0,002393393 | G > AG | 0,080159517 |
| **cg25837126** | chr5 | 37901279 | 37901279 | 1 | - | ENSG00000168621.15 | GDNF | 61237 | 1,43614419353219E-06 | 0,002393393 | G > AG | 0,114924526 |
| **cg03991547** | chr13 | 112160805 | 112160805 | 1 | + | ENSG00000182968.5 | SOX1 | 93657 | 1,44743670014072E-06 | 0,002396557 | G > AG | 0,076402343 |
| **cg25925441** | chr2 | 153479532 | 153479532 | 1 | - | ENSG00000177519.4 | RPRM | 769 | 1,4522642342058E-06 | 0,002399356 | G > AG | 0,099149115 |
| **cg17573903** | chr12 | 959431 | 959431 | 1 | - | ENSG00000002016.18 | RAD52 | 30623 | 1,45894141874129E-06 | 0,002403072 | G > AG | 0,06430547 |
| **cg11583041** | chr6 | 169340686 | 169340686 | 1 | + | ENSG00000185127.6 | C6orf120 | 361503 | 1,46953385952579E-06 | 0,002408541 | G > AG | 0,072383432 |
| **cg09529221** | chr6 | 136528081 | 136528081 | 1 | - | ENSG00000135525.19 | MAP7 | 22739 | 1,47041786743587E-06 | 0,002408541 | G > AG | 0,099586201 |
| **cg13492484** | chr9 | 21549616 | 21549616 | 1 | - | ENSG00000184995.7 | IFNE | 67302 | 1,48690754486293E-06 | 0,002425165 | G > AG | 0,140180318 |
| **cg13601550** | chrX | 20058188 | 20058188 | 1 | - | ENSG00000184368.16 | MAP7D2 | 58720 | 1,48380563796345E-06 | 0,002425165 | G > AG | 0,106398788 |
| **cg11945276** | chr4 | 1562020 | 1562020 | 1 | - | ENSG00000174137.13 | FAM53A | 122242 | 1,51423438555612E-06 | 0,002448849 | G > AG | 0,10073262 |
| **cg09644829** | chr8 | 66785695 | 66785695 | 1 | + | ENSG00000104205.16 | SGK3 | 72962 | 1,51344799679438E-06 | 0,002448849 | G > AG | 0,092096221 |
| **cg02831944** | chr2 | 54438487 | 54438487 | 1 | - | ENSG00000178021.11 | TSPYL6 | 182257 | 1,51407906434705E-06 | 0,002448849 | G > AG | 0,086688572 |
| **cg12184450** | chr17 | 37328147 | 37328147 | 1 | + | ENSG00000278505.5 | C17orf78 | 47837 | 1,5278999533066E-06 | 0,002458073 | G > AG | 0,069462396 |
| **cg07439336** | chr4 | 181662976 | 181662976 | 1 | - | ENSG00000129187.15 | DCTD | 1254961 | 1,52886733137512E-06 | 0,002458073 | G > AG | 0,116818264 |
| **cg23704362** | chr8 | 66493507 | 66493507 | 1 | - | ENSG00000185697.17 | MYBL1 | 120741 | 1,52957788854958E-06 | 0,002458073 | G > AG | 0,082374159 |
| **cg22506548** | chr1 | 3080385 | 3080385 | 1 | + | ENSG00000142611.17 | PRDM16 | 11218 | 1,53755393416375E-06 | 0,00246571 | G > AG | 0,104286791 |
| **cg15256973** | chr18 | 45517285 | 45517285 | 1 | - | ENSG00000152223.15 | EPG5 | 450045 | 1,54977518682505E-06 | 0,00248011 | G > AG | 0,098013236 |
| **cg24020009** | chr1 | 116849064 | 116849064 | 1 | + | ENSG00000134247.10 | PTGFRN | 60851 | 1,55681051939973E-06 | 0,002486167 | G > AG | 0,125190699 |
| **cg24077260** | chr4 | 182147264 | 182147264 | 1 | - | ENSG00000129187.15 | DCTD | 770673 | 1,5752972576432E-06 | 0,00250523 | G > AG | 0,082560021 |
| **cg04911969** | chr11 | 91308638 | 91308638 | 1 | - | ENSG00000110172.12 | CHORDC1 | 1085560 | 1,58536759101955E-06 | 0,002516014 | G > AG | 0,056944017 |
| **cg10515232** | chr2 | 239140657 | 239140657 | 1 | - | ENSG00000068024.17 | HDAC4 | 260998 | 1,60569535663194E-06 | 0,002537805 | G > AG | 0,122203715 |
| **cg06200710** | chr22 | 49503899 | 49503899 | 1 | + | ENSG00000100426.7 | ZBED4 | 349944 | 1,60573386629576E-06 | 0,002537805 | G > AG | 0,06702637 |
| **cg15733849** | chr9 | 84672079 | 84672079 | 1 | + | ENSG00000148053.17 | NTRK2 | 3529 | 1,63028178569295E-06 | 0,00257129 | G > AG | 0,099112147 |
| **cg09632065** | chr4 | 42673239 | 42673239 | 1 | + | ENSG00000215203.3 | GRXCR1 | 219473 | 1,6420245911463E-06 | 0,002579175 | G > AG | 0,080337006 |
| **cg17541779** | chr6 | 157693989 | 157693989 | 1 | + | ENSG00000130340.17 | SNX9 | 6397 | 1,64072525795265E-06 | 0,002579175 | G > AG | 0,072558283 |
| **cg25584787** | chr5 | 94358149 | 94358149 | 1 | + | ENSG00000133302.13 | SLF1 | 260519 | 1,65253470383003E-06 | 0,002590364 | G > AG | 0,111087339 |
| **cg10956362** | chr20 | 37591161 | 37591161 | 1 | - | ENSG00000166619.15 | BLCAP | 63229 | 1,6572119413403E-06 | 0,002592384 | G > AG | 0,09566029 |
| **cg20098452** | chr9 | 15735799 | 15735799 | 1 | - | ENSG00000164985.15 | PSIP1 | 224803 | 1,66788343857204E-06 | 0,002603753 | G > AG | 0,064759258 |
| **cg14909201** | chr13 | 111095673 | 111095673 | 1 | + | ENSG00000102606.19 | ARHGEF7 | 18885 | 1,67586287592549E-06 | 0,002603863 | G > AG | 0,102554901 |
| **cg05783974** | chr3 | 167981448 | 167981448 | 1 | - | ENSG00000173905.9 | GOLIM4 | 114477 | 1,67821793182616E-06 | 0,002603863 | G > AG | 0,096570997 |

| | | | | | | | | | | | | |
|---|---|---|---|---|---|---|---|---|---|---|---|---|
| **cg14300559** | chr11 | 133903068 | 133903068 | 1 | - | ENSG00000080854.16 | IGSF9B | 53901 | 1,68440166691285E-06 | 0,002603863 | G > AG | 0,071085949 |
| **cg25838711** | chr3 | 157920976 | 157920976 | 1 | - | ENSG00000168779.20 | SHOX2 | 185445 | 1,68837791844308E-06 | 0,002603863 | G > AG | 0,084064859 |
| **cg00002366** | chr4 | 90815784 | 90815784 | 1 | - | ENSG00000145335.17 | SNCA | 977468 | 1,67284096079553E-06 | 0,002603863 | G > AG | 0,125354377 |
| **cg04515263** | chr3 | 9311276 | 9311276 | 1 | - | ENSG00000196220.16 | SRGAP3 | 51778 | 1,68536683813487E-06 | 0,002603863 | G > AG | 0,118394573 |
| **cg19135279** | chr13 | 108111183 | 108111183 | 1 | + | ENSG00000139826.6 | ABHD13 | 107208 | 1,69263932384234E-06 | 0,002605182 | G > AG | 0,082394714 |
| **cg19412808** | chr1 | 69451077 | 69451077 | 1 | - | ENSG00000066557.6 | LRRC40 | 754503 | 1,70749009887195E-06 | 0,002615596 | G > AG | 0,09880885 |
| **cg00775463** | chr2 | 237559259 | 237559259 | 1 | - | ENSG00000124839.13 | RAB17 | 42356 | 1,70966306115699E-06 | 0,002615596 | G > AG | 0,093760688 |
| **cg08390519** | chr4 | 139795408 | 139795408 | 1 | - | ENSG00000145391.14 | SETD7 | 188708 | 1,71398090537869E-06 | 0,002616968 | G > AG | 0,098413156 |
| **cg13653793** | chr4 | 165384083 | 165384083 | 1 | + | ENSG00000109472.14 | CPE | 22890 | 1,72829039860619E-06 | 0,002633559 | G > AG | 0,080289437 |
| **cg23185751** | chr7 | 30680547 | 30680547 | 1 | - | ENSG00000106113.19 | CRHR2 | 19583 | 1,74493414951294E-06 | 0,00264837 | G > AG | 0,071686416 |
| **cg04824591** | chr3 | 183121962 | 183121962 | 1 | - | ENSG00000078070.13 | MCCC1 | 5886 | 1,75197556551456E-06 | 0,002653791 | G > AG | 0,070538724 |
| **cg00930430** | chr10 | 125690296 | 125690296 | 1 | + | ENSG00000107938.18 | EDRF1 | 29218 | 1,76304517884095E-06 | 0,002665281 | G > AG | 0,131298664 |
| **cg23540393** | chr18 | 49614189 | 49614189 | 1 | - | ENSG00000265681.8 | RPL17 | 121680 | 1,77907497319177E-06 | 0,002684209 | G > AG | 0,096965212 |
| **cg02758922** | chr13 | 72508561 | 72508561 | 1 | + | ENSG00000136122.18 | BORA | 219187 | 1,79284257733956E-06 | 0,002699657 | G > AG | 0,086038515 |
| **cg22011557** | chr8 | 52629081 | 52629081 | 1 | - | ENSG00000196711.9 | ALKAL1 | 63650 | 1,8006470924899E-06 | 0,002706082 | G > AG | 0,116570921 |
| **cg08174210** | chr10 | 16492410 | 16492410 | 1 | + | ENSG00000165983.15 | PTER | 55468 | 1,81506590539626E-06 | 0,002717075 | G > AG | 0,090756654 |
| **cg10415870** | chr6 | 165937202 | 165937202 | 1 | - | ENSG00000112541.18 | PDE10A | 50877 | 1,82270182585708E-06 | 0,002723176 | G > AG | 0,08898348 |
| **cg10178907** | chr7 | 156659007 | 156659007 | 1 | + | ENSG00000105982.17 | RNF32 | 18727 | 1,86637582804506E-06 | 0,002782991 | G > AG | 0,079178409 |
| **cg10134641** | chr4 | 78168276 | 78168276 | 1 | + | ENSG00000138759.20 | FRAS1 | 110954 | 1,88147939415463E-06 | 0,002800054 | G > AG | 0,062899406 |
| **cg27050407** | chr6 | 24832500 | 24832500 | 1 | + | ENSG00000112312.10 | GMNN | 57570 | 1,90701978062843E-06 | 0,002832553 | G > AG | 0,074988246 |
| **cg22473662** | chr17 | 37068394 | 37068394 | 1 | + | ENSG00000275700.6 | AATF | 119470 | 1,9111065344911E-06 | 0,002833122 | G > AG | 0,108622087 |
| **cg03407080** | chr2 | 120597139 | 120597139 | 1 | + | ENSG00000074047.22 | GLI2 | 138483 | 1,93779824707868E-06 | 0,002867135 | G > AG | 0,094841294 |
| **cg24083720** | chr2 | 123687844 | 123687844 | 1 | - | ENSG00000155438.12 | NIFK | 1950932 | 1,94506851980889E-06 | 0,002872336 | G > AG | 0,064234286 |
| **cg01333785** | chr5 | 65364482 | 65364482 | 1 | + | ENSG00000113593.12 | PPWD1 | 198753 | 1,9547602385563E-06 | 0,002881086 | G > AG | 0,083230989 |
| **cg12548176** | chr12 | 55246329 | 55246329 | 1 | - | ENSG00000188324.4 | OR6C6 | 50241 | 1,96036425319457E-06 | 0,002883789 | G > AG | 0,081092387 |
| **cg21142398** | chr1 | 53734405 | 53734405 | 1 | - | ENSG00000174332.5 | GLIS1 | 3702 | 1,97493177108214E-06 | 0,002899643 | G > AG | 0,068419579 |
| **cg21854617** | chr8 | 48683091 | 48683091 | 1 | + | ENSG00000168333.14 | PPDPFL | 371219 | 2,00879056879717E-06 | 0,002938076 | G > AG | 0,090423332 |
| **cg23989584** | chr11 | 120167813 | 120167813 | 1 | - | ENSG00000137699.17 | TRIM29 | 17717 | 2,01341671866322E-06 | 0,002939223 | G > AG | 0,091369782 |
| **cg02821895** | chr8 | 122619648 | 122619648 | 1 | - | ENSG00000136986.10 | DERL1 | 422655 | 2,0266953512196E-06 | 0,002952972 | G > AG | 0,119813259 |
| **cg05572616** | chr4 | 86841843 | 86841843 | 1 | - | ENSG00000145283.8 | SLC10A6 | 7542 | 2,03332427589835E-06 | 0,002956998 | G > AG | 0,117144192 |
| **cg17216165** | chr3 | 11434310 | 11434310 | 1 | + | ENSG00000197548.12 | ATG7 | 162002 | 2,06816181572458E-06 | 0,003001954 | G > AG | 0,09978709 |

| | | | | | | | | | | | | |
|---|---|---|---|---|---|---|---|---|---|---|---|---|
| **cg23790554** | chr8 | 1439327 | 1439327 | 1 | - | ENSG00000104714.14 | ERICH1 | 701220 | 2,09615276483005E-06 | 0,003023686 | G > AG | 0,078060713 |
| **cg05028307** | chr10 | 26252082 | 26252082 | 1 | + | ENSG00000136750.13 | GAD2 | 35418 | 2,11080385020789E-06 | 0,003023686 | G > AG | 0,079940983 |
| **cg19892363** | chr14 | 94868151 | 94868151 | 1 | + | ENSG00000196136.18 | SERPINA3 | 255768 | 2,10653719441881E-06 | 0,003023686 | G > AG | 0,087408693 |
| **cg11023224** | chr12 | 129446879 | 129446879 | 1 | - | ENSG00000151952.16 | TMEM132D | 457147 | 2,10979016836025E-06 | 0,003023686 | G > AG | 0,06978176 |
| **cg09341695** | chr6 | 20545073 | 20545073 | 1 | - | ENSG00000172197.11 | MBOAT1 | 332603 | 2,13214130932485E-06 | 0,003048543 | G > AG | 0,084153718 |
| **cg11142925** | chr10 | 55312118 | 55312118 | 1 | - | ENSG00000150275.20 | PCDH15 | 315825 | 2,17607593582695E-06 | 0,003099773 | G > AG | 0,098094397 |
| **cg16660677** | chr1 | 169734091 | 169734091 | 1 | - | ENSG00000188404.10 | SELL | 22388 | 2,17354430053192E-06 | 0,003099773 | G > AG | 0,119613405 |
| **cg20575845** | chr18 | 21303837 | 21303837 | 1 | - | ENSG00000067900.8 | ROCK1 | 192023 | 2,20418307430636E-06 | 0,003122367 | G > AG | 0,10357996 |
| **cg03181006** | chr4 | 61462768 | 61462768 | 1 | - | ENSG00000205678.8 | TECRL | 2946701 | 2,20266448533289E-06 | 0,003122367 | G > AG | 0,146679409 |
| **cg22910862** | chr8 | 90110397 | 90110397 | 1 | + | ENSG00000104325.7 | DECR1 | 108993 | 2,23851654275031E-06 | 0,003165142 | G > AG | 0,090291827 |
| **cg08036078** | chr2 | 170463000 | 170463000 | 1 | + | ENSG00000204335.4 | SP5 | 252336 | 2,24578061224931E-06 | 0,003169554 | G > AG | 0,105556476 |
| **cg01593596** | chr3 | 11496238 | 11496238 | 1 | + | ENSG00000197548.12 | ATG7 | 223930 | 2,25574258051092E-06 | 0,003177751 | G > AG | 0,074135942 |
| **cg06779606** | chr7 | 3001445 | 3001445 | 1 | - | ENSG00000198286.10 | CARD11 | 42423 | 2,26800369123168E-06 | 0,00318915 | G > AG | 0,105082954 |
| **cg16911196** | chr11 | 113908394 | 113908394 | 1 | - | ENSG00000048028.11 | USP28 | 32823 | 2,28861179912531E-06 | 0,003200478 | G > AG | 0,101792292 |
| **cg22846582** | chr12 | 52454430 | 52454430 | 1 | - | ENSG00000185479.6 | KRT6B | 2283 | 2,3012282373112E-06 | 0,003206398 | G > AG | 0,109568829 |
| **cg25930440** | chr8 | 143140921 | 143140921 | 1 | - | ENSG00000176956.13 | LY6H | 19791 | 2,31396992033575E-06 | 0,00321829 | G > AG | 0,098841858 |
| **cg11662428** | chr7 | 126657973 | 126657973 | 1 | - | ENSG00000179603.18 | GRM8 | 595121 | 2,32217561011106E-06 | 0,003223841 | G > AG | 0,100597175 |
| **cg24877842** | chr3 | 42160406 | 42160406 | 1 | - | ENSG00000187094.12 | CCK | 105780 | 2,327805322694E-06 | 0,003225802 | G > AG | 0,131612185 |
| **cg00153406** | chr1 | 7091877 | 7091877 | 1 | + | ENSG00000171735.19 | CAMTA1 | 306424 | 2,33388643555467E-06 | 0,00322838 | G > AG | 0,107519881 |
| **cg05369808** | chr3 | 114417185 | 114417185 | 1 | + | ENSG00000181847.12 | TIGIT | 140273 | 2,36475624280535E-06 | 0,003265177 | G > AG | 0,091852234 |
| **cg11134801** | chr14 | 101062017 | 101062017 | 1 | + | ENSG00000185559.16 | DLK1 | 336313 | 2,37738562105121E-06 | 0,003276701 | G > AG | 0,084360628 |
| **cg12989634** | chr3 | 82294927 | 82294927 | 1 | + | ENSG00000175161.14 | CADM2 | 2664061 | 2,38708274869472E-06 | 0,003278252 | G > AG | 0,098976231 |
| **cg12221623** | chr6 | 84688192 | 84688192 | 1 | + | ENSG00000135324.6 | MRAP2 | 654421 | 2,38676011723785E-06 | 0,003278252 | G > AG | 0,118828236 |
| **cg14648289** | chr4 | 152595709 | 152595709 | 1 | + | ENSG00000164144.16 | ARFIP1 | 184227 | 2,40189249631532E-06 | 0,003289646 | G > AG | 0,104785406 |
| **cg14300943** | chr9 | 27534291 | 27534291 | 1 | - | ENSG00000120162.10 | MOB3B | 4476 | 2,40558512112713E-06 | 0,003289646 | G > AG | 0,099298459 |
| **cg15463580** | chr19 | 6113934 | 6113934 | 1 | - | ENSG00000087903.13 | RFX2 | 85639 | 2,41221611046911E-06 | 0,003289646 | G > AG | 0,087455182 |
| **cg20023163** | chr14 | 103881558 | 103881558 | 1 | + | ENSG00000156414.19 | TDRD9 | 46897 | 2,4125808551064E-06 | 0,003289646 | G > AG | 0,100663261 |
| **cg14732655** | chr18 | 73519062 | 73519062 | 1 | + | ENSG00000075336.12 | TIMM21 | 629460 | 2,43301967063951E-06 | 0,003311612 | G > AG | 0,112549537 |
| **cg03397816** | chr3 | 15505551 | 15505551 | 1 | - | ENSG00000206561.14 | COLQ | 16201 | 2,44410377197767E-06 | 0,003314902 | G > AG | 0,082383806 |
| **cg24843615** | chr2 | 17815828 | 17815828 | 1 | + | ENSG00000151379.4 | MSGN1 | 631 | 2,44237062310403E-06 | 0,003314902 | G > AG | 0,133930205 |
| **cg11022539** | chr8 | 138600799 | 138600799 | 1 | - | ENSG00000147724.12 | FAM135B | 103537 | 2,45609964955355E-06 | 0,0033194 | G > AG | 0,113334694 |

| | | | | | | | | | | | | |
|---|---|---|---|---|---|---|---|---|---|---|---|---|
| **cg10104252** | chr12 | 3454733 | 3454733 | 1 | - | ENSG00000130038.10 | CRACR2A | 310087 | 2,4664852851378E-06 | 0,003327558 | G > AG | 0,113392192 |
| **cg08160063** | chr16 | 7654959 | 7654959 | 1 | - | ENSG00000232258.6 | TMEM114 | 935235 | 2,49413769907217E-06 | 0,003355092 | G > AG | 0,070986504 |
| **cg10480055** | chr6 | 125492565 | 125492565 | 1 | - | ENSG00000111906.18 | HDDC2 | 190486 | 2,52576159890762E-06 | 0,003389593 | G > AG | 0,080241726 |
| **cg09334731** | chr1 | 6960747 | 6960747 | 1 | + | ENSG00000171735.19 | CAMTA1 | 175294 | 2,53365083534583E-06 | 0,003394226 | G > AG | 0,115341512 |
| **cg04345513** | chr2 | 236814780 | 236814780 | 1 | + | ENSG00000144476.6 | ACKR3 | 246994 | 2,56679208063563E-06 | 0,003420652 | G > AG | 0,095952775 |
| **cg03962250** | chr13 | 95559303 | 95559303 | 1 | + | ENSG00000102580.15 | DNAJC3 | 117835 | 2,56396502362114E-06 | 0,003420652 | G > AG | 0,106503043 |
| **cg02216535** | chr4 | 84387776 | 84387776 | 1 | - | ENSG00000163623.10 | NKX6-1 | 111517 | 2,56589077429934E-06 | 0,003420652 | G > AG | 0,096878532 |
| **cg14547509** | chr9 | 14318499 | 14318499 | 1 | + | ENSG00000164975.15 | SNAPC3 | 1104204 | 2,60969597434581E-06 | 0,003465752 | G > AG | 0,112833752 |
| **cg17017848** | chr11 | 15629408 | 15629408 | 1 | - | ENSG00000110680.13 | CALCA | 657056 | 2,62426995686371E-06 | 0,003473048 | G > AG | 0,060228417 |
| **cg16123687** | chr10 | 12702627 | 12702627 | 1 | - | ENSG00000151468.11 | CCDC3 | 397026 | 2,62009115976502E-06 | 0,003473048 | G > AG | 0,071651146 |
| **cg23987565** | chr15 | 83180390 | 83180390 | 1 | - | ENSG00000166503.9 | HDGFL3 | 27434 | 2,65542089327606E-06 | 0,003478756 | G > AG | 0,081581194 |
| **cg12661889** | chr8 | 65419534 | 65419534 | 1 | + | ENSG00000066855.16 | MTFR1 | 225199 | 2,6432568921589E-06 | 0,003478756 | G > AG | 0,061606257 |
| **cg05691475** | chr1 | 188778730 | 188778730 | 1 | + | ENSG00000116711.10 | PLA2G4A | 1949782 | 2,64859427632985E-06 | 0,003478756 | G > AG | 0,089767039 |
| **cg02419554** | chr2 | 10194379 | 10194379 | 1 | + | ENSG00000171848.16 | RRM2 | 73682 | 2,6453962374136E-06 | 0,003478756 | G > AG | 0,070090302 |
| **cg22894655** | chr17 | 71927080 | 71927080 | 1 | + | ENSG00000125398.8 | SOX9 | 193939 | 2,65932036094811E-06 | 0,003478756 | G > AG | 0,093987066 |
| **cg13440411** | chr5 | 150578362 | 150578362 | 1 | + | ENSG00000171992.13 | SYNPO | 22717 | 2,65322265491937E-06 | 0,003478756 | G > AG | 0,043149445 |
| **cg11426250** | chr17 | 10099247 | 10099247 | 1 | - | ENSG00000007237.19 | GAS7 | 99360 | 2,6652155015646E-06 | 0,003479083 | G > AG | 0,099381165 |
| **cg05804170** | chr1 | 3204950 | 3204950 | 1 | - | ENSG00000215912.12 | TTC34 | 403232 | 2,67213225099092E-06 | 0,00348217 | G > AG | 0,086236669 |
| **cg27422141** | chr13 | 67229119 | 67229119 | 1 | + | ENSG00000237378.3 | RP11-473M10.3 | 3482379 | 2,6778805872144E-06 | 0,003483726 | G > AG | 0,110556789 |
| **cg02632952** | chr3 | 190708735 | 190708735 | 1 | - | ENSG00000205835.9 | GMNC | 183695 | 2,70263682586815E-06 | 0,003509963 | G > AG | 0,132900612 |
| **cg13407144** | chr10 | 33179409 | 33179409 | 1 | + | ENSG00000216937.13 | CCDC7 | 733270 | 2,72852513404563E-06 | 0,003513756 | G > AG | 0,087275704 |
| **cg20741534** | chr15 | 85479267 | 85479267 | 1 | - | ENSG00000183655.13 | KLHL25 | 315659 | 2,72613838737117E-06 | 0,003513756 | G > AG | 0,097354966 |
| **cg25870763** | chr21 | 39667282 | 39667282 | 1 | - | ENSG00000157578.14 | LCA5L | 221476 | 2,71874808582416E-06 | 0,003513756 | G > AG | 0,085870042 |
| **cg07749951** | chr16 | 79318539 | 79318539 | 1 | - | ENSG00000178573.7 | MAF | 282199 | 2,71197090637537E-06 | 0,003513756 | G > AG | 0,104539973 |
| **cg24355809** | chr17 | 843334 | 843334 | 1 | + | ENSG00000171861.11 | MRM3 | 60982 | 2,73975464727272E-06 | 0,003522288 | G > AG | 0,079665938 |
| **cg10309230** | chr13 | 99590260 | 99590260 | 1 | - | ENSG00000169508.7 | GPR183 | 282860 | 2,75153777361954E-06 | 0,003531501 | G > AG | 0,063218825 |
| **cg11710027** | chr2 | 143399495 | 143399495 | 1 | - | ENSG00000121964.15 | GTDC1 | 933074 | 2,7570558189768E-06 | 0,003532656 | G > AG | 0,106768471 |
| **cg18027452** | chr12 | 105825228 | 105825228 | 1 | + | ENSG00000166046.11 | TCP11L2 | 476700 | 2,7694129008529E-06 | 0,003542555 | G > AG | 0,078009337 |
| **cg17202519** | chr11 | 84923537 | 84923537 | 1 | - | ENSG00000150672.18 | DLG2 | 704386 | 2,79492327442135E-06 | 0,003543174 | G > AG | 0,115523002 |
| **cg15089567** | chr7 | 1917681 | 1917681 | 1 | - | ENSG00000002822.16 | MAD1L1 | 315563 | 2,78736561834253E-06 | 0,003543174 | G > AG | 0,140256583 |
| **cg25590081** | chr12 | 95551722 | 95551722 | 1 | + | ENSG00000111142.14 | METAP2 | 78203 | 2,78413731331831E-06 | 0,003543174 | G > AG | 0,114910961 |

| | | | | | | | | | | | | |
|---|---|---|---|---|---|---|---|---|---|---|---|---|
| **cg19269749** | chr2 | 127193838 | 127193838 | 1 | + | ENSG00000115718.18 | PROC | 224588 | 2,79726188235712E-06 | 0,003543174 | G > AG | 0,105929106 |
| **cg17707550** | chr6 | 153059280 | 153059280 | 1 | + | ENSG00000146469.13 | VIP | 308484 | 2,7770824696009E-06 | 0,003543174 | G > AG | 0,131427766 |
| **cg00991554** | chr1 | 82185357 | 82185357 | 1 | - | ENSG00000137941.17 | TTLL7 | 1813794 | 2,80236847632032E-06 | 0,003543235 | G > AG | 0,091463754 |
| **cg25343783** | chr20 | 47972550 | 47972550 | 1 | + | ENSG00000124151.19 | NCOA3 | 470664 | 2,81795504685833E-06 | 0,003557063 | G > AG | 0,121323196 |
| **cg24969531** | chr20 | 10741366 | 10741366 | 1 | - | ENSG00000101384.12 | JAG1 | 67366 | 2,82756188060416E-06 | 0,003563309 | G > AG | 0,067959416 |
| **cg14290628** | chr13 | 109495286 | 109495286 | 1 | - | ENSG00000185950.9 | IRS2 | 291298 | 2,84576626970636E-06 | 0,003580352 | G > AG | 0,114884688 |
| **cg20576349** | chr8 | 105686290 | 105686290 | 1 | + | ENSG00000164830.19 | OXR1 | 583853 | 2,8712492483062E-06 | 0,003606481 | G > AG | 0,090048783 |
| **cg18523616** | chr13 | 25820130 | 25820130 | 1 | + | ENSG00000132964.12 | CDK8 | 433973 | 2,88279003336919E-06 | 0,003615041 | G > AG | 0,069041264 |
| **cg06571834** | chr13 | 48486313 | 48486313 | 1 | - | ENSG00000139679.16 | LPAR6 | 41608 | 2,89959875620118E-06 | 0,003630169 | G > AG | 0,091253666 |
| **cg19175123** | chr10 | 101651989 | 101651989 | 1 | + | ENSG00000166171.13 | DPCD | 81430 | 2,90959773500468E-06 | 0,003636735 | G > AG | 0,075699477 |
| **cg16540350** | chr11 | 68774395 | 68774395 | 1 | + | ENSG00000069482.7 | GAL | 90617 | 2,91769875653334E-06 | 0,003640911 | G > AG | 0,088604444 |
| **cg13157780** | chr8 | 133831479 | 133831479 | 1 | + | ENSG00000104415.14 | CCN4 | 640441 | 2,93249801713076E-06 | 0,003653419 | G > AG | 0,074660032 |
| **cg11372668** | chr16 | 67987672 | 67987672 | 1 | + | ENSG00000167264.18 | DUS2 | 73 | 2,97467832186837E-06 | 0,003699943 | G > AG | 0,073102697 |
| **cg13709646** | chr9 | 134922099 | 134922099 | 1 | + | ENSG00000160339.16 | FCN2 | 41290 | 3,00646729321948E-06 | 0,003720575 | G > AG | 0,087422225 |
| **cg06418837** | chr1 | 211474114 | 211474114 | 1 | - | ENSG00000198570.7 | RD3 | 18049 | 3,00459724857237E-06 | 0,003720575 | G > AG | 0,065371204 |
| **cg13036445** | chr5 | 38468715 | 38468715 | 1 | + | ENSG00000164318.18 | EGFLAM | 210307 | 3,02789612735182E-06 | 0,003724312 | G > AG | 0,113565514 |
| **cg03600196** | chr2 | 142920886 | 142920886 | 1 | + | ENSG00000115919.15 | KYNU | 43230 | 3,02315603254703E-06 | 0,003724312 | G > AG | 0,087207596 |
| **cg20143026** | chr15 | 84853282 | 84853282 | 1 | - | ENSG00000140612.14 | SEC11A | 136821 | 3,02835225780788E-06 | 0,003724312 | G > AG | 0,086382327 |
| **cg23664795** | chr10 | 92751213 | 92751213 | 1 | + | ENSG00000152804.11 | HHEX | 61259 | 3,06217432826049E-06 | 0,003759863 | G > AG | 0,108642052 |
| **cg10725555** | chr5 | 71748482 | 71748482 | 1 | - | ENSG00000113048.17 | MRPS27 | 572165 | 3,08038883918059E-06 | 0,003770124 | G > AG | 0,09919538 |
| **cg26154608** | chr22 | 25729139 | 25729139 | 1 | + | ENSG00000133454.16 | MYO18B | 13004 | 3,10168287789702E-06 | 0,003790122 | G > AG | 0,073511284 |
| **cg09263768** | chr7 | 134284206 | 134284206 | 1 | - | ENSG00000205060.11 | SLC35B4 | 32725 | 3,11290536335594E-06 | 0,003797768 | G > AG | 0,107522857 |
| **cg24693865** | chr17 | 16372060 | 16372060 | 1 | + | ENSG00000170315.14 | UBB | 8737 | 3,11794892549201E-06 | 0,003797864 | G > AG | 0,088131322 |
| **cg17145219** | chr16 | 66511279 | 66511279 | 1 | - | ENSG00000260851.7 | RP11-403P17.5 | 6034 | 3,13101339794628E-06 | 0,003805146 | G > AG | 0,087870262 |
| **cg13359296** | chr9 | 3956347 | 3956347 | 1 | + | ENSG00000106688.12 | SLC1A1 | 534120 | 3,1338755899818E-06 | 0,003805146 | G > AG | 0,067478094 |
| **cg19664566** | chr14 | 71248310 | 71248310 | 1 | + | ENSG00000197555.10 | SIPA1L1 | 72138 | 3,14196028234105E-06 | 0,003808916 | G > AG | 0,100469013 |
| **cg15782751** | chr10 | 129708415 | 129708415 | 1 | + | ENSG00000170430.10 | MGMT | 241226 | 3,1716902461384E-06 | 0,003838873 | G > AG | 0,183523864 |
| **cg26120372** | chr2 | 223489207 | 223489207 | 1 | - | ENSG00000171951.5 | SCG2 | 113155 | 3,1917255704846E-06 | 0,00385702 | G > AG | 0,103830768 |
| **cg10663897** | chr2 | 161123641 | 161123641 | 1 | - | ENSG00000153250.20 | RBMS1 | 629833 | 3,21096674991858E-06 | 0,003874152 | G > AG | 0,118170707 |
| **cg04427640** | chr7 | 100948182 | 100948182 | 1 | - | ENSG00000087085.16 | ACHE | 51207 | 3,25290067642782E-06 | 0,003887952 | G > AG | 0,121553146 |
| **cg00892802** | chr1 | 66563917 | 66563917 | 1 | - | ENSG00000172410.5 | INSL5 | 237360 | 3,24890941966331E-06 | 0,003887952 | G > AG | 0,103880659 |

| | | | | | | | | | | | | |
|---|---|---|---|---|---|---|---|---|---|---|---|---|
| **cg00375025** | chr3 | 156143143 | 156143143 | 1 | + | ENSG00000169282.18 | KCNAB1 | 105443 | 3,23882130965138E-06 | 0,003887952 | G > AG | 0,069100301 |
| **cg17973038** | chr8 | 98382840 | 98382840 | 1 | + | ENSG00000156486.8 | KCNS2 | 44117 | 3,25027772006228E-06 | 0,003887952 | G > AG | 0,09882269 |
| **cg18048027** | chr2 | 129744959 | 129744959 | 1 | - | ENSG00000196604.13 | POTEF | 384264 | 3,24615330456831E-06 | 0,003887952 | G > AG | 0,087995562 |
| **cg09162970** | chr6 | 11606495 | 11606495 | 1 | + | ENSG00000205269.6 | TMEM170B | 68747 | 3,22962023656811E-06 | 0,003887952 | G > AG | 0,123987248 |
| **cg03028721** | chr6 | 121435657 | 121435657 | 1 | + | ENSG00000152661.9 | GJA1 | 63 | 3,26640060271288E-06 | 0,003897997 | G > AG | 0,154473159 |
| **cg07734108** | chr5 | 16727795 | 16727795 | 1 | + | ENSG00000176788.9 | BASP1 | 337802 | 3,2811259148357E-06 | 0,003909471 | G > AG | 0,10711943 |
| **cg02176460** | chr17 | 80169998 | 80169998 | 1 | + | ENSG00000141527.18 | CARD14 | 7 | 3,30429016771701E-06 | 0,003930948 | G > AG | 0,107246253 |
| **cg12733117** | chr8 | 74284581 | 74284581 | 1 | + | ENSG00000104381.14 | GDAP1 | 36031 | 3,31121814703352E-06 | 0,003933073 | G > AG | 0,106003889 |
| **cg19432505** | chr4 | 176269086 | 176269086 | 1 | - | ENSG00000164122.9 | ASB5 | 8486 | 3,35431391262698E-06 | 0,003956912 | G > AG | 0,067925656 |
| **cg08868674** | chr19 | 41159471 | 41159471 | 1 | + | ENSG00000167600.14 | CYP2S1 | 33738 | 3,34931334218522E-06 | 0,003956912 | G > AG | 0,081383393 |
| **cg14638934** | chr10 | 8591671 | 8591671 | 1 | - | ENSG00000151657.12 | KIN | 803677 | 3,35478389727653E-06 | 0,003956912 | G > AG | 0,104898967 |
| **cg06318355** | chr6 | 124476323 | 124476323 | 1 | + | ENSG00000146373.17 | RNF217 | 486113 | 3,35715164669452E-06 | 0,003956912 | G > AG | 0,120576035 |
| **cg10045883** | chr6 | 123352836 | 123352836 | 1 | - | ENSG00000186439.14 | TRDN | 284258 | 3,36412193349816E-06 | 0,003959027 | G > AG | 0,109846403 |
| **cg17847459** | chr6 | 157506713 | 157506713 | 1 | - | ENSG00000215712.11 | TMEM242 | 183111 | 3,37310356771791E-06 | 0,003963499 | G > AG | 0,102393193 |
| **cg01654432** | chr10 | 61475589 | 61475589 | 1 | + | ENSG00000183346.9 | CABCOCO1 | 187339 | 3,37995854449042E-06 | 0,003964093 | G > AG | 0,072766882 |
| **cg05556849** | chr14 | 22279915 | 22279915 | 1 | - | ENSG00000129562.11 | DAD1 | 309310 | 3,38397298157875E-06 | 0,003964093 | G > AG | 0,091393828 |
| **cg04215000** | chr2 | 225117583 | 225117583 | 1 | - | ENSG00000135905.20 | DOCK10 | 75114 | 3,39675160816176E-06 | 0,003966912 | G > AG | 0,062966935 |
| **cg15158924** | chr4 | 21709325 | 21709325 | 1 | - | ENSG00000185774.16 | KCNIP4 | 239448 | 3,39299033092187E-06 | 0,003966912 | G > AG | 0,111745201 |
| **cg06573088** | chr6 | 17492490 | 17492490 | 1 | + | ENSG00000112186.12 | CAP2 | 98986 | 3,40745717956675E-06 | 0,003967741 | G > AG | 0,107278167 |
| **cg17260002** | chr17 | 6799945 | 6799945 | 1 | + | ENSG00000177294.7 | FBXO39 | 23731 | 3,40826752860026E-06 | 0,003967741 | G > AG | 0,104920731 |
| **cg02374420** | chr2 | 6370933 | 6370933 | 1 | + | ENSG00000134321.13 | RSAD2 | 494623 | 3,41302188819096E-06 | 0,003967741 | G > AG | 0,075676755 |
| **cg18014277** | chr10 | 26567735 | 26567735 | 1 | + | ENSG00000077420.16 | APBB1IP | 129395 | 3,44183406383788E-06 | 0,003983289 | G > AG | 0,106375675 |
| **cg16094493** | chr7 | 43467753 | 43467753 | 1 | - | ENSG00000106603.20 | COA1 | 261965 | 3,447225922323E-06 | 0,003983289 | G > AG | 0,084632299 |
| **cg05900542** | chr3 | 46356366 | 46356366 | 1 | - | ENSG00000012223.13 | LTF | 128869 | 3,44263612274343E-06 | 0,003983289 | G > AG | 0,085599849 |
| **cg03133301** | chr13 | 94128597 | 94128597 | 1 | - | ENSG00000080166.16 | DCT | 351086 | 3,45719741366464E-06 | 0,003988786 | G > AG | 0,109477202 |
| **cg23309843** | chr5 | 127652133 | 127652133 | 1 | - | ENSG00000164241.14 | C5orf63 | 578626 | 3,48039628604624E-06 | 0,004009505 | G > AG | 0,126260777 |
| **cg08782448** | chr16 | 9567829 | 9567829 | 1 | + | ENSG00000182831.12 | C16orf72 | 476186 | 3,53015669081852E-06 | 0,004057533 | G > AG | 0,124932428 |
| **cg25278456** | chr11 | 7539161 | 7539161 | 1 | - | ENSG00000166394.15 | CYB5R2 | 138062 | 3,54860867616004E-06 | 0,004057533 | G > AG | 0,094659364 |
| **cg08086873** | chr8 | 26007352 | 26007352 | 1 | - | ENSG00000221818.9 | EBF2 | 38062 | 3,54361975405694E-06 | 0,004057533 | G > AG | 0,082893171 |
| **cg01670430** | chr7 | 2733376 | 2733376 | 1 | - | ENSG00000146535.14 | GNA12 | 110933 | 3,54545386017879E-06 | 0,004057533 | G > AG | 0,083190456 |
| **cg06548300** | chr13 | 101840954 | 101840954 | 1 | - | ENSG00000102452.18 | NALCN | 424445 | 3,57894614918163E-06 | 0,004077286 | G > AG | 0,108632789 |

| | | | | | | | | | | | | |
|---|---|---|---|---|---|---|---|---|---|---|---|---|
| **cg24693836** | chr22 | 43706693 | 43706693 | 1 | + | ENSG00000100344.11 | PNPLA3 | 217098 | 3,5755719241252E-06 | 0,004077286 | G > AG | 0,091050335 |
| **cg24091191** | chr16 | 12440011 | 12440011 | 1 | + | ENSG00000237515.9 | SHISA9 | 461586 | 3,58187468518115E-06 | 0,004077286 | G > AG | 0,09909436 |
| **cg02717484** | chr3 | 32894061 | 32894061 | 1 | + | ENSG00000183813.7 | CCR4 | 57582 | 3,60559246910614E-06 | 0,004083429 | G > AG | 0,115910013 |
| **cg16223546** | chr5 | 159341968 | 159341968 | 1 | - | ENSG00000113302.5 | IL12B | 11480 | 3,60021419846008E-06 | 0,004083429 | G > AG | 0,121202028 |
| **cg25596109** | chr20 | 64196983 | 64196983 | 1 | - | ENSG00000125522.4 | NPBWR2 | 89417 | 3,61071532527338E-06 | 0,004083429 | G > AG | 0,072068904 |
| **cg13326076** | chr12 | 75982472 | 75982472 | 1 | - | ENSG00000139289.14 | PHLDA1 | 49305 | 3,61396210224417E-06 | 0,004083429 | G > AG | 0,097563531 |
| **cg16326962** | chr1 | 183270753 | 183270753 | 1 | + | ENSG00000058085.15 | LAMC2 | 84516 | 3,63381305810919E-06 | 0,004099803 | G > AG | 0,123749476 |
| **cg08325898** | chr8 | 70184664 | 70184664 | 1 | - | ENSG00000147596.4 | PRDM14 | 112970 | 3,66122437285375E-06 | 0,004124646 | G > AG | 0,146401804 |
| **cg19623020** | chr9 | 16708841 | 16708841 | 1 | + | ENSG00000044459.15 | CNTLN | 426140 | 3,67478409936965E-06 | 0,004127764 | G > AG | 0,081729823 |
| **cg12607957** | chr8 | 57703383 | 57703383 | 1 | + | ENSG00000169122.11 | FAM110B | 291125 | 3,66995577576078E-06 | 0,004127764 | G > AG | 0,078116777 |
| **cg26035347** | chr12 | 93274440 | 93274440 | 1 | - | ENSG00000177889.10 | UBE2N | 167508 | 3,68214628721616E-06 | 0,004129969 | G > AG | 0,08824537 |
| **cg07341914** | chr4 | 95051624 | 95051624 | 1 | - | ENSG00000182168.15 | UNC5C | 497583 | 3,6975937383615E-06 | 0,004135168 | G > AG | 0,081498377 |
| **cg11189709** | chr9 | 124887680 | 124887680 | 1 | + | ENSG00000136950.13 | ARPC5L | 25551 | 3,72327694915297E-06 | 0,004151751 | G > AG | 0,083234256 |
| **cg24619462** | chr2 | 16228350 | 16228350 | 1 | + | ENSG00000134323.12 | MYCN | 287801 | 3,71832461921895E-06 | 0,004151751 | G > AG | 0,064973547 |
| **cg17080727** | chr5 | 127740110 | 127740110 | 1 | - | ENSG00000164241.14 | C5orf63 | 666603 | 3,75222953037856E-06 | 0,004171873 | G > AG | 0,083093567 |
| **cg08747277** | chr6 | 95600250 | 95600250 | 1 | - | ENSG00000112218.9 | GPR63 | 1237228 | 3,74692352461596E-06 | 0,004171873 | G > AG | 0,096128889 |
| **cg02617540** | chr2 | 29803460 | 29803460 | 1 | - | ENSG00000171094.18 | ALK | 118127 | 3,82020522958243E-06 | 0,004222899 | G > AG | 0,060832227 |
| **cg05593906** | chr3 | 139710168 | 139710168 | 1 | + | ENSG00000158258.16 | CLSTN2 | 225016 | 3,81907550302313E-06 | 0,004222899 | G > AG | 0,109139621 |
| **cg06014707** | chr8 | 74755383 | 74755383 | 1 | - | ENSG00000104369.5 | JPH1 | 433842 | 3,81174933310775E-06 | 0,004222899 | G > AG | 0,111628203 |
| **cg15646593** | chr4 | 120763198 | 120763198 | 1 | - | ENSG00000138738.11 | PRDM5 | 159673 | 3,81476414093664E-06 | 0,004222899 | G > AG | 0,067100319 |
| **cg25083402** | chr14 | 103926058 | 103926058 | 1 | - | ENSG00000156411.10 | ATP5MJ | 2212 | 3,83721551571065E-06 | 0,004229479 | G > AG | 0,113588844 |
| **cg11439662** | chr8 | 110060055 | 110060055 | 1 | - | ENSG00000164794.9 | KCNV1 | 84283 | 3,85519187181166E-06 | 0,004237082 | G > AG | 0,063796144 |
| **cg06456216** | chr4 | 61519437 | 61519437 | 1 | - | ENSG00000205678.8 | TECRL | 2890032 | 3,84968260308143E-06 | 0,004237082 | G > AG | 0,082029709 |
| **cg08004360** | chr5 | 56530210 | 56530210 | 1 | - | ENSG00000164512.18 | ANKRD55 | 296879 | 3,87634404817527E-06 | 0,004242045 | G > AG | 0,147240347 |
| **cg13597025** | chr6 | 165674744 | 165674744 | 1 | - | ENSG00000112541.18 | PDE10A | 313335 | 3,87408398705872E-06 | 0,004242045 | G > AG | 0,102129974 |
| **cg12786481** | chr9 | 19028869 | 19028869 | 1 | + | ENSG00000155876.6 | RRAGA | 20557 | 3,87480044120206E-06 | 0,004242045 | G > AG | 0,124642515 |
| **cg22006654** | chr2 | 146635031 | 146635031 | 1 | - | ENSG00000115947.14 | ORC4 | 1386574 | 3,89125603200183E-06 | 0,00425228 | G > AG | 0,110702781 |
| **cg24938632** | chr17 | 48752763 | 48752763 | 1 | - | ENSG00000159184.8 | HOXB13 | 24012 | 3,91062091822461E-06 | 0,004261267 | G > AG | 0,069363051 |
| **cg09289651** | chr6 | 9200163 | 9200163 | 1 | - | ENSG00000124786.12 | SLC35B3 | 764601 | 3,9059655596994E-06 | 0,004261267 | G > AG | 0,094553783 |
| **cg19580776** | chr14 | 62179872 | 62179872 | 1 | + | ENSG00000139973.17 | SYT16 | 367899 | 3,92600893014235E-06 | 0,004271949 | G > AG | 0,055017671 |
| **cg06702990** | chr11 | 92829561 | 92829561 | 1 | + | ENSG00000134640.3 | MTNR1B | 140089 | 3,94018337209911E-06 | 0,004281282 | G > AG | 0,068557255 |

| | | | | | | | | | | | | |
|---|---|---|---|---|---|---|---|---|---|---|---|---|
| **cg20771998** | chr17 | 70661422 | 70661422 | 1 | + | ENSG00000123700.5 | KCNJ2 | 492750 | 3,9761255932372E-06 | 0,004302004 | G > AG | 0,125734913 |
| **cg03801287** | chr15 | 47727034 | 47727034 | 1 | + | ENSG00000188467.11 | SLC24A5 | 393955 | 3,97358070097625E-06 | 0,004302004 | G > AG | 0,099996919 |
| **cg27175191** | chr3 | 109119541 | 109119541 | 1 | + | ENSG00000241224.8 | C3orf85 | 1290 | 4,00769820928141E-06 | 0,004324378 | G > AG | 0,117523537 |
| **cg16112285** | chr20 | 23133233 | 23133233 | 1 | + | ENSG00000132671.6 | SSTR4 | 97922 | 4,00811141042205E-06 | 0,004324378 | G > AG | 0,078006328 |
| **cg00118369** | chr17 | 9974245 | 9974245 | 1 | - | ENSG00000109047.8 | RCVRN | 68973 | 4,08861013355686E-06 | 0,00439882 | G > AG | 0,111956195 |
| **cg16468389** | chr17 | 48754380 | 48754380 | 1 | + | ENSG00000229637.4 | PRAC2 | 31196 | 4,12092456648536E-06 | 0,004414958 | G > AG | 0,109511862 |
| **cg06975043** | chr9 | 119284747 | 119284747 | 1 | + | ENSG00000214654.8 | B3GALT9 | 1507655 | 4,15809002661494E-06 | 0,004438069 | G > AG | 0,127529916 |
| **cg08317144** | chr9 | 14563981 | 14563981 | 1 | + | ENSG00000164975.15 | SNAPC3 | 858722 | 4,15990217297196E-06 | 0,004438069 | G > AG | 0,06870802 |
| **cg03967240** | chr7 | 28390240 | 28390240 | 1 | + | ENSG00000146592.17 | CREB5 | 90920 | 4,17214156032783E-06 | 0,004444928 | G > AG | 0,156978473 |
| **cg10876051** | chr17 | 15464313 | 15464313 | 1 | + | ENSG00000187607.16 | ZNF286A | 235263 | 4,1826128449552E-06 | 0,004449886 | G > AG | 0,076395411 |
| **cg19349346** | chr14 | 32007288 | 32007288 | 1 | - | ENSG00000214943.5 | GPR33 | 519248 | 4,20436672946148E-06 | 0,004466817 | G > AG | 0,084092215 |
| **cg02612270** | chr16 | 965103 | 965103 | 1 | - | ENSG00000103227.19 | LMF1 | 16216 | 4,21578614182216E-06 | 0,004472737 | G > AG | 0,098858392 |
| **cg21222888** | chr13 | 93840802 | 93840802 | 1 | - | ENSG00000080166.16 | DCT | 638881 | 4,24236426032981E-06 | 0,004494702 | G > AG | 0,128281409 |
| **cg13551535** | chr4 | 79050308 | 79050308 | 1 | + | ENSG00000138756.19 | BMP2K | 273967 | 4,25675677868487E-06 | 0,004499124 | G > AG | 0,151204595 |
| **cg12014037** | chr7 | 157999415 | 157999415 | 1 | - | ENSG00000155093.19 | PTPRN2 | 588409 | 4,27271015766841E-06 | 0,004499124 | G > AG | 0,086320828 |
| **cg10868668** | chr4 | 38605321 | 38605321 | 1 | - | ENSG00000174123.11 | TLR10 | 177670 | 4,26363295269257E-06 | 0,004499124 | G > AG | 0,097966927 |
| **cg12561408** | chr6 | 153129199 | 153129199 | 1 | + | ENSG00000146469.13 | VIP | 378403 | 4,27894235561993E-06 | 0,004499124 | G > AG | 0,108416423 |
| **cg14398228** | chr14 | 96863583 | 96863583 | 1 | + | ENSG00000100749.9 | VRK1 | 66280 | 4,29359132234214E-06 | 0,004499124 | G > AG | 0,091420926 |
| **cg03062642** | chr10 | 120885078 | 120885078 | 1 | + | ENSG00000120008.16 | WDR11 | 33774 | 4,29160463954419E-06 | 0,004499124 | G > AG | 0,061445809 |
| **cg11504954** | chr17 | 15406479 | 15406479 | 1 | + | ENSG00000187607.16 | ZNF286A | 293097 | 4,26737106903867E-06 | 0,004499124 | G > AG | 0,086841998 |
| **cg17453024** | chr6 | 47174629 | 47174629 | 1 | - | ENSG00000153292.16 | ADGRF1 | 132278 | 4,30374586052624E-06 | 0,004503595 | G > AG | 0,107279367 |
| **cg13121144** | chr15 | 32926455 | 32926455 | 1 | + | ENSG00000166923.12 | GREM1 | 208452 | 4,31117668675098E-06 | 0,004505208 | G > AG | 0,083576145 |
| **cg20344367** | chr5 | 406778 | 406778 | 1 | - | ENSG00000066230.12 | SLC9A3 | 117672 | 4,32936977449866E-06 | 0,004518048 | G > AG | 0,069039129 |
| **cg20922371** | chr2 | 46330571 | 46330571 | 1 | + | ENSG00000116016.14 | EPAS1 | 36905 | 4,34640540096335E-06 | 0,004528619 | G > AG | 0,119114907 |
| **cg20024732** | chr4 | 138104410 | 138104410 | 1 | + | ENSG00000151014.6 | NOCT | 911370 | 4,3513400473089E-06 | 0,004528619 | G > AG | 0,105258734 |
| **cg19653417** | chr12 | 132170379 | 132170379 | 1 | - | ENSG00000185163.10 | DDX51 | 26059 | 4,36471238229279E-06 | 0,004536364 | G > AG | 0,118667617 |
| **cg04180823** | chr6 | 167238540 | 167238540 | 1 | + | ENSG00000112494.10 | UNC93A | 32628 | 4,44908149873403E-06 | 0,00461152 | G > AG | 0,072361996 |
| **cg10620434** | chr1 | 5526297 | 5526297 | 1 | + | ENSG00000069424.15 | KCNAB2 | 464629 | 4,45957600558422E-06 | 0,004613816 | G > AG | 0,06048402 |
| **cg19057390** | chr8 | 69891758 | 69891758 | 1 | + | ENSG00000137573.14 | SULF1 | 425135 | 4,46336019354206E-06 | 0,004613816 | G > AG | 0,098853053 |
| **cg10251446** | chr3 | 187907036 | 187907036 | 1 | - | ENSG00000113916.18 | BCL6 | 161310 | 4,49426844585243E-06 | 0,004639497 | G > AG | 0,073196124 |
| **cg16964439** | chr9 | 84672092 | 84672092 | 1 | + | ENSG00000148053.17 | NTRK2 | 3542 | 4,5332529606483E-06 | 0,004673434 | G > AG | 0,076062858 |

| | | | | | | | | | | | | |
|---|---|---|---|---|---|---|---|---|---|---|---|---|
| **cg25180625** | chr20 | 34180646 | 34180646 | 1 | + | ENSG00000101440.10 | ASIP | 13922 | 4,54122509408177E-06 | 0,004675352 | G > AG | 0,086407589 |
| **cg11294721** | chrX | 13622582 | 13622582 | 1 | + | ENSG00000176896.8 | TCEANC | 30606 | 4,55806761884405E-06 | 0,004686385 | G > AG | 0,115532675 |
| **cg26992381** | chr3 | 71429001 | 71429001 | 1 | + | ENSG00000170837.3 | GPR27 | 324853 | 4,56693311959503E-06 | 0,004689197 | G > AG | 0,138556523 |
| **cg25850484** | chr1 | 17232853 | 17232853 | 1 | + | ENSG00000142619.4 | PADI3 | 16244 | 4,58320470537431E-06 | 0,004693305 | G > AG | 0,070978784 |
| **cg06251435** | chr18 | 72013692 | 72013692 | 1 | + | ENSG00000170677.6 | SOCS6 | 1724648 | 4,59357134091462E-06 | 0,004697632 | G > AG | 0,107629751 |
| **cg18086445** | chr3 | 74080091 | 74080091 | 1 | - | ENSG00000113805.8 | CNTN3 | 441050 | 4,61268205501616E-06 | 0,004710877 | G > AG | 0,062196541 |
| **cg04633888** | chr17 | 79919271 | 79919271 | 1 | - | ENSG00000141582.15 | CBX4 | 79830 | 4,63823850913807E-06 | 0,004730662 | G > AG | 0,113812757 |
| **cg26908340** | chr8 | 58273262 | 58273262 | 1 | + | ENSG00000215114.10 | UBXN2B | 138096 | 4,65587373844353E-06 | 0,004742325 | G > AG | 0,119021729 |
| **cg18381465** | chr8 | 123531989 | 123531989 | 1 | + | ENSG00000156795.8 | NTAQ1 | 115264 | 4,67334164044467E-06 | 0,004753788 | G > AG | 0,067518354 |
| **cg12863955** | chr1 | 14254622 | 14254622 | 1 | - | ENSG00000162494.6 | LRRC38 | 740618 | 4,68894425069347E-06 | 0,004757007 | G > AG | 0,116930518 |
| **cg18370772** | chr8 | 26077775 | 26077775 | 1 | + | ENSG00000221914.11 | PPP2R2A | 213732 | 4,72446204701109E-06 | 0,004774046 | G > AG | 0,149685128 |
| **cg04803335** | chr3 | 42923670 | 42923670 | 1 | + | ENSG00000182983.15 | ZNF662 | 17529 | 4,75097050938807E-06 | 0,004788182 | G > AG | 0,10127557 |
| **cg23375121** | chr18 | 26941719 | 26941719 | 1 | + | ENSG00000141384.13 | TAF4B | 715275 | 4,78598477672494E-06 | 0,004799644 | G > AG | 0,136554397 |
| **cg19353326** | chr13 | 112160769 | 112160769 | 1 | - | ENSG00000126216.15 | TUBGCP3 | 427437 | 4,78744086705376E-06 | 0,004799644 | G > AG | 0,058318176 |
| **cg26603183** | chr4 | 95051593 | 95051593 | 1 | + | ENSG00000138696.11 | BMPR1B | 293639 | 4,80285674233707E-06 | 0,00480251 | G > AG | 0,099569164 |
| **cg06097438** | chr16 | 55641571 | 55641571 | 1 | - | ENSG00000198848.13 | CES1 | 191767 | 4,82949300616614E-06 | 0,00482284 | G > AG | 0,103046395 |
| **cg02455045** | chr2 | 13022397 | 13022397 | 1 | + | ENSG00000071575.12 | TRIB2 | 305488 | 4,86832647656166E-06 | 0,004855282 | G > AG | 0,116014267 |
| **cg15721474** | chr5 | 173195860 | 173195860 | 1 | + | ENSG00000113734.18 | BNIP1 | 51419 | 4,88893968259081E-06 | 0,004856843 | G > AG | 0,072644105 |
| **cg02297168** | chr13 | 30192775 | 30192775 | 1 | - | ENSG00000102781.14 | KATNAL1 | 114777 | 4,88867698238411E-06 | 0,004856843 | G > AG | 0,071108333 |
| **cg11923916** | chr17 | 9646944 | 9646944 | 1 | + | ENSG00000154914.17 | USP43 | 2247 | 4,88565205356697E-06 | 0,004856843 | G > AG | 0,147635126 |
| **cg18167201** | chr20 | 48660162 | 48660162 | 1 | + | ENSG00000124198.10 | ARFGEF2 | 261548 | 4,91364713116378E-06 | 0,004875057 | G > AG | 0,093946004 |
| **cg02612927** | chr20 | 31473081 | 31473081 | 1 | - | ENSG00000180383.4 | DEFB124 | 3677 | 4,96694331191459E-06 | 0,004921551 | G > AG | 0,065097484 |
| **cg18558488** | chr13 | 28854743 | 28854743 | 1 | - | ENSG00000139508.15 | SLC46A3 | 135772 | 4,98008216528763E-06 | 0,004928186 | G > AG | 0,104870169 |
| **cg15264281** | chr10 | 77305594 | 77305594 | 1 | - | ENSG00000156113.24 | KCNMA1 | 332776 | 4,99303423239253E-06 | 0,004928252 | G > AG | 0,098897166 |
| **cg04786038** | chr18 | 2147654 | 2147654 | 1 | - | ENSG00000101574.15 | METTL4 | 423856 | 4,98902279559772E-06 | 0,004928252 | G > AG | 0,124714163 |
| **cg03328571** | chr2 | 169358293 | 169358293 | 1 | + | ENSG00000163093.12 | BBS5 | 121186 | 5,02613189569313E-06 | 0,004954528 | G > AG | 0,067409628 |
| **cg20200779** | chr15 | 31296882 | 31296882 | 1 | + | ENSG00000169926.11 | KLF13 | 29952 | 5,05185340580927E-06 | 0,004973474 | G > AG | 0,108664208 |
| **cg11454747** | chr1 | 15049347 | 15049347 | 1 | - | ENSG00000132906.18 | CASP9 | 477188 | 5,07883088388769E-06 | 0,004987196 | G > AG | 0,060615484 |
| **cg03939372** | chr2 | 191552400 | 191552400 | 1 | - | ENSG00000168497.5 | CAVIN2 | 294689 | 5,07342746622729E-06 | 0,004987196 | G > AG | 0,087294615 |
| **cg13205617** | chr2 | 2173759 | 2173759 | 1 | - | ENSG00000186487.20 | MYT1L | 157906 | 5,08648120379217E-06 | 0,004988304 | G > AG | 0,093929862 |
| **cg24776259** | chr19 | 54008750 | 54008750 | 1 | + | ENSG00000130433.8 | CACNG6 | 17114 | 5,11727446239861E-06 | 0,005011643 | G > AG | 0,102219071 |

| | | | | | | | | | | | | |
|---|---|---|---|---|---|---|---|---|---|---|---|---|
| **cg09608958** | chr3 | 18988258 | 18988258 | 1 | + | ENSG00000183960.9 | KCNH8 | 160251 | 5,12588779207602E-06 | 0,005011643 | G > AG | 0,097527037 |
| **cg01255459** | chr1 | 117237935 | 117237935 | 1 | + | ENSG00000198162.12 | MAN1A2 | 129513 | 5,1629373821888E-06 | 0,005037452 | G > AG | 0,115700982 |
| **cg04957384** | chr16 | 76778524 | 76778524 | 1 | + | ENSG00000103111.15 | MON1B | 412310 | 5,24516880243419E-06 | 0,005098176 | G > AG | 0,083213111 |
| **cg17452354** | chr2 | 192585688 | 192585688 | 1 | - | ENSG00000144339.12 | TMEFF2 | 390754 | 5,23995700585629E-06 | 0,005098176 | G > AG | 0,098398484 |
| **cg21449559** | chr20 | 52801665 | 52801665 | 1 | - | ENSG00000020256.20 | ZFP64 | 597356 | 5,23560923673141E-06 | 0,005098176 | G > AG | 0,079671576 |
| **cg22797339** | chr17 | 62955930 | 62955930 | 1 | - | ENSG00000173838.12 | MARCHF10 | 147585 | 5,26250364236759E-06 | 0,005104773 | G > AG | 0,116053541 |
| **cg13090007** | chr8 | 141900232 | 141900232 | 1 | + | ENSG00000226490.2 | AC138647.1 | 385595 | 5,28505778900553E-06 | 0,00511744 | G > AG | 0,062070758 |
| **cg02523906** | chr2 | 21758143 | 21758143 | 1 | - | ENSG00000084674.15 | APOB | 714069 | 5,31445353315352E-06 | 0,005133181 | G > AG | 0,066518749 |
| **cg16232933** | chr11 | 40167101 | 40167101 | 1 | - | ENSG00000148948.8 | LRRC4C | 1292673 | 5,3281564114163E-06 | 0,005133181 | G > AG | 0,1030655 |
| **cg16550158** | chr11 | 69267595 | 69267595 | 1 | - | ENSG00000172935.9 | MRGPRF | 254212 | 5,32374007702928E-06 | 0,005133181 | G > AG | 0,085247563 |
| **cg02187357** | chr6 | 37256440 | 37256440 | 1 | - | ENSG00000286105.1 | RP3-355M6.3 | 1198 | 5,31903201990211E-06 | 0,005133181 | G > AG | 0,076763068 |
| **cg21341391** | chr17 | 53755132 | 53755132 | 1 | - | ENSG00000166260.13 | COX11 | 1213572 | 5,37234198442931E-06 | 0,005156157 | G > AG | 0,086824061 |
| **cg05305278** | chr15 | 45510827 | 45510827 | 1 | - | ENSG00000104154.7 | SLC30A4 | 11929 | 5,37896766176067E-06 | 0,005156157 | G > AG | 0,090517641 |
| **cg00032637** | chr2 | 227295993 | 227295993 | 1 | - | ENSG00000168955.4 | TM4SF20 | 86003 | 5,39357462329235E-06 | 0,005161327 | G > AG | 0,087530327 |
| **cg22572902** | chr4 | 95051639 | 95051639 | 1 | - | ENSG00000182168.15 | UNC5C | 497568 | 5,40460332009135E-06 | 0,005161327 | G > AG | 0,069324916 |
| **cg17321346** | chr2 | 68727043 | 68727043 | 1 | - | ENSG00000163217.2 | BMP10 | 144355 | 5,43582899426085E-06 | 0,005171778 | G > AG | 0,095804896 |
| **cg05061672** | chr5 | 96247737 | 96247737 | 1 | + | ENSG00000153113.24 | CAST | 277529 | 5,42440556427504E-06 | 0,005171778 | G > AG | 0,115329096 |
| **cg25377605** | chr8 | 10656207 | 10656207 | 1 | - | ENSG00000183638.6 | RP1L1 | 55981 | 5,43334354041423E-06 | 0,005171778 | G > AG | 0,085909429 |
| **cg02081722** | chr7 | 100463988 | 100463988 | 1 | + | ENSG00000166924.9 | NYAP1 | 19938 | 5,46420097008295E-06 | 0,005192313 | G > AG | 0,062824739 |
| **cg02992887** | chr1 | 184399704 | 184399704 | 1 | + | ENSG00000116667.15 | C1orf21 | 12676 | 5,4791569610659E-06 | 0,00519948 | G > AG | 0,087996902 |
| **cg26834993** | chr1 | 119196584 | 119196584 | 1 | - | ENSG00000116874.12 | WARS2 | 55929 | 5,48533768025267E-06 | 0,00519948 | G > AG | 0,082389132 |
| **cg17647537** | chr11 | 113908235 | 113908235 | 1 | - | ENSG00000048028.11 | USP28 | 32664 | 5,54467127116094E-06 | 0,005249217 | G > AG | 0,07164339 |
| **cg11354777** | chr2 | 67562050 | 67562050 | 1 | - | ENSG00000197223.12 | C1D | 548899 | 5,59541355021772E-06 | 0,005290708 | G > AG | 0,083590644 |
| **cg09229183** | chr2 | 191247601 | 191247601 | 1 | + | ENSG00000128641.19 | MYO1B | 2417 | 5,62024573406743E-06 | 0,005303036 | G > AG | 0,099588408 |
| **cg14377849** | chr1 | 56073946 | 56073946 | 1 | + | ENSG00000162409.11 | PRKAA2 | 571367 | 5,62231684042483E-06 | 0,005303036 | G > AG | 0,071675732 |
| **cg21023534** | chr12 | 28938814 | 28938814 | 1 | + | ENSG00000064763.11 | FAR2 | 210288 | 5,63545228262367E-06 | 0,005303402 | G > AG | 0,083613153 |
| **cg06607919** | chr2 | 80322672 | 80322672 | 1 | - | ENSG00000162951.11 | LRRTM1 | 17919 | 5,63657129097433E-06 | 0,005303402 | G > AG | 0,082593929 |
| **cg01869955** | chr8 | 1061835 | 1061835 | 1 | + | ENSG00000198010.13 | DLGAP2 | 324208 | 5,6639997241295E-06 | 0,005322662 | G > AG | 0,082096567 |
| **cg19664449** | chr14 | 71236697 | 71236697 | 1 | - | ENSG00000006432.16 | MAP3K9 | 427162 | 5,6847446159826E-06 | 0,005335602 | G > AG | 0,097314643 |
| **cg06624056** | chr7 | 28279880 | 28279880 | 1 | - | ENSG00000153814.13 | JAZF1 | 99084 | 5,69830360109816E-06 | 0,005341774 | G > AG | 0,07791112 |
| **cg10699939** | chr8 | 73085973 | 73085973 | 1 | + | ENSG00000147601.15 | TERF1 | 77118 | 5,71221159545726E-06 | 0,005348258 | G > AG | 0,077183888 |

| | | | | | | | | | | | | |
|---|---|---|---|---|---|---|---|---|---|---|---|---|
| **cg10157574** | chr8 | 32720433 | 32720433 | 1 | - | ENSG00000172728.16 | FUT10 | 752714 | 5,74426387946355E-06 | 0,005371693 | G > AG | 0,130110428 |
| **cg04482712** | chr17 | 74761154 | 74761154 | 1 | + | ENSG00000109062.12 | SLC9A3R1 | 12527 | 5,78296500530526E-06 | 0,005401281 | G > AG | 0,084361862 |
| **cg07182708** | chr2 | 120392248 | 120392248 | 1 | + | ENSG00000163083.6 | INHBB | 46113 | 5,79740003079299E-06 | 0,00540816 | G > AG | 0,098090552 |
| **cg17824906** | chr12 | 1590845 | 1590845 | 1 | + | ENSG00000111186.13 | WNT5B | 60955 | 5,81047364601184E-06 | 0,005413754 | G > AG | 0,096107348 |
| **cg00084169** | chr1 | 3335456 | 3335456 | 1 | - | ENSG00000162591.16 | MEGF6 | 276053 | 5,86095810203355E-06 | 0,00544091 | G > AG | 0,094561806 |
| **cg03627237** | chr2 | 147487840 | 147487840 | 1 | - | ENSG00000115947.14 | ORC4 | 533765 | 5,86978782957367E-06 | 0,005442501 | G > AG | 0,11356843 |
| **cg10818074** | chr7 | 28587327 | 28587327 | 1 | - | ENSG00000255690.3 | TRIL | 371004 | 5,87924709732513E-06 | 0,005444673 | G > AG | 0,105070155 |
| **cg12766091** | chr8 | 79903894 | 79903894 | 1 | - | ENSG00000147586.10 | MRPS28 | 126396 | 5,91239505739854E-06 | 0,005462145 | G > AG | 0,085369888 |
| **cg16020380** | chr11 | 11347508 | 11347508 | 1 | + | ENSG00000170242.19 | USP47 | 493914 | 5,90608132708718E-06 | 0,005462145 | G > AG | 0,110993069 |
| **cg19273865** | chr14 | 22487188 | 22487188 | 1 | - | ENSG00000129562.11 | DAD1 | 102037 | 5,92480498670634E-06 | 0,005467007 | G > AG | 0,103258815 |
| **cg09825321** | chr3 | 139944859 | 139944859 | 1 | + | ENSG00000158258.16 | CLSTN2 | 9675 | 5,97733644664642E-06 | 0,005507065 | G > AG | 0,082965635 |
| **cg14234912** | chr11 | 103806775 | 103806775 | 1 | + | ENSG00000170967.5 | DDI1 | 229864 | 5,98261617739007E-06 | 0,005507065 | G > AG | 0,08595199 |
| **cg01923516** | chr7 | 3949535 | 3949535 | 1 | - | ENSG00000198286.10 | CARD11 | 905667 | 6,00232511273213E-06 | 0,005511942 | G > AG | 0,078622782 |
| **cg21765730** | chr6 | 155456041 | 155456041 | 1 | + | ENSG00000171217.5 | CLDN20 | 192029 | 5,99941638822451E-06 | 0,005511942 | G > AG | 0,107462259 |
| **cg25318933** | chr20 | 53613502 | 53613502 | 1 | + | ENSG00000101132.10 | PFDN4 | 594584 | 6,05281197852691E-06 | 0,005544991 | G > AG | 0,067721486 |
| **cg27052685** | chr2 | 104052707 | 104052707 | 1 | - | ENSG00000135966.13 | TGFBRAP1 | 1277029 | 6,10257452198433E-06 | 0,005583891 | G > AG | 0,058248265 |
| **cg26872907** | chr6 | 161375822 | 161375822 | 1 | - | ENSG00000026652.15 | AGPAT4 | 101760 | 6,1864113615843E-06 | 0,005640361 | G > AG | 0,124341243 |
| **cg20578405** | chr4 | 158521433 | 158521433 | 1 | + | ENSG00000171503.13 | ETFDH | 150534 | 6,20699390971371E-06 | 0,005648896 | G > AG | 0,127749906 |
| **cg10991752** | chr6 | 155524180 | 155524180 | 1 | + | ENSG00000171217.5 | CLDN20 | 260168 | 6,2502542343168E-06 | 0,00567153 | G > AG | 0,120769713 |
| **cg23523548** | chr11 | 104609692 | 104609692 | 1 | + | ENSG00000170967.5 | DDI1 | 573053 | 6,24301018604416E-06 | 0,00567153 | G > AG | 0,078405809 |
| **cg03509898** | chr14 | 103724413 | 103724413 | 1 | + | ENSG00000100711.14 | ZFYVE21 | 8684 | 6,26624030813858E-06 | 0,005679299 | G > AG | 0,125223666 |
| **cg24754626** | chr2 | 16727499 | 16727499 | 1 | - | ENSG00000197872.11 | CYRIA | 61167 | 6,35434611001285E-06 | 0,005731986 | G > AG | 0,06119997 |
| **cg00600584** | chr3 | 189630025 | 189630025 | 1 | + | ENSG00000073282.14 | TP63 | 1363 | 6,34795303535976E-06 | 0,005731986 | G > AG | 0,108049899 |
| **cg07407218** | chr1 | 50330628 | 50330628 | 1 | - | ENSG00000142700.12 | DMRTA2 | 92816 | 6,40066078467609E-06 | 0,005733383 | G > AG | 0,094649126 |
| **cg06516640** | chr12 | 26722017 | 26722017 | 1 | + | ENSG00000111790.14 | FGFR1OP2 | 216452 | 6,38859140601248E-06 | 0,005733383 | G > AG | 0,109550939 |
| **cg07447910** | chr9 | 69205120 | 69205120 | 1 | - | ENSG00000165059.8 | PRKACG | 191006 | 6,39111768791513E-06 | 0,005733383 | G > AG | 0,06275544 |
| **cg07853663** | chr17 | 72027230 | 72027230 | 1 | + | ENSG00000125398.8 | SOX9 | 93789 | 6,4008661122866E-06 | 0,005733383 | G > AG | 0,098241185 |
| **cg25750291** | chr21 | 28633027 | 28633027 | 1 | + | ENSG00000156256.15 | USP16 | 391601 | 6,36356351219571E-06 | 0,005733383 | G > AG | 0,076700493 |
| **cg12919390** | chr8 | 101157869 | 101157869 | 1 | - | ENSG00000120963.12 | ZNF706 | 48325 | 6,4604164535024E-06 | 0,005779956 | G > AG | 0,163867205 |
| **cg08340618** | chr9 | 135639754 | 135639754 | 1 | + | ENSG00000148386.9 | LCN9 | 23567 | 6,48166929547906E-06 | 0,005782057 | G > AG | 0,106776842 |
| **cg11971852** | chr2 | 32453778 | 32453778 | 1 | - | ENSG00000091106.19 | NLRC4 | 188045 | 6,5005587565846E-06 | 0,005782057 | G > AG | 0,076197432 |

| | | | | | | | | | | | | |
|---|---|---|---|---|---|---|---|---|---|---|---|---|
| **cg15620850** | chr6 | 25831936 | 25831936 | 1 | - | ENSG00000124568.12 | SLC17A1 | 117 | 6,50037140540687E-06 | 0,005782057 | G > AG | 0,119502851 |
| **cg20286290** | chr15 | 40910313 | 40910313 | 1 | + | ENSG00000104142.11 | VPS18 | 15864 | 6,48805305415386E-06 | 0,005782057 | G > AG | 0,109877556 |
| **cg13849759** | chr9 | 13860786 | 13860786 | 1 | + | ENSG00000153714.6 | LURAP1L | 1085767 | 6,51744966987732E-06 | 0,005790348 | G > AG | 0,060133768 |
| **cg16408679** | chr2 | 54290245 | 54290245 | 1 | - | ENSG00000178021.11 | TSPYL6 | 34015 | 6,53408726463134E-06 | 0,005798395 | G > AG | 0,080392998 |
| **cg20749999** | chr15 | 86399199 | 86399199 | 1 | + | ENSG00000273540.5 | AGBL1 | 319329 | 6,61956141012244E-06 | 0,005808424 | G > AG | 0,057755754 |
| **cg03557384** | chr3 | 182570951 | 182570951 | 1 | + | ENSG00000058063.16 | ATP11B | 222551 | 6,62781328600312E-06 | 0,005808424 | G > AG | 0,09094514 |
| **cg24206431** | chr17 | 80187668 | 80187668 | 1 | + | ENSG00000141527.18 | CARD14 | 17677 | 6,59333990343362E-06 | 0,005808424 | G > AG | 0,088881479 |
| **cg00245560** | chr1 | 12368904 | 12368904 | 1 | - | ENSG00000162496.9 | DHRS3 | 249307 | 6,60132243389782E-06 | 0,005808424 | G > AG | 0,093913423 |
| **cg21574556** | chr5 | 109015231 | 109015231 | 1 | + | ENSG00000151422.13 | FER | 267391 | 6,63438817969648E-06 | 0,005808424 | G > AG | 0,079453119 |
| **cg08519239** | chr7 | 126937380 | 126937380 | 1 | - | ENSG00000179603.18 | GRM8 | 315714 | 6,5791841706924E-06 | 0,005808424 | G > AG | 0,112395203 |
| **cg17529125** | chr5 | 154576584 | 154576584 | 1 | - | ENSG00000113196.3 | HAND1 | 98356 | 6,59930839423608E-06 | 0,005808424 | G > AG | 0,088890274 |
| **cg15303300** | chr1 | 163875719 | 163875719 | 1 | + | ENSG00000143228.13 | NUF2 | 609144 | 6,61071299795123E-06 | 0,005808424 | G > AG | 0,117758091 |
| **cg11067226** | chr13 | 24529857 | 24529857 | 1 | - | ENSG00000102699.6 | PARP4 | 17078 | 6,6309273256293E-06 | 0,005808424 | G > AG | 0,058030807 |
| **cg10181156** | chr1 | 171435291 | 171435291 | 1 | + | ENSG00000117523.17 | PRRC2C | 50238 | 6,63650784356479E-06 | 0,005808424 | G > AG | 0,074758734 |
| **cg03925904** | chr4 | 186271585 | 186271585 | 1 | - | ENSG00000272297.2 | RP11-215A19.2 | 283744 | 6,5667823123807E-06 | 0,005808424 | G > AG | 0,086967222 |
| **cg07697770** | chr5 | 136054016 | 136054016 | 1 | + | ENSG00000120708.17 | TGFBI | 25029 | 6,63349594702287E-06 | 0,005808424 | G > AG | 0,090894788 |
| **cg04777769** | chr6 | 170234562 | 170234562 | 1 | + | ENSG00000112584.14 | FAM120B | 56140 | 6,67704810569753E-06 | 0,005837227 | G > AG | 0,050566838 |
| **cg14261176** | chr9 | 84239654 | 84239654 | 1 | + | ENSG00000178966.17 | RMI1 | 258857 | 6,69753229807E-06 | 0,00584845 | G > AG | 0,098562186 |
| **cg03455736** | chr22 | 49488848 | 49488848 | 1 | - | ENSG00000100425.18 | BRD1 | 338665 | 6,72938561219967E-06 | 0,005869565 | G > AG | 0,117843953 |
| **cg22903834** | chr13 | 35957904 | 35957904 | 1 | + | ENSG00000133101.10 | CCNA1 | 473615 | 6,74688740639646E-06 | 0,005878128 | G > AG | 0,049728313 |
| **cg23343735** | chr18 | 23495788 | 23495788 | 1 | - | ENSG00000134490.14 | TMEM241 | 57826 | 6,75771933488064E-06 | 0,005880867 | G > AG | 0,073255003 |
| **cg24860782** | chr2 | 195213031 | 195213031 | 1 | - | ENSG00000118997.14 | DNAH7 | 855807 | 6,77490115252153E-06 | 0,00588912 | G > AG | 0,072028373 |
| **cg03032512** | chr17 | 79034336 | 79034336 | 1 | - | ENSG00000171302.17 | CANT1 | 24468 | 6,82422116966758E-06 | 0,005925258 | G > AG | 0,115319173 |
| **cg12391588** | chr8 | 32128212 | 32128212 | 1 | - | ENSG00000172733.12 | PURG | 1094496 | 6,83787523872266E-06 | 0,005930382 | G > AG | 0,099031913 |
| **cg12718582** | chr8 | 1364709 | 1364709 | 1 | + | ENSG00000182372.10 | CLN8 | 391068 | 6,84772401382698E-06 | 0,005932198 | G > AG | 0,113560856 |
| **cg01078582** | chr21 | 40840675 | 40840675 | 1 | - | ENSG00000171587.15 | DSCAM | 6484 | 6,86904079529402E-06 | 0,005937217 | G > AG | 0,08347406 |
| **cg02642212** | chr4 | 88467596 | 88467596 | 1 | + | ENSG00000138646.9 | HERC5 | 10478 | 6,86495202185652E-06 | 0,005937217 | G > AG | 0,073252644 |
| **cg20063095** | chr2 | 134219570 | 134219570 | 1 | - | ENSG00000152128.13 | TMEM163 | 499431 | 6,89179247255153E-06 | 0,005950159 | G > AG | 0,086051009 |
| **cg21094614** | chr7 | 16652227 | 16652227 | 1 | + | ENSG00000136261.15 | BZW2 | 6097 | 6,93595307336687E-06 | 0,005969389 | G > AG | 0,061938476 |
| **cg14513578** | chr2 | 138658723 | 138658723 | 1 | + | ENSG00000144228.9 | SPOPL | 156954 | 6,94527993948829E-06 | 0,005969389 | G > AG | 0,054072341 |
| **cg05533607** | chr3 | 132630599 | 132630599 | 1 | + | ENSG00000081307.15 | UBA5 | 23846 | 6,93997983455956E-06 | 0,005969389 | G > AG | 0,085966746 |

| | | | | | | | | | | | | |
|---|---|---|---|---|---|---|---|---|---|---|---|---|
| **cg25026693** | chr11 | 122733485 | 122733485 | 1 | + | ENSG00000154127.10 | UBASH3B | 77764 | 6,95502431833124E-06 | 0,005971055 | G > AG | 0,100751482 |
| **cg15117233** | chr20 | 53756437 | 53756437 | 1 | - | ENSG00000171940.14 | ZNF217 | 146529 | 6,96686743805139E-06 | 0,005974517 | G > AG | 0,072353113 |
| **cg09662930** | chr15 | 81752798 | 81752798 | 1 | - | ENSG00000183496.6 | MEX3B | 293322 | 6,97747952809067E-06 | 0,005976917 | G > AG | 0,098880556 |
| **cg05439767** | chr3 | 124037160 | 124037160 | 1 | - | ENSG00000065371.17 | ROPN1 | 44981 | 6,98614641471501E-06 | 0,005977647 | G > AG | 0,110813925 |
| **cg05114674** | chr1 | 245440381 | 245440381 | 1 | - | ENSG00000153187.20 | HNRNPU | 575820 | 7,00451557148452E-06 | 0,005979986 | G > AG | 0,058507157 |
| **cg02411244** | chr7 | 52147091 | 52147091 | 1 | + | ENSG00000221900.6 | POM121L12 | 888541 | 7,02543915042385E-06 | 0,005991163 | G > AG | 0,105010529 |
| **cg15315515** | chr6 | 45813084 | 45813084 | 1 | - | ENSG00000112782.19 | CLIC5 | 267265 | 7,04121931401439E-06 | 0,005991262 | G > AG | 0,08249213 |
| **cg22301097** | chr4 | 163702936 | 163702936 | 1 | - | ENSG00000151005.5 | TKTL2 | 229181 | 7,03592020722748E-06 | 0,005991262 | G > AG | 0,081106168 |
| **cg03826600** | chr2 | 174663269 | 174663269 | 1 | - | ENSG00000115935.18 | WIPF1 | 19648 | 7,05850226524598E-06 | 0,005999294 | G > AG | 0,071331377 |
| **cg13332142** | chr3 | 27533787 | 27533787 | 1 | - | ENSG00000033867.16 | SLC4A7 | 49366 | 7,09806887112514E-06 | 0,006026227 | G > AG | 0,117174923 |
| **cg20538093** | chr16 | 55956097 | 55956097 | 1 | + | ENSG00000087258.16 | GNAO1 | 235292 | 7,10947413715243E-06 | 0,006026468 | G > AG | 0,073927379 |
| **cg12521187** | chr15 | 77696315 | 77696315 | 1 | + | ENSG00000140382.15 | HMG20A | 275904 | 7,13774418896387E-06 | 0,006026468 | G > AG | 0,093602324 |
| **cg19535073** | chr7 | 41704008 | 41704008 | 1 | - | ENSG00000122641.11 | INHBA | 1827 | 7,13501239421703E-06 | 0,006026468 | G > AG | 0,110341309 |
| **cg07413747** | chr10 | 19932153 | 19932153 | 1 | + | ENSG00000120594.17 | PLXDC2 | 115915 | 7,13350448060083E-06 | 0,006026468 | G > AG | 0,097481144 |
| **cg25325918** | chr2 | 119655684 | 119655684 | 1 | + | ENSG00000144120.13 | TMEM177 | 23482 | 7,11565867370285E-06 | 0,006026468 | G > AG | 0,076280044 |
| **cg21086148** | chr7 | 71871313 | 71871313 | 1 | + | ENSG00000185274.12 | GALNT17 | 739170 | 7,16879175446693E-06 | 0,00603935 | G > AG | 0,134354761 |
| **cg19895325** | chr18 | 55908951 | 55908951 | 1 | + | ENSG00000091157.14 | WDR7 | 742391 | 7,16460992203724E-06 | 0,00603935 | G > AG | 0,076238283 |
| **cg17926729** | chr12 | 93887345 | 93887345 | 1 | + | ENSG00000169372.13 | CRADD | 209971 | 7,1969859246448E-06 | 0,00605592 | G > AG | 0,051362292 |
| **cg10153023** | chr6 | 138194419 | 138194419 | 1 | - | ENSG00000254440.3 | PBOV1 | 24073 | 7,20429372685586E-06 | 0,00605592 | G > AG | 0,129498577 |
| **cg07415610** | chr8 | 76431471 | 76431471 | 1 | - | ENSG00000164751.15 | PEX2 | 569574 | 7,26075474442621E-06 | 0,0060921 | G > AG | 0,129708122 |
| **cg09863968** | chr6 | 98274979 | 98274979 | 1 | + | ENSG00000184486.10 | POU3F2 | 559594 | 7,26326243546023E-06 | 0,0060921 | G > AG | 0,130584858 |
| **cg03678595** | chr14 | 77557512 | 77557512 | 1 | - | ENSG00000100593.18 | ISM2 | 58695 | 7,29031783661151E-06 | 0,006101412 | G > AG | 0,073810391 |
| **cg16050285** | chr11 | 14942948 | 14942948 | 1 | - | ENSG00000110680.13 | CALCA | 29404 | 7,31107562402653E-06 | 0,006112098 | G > AG | 0,068369702 |
| **cg12260808** | chr20 | 35193926 | 35193926 | 1 | - | ENSG00000088298.13 | EDEM2 | 46589 | 7,36035258253344E-06 | 0,006123901 | G > AG | 0,09565 |
| **cg14588317** | chr3 | 59263086 | 59263086 | 1 | + | ENSG00000168301.13 | KCTD6 | 770991 | 7,35632265233184E-06 | 0,006123901 | G > AG | 0,065866806 |
| **cg22955177** | chr6 | 161482697 | 161482697 | 1 | + | ENSG00000085511.20 | MAP3K4 | 490971 | 7,34511620852661E-06 | 0,006123901 | G > AG | 0,121793185 |
| **cg22070156** | chr13 | 101452640 | 101452640 | 1 | - | ENSG00000102452.18 | NALCN | 36131 | 7,36522263452451E-06 | 0,006123901 | G > AG | 0,065610837 |
| **cg18278907** | chr15 | 67792821 | 67792821 | 1 | - | ENSG00000129007.16 | CALML4 | 413259 | 7,39169542491778E-06 | 0,006139239 | G > AG | 0,106487195 |
| **cg06058752** | chr3 | 197216404 | 197216404 | 1 | - | ENSG00000075711.21 | DLG1 | 82927 | 7,46433038479177E-06 | 0,006187196 | G > AG | 0,123489736 |
| **cg21919921** | chr3 | 168870778 | 168870778 | 1 | + | ENSG00000085274.16 | MYNN | 902617 | 7,46561337195367E-06 | 0,006187196 | G > AG | 0,072416838 |
| **cg07919456** | chr11 | 106249859 | 106249859 | 1 | - | ENSG00000182359.15 | KBTBD3 | 172399 | 7,50151298794247E-06 | 0,006206775 | G > AG | 0,094380898 |

| | | | | | | | | | | | | |
|---|---|---|---|---|---|---|---|---|---|---|---|---|
| **cg11243859** | chr8 | 48577024 | 48577024 | 1 | + | ENSG00000168333.14 | PPDPFL | 477286 | 7,50546591952055E-06 | 0,006206775 | G > AG | 0,090838698 |
| **cg23812853** | chr2 | 99602747 | 99602747 | 1 | - | ENSG00000135945.10 | REV1 | 112711 | 7,51960397105485E-06 | 0,006211752 | G > AG | 0,062394562 |
| **cg17487559** | chr14 | 91732663 | 91732663 | 1 | + | ENSG00000165934.13 | CPSF2 | 389305 | 7,56097582419374E-06 | 0,00623919 | G > AG | 0,077070386 |
| **cg00397764** | chr10 | 114179958 | 114179958 | 1 | - | ENSG00000165813.20 | CCDC186 | 5725 | 7,58982724941949E-06 | 0,006255701 | G > AG | 0,088316173 |
| **cg09304047** | chr2 | 53520403 | 53520403 | 1 | + | ENSG00000143942.5 | CHAC2 | 247400 | 7,60114388258199E-06 | 0,006255701 | G > AG | 0,09832535 |
| **cg18144687** | chr12 | 115570201 | 115570201 | 1 | + | ENSG00000258102.5 | MAP1LC3B2 | 977903 | 7,605518537431E-06 | 0,006255701 | G > AG | 0,098749867 |
| **cg07355627** | chr4 | 169921325 | 169921325 | 1 | + | ENSG00000109572.14 | CLCN3 | 308693 | 7,6291603369965E-06 | 0,006261681 | G > AG | 0,100957719 |
| **cg14155196** | chr8 | 22868391 | 22868391 | 1 | + | ENSG00000008853.17 | RHOBTB2 | 119025 | 7,62616796507059E-06 | 0,006261681 | G > AG | 0,06060126 |
| **cg24766543** | chr6 | 54929757 | 54929757 | 1 | + | ENSG00000168143.9 | FAM83B | 82987 | 7,76720579634806E-06 | 0,006361332 | G > AG | 0,078829514 |
| **cg12658800** | chr5 | 73672812 | 73672812 | 1 | - | ENSG00000164331.10 | ANKRA2 | 107144 | 7,80200117836952E-06 | 0,006365228 | G > AG | 0,075562789 |
| **cg03655701** | chr1 | 24559173 | 24559173 | 1 | + | ENSG00000184454.7 | NCMAP | 3087 | 7,79794936938536E-06 | 0,006365228 | G > AG | 0,091478077 |
| **cg07483116** | chr4 | 102042708 | 102042708 | 1 | + | ENSG00000109320.13 | NFKB1 | 458622 | 7,80169507726624E-06 | 0,006365228 | G > AG | 0,106508771 |
| **cg26258213** | chr10 | 125531752 | 125531752 | 1 | - | ENSG00000175018.13 | TEX36 | 151412 | 7,80524736453261E-06 | 0,006365228 | G > AG | 0,063972804 |
| **cg11612377** | chr22 | 42376900 | 42376900 | 1 | - | ENSG00000100207.21 | TCF20 | 33283 | 7,81754598551698E-06 | 0,006368468 | G > AG | 0,098257866 |
| **cg22950986** | chr3 | 133444333 | 133444333 | 1 | + | ENSG00000170819.5 | BFSP2 | 44278 | 7,8431886585672E-06 | 0,006378035 | G > AG | 0,091930076 |
| **cg12725334** | chr3 | 19914363 | 19914363 | 1 | + | ENSG00000144566.11 | RAB5A | 32733 | 7,87969928930558E-06 | 0,006398657 | G > AG | 0,080190151 |
| **cg05192402** | chr8 | 20818856 | 20818856 | 1 | + | ENSG00000147416.11 | ATP6V1B2 | 621476 | 7,90835649704467E-06 | 0,006401541 | G > AG | 0,085897277 |
| **cg11154727** | chr5 | 85893820 | 85893820 | 1 | + | ENSG00000127184.13 | COX7C | 724107 | 7,90773493612249E-06 | 0,006401541 | G > AG | 0,066609324 |
| **cg07874609** | chr2 | 119175115 | 119175115 | 1 | - | ENSG00000144119.4 | C1QL2 | 16363 | 7,98849550498821E-06 | 0,006432377 | G > AG | 0,086759018 |
| **cg24504068** | chr2 | 172889429 | 172889429 | 1 | - | ENSG00000115844.11 | DLX2 | 786528 | 7,97167988553175E-06 | 0,006432377 | G > AG | 0,089242407 |
| **cg23578867** | chr5 | 34863076 | 34863076 | 1 | - | ENSG00000113456.19 | RAD1 | 55914 | 7,97410607589971E-06 | 0,006432377 | G > AG | 0,04260915 |
| **cg02144950** | chr4 | 66565361 | 66565361 | 1 | + | ENSG00000035720.8 | STAP1 | 993365 | 7,98312753976184E-06 | 0,006432377 | G > AG | 0,152701434 |
| **cg12483269** | chr8 | 42377751 | 42377751 | 1 | + | ENSG00000078668.14 | VDAC3 | 13872 | 7,97032457313383E-06 | 0,006432377 | G > AG | 0,088139841 |
| **cg24485927** | chr2 | 795977 | 795977 | 1 | - | ENSG00000151353.15 | TMEM18 | 118570 | 8,01413013554412E-06 | 0,006446233 | G > AG | 0,090178637 |
| **cg04667734** | chr3 | 25454652 | 25454652 | 1 | + | ENSG00000077092.19 | RARB | 280321 | 8,02292484631998E-06 | 0,006446528 | G > AG | 0,107203347 |
| **cg19931260** | chr14 | 98633461 | 98633461 | 1 | - | ENSG00000127152.18 | BCL11B | 638737 | 8,10925596272209E-06 | 0,006495427 | G > AG | 0,098352899 |
| **cg20459739** | chr15 | 59739796 | 59739796 | 1 | + | ENSG00000140297.13 | GCNT3 | 144922 | 8,10145042211186E-06 | 0,006495427 | G > AG | 0,070989655 |
| **cg14072063** | chr4 | 187413363 | 187413363 | 1 | - | ENSG00000083857.14 | FAT1 | 686640 | 8,22241751913443E-06 | 0,006579179 | G > AG | 0,062535246 |
| **cg10125925** | chr2 | 11398186 | 11398186 | 1 | + | ENSG00000196208.14 | GREB1 | 84154 | 8,31613153713938E-06 | 0,006600293 | G > AG | 0,070132124 |
| **cg26825170** | chr11 | 96102984 | 96102984 | 1 | + | ENSG00000183340.7 | JRKL | 287004 | 8,28447677075895E-06 | 0,006600293 | G > AG | 0,080221256 |
| **cg19237900** | chr20 | 51354516 | 51354516 | 1 | + | ENSG00000124217.5 | MOCS3 | 395699 | 8,28190796538495E-06 | 0,006600293 | G > AG | 0,108431219 |

| | | | | | | | | | | | | |
|---|---|---|---|---|---|---|---|---|---|---|---|---|
| **cg27081488** | chr22 | 41487504 | 41487504 | 1 | - | ENSG00000100410.8 | PHF5A | 18811 | 8,29291706708835E-06 | 0,006600293 | G > AG | 0,111693537 |
| **cg07364240** | chr3 | 196661816 | 196661816 | 1 | + | ENSG00000163964.18 | PIGX | 22042 | 8,29196431124968E-06 | 0,006600293 | G > AG | 0,106465309 |
| **cg09993945** | chr10 | 68722533 | 68722533 | 1 | - | ENSG00000122912.15 | SLC25A16 | 195009 | 8,31783181295891E-06 | 0,006600293 | G > AG | 0,098742464 |
| **cg11275049** | chr3 | 78507000 | 78507000 | 1 | + | ENSG00000185008.17 | ROBO2 | 2600306 | 8,33309564044303E-06 | 0,006603768 | G > AG | 0,115147354 |
| **cg07969217** | chr12 | 72057495 | 72057495 | 1 | + | ENSG00000072657.10 | TRHDE | 29770 | 8,33947738106587E-06 | 0,006603768 | G > AG | 0,093107897 |
| **cg13907014** | chr1 | 87715234 | 87715234 | 1 | + | ENSG00000143013.13 | LMO4 | 386355 | 8,34918012410458E-06 | 0,006604614 | G > AG | 0,049488787 |
| **cg02305203** | chr4 | 56682107 | 56682107 | 1 | - | ENSG00000171476.22 | HOPX | 207 | 8,3807371584918E-06 | 0,006622729 | G > AG | 0,098044303 |
| **cg17923947** | chr14 | 50117287 | 50117287 | 1 | + | ENSG00000125375.17 | DMAC2L | 195036 | 8,41225323127574E-06 | 0,006638767 | G > AG | 0,098407823 |
| **cg07895745** | chr5 | 38796215 | 38796215 | 1 | - | ENSG00000113594.10 | LIFR | 187860 | 8,41839056999249E-06 | 0,006638767 | G > AG | 0,080879205 |
| **cg07758192** | chr15 | 84950449 | 84950449 | 1 | + | ENSG00000073417.15 | PDE8A | 29990 | 8,48017296927039E-06 | 0,006680602 | G > AG | 0,0806936 |
| **cg26201819** | chr18 | 45255415 | 45255415 | 1 | - | ENSG00000152223.15 | EPG5 | 711915 | 8,51640405884586E-06 | 0,00668162 | G > AG | 0,063291514 |
| **cg20348746** | chr2 | 187264082 | 187264082 | 1 | + | ENSG00000144369.13 | FAM171B | 570023 | 8,50966618395335E-06 | 0,00668162 | G > AG | 0,056440853 |
| **cg08225841** | chr1 | 56639226 | 56639226 | 1 | - | ENSG00000162407.9 | PLPP3 | 6076 | 8,50445962319782E-06 | 0,00668162 | G > AG | 0,13260289 |
| **cg09446373** | chr14 | 61517195 | 61517195 | 1 | + | ENSG00000258989.1 | RP11-47I22.4 | 11932 | 8,50759538297034E-06 | 0,00668162 | G > AG | 0,071327816 |
| **cg00449067** | chr2 | 187404716 | 187404716 | 1 | - | ENSG00000064989.13 | CALCRL | 43745 | 8,53451731348045E-06 | 0,00668897 | G > AG | 0,057303348 |
| **cg26630411** | chr16 | 47857221 | 47857221 | 1 | - | ENSG00000140798.16 | ABCC12 | 298798 | 8,56836565158844E-06 | 0,00669492 | G > AG | 0,073370755 |
| **cg02090654** | chr7 | 127058290 | 127058290 | 1 | + | ENSG00000004059.11 | ARF5 | 530095 | 8,55565682213315E-06 | 0,00669492 | G > AG | 0,10053728 |
| **cg05898830** | chr3 | 183153412 | 183153412 | 1 | - | ENSG00000078081.8 | LAMP3 | 10428 | 8,5596727968654E-06 | 0,00669492 | G > AG | 0,077801084 |
| **cg01999399** | chr3 | 195810637 | 195810637 | 1 | + | ENSG00000176945.17 | MUC20 | 89754 | 8,58620314734801E-06 | 0,006702012 | G > AG | 0,070943099 |
| **cg08803553** | chr13 | 68369840 | 68369840 | 1 | - | ENSG00000184226.15 | PCDH9 | 1139394 | 8,60824347972151E-06 | 0,006712366 | G > AG | 0,110902512 |
| **cg07970217** | chr17 | 72028567 | 72028567 | 1 | - | ENSG00000133195.11 | SLC39A11 | 1064146 | 8,62513680693572E-06 | 0,00671869 | G > AG | 0,051987852 |
| **cg24420884** | chr1 | 6354478 | 6354478 | 1 | - | ENSG00000097021.20 | ACOT7 | 39290 | 8,68060092094665E-06 | 0,006726303 | G > AG | 0,077428235 |
| **cg12583073** | chr3 | 72262785 | 72262785 | 1 | + | ENSG00000170837.3 | GPR27 | 508931 | 8,696461664712E-06 | 0,006726303 | G > AG | 0,086374052 |
| **cg01957834** | chr6 | 71393341 | 71393341 | 1 | + | ENSG00000119900.9 | OGFRL1 | 104531 | 8,68752110763432E-06 | 0,006726303 | G > AG | 0,104651156 |
| **cg01306259** | chr7 | 132422643 | 132422643 | 1 | - | ENSG00000221866.9 | PLXNA4 | 226046 | 8,69322171339912E-06 | 0,006726303 | G > AG | 0,119501799 |
| **cg24841501** | chr18 | 23932404 | 23932404 | 1 | + | ENSG00000168234.13 | TTC39C | 60368 | 8,66333242793659E-06 | 0,006726303 | G > AG | 0,111763318 |
| **cg25090542** | chr20 | 24800802 | 24800802 | 1 | + | ENSG00000077984.6 | CST7 | 148466 | 8,73338265210854E-06 | 0,006734431 | G > AG | 0,083841081 |
| **cg11190805** | chr7 | 70569030 | 70569030 | 1 | + | ENSG00000185274.12 | GALNT17 | 563113 | 8,72584862773084E-06 | 0,006734431 | G > AG | 0,108126841 |
| **cg13812829** | chr5 | 135085545 | 135085545 | 1 | + | ENSG00000152705.8 | CATSPER3 | 117639 | 8,74661091395353E-06 | 0,006737839 | G > AG | 0,086807914 |
| **cg12181306** | chr8 | 69712315 | 69712315 | 1 | + | ENSG00000137573.14 | SULF1 | 245692 | 8,77385523838291E-06 | 0,006752027 | G > AG | 0,088813499 |
| **cg13335506** | chr9 | 967032 | 967032 | 1 | - | ENSG00000172785.18 | CBWD1 | 787884 | 8,81107946790781E-06 | 0,006767058 | G > AG | 0,131542429 |

| | | | | | | | | | | | | |
|---|---|---|---|---|---|---|---|---|---|---|---|---|
| **cg04099793** | chr1 | 2917543 | 2917543 | 1 | - | ENSG00000215912.12 | TTC34 | 115825 | 8,83853277835249E-06 | 0,006774539 | G > AG | 0,066552267 |
| **cg16900740** | chr2 | 145769186 | 145769186 | 1 | - | ENSG00000169554.22 | ZEB2 | 1248128 | 8,83777938612446E-06 | 0,006774539 | G > AG | 0,104807608 |
| **cg19937288** | chr14 | 100733536 | 100733536 | 1 | + | ENSG00000185559.16 | DLK1 | 7832 | 8,87170240070494E-06 | 0,006779583 | G > AG | 0,072039341 |
| **cg27481083** | chr10 | 110323857 | 110323857 | 1 | + | ENSG00000119950.21 | MXI1 | 116253 | 8,8640692840845E-06 | 0,006779583 | G > AG | 0,088893394 |
| **cg05570616** | chr3 | 137020461 | 137020461 | 1 | + | ENSG00000174564.13 | IL20RB | 74232 | 8,89110811306319E-06 | 0,006784259 | G > AG | 0,081870225 |
| **cg19224418** | chr13 | 113856955 | 113856955 | 1 | + | ENSG00000184497.13 | TMEM255B | 97730 | 8,89555910827216E-06 | 0,006784259 | G > AG | 0,074948755 |
| **cg18716548** | chr13 | 48110974 | 48110974 | 1 | - | ENSG00000136146.15 | MED4 | 15842 | 8,93169631877798E-06 | 0,006791506 | G > AG | 0,129690368 |
| **cg15417269** | chr2 | 103056685 | 103056685 | 1 | + | ENSG00000170417.16 | TMEM182 | 319781 | 8,9298106858699E-06 | 0,006791506 | G > AG | 0,079769856 |
| **cg26204978** | chr1 | 20413426 | 20413426 | 1 | - | ENSG00000162545.6 | CAMK2N1 | 72785 | 9,02565741437976E-06 | 0,006849335 | G > AG | 0,113265853 |
| **cg04417513** | chr2 | 240973290 | 240973290 | 1 | + | ENSG00000162804.14 | SNED1 | 25327 | 9,08310645337647E-06 | 0,0068861 | G > AG | 0,07945856 |
| **cg03443220** | chr13 | 75800862 | 75800862 | 1 | + | ENSG00000136153.20 | LMO7 | 180429 | 9,10450753611502E-06 | 0,006888039 | G > AG | 0,094390384 |
| **cg08162457** | chr4 | 39228464 | 39228464 | 1 | - | ENSG00000035928.16 | RFC1 | 137912 | 9,11267786807708E-06 | 0,006888039 | G > AG | 0,10899126 |
| **cg22926422** | chr13 | 27988713 | 27988713 | 1 | - | ENSG00000183463.6 | URAD | 19 | 9,09569656487842E-06 | 0,006888039 | G > AG | 0,071955646 |
| **cg25441466** | chr13 | 107964642 | 107964642 | 1 | + | ENSG00000139826.6 | ABHD13 | 253749 | 9,13898126139258E-06 | 0,006901102 | G > AG | 0,135864034 |
| **cg19017633** | chr13 | 95100735 | 95100735 | 1 | - | ENSG00000125257.16 | ABCC4 | 200741 | 9,15388249503813E-06 | 0,006905537 | G > AG | 0,110711535 |
| **cg11307919** | chr3 | 48248330 | 48248330 | 1 | - | ENSG00000172113.10 | NME6 | 53356 | 9,18313934902154E-06 | 0,006918112 | G > AG | 0,073029207 |
| **cg18914863** | chr12 | 95740205 | 95740205 | 1 | - | ENSG00000074527.13 | NTN4 | 50985 | 9,27049526373329E-06 | 0,006972878 | G > AG | 0,076498394 |
| **cg23941527** | chr5 | 93532674 | 93532674 | 1 | - | ENSG00000248483.7 | POU5F2 | 208927 | 9,28166866384216E-06 | 0,006974425 | G > AG | 0,111088588 |
| **cg26642435** | chr5 | 163443780 | 163443780 | 1 | - | ENSG00000170584.11 | NUDCD2 | 16323 | 9,32269174060626E-06 | 0,006998376 | G > AG | 0,04969097 |
| **cg10698161** | chr7 | 79877107 | 79877107 | 1 | - | ENSG00000187391.22 | MAGI2 | 423439 | 9,36930108145448E-06 | 0,007019587 | G > AG | 0,071817724 |
| **cg12769917** | chr8 | 80301841 | 80301841 | 1 | - | ENSG00000076554.16 | TPD52 | 70608 | 9,36885457095748E-06 | 0,007019587 | G > AG | 0,089242058 |
| **cg22408697** | chr2 | 98356019 | 98356019 | 1 | + | ENSG00000144191.12 | CNGA3 | 9832 | 9,40020769379261E-06 | 0,007035851 | G > AG | 0,080877703 |
| **cg18549123** | chr13 | 28014488 | 28014488 | 1 | - | ENSG00000183463.6 | URAD | 25794 | 9,42752937856369E-06 | 0,007049403 | G > AG | 0,084910735 |
| **cg21149863** | chr7 | 13725365 | 13725365 | 1 | + | ENSG00000122644.13 | ARL4A | 1038510 | 9,45550475289338E-06 | 0,007056526 | G > AG | 0,098216199 |
| **cg26973852** | chr5 | 107254536 | 107254536 | 1 | + | ENSG00000151422.13 | FER | 1493304 | 9,48153264402215E-06 | 0,007059607 | G > AG | 0,067525456 |
| **cg23612423** | chr12 | 54956435 | 54956435 | 1 | - | ENSG00000135426.16 | TESPA1 | 28328 | 9,48269695422099E-06 | 0,007059607 | G > AG | 0,113861516 |
| **cg04258189** | chr11 | 125201585 | 125201585 | 1 | - | ENSG00000150433.10 | TMEM218 | 89821 | 9,48731975107902E-06 | 0,007059607 | G > AG | 0,054062622 |
| **cg11945884** | chr2 | 169953315 | 169953315 | 1 | + | ENSG00000144357.18 | UBR3 | 125862 | 9,53009250262336E-06 | 0,007077665 | G > AG | 0,110982388 |
| **cg25598038** | chr10 | 103929411 | 103929411 | 1 | - | ENSG00000107960.11 | STN 1,00 | 11226 | 9,56558236145017E-06 | 0,007097131 | G > AG | 0,096444856 |
| **cg11801842** | chr22 | 47629619 | 47629619 | 1 | - | ENSG00000100422.14 | CERK | 891366 | 9,64674841623059E-06 | 0,007129691 | G > AG | 0,126335495 |
| **cg17031787** | chr6 | 32081458 | 32081458 | 1 | + | ENSG00000231852.9 | CYP21A2 | 43132 | 9,64107406831051E-06 | 0,007129691 | G > AG | 0,085207396 |

| | | | | | | | | | | | | |
|---|---|---|---|---|---|---|---|---|---|---|---|---|
| **cg17195430** | chr5 | 44814550 | 44814550 | 1 | - | ENSG00000070193.5 | FGF10 | 424843 | 9,63564414290628E-06 | 0,007129691 | G > AG | 0,107793174 |
| **cg18574995** | chr12 | 8663038 | 8663038 | 1 | + | ENSG00000166532.16 | RIMKLB | 18561 | 9,64202301315955E-06 | 0,007129691 | G > AG | 0,103137372 |
| **cg03253615** | chr2 | 133517966 | 133517966 | 1 | + | ENSG00000152127.9 | MGAT5 | 602016 | 9,66969065643015E-06 | 0,007139749 | G > AG | 0,080586334 |
| **cg07966558** | chr17 | 79362071 | 79362071 | 1 | + | ENSG00000167280.17 | ENGASE | 287248 | 9,68357302537598E-06 | 0,007143104 | G > AG | 0,103762418 |
| **cg11265882** | chr4 | 18012314 | 18012314 | 1 | + | ENSG00000109805.10 | NCAPG | 201336 | 9,7018973753882E-06 | 0,007149726 | G > AG | 0,15542379 |
| **cg10613224** | chr2 | 201806860 | 201806860 | 1 | + | ENSG00000138395.17 | CDK15 | 16400 | 9,83469462532406E-06 | 0,007205937 | G > AG | 0,126793787 |
| **cg14452322** | chr9 | 130142326 | 130142326 | 1 | - | ENSG00000187239.17 | FNBP1 | 99131 | 9,81433215272373E-06 | 0,007205937 | G > AG | 0,052505923 |
| **cg02076598** | chr22 | 25027834 | 25027834 | 1 | + | ENSG00000197077.14 | KIAA1671 | 75119 | 9,81793211917027E-06 | 0,007205937 | G > AG | 0,115582553 |
| **cg14102355** | chr20 | 51811182 | 51811182 | 1 | - | ENSG00000101115.13 | SALL4 | 8660 | 9,79831910366571E-06 | 0,007205937 | G > AG | 0,128100781 |
| **cg18670716** | chr13 | 42576210 | 42576210 | 1 | - | ENSG00000133106.15 | EPSTI1 | 416062 | 9,86246766795039E-06 | 0,00720743 | G > AG | 0,115766007 |
| **cg07471527** | chr4 | 184251485 | 184251485 | 1 | + | ENSG00000164306.11 | PRIMPOL | 398181 | 9,85507102275438E-06 | 0,00720743 | G > AG | 0,093755216 |
| **cg13427006** | chr9 | 13444409 | 13444409 | 1 | + | ENSG00000153714.6 | LURAP1L | 669390 | 9,88709255982103E-06 | 0,007210833 | G > AG | 0,05503496 |
| **cg00169776** | chr1 | 3031948 | 3031948 | 1 | - | ENSG00000215912.12 | TTC34 | 230230 | 9,88850917244863E-06 | 0,007210833 | G > AG | 0,120962965 |
| **cg16422596** | chr4 | 32499388 | 32499388 | 1 | + | ENSG00000169851.15 | PCDH7 | 1778974 | 9,92717383981022E-06 | 0,007232133 | G > AG | 0,072111468 |
| **cg13706784** | chr16 | 964975 | 964975 | 1 | - | ENSG00000103227.19 | LMF1 | 16344 | 9,94544289803037E-06 | 0,007238549 | G > AG | 0,129546506 |
| **cg21449646** | chr8 | 19608125 | 19608125 | 1 | + | ENSG00000104613.12 | INTS10 | 209265 | 9,96455585560886E-06 | 0,007242577 | G > AG | 0,069231093 |
| **cg08787261** | chr19 | 4671090 | 4671090 | 1 | - | ENSG00000074842.8 | MYDGF | 727 | 1,00163431602084E-05 | 0,007255634 | G > AG | 0,101404702 |
| **cg21011326** | chr16 | 2823459 | 2823459 | 1 | + | ENSG00000007038.11 | PRSS21 | 6280 | 1,0003166355876E-05 | 0,007255634 | G > AG | 0,097235224 |
| **cg27000059** | chr15 | 95036036 | 95036036 | 1 | - | ENSG00000182175.14 | RGMA | 1946831 | 1,00127898596664E-05 | 0,007255634 | G > AG | 0,079615194 |
| **cg14059881** | chr6 | 140542845 | 140542845 | 1 | + | ENSG00000112406.5 | HECA | 1407766 | 1,00792242512034E-05 | 0,007273632 | G > AG | 0,093743245 |
| **cg24460327** | chr1 | 217403211 | 217403211 | 1 | + | ENSG00000162814.11 | SPATA17 | 228112 | 1,00732705293098E-05 | 0,007273632 | G > AG | 0,076625844 |
| **cg07632981** | chr5 | 5344660 | 5344660 | 1 | + | ENSG00000164151.12 | ICE1 | 76003 | 1,01167752457927E-05 | 0,00729385 | G > AG | 0,101012269 |
| **cg12033914** | chr3 | 179903669 | 179903669 | 1 | + | ENSG00000058056.10 | USP13 | 250638 | 1,01328849655351E-05 | 0,007298585 | G > AG | 0,08872194 |
| **cg07544152** | chr10 | 17953223 | 17953223 | 1 | + | ENSG00000148482.12 | SLC39A12 | 1385 | 1,01496267981206E-05 | 0,007303767 | G > AG | 0,071162094 |
| **cg27266061** | chr15 | 43206594 | 43206594 | 1 | + | ENSG00000166946.14 | CCNDBP1 | 21477 | 1,02217125225323E-05 | 0,007341827 | G > AG | 0,06504214 |
| **cg22851420** | chr1 | 39683978 | 39683978 | 1 | - | ENSG00000116983.13 | HPCAL4 | 7508 | 1,02957646648641E-05 | 0,00737327 | G > AG | 0,145192565 |
| **cg12757676** | chr2 | 100183096 | 100183096 | 1 | + | ENSG00000204640.1 | NMS | 287385 | 1,03099442180063E-05 | 0,00737327 | G > AG | 0,088793243 |
| **cg25418694** | chr1 | 47934666 | 47934666 | 1 | + | ENSG00000117834.13 | SLC5A9 | 288018 | 1,02941629300976E-05 | 0,00737327 | G > AG | 0,083662756 |
| **cg04579753** | chr3 | 13950480 | 13950480 | 1 | - | ENSG00000154764.6 | WNT7A | 70408 | 1,03136841601411E-05 | 0,00737327 | G > AG | 0,106253907 |
| **cg22358705** | chr17 | 27455710 | 27455710 | 1 | - | ENSG00000007171.18 | NOS2 | 344820 | 1,03585187688942E-05 | 0,007398408 | G > AG | 0,05520249 |
| **cg21143225** | chr2 | 235331338 | 235331338 | 1 | + | ENSG00000157985.19 | AGAP1 | 162704 | 1,04244155702225E-05 | 0,007431596 | G > AG | 0,079635609 |

| | | | | | | | | | | | | |
|---|---|---|---|---|---|---|---|---|---|---|---|---|
| **cg03344098** | chrX | 149146219 | 149146219 | 1 | - | ENSG00000287585.1 | RP11-559E14.1 | 269277 | 1,04531044440712E-05 | 0,00744511 | G > AG | 0,085056029 |
| **cg15671845** | chr7 | 41701722 | 41701722 | 1 | + | ENSG00000106591.4 | MRPL32 | 1230653 | 1,04866566058427E-05 | 0,007448202 | G > AG | 0,116597024 |
| **cg06917199** | chr15 | 34800077 | 34800077 | 1 | + | ENSG00000184507.16 | NUTM1 | 456763 | 1,04802601300915E-05 | 0,007448202 | G > AG | 0,087398386 |
| **cg26781758** | chr20 | 37343094 | 37343094 | 1 | + | ENSG00000197122.12 | SRC | 1590 | 1,0485741195516E-05 | 0,007448202 | G > AG | 0,05514019 |
| **cg18336525** | chr19 | 33246268 | 33246268 | 1 | + | ENSG00000130881.14 | LRP3 | 68666 | 1,05535092293594E-05 | 0,007467948 | G > AG | 0,076795528 |
| **cg02132307** | chr10 | 128429567 | 128429567 | 1 | - | ENSG00000148773.14 | MKI67 | 303143 | 1,05351553942325E-05 | 0,007467948 | G > AG | 0,069674328 |
| **cg09911329** | chr6 | 105676227 | 105676227 | 1 | - | ENSG00000085377.15 | PREP | 222164 | 1,0568672346527E-05 | 0,007471766 | G > AG | 0,079498286 |
| **cg03049399** | chr11 | 82718798 | 82718798 | 1 | + | ENSG00000165490.13 | DDIAS | 181176 | 1,06592799064245E-05 | 0,007528865 | G > AG | 0,077613355 |
| **cg07077965** | chr2 | 176638864 | 176638864 | 1 | + | ENSG00000128654.14 | MTX2 | 369470 | 1,07121795227758E-05 | 0,007552282 | G > AG | 0,102325041 |
| **cg01582822** | chr18 | 44245353 | 44245353 | 1 | + | ENSG00000152217.20 | SETBP1 | 434819 | 1,07071020473089E-05 | 0,007552282 | G > AG | 0,099295145 |
| **cg00974578** | chr6 | 143595348 | 143595348 | 1 | - | ENSG00000001036.14 | FUCA2 | 83627 | 1,07762238369264E-05 | 0,007587201 | G > AG | 0,066311566 |
| **cg03979421** | chr2 | 197576833 | 197576833 | 1 | - | ENSG00000144381.18 | HSPD1 | 60095 | 1,07887187662701E-05 | 0,007587201 | G > AG | 0,125336519 |
| **cg08162952** | chr11 | 125931561 | 125931561 | 1 | + | ENSG00000283703.2 | VSIG10L2 | 14494 | 1,07914643247829E-05 | 0,007587201 | G > AG | 0,092420503 |
| **cg06804657** | chr6 | 145347987 | 145347987 | 1 | + | ENSG00000152822.14 | GRM1 | 679658 | 1,08190769516876E-05 | 0,00759963 | G > AG | 0,089849113 |
| **cg24367037** | chr19 | 29835404 | 29835404 | 1 | + | ENSG00000105173.14 | CCNE1 | 23414 | 1,0855422654582E-05 | 0,007614117 | G > AG | 0,068589392 |
| **cg01679364** | chr1 | 208613622 | 208613622 | 1 | - | ENSG00000076356.7 | PLXNA2 | 369237 | 1,08596087237532E-05 | 0,007614117 | G > AG | 0,084801954 |
| **cg07137983** | chr6 | 135387891 | 135387891 | 1 | - | ENSG00000135541.22 | AHI1 | 110544 | 1,09067735261071E-05 | 0,007616915 | G > AG | 0,103889864 |
| **cg05237114** | chr8 | 57993113 | 57993113 | 1 | - | ENSG00000167910.4 | CYP7A1 | 507051 | 1,09133869331939E-05 | 0,007616915 | G > AG | 0,094496112 |
| **cg26769081** | chr4 | 182212293 | 182212293 | 1 | - | ENSG00000129187.15 | DCTD | 705644 | 1,08996130620683E-05 | 0,007616915 | G > AG | 0,115018887 |
| **cg17554250** | chr2 | 219844653 | 219844653 | 1 | - | ENSG00000124006.15 | OBSL1 | 272793 | 1,08792338226966E-05 | 0,007616915 | G > AG | 0,109548135 |
| **cg22823987** | chr5 | 56585778 | 56585778 | 1 | + | ENSG00000095015.6 | MAP3K1 | 229770 | 1,09402380501893E-05 | 0,007621747 | G > AG | 0,107153349 |
| **cg25054860** | chr20 | 21089537 | 21089537 | 1 | + | ENSG00000088970.16 | KIZ | 36445 | 1,09851519625981E-05 | 0,007646074 | G > AG | 0,128819914 |
| **cg13156098** | chr8 | 133642582 | 133642582 | 1 | + | ENSG00000104415.14 | CCN4 | 451544 | 1,10017614314285E-05 | 0,007650673 | G > AG | 0,066820833 |
| **cg14254615** | chr9 | 126058832 | 126058832 | 1 | + | ENSG00000196814.15 | MVB12B | 267996 | 1,10250424828296E-05 | 0,007659899 | G > AG | 0,080709337 |
| **cg22513076** | chr17 | 39848265 | 39848265 | 1 | - | ENSG00000161405.17 | IKZF3 | 16048 | 1,10363884463033E-05 | 0,007660824 | G > AG | 0,071693062 |
| **cg24515979** | chr19 | 39470729 | 39470729 | 1 | - | ENSG00000105193.9 | RPS16 | 34779 | 1,10576149069699E-05 | 0,007668599 | G > AG | 0,096559725 |
| **cg10529470** | chr5 | 172339419 | 172339419 | 1 | + | ENSG00000214360.5 | EFCAB9 | 145248 | 1,11741682505192E-05 | 0,007740201 | G > AG | 0,076669841 |
| **cg14551162** | chr3 | 152639266 | 152639266 | 1 | - | ENSG00000152580.9 | IGSF10 | 1178204 | 1,12064604603054E-05 | 0,007743743 | G > AG | 0,094948107 |
| **cg25372239** | chr1 | 3213986 | 3213986 | 1 | - | ENSG00000162591.16 | MEGF6 | 397523 | 1,11994597821153E-05 | 0,007743743 | G > AG | 0,082437308 |
| **cg25174728** | chr14 | 66485314 | 66485314 | 1 | + | ENSG00000171723.16 | GPHN | 22092 | 1,12191147411639E-05 | 0,007745491 | G > AG | 0,086694461 |
| **cg14655541** | chr9 | 134208620 | 134208620 | 1 | + | ENSG00000196363.10 | WDR5 | 73256 | 1,12435020798872E-05 | 0,007748341 | G > AG | 0,060818829 |

| | | | | | | | | | | | | |
|---|---|---|---|---|---|---|---|---|---|---|---|---|
| **cg11471498** | chr2 | 110667357 | 110667357 | 1 | - | ENSG00000169679.15 | BUB1 | 10707 | 1,12906328649172E-05 | 0,007773817 | G > AG | 0,109715307 |
| **cg14453145** | chr2 | 241859744 | 241859744 | 1 | + | ENSG00000188011.5 | RTP5 | 9855 | 1,13494601899828E-05 | 0,007807294 | G > AG | 0,115851636 |
| **cg03757901** | chr2 | 166901628 | 166901628 | 1 | + | ENSG00000163092.21 | XIRP2 | 13149 | 1,13802138369822E-05 | 0,007814395 | G > AG | 0,085018706 |
| **cg02614428** | chr10 | 77081659 | 77081659 | 1 | + | ENSG00000138326.21 | RPS24 | 952100 | 1,14457457016195E-05 | 0,007842367 | G > AG | 0,115559494 |
| **cg16431025** | chr6 | 25629278 | 25629278 | 1 | + | ENSG00000079689.14 | SCGN | 22922 | 1,14517066187169E-05 | 0,007842367 | G > AG | 0,075658155 |
| **cg18899818** | chr6 | 87095315 | 87095315 | 1 | - | ENSG00000135346.9 | CGA | 208 | 1,15359334283608E-05 | 0,007892981 | G > AG | 0,118254389 |
| **cg07321742** | chr20 | 8715419 | 8715419 | 1 | + | ENSG00000101333.18 | PLCB4 | 353343 | 1,15512823103434E-05 | 0,00789642 | G > AG | 0,076524472 |
| **cg00589411** | chr2 | 212227946 | 212227946 | 1 | - | ENSG00000178568.15 | ERBB4 | 310896 | 1,15694094717114E-05 | 0,007900164 | G > AG | 0,116579045 |
| **cg00050482** | chr16 | 73173314 | 73173314 | 1 | - | ENSG00000140836.17 | ZFHX3 | 718558 | 1,15774153018748E-05 | 0,007900164 | G > AG | 0,089936276 |
| **cg12322456** | chr5 | 180440149 | 180440149 | 1 | + | ENSG00000113300.13 | CNOT6 | 54229 | 1,16376414438949E-05 | 0,00792438 | G > AG | 0,072676245 |
| **cg05162870** | chr3 | 134852324 | 134852324 | 1 | - | ENSG00000174611.12 | KY | 200687 | 1,16245869559864E-05 | 0,00792438 | G > AG | 0,074164737 |
| **cg23716516** | chr1 | 186566606 | 186566606 | 1 | + | ENSG00000157181.16 | ODR4 | 190769 | 1,16516428004171E-05 | 0,00792438 | G > AG | 0,126113026 |
| **cg03003919** | chr8 | 84175678 | 84175678 | 1 | - | ENSG00000176731.12 | RBIS | 1044744 | 1,16543410658716E-05 | 0,00792438 | G > AG | 0,084848825 |
| **cg14025373** | chr8 | 1048462 | 1048462 | 1 | - | ENSG00000104714.14 | ERICH1 | 310355 | 1,1676291757503E-05 | 0,00792919 | G > AG | 0,082113998 |
| **cg01966066** | chr17 | 10069585 | 10069585 | 1 | - | ENSG00000007237.19 | GAS7 | 129022 | 1,16821458837889E-05 | 0,00792919 | G > AG | 0,113916003 |
| **cg03489615** | chr7 | 46860328 | 46860328 | 1 | + | ENSG00000146678.10 | IGFBP1 | 971969 | 1,17335921641657E-05 | 0,007951947 | G > AG | 0,067776677 |
| **cg16491547** | chr10 | 113104879 | 113104879 | 1 | + | ENSG00000148737.17 | TCF7L2 | 154633 | 1,17364658893281E-05 | 0,007951947 | G > AG | 0,077537156 |
| **cg09305825** | chr1 | 102628516 | 102628516 | 1 | - | ENSG00000060718.22 | COL11A1 | 480357 | 1,17518705160332E-05 | 0,007955338 | G > AG | 0,09572058 |
| **cg24699271** | chr3 | 4596900 | 4596900 | 1 | - | ENSG00000144455.14 | SUMF1 | 129626 | 1,17717316280262E-05 | 0,007961737 | G > AG | 0,066113443 |
| **cg18558487** | chr13 | 28854635 | 28854635 | 1 | - | ENSG00000139508.15 | SLC46A3 | 135664 | 1,18152014083369E-05 | 0,007984079 | G > AG | 0,081035536 |
| **cg09366519** | chr1 | 209704625 | 209704625 | 1 | - | ENSG00000196878.15 | LAMB3 | 52199 | 1,18386567539216E-05 | 0,007988151 | G > AG | 0,064085934 |
| **cg24631102** | chr10 | 127551380 | 127551380 | 1 | + | ENSG00000214285.3 | NPS | 2072 | 1,18472567578376E-05 | 0,007988151 | G > AG | 0,077691423 |
| **cg04209885** | chr10 | 132296251 | 132296251 | 1 | + | ENSG00000148814.18 | LRRC27 | 35902 | 1,19344833687883E-05 | 0,008036286 | G > AG | 0,059825029 |
| **cg24693364** | chr10 | 13827492 | 13827492 | 1 | - | ENSG00000165626.18 | BEND7 | 298517 | 1,19817507365073E-05 | 0,008046864 | G > AG | 0,08455959 |
| **cg07242369** | chr4 | 94298776 | 94298776 | 1 | + | ENSG00000163104.18 | SMARCAD1 | 91166 | 1,19705550606568E-05 | 0,008046864 | G > AG | 0,105961108 |
| **cg04202610** | chr2 | 223815122 | 223815122 | 1 | - | ENSG00000152056.17 | AP1S3 | 22906 | 1,20513531550238E-05 | 0,008086509 | G > AG | 0,135859759 |
| **cg04403850** | chr7 | 959345 | 959345 | 1 | - | ENSG00000105963.15 | ADAP1 | 3937 | 1,20816416777861E-05 | 0,008090519 | G > AG | 0,106995432 |
| **cg24432193** | chr6 | 101614858 | 101614858 | 1 | - | ENSG00000112249.14 | ASCC3 | 733485 | 1,20744199241227E-05 | 0,008090519 | G > AG | 0,082622222 |
| **cg09424398** | chr16 | 80900443 | 80900443 | 1 | + | ENSG00000166451.14 | CENPN | 106108 | 1,21151410767576E-05 | 0,008090519 | G > AG | 0,119736053 |
| **cg02281238** | chr18 | 49369110 | 49369110 | 1 | + | ENSG00000101670.12 | LIPG | 191588 | 1,21039281457466E-05 | 0,008090519 | G > AG | 0,075077277 |
| **cg24994790** | chr20 | 14361964 | 14361964 | 1 | + | ENSG00000172264.18 | MACROD2 | 366596 | 1,2104100372302E-05 | 0,008090519 | G > AG | 0,094799174 |

| | | | | | | | | | | | | |
|---|---|---|---|---|---|---|---|---|---|---|---|---|
| **cg04064631** | chr7 | 67391393 | 67391393 | 1 | - | ENSG00000126524.10 | SBDS | 395805 | 1,21207884959703E-05 | 0,008090519 | G > AG | 0,071420849 |
| **cg16837310** | chr4 | 184284398 | 184284398 | 1 | + | ENSG00000164306.11 | PRIMPOL | 365268 | 1,21557853925007E-05 | 0,008106805 | G > AG | 0,092648012 |
| **cg03596645** | chr17 | 39255100 | 39255100 | 1 | + | ENSG00000108298.12 | RPL19 | 54818 | 1,23271033263431E-05 | 0,008199612 | G > AG | 0,091867631 |
| **cg16167369** | chr11 | 31903636 | 31903636 | 1 | + | ENSG00000285283.1 | RP1-65P5.6 | 91246 | 1,23744222885591E-05 | 0,008223936 | G > AG | 0,061973177 |
| **cg21068293** | chr15 | 74204235 | 74204235 | 1 | + | ENSG00000140481.15 | CCDC33 | 1531 | 1,24050678091386E-05 | 0,008230002 | G > AG | 0,070108461 |
| **cg17455324** | chr12 | 32542001 | 32542001 | 1 | - | ENSG00000139131.13 | YARS2 | 213897 | 1,23956923547686E-05 | 0,008230002 | G > AG | 0,094429914 |
| **cg04527612** | chr2 | 67130508 | 67130508 | 1 | - | ENSG00000197223.12 | C1D | 980441 | 1,2442938084051E-05 | 0,008242398 | G > AG | 0,078066027 |
| **cg19570267** | chr2 | 219729426 | 219729426 | 1 | + | ENSG00000114923.17 | SLC4A3 | 102033 | 1,24564232866058E-05 | 0,008242627 | G > AG | 0,103831244 |
| **cg17450053** | chr3 | 150903905 | 150903905 | 1 | + | ENSG00000144893.12 | MED12L | 181791 | 1,24730928225469E-05 | 0,008246523 | G > AG | 0,077814631 |
| **cg09237746** | chr6 | 18507072 | 18507072 | 1 | - | ENSG00000124795.17 | DEK | 242523 | 1,25687568603106E-05 | 0,008301294 | G > AG | 0,078382892 |
| **cg26584481** | chrX | 657730 | 657730 | 1 | - | ENSG00000167393.18 | PPP2R3B | 270774 | 1,25776392549327E-05 | 0,008301294 | G > AG | 0,110656871 |
| **cg27624408** | chr20 | 58544508 | 58544508 | 1 | - | ENSG00000198768.11 | APCDD1L | 29108 | 1,26170873061765E-05 | 0,008320151 | G > AG | 0,068427232 |
| **cg21572511** | chr8 | 85351033 | 85351033 | 1 | + | ENSG00000164879.7 | CA3 | 22402 | 1,26400947117163E-05 | 0,008325529 | G > AG | 0,088084248 |
| **cg19821125** | chr14 | 89139898 | 89139898 | 1 | + | ENSG00000165533.19 | TTC8 | 315746 | 1,26470101705181E-05 | 0,008325529 | G > AG | 0,049217597 |
| **cg25688251** | chr16 | 83887796 | 83887796 | 1 | + | ENSG00000103150.7 | MLYCD | 11318 | 1,26691652391119E-05 | 0,008325784 | G > AG | 0,069063715 |
| **cg12294112** | chr12 | 77163063 | 77163063 | 1 | + | ENSG00000067798.16 | NAV3 | 161577 | 1,26661465237939E-05 | 0,008325784 | G > AG | 0,150836442 |
| **cg17337307** | chr6 | 19380550 | 19380550 | 1 | - | ENSG00000172197.11 | MBOAT1 | 831920 | 1,27139880905875E-05 | 0,008348068 | G > AG | 0,07474216 |
| **cg23497078** | chr18 | 45517401 | 45517401 | 1 | - | ENSG00000152223.15 | EPG5 | 449929 | 1,27506481743644E-05 | 0,008357791 | G > AG | 0,098939014 |
| **cg15096829** | chr1 | 202122877 | 202122877 | 1 | + | ENSG00000170075.10 | GPR37L1 | 8 | 1,27487594143821E-05 | 0,008357791 | G > AG | 0,090933673 |
| **cg15684236** | chr12 | 126742596 | 126742596 | 1 | + | ENSG00000181234.10 | TMEM132C | 1524573 | 1,2808797962886E-05 | 0,008381543 | G > AG | 0,067631013 |
| **cg00997054** | chr3 | 32862799 | 32862799 | 1 | - | ENSG00000188167.9 | TMPPE | 234348 | 1,28332821180758E-05 | 0,008390387 | G > AG | 0,070676532 |
| **cg11281485** | chr5 | 125550347 | 125550347 | 1 | + | ENSG00000155324.10 | GRAMD2B | 809784 | 1,28448031423559E-05 | 0,008390748 | G > AG | 0,101540856 |
| **cg26145091** | chr22 | 25025441 | 25025441 | 1 | - | ENSG00000206069.7 | TMEM211 | 78745 | 1,30167961135589E-05 | 0,008481372 | G > AG | 0,071771191 |
| **cg11369836** | chr10 | 69908511 | 69908511 | 1 | - | ENSG00000042286.15 | AIFM2 | 224424 | 1,30820362469203E-05 | 0,008494937 | G > AG | 0,079618307 |
| **cg12374044** | chr13 | 107202685 | 107202685 | 1 | - | ENSG00000134884.15 | ARGLU1 | 634547 | 1,30749506322003E-05 | 0,008494937 | G > AG | 0,0993134 |
| **cg09432202** | chr6 | 163347519 | 163347519 | 1 | - | ENSG00000185345.23 | PRKN | 619743 | 1,30630948151276E-05 | 0,008494937 | G > AG | 0,062378563 |
| **cg20786944** | chr21 | 28233700 | 28233700 | 1 | - | ENSG00000156239.12 | N6AMT1 | 651672 | 1,31553276186865E-05 | 0,008535284 | G > AG | 0,095910246 |
| **cg17315703** | chr8 | 29721512 | 29721512 | 1 | + | ENSG00000104660.19 | LEPROTL1 | 373895 | 1,31813723798727E-05 | 0,008539639 | G > AG | 0,068029221 |
| **cg09975822** | chr6 | 112298447 | 112298447 | 1 | + | ENSG00000251258.2 | RFPL4B | 48882 | 1,31926881680876E-05 | 0,008539639 | G > AG | 0,075085118 |
| **cg02745692** | chr9 | 91060644 | 91060644 | 1 | + | ENSG00000165025.15 | SYK | 258858 | 1,31955318972475E-05 | 0,008539639 | G > AG | 0,131637983 |
| **cg25919521** | chr5 | 93681468 | 93681468 | 1 | + | ENSG00000175745.14 | NR2F1 | 98247 | 1,32088298384567E-05 | 0,008541019 | G > AG | 0,071853935 |

| | | | | | | | | | | | | |
|---|---|---|---|---|---|---|---|---|---|---|---|---|
| **cg23199907** | chr13 | 32731829 | 32731829 | 1 | - | ENSG00000244754.9 | N4BP2L2 | 192943 | 1,32768996720983E-05 | 0,008548902 | G > AG | 0,079793704 |
| **cg05922437** | chr15 | 70582587 | 70582587 | 1 | - | ENSG00000137831.15 | UACA | 180972 | 1,32754663145005E-05 | 0,008548902 | G > AG | 0,068144022 |
| **cg25573906** | chr3 | 123784190 | 123784190 | 1 | - | ENSG00000065534.19 | MYLK | 100143 | 1,33142803771097E-05 | 0,008565761 | G > AG | 0,093358475 |
| **cg23510570** | chr18 | 46771369 | 46771369 | 1 | + | ENSG00000167216.18 | KATNAL2 | 146122 | 1,33434922117327E-05 | 0,00857038 | G > AG | 0,111427502 |
| **cg11920099** | chr7 | 151868029 | 151868029 | 1 | - | ENSG00000106617.15 | PRKAG2 | 9097 | 1,33438687074453E-05 | 0,00857038 | G > AG | 0,076077184 |
| **cg05316110** | chr3 | 106699125 | 106699125 | 1 | - | ENSG00000114423.23 | CBLB | 829572 | 1,33615580218075E-05 | 0,008574542 | G > AG | 0,087630824 |
| **cg14783517** | chr1 | 119637387 | 119637387 | 1 | - | ENSG00000143067.5 | ZNF697 | 10880 | 1,33945815006724E-05 | 0,008588529 | G > AG | 0,082543674 |
| **cg27572696** | chr10 | 32681391 | 32681391 | 1 | + | ENSG00000216937.13 | CCDC7 | 235252 | 1,34179651749218E-05 | 0,008596317 | G > AG | 0,072303668 |
| **cg24818118** | chr12 | 106284387 | 106284387 | 1 | + | ENSG00000166046.11 | TCP11L2 | 17541 | 1,34574722230008E-05 | 0,008614413 | G > AG | 0,08510213 |
| **cg03466127** | chr2 | 127419523 | 127419523 | 1 | + | ENSG00000115718.18 | PROC | 1097 | 1,35117761532297E-05 | 0,008629315 | G > AG | 0,096980512 |
| **cg06960850** | chr4 | 111885131 | 111885131 | 1 | - | ENSG00000145365.11 | TIFA | 400774 | 1,35213983616195E-05 | 0,008629315 | G > AG | 0,068228973 |
| **cg18947376** | chr22 | 17832006 | 17832006 | 1 | + | ENSG00000099968.18 | BCL2L13 | 203152 | 1,35574060474305E-05 | 0,008635855 | G > AG | 0,070542783 |
| **cg22331723** | chr14 | 67446831 | 67446831 | 1 | - | ENSG00000100558.9 | PLEK2 | 34663 | 1,3558706355749E-05 | 0,008635855 | G > AG | 0,08773237 |
| **cg17063595** | chr5 | 154118904 | 154118904 | 1 | - | ENSG00000055147.19 | FAM114A2 | 79967 | 1,35958038098469E-05 | 0,008652279 | G > AG | 0,062927408 |
| **cg14905382** | chr1 | 217881732 | 217881732 | 1 | + | ENSG00000162814.11 | SPATA17 | 250409 | 1,36161156299939E-05 | 0,008658002 | G > AG | 0,078089907 |
| **cg04118554** | chr2 | 215218722 | 215218722 | 1 | - | ENSG00000144452.15 | ABCA12 | 80095 | 1,37129188074129E-05 | 0,008712314 | G > AG | 0,07495667 |
| **cg09288558** | chr17 | 43638123 | 43638123 | 1 | + | ENSG00000231256.8 | CFAP97D1 | 142311 | 1,37246662329232E-05 | 0,008712541 | G > AG | 0,08640655 |
| **cg16843910** | chr8 | 903424 | 903424 | 1 | + | ENSG00000198010.13 | DLGAP2 | 165797 | 1,37717245117123E-05 | 0,008729038 | G > AG | 0,112217241 |
| **cg23234561** | chr18 | 9877411 | 9877411 | 1 | + | ENSG00000168454.12 | TXNDC2 | 8314 | 1,37734755846695E-05 | 0,008729038 | G > AG | 0,104676484 |
| **cg14930065** | chr8 | 37815089 | 37815089 | 1 | - | ENSG00000104221.13 | BRF2 | 34773 | 1,38207639291883E-05 | 0,008751756 | G > AG | 0,072518393 |
| **cg25185907** | chr10 | 31104356 | 31104356 | 1 | + | ENSG00000148516.23 | ZEB1 | 214138 | 1,38917190395526E-05 | 0,008789411 | G > AG | 0,071063437 |
| **cg15788537** | chr11 | 8357280 | 8357280 | 1 | - | ENSG00000166407.14 | LMO1 | 88563 | 1,39141431195709E-05 | 0,008796324 | G > AG | 0,122702123 |
| **cg11270449** | chr13 | 46094696 | 46094696 | 1 | - | ENSG00000080618.17 | CPB2 | 10338 | 1,39632950213141E-05 | 0,008799735 | G > AG | 0,051251658 |
| **cg14172765** | chr2 | 76323713 | 76323713 | 1 | + | ENSG00000115364.14 | MRPL19 | 676931 | 1,39655544567536E-05 | 0,008799735 | G > AG | 0,074207234 |
| **cg27537673** | chr9 | 14516069 | 14516069 | 1 | - | ENSG00000147862.17 | NFIB | 117085 | 1,3962193425588E-05 | 0,008799735 | G > AG | 0,084724439 |
| **cg13602478** | chr4 | 159398442 | 159398442 | 1 | - | ENSG00000164123.7 | C4orf45 | 359681 | 1,40112411948083E-05 | 0,008803972 | G > AG | 0,063272844 |
| **cg04959549** | chr4 | 26298500 | 26298500 | 1 | - | ENSG00000163394.6 | CCKAR | 191985 | 1,3984772674472E-05 | 0,008803972 | G > AG | 0,15195183 |
| **cg12897600** | chr7 | 126699182 | 126699182 | 1 | - | ENSG00000179603.18 | GRM8 | 553912 | 1,40036052982678E-05 | 0,008803972 | G > AG | 0,12701804 |
| **cg07850246** | chr11 | 4578407 | 4578407 | 1 | - | ENSG00000167333.13 | TRIM68 | 29825 | 1,40183155763432E-05 | 0,008803972 | G > AG | 0,061403493 |
| **cg00778996** | chr8 | 6417977 | 6417977 | 1 | - | ENSG00000091879.14 | ANGPT2 | 145433 | 1,40686909888477E-05 | 0,008828361 | G > AG | 0,054574901 |
| **cg24973733** | chr11 | 122817013 | 122817013 | 1 | - | ENSG00000188909.5 | BSX | 164822 | 1,4111051139104E-05 | 0,008840438 | G > AG | 0,104674646 |

| | | | | | | | | | | | | |
|---|---|---|---|---|---|---|---|---|---|---|---|---|
| **cg07278020** | chr2 | 164569494 | 164569494 | 1 | + | ENSG00000136531.19 | SCN2A | 625498 | 1,41055532417953E-05 | 0,008840438 | G > AG | 0,107722554 |
| **cg22031783** | chr1 | 3212042 | 3212042 | 1 | + | ENSG00000142611.17 | PRDM16 | 142875 | 1,41854655343383E-05 | 0,008879786 | G > AG | 0,081355061 |
| **cg17692087** | chr22 | 35051060 | 35051060 | 1 | + | ENSG00000175329.13 | ISX | 15075 | 1,42025256195656E-05 | 0,008883196 | G > AG | 0,08246341 |
| **cg00886293** | chr20 | 51339873 | 51339873 | 1 | + | ENSG00000124217.5 | MOCS3 | 381056 | 1,42180742721977E-05 | 0,008885655 | G > AG | 0,097674828 |
| **cg13787775** | chr1 | 204602130 | 204602130 | 1 | + | ENSG00000198625.13 | MDM4 | 85752 | 1,42401210704119E-05 | 0,008892169 | G > AG | 0,093104313 |
| **cg22529989** | chr11 | 31238281 | 31238281 | 1 | + | ENSG00000170946.15 | DNAJC24 | 131558 | 1,42662810698023E-05 | 0,008897902 | G > AG | 0,048529036 |
| **cg24986119** | chr20 | 13262124 | 13262124 | 1 | + | ENSG00000101230.6 | ISM1 | 40851 | 1,42958310979925E-05 | 0,008897902 | G > AG | 0,085407731 |
| **cg03421195** | chr10 | 7475253 | 7475253 | 1 | + | ENSG00000151655.19 | ITIH2 | 228062 | 1,42925315435406E-05 | 0,008897902 | G > AG | 0,077131443 |
| **cg16121641** | chr7 | 37665984 | 37665984 | 1 | - | ENSG00000155849.16 | ELMO1 | 216760 | 1,43153639258833E-05 | 0,008902816 | G > AG | 0,074941248 |
| **cg16206813** | chr5 | 3122048 | 3122048 | 1 | - | ENSG00000170561.13 | IRX2 | 370370 | 1,43995317232712E-05 | 0,008947885 | G > AG | 0,104496849 |
| **cg15587274** | chr10 | 112792822 | 112792822 | 1 | - | ENSG00000023041.12 | ZDHHC6 | 345249 | 1,44297230998708E-05 | 0,008959368 | G > AG | 0,100714237 |
| **cg17360736** | chr6 | 40733541 | 40733541 | 1 | - | ENSG00000156564.9 | LRFN2 | 146176 | 1,44420565914019E-05 | 0,008959754 | G > AG | 0,094054862 |
| **cg24007402** | chr10 | 62770487 | 62770487 | 1 | + | ENSG00000181915.5 | ADO | 34232 | 1,44568874007643E-05 | 0,008961686 | G > AG | 0,100428022 |
| **cg07017486** | chr11 | 100692664 | 100692664 | 1 | - | ENSG00000082175.15 | PGR | 437150 | 1,44702894326529E-05 | 0,008962731 | G > AG | 0,065200209 |
| **cg06969458** | chr4 | 112818735 | 112818735 | 1 | + | ENSG00000145362.21 | ANK2 | 704 | 1,45679560139074E-05 | 0,008977937 | G > AG | 0,118555135 |
| **cg15330301** | chr15 | 28985515 | 28985515 | 1 | - | ENSG00000261649.6 | GOLGA6L7 | 136839 | 1,45280167372696E-05 | 0,008977937 | G > AG | 0,05925259 |
| **cg04110962** | chr11 | 132712913 | 132712913 | 1 | + | ENSG00000182667.15 | NTM | 1342436 | 1,45769968198663E-05 | 0,008977937 | G > AG | 0,070626948 |
| **cg18553350** | chr13 | 28390061 | 28390061 | 1 | + | ENSG00000152520.14 | PAN3 | 251556 | 1,45733306452925E-05 | 0,008977937 | G > AG | 0,107704808 |
| **cg06774144** | chr1 | 198837467 | 198837467 | 1 | + | ENSG00000081237.20 | PTPRC | 199011 | 1,45589654849234E-05 | 0,008977937 | G > AG | 0,052661658 |
| **cg20515885** | chr15 | 65055406 | 65055406 | 1 | + | ENSG00000186198.4 | SLC51B | 10020 | 1,45522355166669E-05 | 0,008977937 | G > AG | 0,062111482 |
| **cg14335783** | chr1 | 117211696 | 117211696 | 1 | + | ENSG00000116830.12 | TTF2 | 151371 | 1,4530571499651E-05 | 0,008977937 | G > AG | 0,097147806 |
| **cg09070316** | chr1 | 64479085 | 64479085 | 1 | - | ENSG00000162434.13 | JAK1 | 588670 | 1,46214072218031E-05 | 0,008998045 | G > AG | 0,070312812 |
| **cg03310503** | chr6 | 45642099 | 45642099 | 1 | - | ENSG00000196284.17 | SUPT3H | 264145 | 1,46702261539746E-05 | 0,009020831 | G > AG | 0,062925153 |
| **cg22399387** | chr4 | 25486341 | 25486341 | 1 | + | ENSG00000053900.11 | ANAPC4 | 109079 | 1,47320998876114E-05 | 0,009051601 | G > AG | 0,06195151 |
| **cg25456620** | chr20 | 57109373 | 57109373 | 1 | - | ENSG00000101144.13 | BMP7 | 157269 | 1,48342440663663E-05 | 0,009065841 | G > AG | 0,076615101 |
| **cg17092502** | chr11 | 15112203 | 15112203 | 1 | - | ENSG00000110680.13 | CALCA | 139851 | 1,48382381533855E-05 | 0,009065841 | G > AG | 0,143267326 |
| **cg12830671** | chr10 | 71575540 | 71575540 | 1 | + | ENSG00000107736.22 | CDH23 | 178621 | 1,48188794345152E-05 | 0,009065841 | G > AG | 0,084285003 |
| **cg12991093** | chr5 | 43649247 | 43649247 | 1 | + | ENSG00000112992.18 | NNT | 46556 | 1,47885453772343E-05 | 0,009065841 | G > AG | 0,113320169 |
| **cg03233301** | chr13 | 33697022 | 33697022 | 1 | + | ENSG00000133119.13 | RFC3 | 121046 | 1,48335876442901E-05 | 0,009065841 | G > AG | 0,07183246 |
| **cg07514693** | chr13 | 97850324 | 97850324 | 1 | - | ENSG00000139797.8 | RNF113B | 326946 | 1,48318089355425E-05 | 0,009065841 | G > AG | 0,049166961 |
| **cg10072849** | chr18 | 62418063 | 62418063 | 1 | + | ENSG00000141655.17 | TNFRSF11A | 92777 | 1,48022594374629E-05 | 0,009065841 | G > AG | 0,063717199 |

| | | | | | | | | | | | | |
|---|---|---|---|---|---|---|---|---|---|---|---|---|
| **cg06592137** | chr17 | 57951939 | 57951939 | 1 | - | ENSG00000180891.13 | CUEDC1 | 3474 | 1,48602757579304E-05 | 0,00907206 | G > AG | 0,07789074 |
| **cg05948157** | chr14 | 74863905 | 74863905 | 1 | + | ENSG00000119689.15 | DLST | 17985 | 1,49149724059109E-05 | 0,009097978 | G > AG | 0,06534874 |
| **cg24487502** | chr19 | 38145770 | 38145770 | 1 | + | ENSG00000167642.13 | SPINT2 | 98264 | 1,49265180746806E-05 | 0,009097978 | G > AG | 0,07845877 |
| **cg01148766** | chr5 | 11251322 | 11251322 | 1 | + | ENSG00000164236.12 | ANKRD33B | 687253 | 1,49806627691069E-05 | 0,009109205 | G > AG | 0,104897819 |
| **cg09423231** | chr2 | 51026346 | 51026346 | 1 | + | ENSG00000242441.8 | GTF2A1L | 2408549 | 1,49690318717194E-05 | 0,009109205 | G > AG | 0,090654423 |
| **cg04687727** | chr3 | 40691580 | 40691580 | 1 | - | ENSG00000168038.11 | ULK4 | 1270551 | 1,49581240704474E-05 | 0,009109205 | G > AG | 0,079497573 |
| **cg09696051** | chr6 | 72273173 | 72273173 | 1 | - | ENSG00000256980.5 | KHDC1L | 952598 | 1,50843198927168E-05 | 0,009157676 | G > AG | 0,081587778 |
| **cg09692449** | chr17 | 41523965 | 41523965 | 1 | - | ENSG00000171346.16 | KRT15 | 1435 | 1,51104066419382E-05 | 0,009166239 | G > AG | 0,133471815 |
| **cg04804145** | chr3 | 43019048 | 43019048 | 1 | + | ENSG00000144649.9 | GASK1A | 39782 | 1,51460822015678E-05 | 0,0091806 | G > AG | 0,064819463 |
| **cg26831100** | chr19 | 16474207 | 16474207 | 1 | - | ENSG00000127527.14 | EPS15L1 | 2121 | 1,52099586692784E-05 | 0,00920473 | G > AG | 0,075934101 |
| **cg05978555** | chr1 | 46175080 | 46175080 | 1 | + | ENSG00000117472.10 | TSPAN1 | 8 | 1,52419486755107E-05 | 0,009216798 | G > AG | 0,066649982 |
| **cg09132781** | chr14 | 57749421 | 57749421 | 1 | - | ENSG00000151812.15 | SLC35F4 | 232774 | 1,52730181972721E-05 | 0,009228291 | G > AG | 0,122477579 |
| **cg09639890** | chr3 | 192417283 | 192417283 | 1 | + | ENSG00000127252.7 | PLAAT1 | 823844 | 1,53322832392513E-05 | 0,009256788 | G > AG | 0,096321129 |
| **cg13460221** | chr5 | 58056397 | 58056397 | 1 | - | ENSG00000145632.15 | PLK2 | 403743 | 1,54393367502695E-05 | 0,009299402 | G > AG | 0,102549581 |
| **cg08088590** | chr5 | 68747102 | 68747102 | 1 | + | ENSG00000145740.19 | SLC30A5 | 346846 | 1,54374904216556E-05 | 0,009299402 | G > AG | 0,065312586 |
| **cg17646411** | chr12 | 54040350 | 54040350 | 1 | + | ENSG00000172789.4 | HOXC5 | 7301 | 1,54854462199143E-05 | 0,009305194 | G > AG | 0,083696902 |
| **cg04893217** | chr9 | 14310316 | 14310316 | 1 | + | ENSG00000164975.15 | SNAPC3 | 1112387 | 1,54751357059871E-05 | 0,009305194 | G > AG | 0,149017824 |
| **cg06096913** | chr13 | 42656240 | 42656240 | 1 | + | ENSG00000120659.15 | TNFSF11 | 93505 | 1,54737855105684E-05 | 0,009305194 | G > AG | 0,083522946 |
| **cg11931139** | chr3 | 32897413 | 32897413 | 1 | + | ENSG00000183813.7 | CCR4 | 54230 | 1,55057669943863E-05 | 0,009310091 | G > AG | 0,037929537 |
| **cg02879457** | chr2 | 60847969 | 60847969 | 1 | - | ENSG00000162927.14 | PUS10 | 170291 | 1,55383543241917E-05 | 0,009318045 | G > AG | 0,079188034 |
| **cg15221192** | chr5 | 61525773 | 61525773 | 1 | - | ENSG00000188725.8 | SMIM15 | 363304 | 1,55433769792765E-05 | 0,009318045 | G > AG | 0,052988456 |
| **cg01946023** | chr1 | 214464471 | 214464471 | 1 | + | ENSG00000117724.13 | CENPF | 138723 | 1,55721146694083E-05 | 0,009321355 | G > AG | 0,083706493 |
| **cg25379426** | chr5 | 39891522 | 39891522 | 1 | - | ENSG00000153071.15 | DAB2 | 429221 | 1,56520271543766E-05 | 0,009344 | G > AG | 0,07820843 |
| **cg05993870** | chr4 | 115671867 | 115671867 | 1 | - | ENSG00000138653.10 | NDST4 | 557990 | 1,56599631837018E-05 | 0,009344 | G > AG | 0,094662802 |
| **cg13529513** | chr9 | 36880058 | 36880058 | 1 | + | ENSG00000147905.18 | ZCCHC7 | 240515 | 1,56529591406312E-05 | 0,009344 | G > AG | 0,078406837 |
| **cg02989724** | chr20 | 24584621 | 24584621 | 1 | + | ENSG00000101463.6 | SYNDIG1 | 114993 | 1,56807635037353E-05 | 0,009347935 | G > AG | 0,068976572 |
| **cg07819175** | chr21 | 28986918 | 28986918 | 1 | + | ENSG00000156256.15 | USP16 | 37710 | 1,5691000074607E-05 | 0,009347935 | G > AG | 0,066540757 |
| **cg16876636** | chr2 | 169637729 | 169637729 | 1 | - | ENSG00000154479.13 | CCDC173 | 56677 | 1,57142631408415E-05 | 0,009354509 | G > AG | 0,068176377 |
| **cg16415646** | chr7 | 41703403 | 41703403 | 1 | + | ENSG00000106591.4 | MRPL32 | 1228972 | 1,57982584104883E-05 | 0,009397197 | G > AG | 0,119911499 |
| **cg02373664** | chr8 | 111027416 | 111027416 | 1 | - | ENSG00000164794.9 | KCNV1 | 1051644 | 1,58548619055098E-05 | 0,009408917 | G > AG | 0,074593371 |
| **cg18055501** | chr6 | 155399925 | 155399925 | 1 | + | ENSG00000171217.5 | CLDN20 | 135913 | 1,58849010562041E-05 | 0,009419436 | G > AG | 0,092004862 |

| | | | | | | | | | | | | |
|---|---|---|---|---|---|---|---|---|---|---|---|---|
| **cg17059507** | chr12 | 49924820 | 49924820 | 1 | - | ENSG00000135472.9 | FAIM2 | 20602 | 1,59230119935228E-05 | 0,009428743 | G > AG | 0,075347211 |
| **cg01694400** | chr6 | 127162955 | 127162955 | 1 | + | ENSG00000146374.14 | RSPO3 | 44285 | 1,59252489692982E-05 | 0,009428743 | G > AG | 0,076511364 |
| **cg13538474** | chr9 | 28364375 | 28364375 | 1 | - | ENSG00000174482.10 | LINGO2 | 305912 | 1,59775042876693E-05 | 0,009446472 | G > AG | 0,080053281 |
| **cg06700142** | chr10 | 49081715 | 49081715 | 1 | + | ENSG00000177354.12 | C10orf71 | 217454 | 1,60178843309824E-05 | 0,00946162 | G > AG | 0,107551111 |
| **cg27510725** | chr8 | 118015219 | 118015219 | 1 | + | ENSG00000164758.7 | MED30 | 494507 | 1,60321109688902E-05 | 0,009462716 | G > AG | 0,05591577 |
| **cg04353193** | chr20 | 17990725 | 17990725 | 1 | + | ENSG00000125871.14 | MGME1 | 21708 | 1,6059377979482E-05 | 0,009471502 | G > AG | 0,097176558 |
| **cg24309040** | chr12 | 111226796 | 111226796 | 1 | - | ENSG00000198324.14 | PHETA1 | 142326 | 1,60787263133903E-05 | 0,009475607 | G > AG | 0,075873921 |
| **cg26463157** | chr16 | 1291289 | 1291289 | 1 | - | ENSG00000007520.4 | TSR3 | 60590 | 1,60961163712573E-05 | 0,009478553 | G > AG | 0,074550001 |
| **cg11378575** | chr2 | 231400293 | 231400293 | 1 | - | ENSG00000115053.17 | NCL | 83349 | 1,61546601731034E-05 | 0,00950571 | G > AG | 0,12232388 |
| **cg09863230** | chr2 | 6506449 | 6506449 | 1 | - | ENSG00000134326.12 | CMPK2 | 360187 | 1,61886760159393E-05 | 0,009518404 | G > AG | 0,083390696 |
| **cg20168495** | chr4 | 147361914 | 147361914 | 1 | - | ENSG00000164169.13 | PRMT9 | 322250 | 1,62171771741592E-05 | 0,009527838 | G > AG | 0,133650027 |
| **cg02627528** | chr2 | 30865733 | 30865733 | 1 | - | ENSG00000162949.16 | CAPN13 | 45190 | 1,62520321107766E-05 | 0,009534604 | G > AG | 0,082442793 |
| **cg00230450** | chr12 | 14823294 | 14823294 | 1 | - | ENSG00000179256.2 | SMCO3 | 9111 | 1,62536212866019E-05 | 0,009534604 | G > AG | 0,07330153 |
| **cg15077577** | chr8 | 8382859 | 8382859 | 1 | - | ENSG00000275342.5 | PRAG1 | 3581 | 1,63152160453218E-05 | 0,009563402 | G > AG | 0,067047618 |
| **cg15429600** | chr10 | 96794759 | 96794759 | 1 | - | ENSG00000155629.15 | PIK3AP1 | 74244 | 1,63528237988802E-05 | 0,009578107 | G > AG | 0,101343441 |
| **cg26915343** | chr2 | 18721573 | 18721573 | 1 | + | ENSG00000170745.12 | KCNS3 | 843727 | 1,64043352441861E-05 | 0,009600927 | G > AG | 0,079170041 |
| **cg22716950** | chr4 | 111651038 | 111651038 | 1 | + | ENSG00000174749.6 | FAM241A | 494415 | 1,64266568274462E-05 | 0,009606641 | G > AG | 0,065365458 |
| **cg08091050** | chr4 | 26206429 | 26206429 | 1 | - | ENSG00000163394.6 | CCKAR | 284056 | 1,64541068630323E-05 | 0,009615343 | G > AG | 0,126049297 |
| **cg12734189** | chr6 | 52724327 | 52724327 | 1 | - | ENSG00000244067.3 | GSTA2 | 39149 | 1,65657907566697E-05 | 0,009634972 | G > AG | 0,09428362 |
| **cg04779473** | chr6 | 105210279 | 105210279 | 1 | + | ENSG00000187772.8 | LIN28B | 273664 | 1,65533032440022E-05 | 0,009634972 | G > AG | 0,083279059 |
| **cg00884004** | chr5 | 31809709 | 31809709 | 1 | + | ENSG00000133401.16 | PDZD2 | 170579 | 1,65451635355692E-05 | 0,009634972 | G > AG | 0,097151361 |
| **cg23657355** | chr1 | 38572359 | 38572359 | 1 | - | ENSG00000116954.8 | RRAGC | 287414 | 1,65758672317012E-05 | 0,009634972 | G > AG | 0,104925681 |
| **cg18154417** | chr1 | 47934401 | 47934401 | 1 | - | ENSG00000269113.4 | TRABD2B | 62985 | 1,65289600214039E-05 | 0,009634972 | G > AG | 0,072342611 |
| **cg25112366** | chr5 | 176743137 | 176743137 | 1 | + | ENSG00000113763.12 | UNC5A | 67381 | 1,66780008496546E-05 | 0,009686978 | G > AG | 0,064923987 |
| **cg03224756** | chr16 | 73192511 | 73192511 | 1 | - | ENSG00000140836.17 | ZFHX3 | 699361 | 1,67048649758817E-05 | 0,009687869 | G > AG | 0,08140572 |
| **cg14157525** | chr16 | 58127138 | 58127138 | 1 | + | ENSG00000102996.5 | MMP15 | 101385 | 1,67433118376763E-05 | 0,00970281 | G > AG | 0,078055403 |
| **cg23015950** | chr17 | 78563349 | 78563349 | 1 | - | ENSG00000187775.17 | DNAH17 | 14048 | 1,68425312670307E-05 | 0,009716144 | G > AG | 0,089897449 |
| **cg08926977** | chr8 | 70781865 | 70781865 | 1 | - | ENSG00000147592.9 | LACTB2 | 112679 | 1,68038436111806E-05 | 0,009716144 | G > AG | 0,117081349 |
| **cg13584406** | chr12 | 76018403 | 76018403 | 1 | - | ENSG00000139289.14 | PHLDA1 | 13374 | 1,68313940245434E-05 | 0,009716144 | G > AG | 0,106868738 |
| **cg01404407** | chr6 | 48080685 | 48080685 | 1 | - | ENSG00000244694.8 | PTCHD4 | 30513 | 1,68126625460334E-05 | 0,009716144 | G > AG | 0,103821943 |
| **cg06298729** | chr11 | 48096489 | 48096489 | 1 | + | ENSG00000149177.13 | PTPRJ | 115932 | 1,68332657662973E-05 | 0,009716144 | G > AG | 0,083253851 |

| | | | | | | | | | | | | |
|---|---|---|---|---|---|---|---|---|---|---|---|---|
| **cg23283358** | chr18 | 13652048 | 13652048 | 1 | + | ENSG00000101654.18 | RNMT | 74611 | 1,67836091447427E-05 | 0,009716144 | G > AG | 0,098320059 |
| **cg23398487** | chr13 | 106756567 | 106756567 | 1 | + | ENSG00000182346.21 | DAOA | 1290701 | 1,69608542271087E-05 | 0,009777029 | G > AG | 0,101791788 |
| **cg07230654** | chr4 | 152329730 | 152329730 | 1 | + | ENSG00000164144.16 | ARFIP1 | 450206 | 1,69830343830709E-05 | 0,009782443 | G > AG | 0,063359006 |
| **cg13786163** | chr6 | 1446236 | 1446236 | 1 | + | ENSG00000137273.6 | FOXF2 | 56661 | 1,70425001539816E-05 | 0,009800032 | G > AG | 0,097189308 |
| **cg21463368** | chr6 | 166849537 | 166849537 | 1 | - | ENSG00000071242.12 | RPS6KA2 | 56915 | 1,70409420482317E-05 | 0,009800032 | G > AG | 0,104519428 |
| **cg01015776** | chr1 | 85818617 | 85818617 | 1 | - | ENSG00000117174.11 | ZNHIT6 | 110183 | 1,70520047721491E-05 | 0,009800032 | G > AG | 0,068408137 |
| **cg26112574** | chr1 | 94299579 | 94299579 | 1 | - | ENSG00000137962.13 | ARHGAP29 | 24510 | 1,70908627227424E-05 | 0,00981499 | G > AG | 0,119327544 |
| **cg25611671** | chr2 | 14616389 | 14616389 | 1 | + | ENSG00000162981.14 | LRATD1 | 16310 | 1,71101584953573E-05 | 0,0098187 | G > AG | 0,098373732 |
| **cg15044147** | chr10 | 50327208 | 50327208 | 1 | - | ENSG00000188611.17 | ASAH2 | 47487 | 1,71775402525073E-05 | 0,009849978 | G > AG | 0,091488583 |
| **cg13850846** | chr9 | 84920368 | 84920368 | 1 | + | ENSG00000148053.17 | NTRK2 | 251818 | 1,71955362310102E-05 | 0,009852911 | G > AG | 0,044707806 |
| **cg21584710** | chr17 | 74310002 | 74310002 | 1 | + | ENSG00000196169.15 | KIF19 | 16207 | 1,72277049602098E-05 | 0,009863955 | G > AG | 0,053753771 |
| **cg27171704** | chr17 | 47708644 | 47708644 | 1 | + | ENSG00000198933.9 | TBKBP1 | 14564 | 1,72506429923564E-05 | 0,009869701 | G > AG | 0,124845565 |
| **cg01941747** | chr1 | 214018455 | 214018455 | 1 | + | ENSG00000117707.16 | PROX1 | 35275 | 1,73751041619255E-05 | 0,009918654 | G > AG | 0,078116705 |
| **cg13537061** | chr7 | 35259109 | 35259109 | 1 | - | ENSG00000164532.11 | TBX20 | 5008 | 1,7373052829436E-05 | 0,009918654 | G > AG | 0,041589004 |
| **cg02063944** | chr13 | 44854934 | 44854934 | 1 | - | ENSG00000083635.8 | NUFIP1 | 134538 | 1,74275304295722E-05 | 0,009933755 | G > AG | 0,104625265 |
| **cg12940993** | chr14 | 20802989 | 20802989 | 1 | - | ENSG00000129538.14 | RNASE1 | 133 | 1,74443656964419E-05 | 0,009935947 | G > AG | 0,12012214 |
| **cg08251860** | chr16 | 81473786 | 81473786 | 1 | + | ENSG00000153815.17 | CMIP | 28979 | 1,75035713808991E-05 | 0,00995978 | G > AG | 0,101332409 |
| **cg12170044** | chr8 | 10488129 | 10488129 | 1 | - | ENSG00000253649.5 | PRSS51 | 59457 | 1,7583432929573E-05 | 0,009985416 | G > AG | 0,055000703 |
