## Supplementary material for "An integrative multi-omics framework identifies epigenetic dysregulation of *HAND2* as a potential primary driver of impaired enteric neural crest cell differentiation in Hirschsprung Disease": Suppl. Table 6

**Suppl. Table 11:** Covariates for MOFA.

| |
|---|
| Sex |
| Trisomy 21 |
| Comorbidities |
| Long-segment Hirschsprung disease (>30cm aganglionosis = via left flexure) |
| Occurrence of stenosis that required bougienage for >10 days |
| Occurrence of enterocolitis (if 3/7 positive: >3x stool/d, fever, meteorism, vomiting, InfektLab+, antibiotics, KH) -preoperatively- |
| Occurrence of enterocolitis (if 3/7 positive: >3x stool/d, fever, meteorism, vomiting, InfektLab+, antibiotics, KH) -postoperative - after 1 year- |
| Occurrence of constipation (Rome III criteria) -postoperatively, overall- |
| Occurrence of constipation (Rome III criteria) -postoperatively - after 1 year- |
| Birth weight (in g) |
| Length of aganglionic piece (in cm) |
| Length of resected piece (in cm) |
| Tissue origin (ganglionic, aganglionic) |
